# Clinical and genomic signatures of rising SARS-CoV-2 Delta breakthrough infections in New York

**DOI:** 10.1101/2021.12.07.21267431

**Authors:** Ralf Duerr, Dacia Dimartino, Christian Marier, Paul Zappile, Samuel Levine, Fritz François, Eduardo Iturrate, Guiqing Wang, Meike Dittmann, Jennifer Lighter, Brian Elbel, Andrea B. Troxel, Keith S. Goldfeld, Adriana Heguy

**Affiliations:** Department of Microbiology, NYU Grossman School of Medicine; Genome Technology Center, Office of Science and Research, NYU Langone Health; Hospital Operations, NYU Langone Health; Department of Medicine, NYU Grossman School of Medicine; Department of Pathology, NYU Grossman School of Medicine; Department of Pediatric Infectious Diseases, NYU Grossman School of Medicine; Department of Population Health, NYU Grossman School of Medicine; NYU Wagner Graduate School of Public Service

## Abstract

In 2021, Delta has become the predominant SARS-CoV-2 variant worldwide. While vaccines effectively prevent COVID-19 hospitalization and death, vaccine breakthrough infections increasingly occur. The precise role of clinical and genomic determinants in Delta infections is not known, and whether they contribute to increased rates of breakthrough infections compared to unvaccinated controls. Here, we show a steep and near complete replacement of circulating variants with Delta between May and August 2021 in metropolitan New York. We observed an increase of the Delta sublineage AY.25, its spike mutation S112L, and nsp12 mutation F192V in breakthroughs. Delta infections were associated with younger age and lower hospitalization rates than Alpha. Delta breakthroughs increased significantly with time since vaccination, and, after adjusting for confounders, they rose at similar rates as in unvaccinated individuals. Our data indicate a limited impact of vaccine escape in favor of Delta’s increased epidemic growth in times of waning vaccine protection.

## Introduction

The SARS-CoV-2 pandemic has been a showcase for observing viral evolution in real time. New variants emerged in various parts of the globe and caused waves of infection that reached different countries in rapid succession. Variants being monitored (VBM), particularly Alpha, Beta, and Gamma, and variant of concerns (VOC) Delta and, most recently, Omicron emerged and subsequently increased despite worldwide vaccination efforts^1–6^. In the United States, vaccinations started in late December 2020, with three vaccines currently employed: two mRNA-based vaccines, BNT162b2 (Pfizer/BioNTech), now FDA-approved, mRNA-1273 (Moderna), and the adenovirus-based Janssen COVID-19 vaccine, JNJ-78436735^7–9^. All three vaccines utilize the spike sequence of SARS-CoV-2 Wuhan-Hu-1 isolated in January 2020^10, 11^, and thus their epitopes are not perfectly matched to those of currently circulating variants. Despite this mismatch, the vaccines are highly effective at preventing symptomatic disease, hospitalization, death, and forward transmission^12–14^. However, post-vaccination infections (vaccine breakthroughs) do occur. The epidemiological data on whether these breakthroughs are driven by properties inherent to specific variants are controversial. Two studies earlier in 2021 indicated increased breakthrough rates of the Beta or Gamma variants following two doses of mRNA vaccines^15, 16^. In contrast, other studies, including our own, found that breakthrough cases had a similar variant distribution as in unvaccinated individuals in the respective cohort at that specific time^17–20^.

In 2021, Delta became the predominant variant globally. Data from the CDC indicate that coincident with the rise of Delta in the US, vaccine effectiveness decreased from 91% to 66%, which is consistent with waning immunity observed in Israel and Qatar^21–23^. Delta has been associated with higher replication and transmission^24, 25^, and *in vitro* studies report slightly decreased neutralizing efficacy of monoclonal antibodies, convalescent sera or sera from vaccinated individuals against Delta, suggesting that some mutations present in Delta may cause partial immune escape^26–28^. Clinical and full SARS-CoV-2 genome data from post-vaccination infections in the era of Delta are still scarce^29^ but will be key to revealing features critical for vaccine effectiveness/escape and to identify commonalities and differences with the upcoming Omicron variant and other future VOCs. Here, we determined the SARS-CoV-2 genetic makeup of 132 post-vaccination infections and their associated clinical characteristics in fully vaccinated individuals within NYU Langone Health (NYULH), a large metropolitan New York healthcare system with hospitals across the region. We assessed the probability of Delta to result in breakthrough infection relative to other variants between May and August 2021.

## Results

### Demographic parameters in our cohort of vaccinated and unvaccinated SARS-CoV-2-infected participants

Between May 1^st^ and August 3^rd^, 2021, a total of n=1613 SARS-CoV-2 infections were recorded in our multicenter healthcare system. The majority of SARS-CoV-2-positive tests (82%) were from unvaccinated individuals (n=1297), whereas 18% were from vaccinated (n=297) among a total of 168,127 fully vaccinated individuals within our system (**Table 1**). We sequenced all breakthrough cases with available specimens and randomly selected as many specimens from unvaccinated individuals as allowed by our internal weekly processing capacity (up to 96), irrespective of demographic or clinical parameters. We obtained high quality SARS-CoV-2 sequences from 132 vaccinated and 283 unvaccinated individuals. The median age (37 versus 42 years) and sex distribution were similar in both groups. The majority of recorded breakthrough infections (63%) occurred at >120 days after full vaccination with a median of 136 days after vaccination (**Table 1**).

**Table 1.**
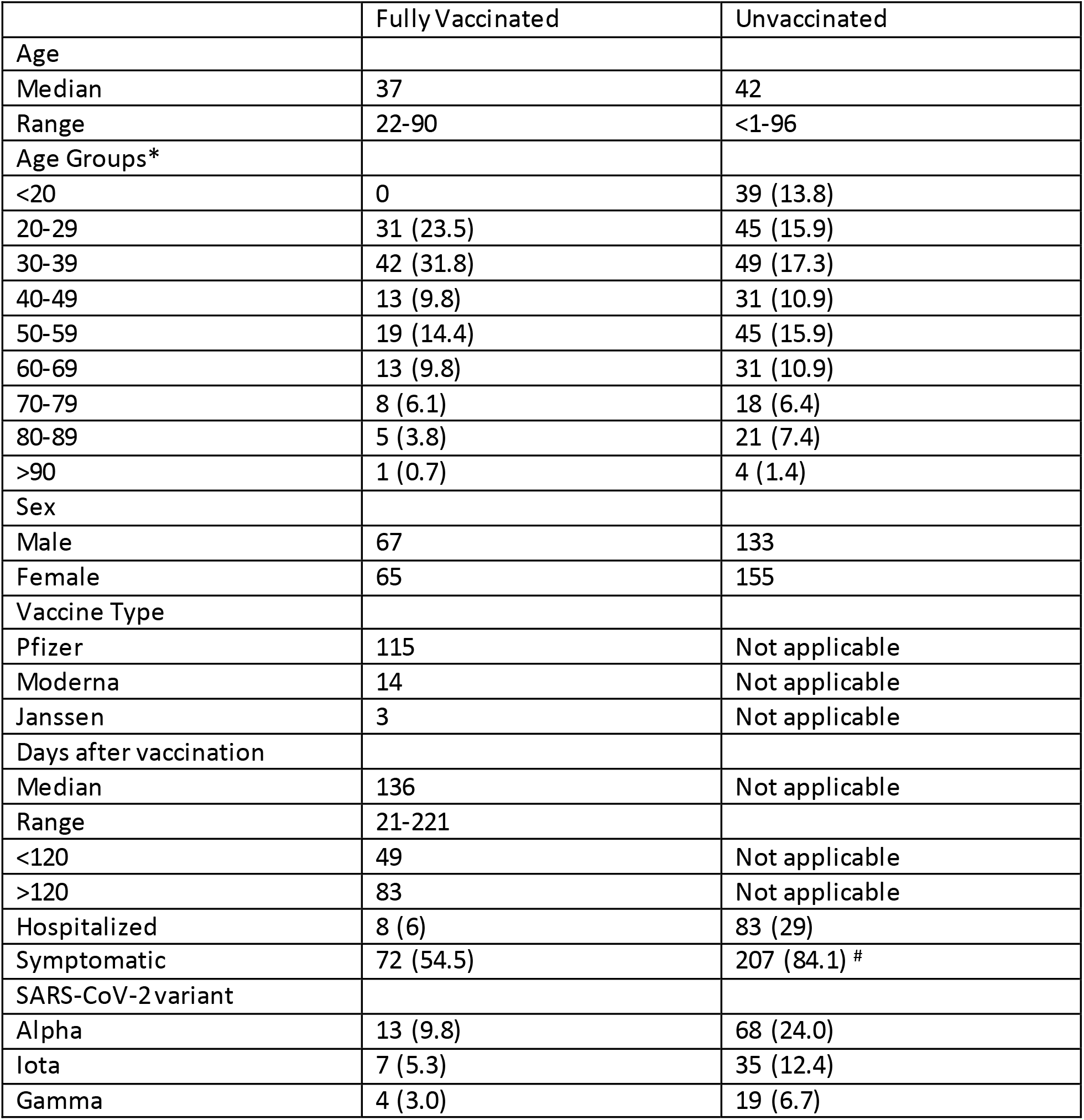
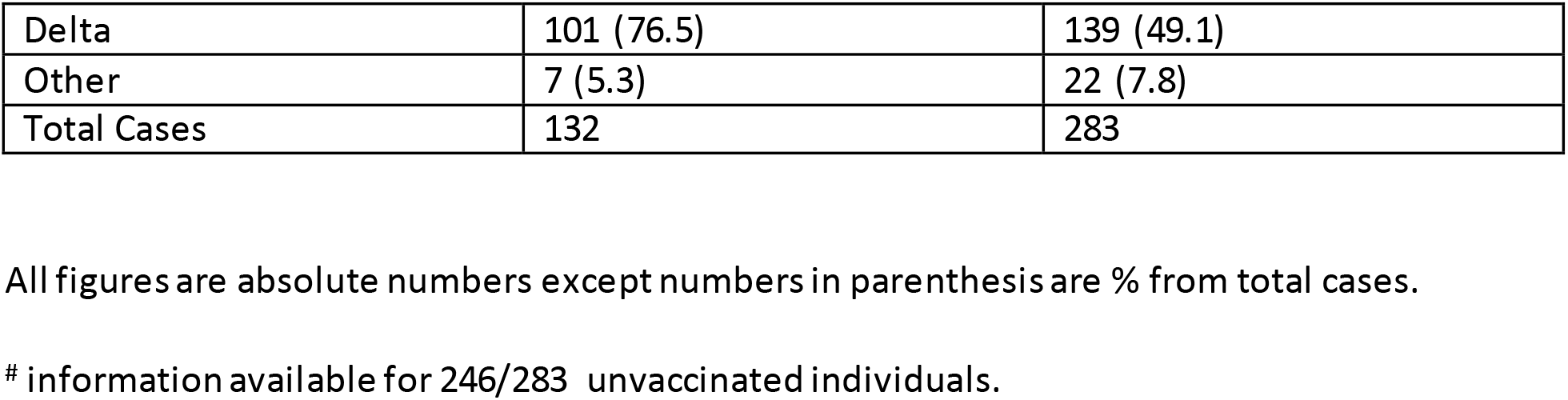
Demographic and clinical data and SARS-CoV-2 lineages of fully vaccinated vs non-vaccinated COVID-19 cases with full SARS-CoV-2 genomes

|  | Fully Vaccinated | Unvaccinated |
| --- | --- | --- |
| Age |  |  |
| Median | 37 | 42 |
| Range | 22-90 | <1-96 |
| Age Groups* |  |  |
| <20 | 0 | 39 (13.8) |
| 20-29 | 31 (23.5) | 45 (15.9) |
| 30-39 | 42 (31.8) | 49 (17.3) |
| 40-49 | 13 (9.8) | 31 (10.9) |
| 50-59 | 19 (14.4) | 45 (15.9) |
| 60-69 | 13 (9.8) | 31 (10.9) |
| 70-79 | 8 (6.1) | 18 (6.4) |
| 80-89 | 5 (3.8) | 21 (7.4) |
| >90 | 1 (0.7) | 4 (1.4) |
| Sex |  |  |
| Male | 67 | 133 |
| Female | 65 | 155 |
| Vaccine Type |  |  |
| Pfizer | 115 | Not applicable |
| Moderna | 14 | Not applicable |
| Janssen | 3 | Not applicable |
| Days after vaccination |  |  |
| Median | 136 | Not applicable |
| Range | 21-221 |  |
| <120 | 49 | Not applicable |
| >120 | 83 | Not applicable |
| Hospitalized | 8 (6) | 83 (29) |
| Symptomatic | 72 (54.5) | 207 (84.1) # |
| SARS-CoV-2 variant |  |  |
| Alpha | 13 (9.8) | 68 (24.0) |
| Iota | 7 (5.3) | 35 (12.4) |
| Gamma | 4 (3.0) | 19 (6.7) |
| Delta | 101 (76.5) | 139 (49.1) |
| Other | 7 (5.3) | 22 (7.8) |
| Total Cases | 132 | 283 |

All figures are absolute numbers except numbers in parenthesis are % from total cases.

### Delta infections were associated with a distinct network of clinical and demographic predictors

More than 45% of the breakthrough infections in our cohort were asymptomatic (n=60); only 6% (n=8) were hospitalized, all of whom were > 50 years (5/8 >70 years) and had co-morbidities. There was one death from metastatic cancer (**Table S1, Figs S1, S2**). The hospitalization rate was significantly higher in the unvaccinated (29%, n=83, *P*<0.0001); for 66 of those, COVID-19 was the main reason for hospitalization, 10 required ICU admission, and seven died from COVID-19 (**Table S2**). To identify differential features of the circulating variants, we compared their clinical and demographic characteristics (**Figs. 1a, b, S1-S3**). Among the SARS-CoV-2-positive cases studied, hospitalizations due to Delta were significantly lower compared with Alpha, Gamma, Iota, and the other variants combined, and significance was maintained against Alpha when assessed separately in vaccinated and unvaccinated individuals (**Fig. S1**). Among Delta infections, we recorded only one COVID-19 related death (0.7%), whereas Alpha had the highest rates with at least four COVID-19-related deaths (5.9%) (**Tables S1, S2**). Delta affected younger individuals, particularly when compared to Alpha. Hospitalizations were associated with older age, specifically in Alpha, Delta, and Iota infections. There was a trend to lower cycle threshold (Ct) values in Delta infections, which translated into a weak but significant inverse correlation between Delta and Ct values in the overall data set and among breakthrough infections (**Figs. 1b, S2a**). The Ct values did not differ in hospitalized versus non-hospitalized. In all, Delta infections exhibited acharacteristic clinical profile regarding age (younger), time of infection (later in the year) and hospitalization rate (lower), all of which significantly correlated with each otherand might be mutual drivers (**Fig. 1b**).

**Figure 1.**
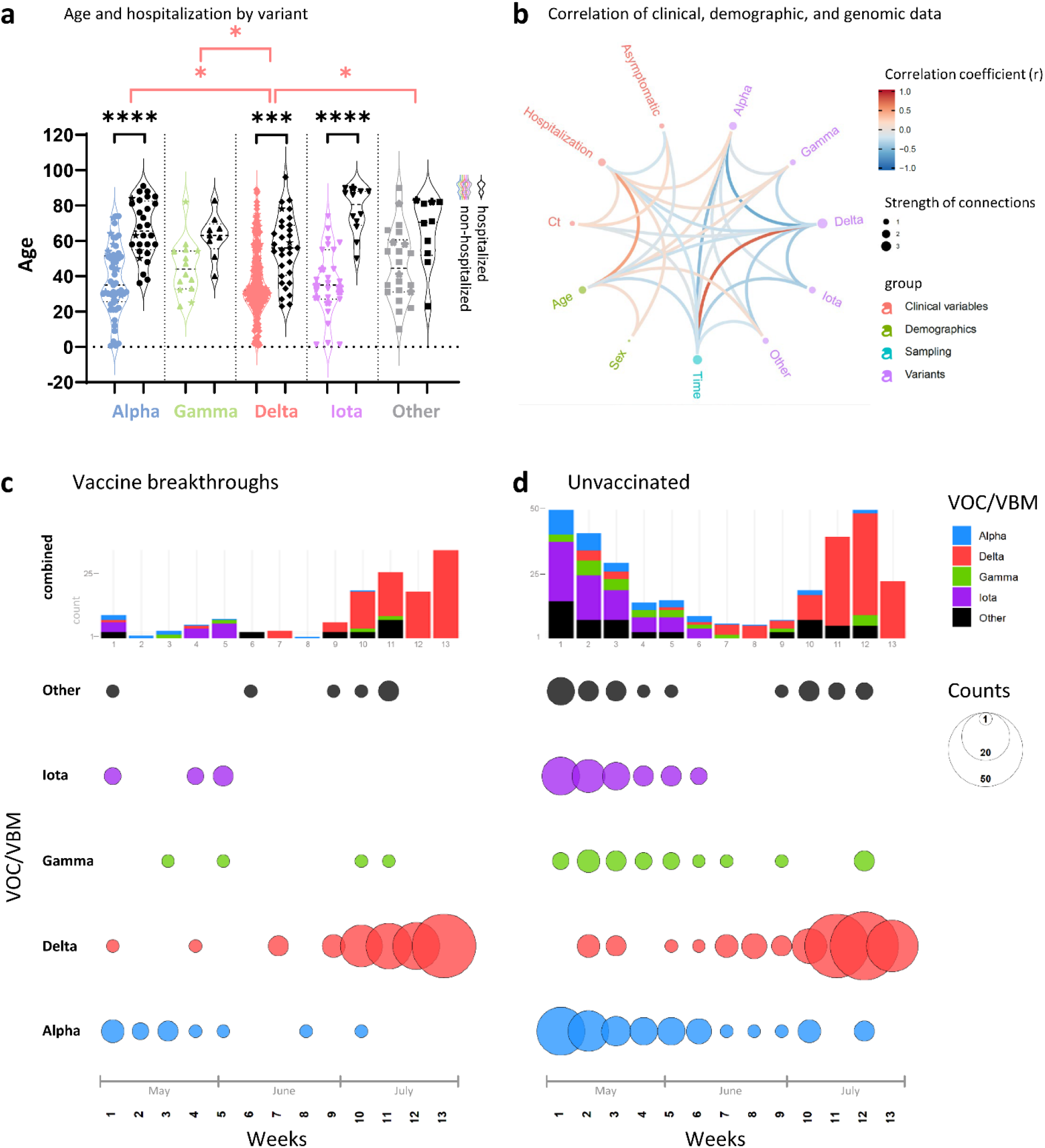
Relationship of clinical, demographic, and genomic data. **a**, Violin plot summarizing the age distribution of the combined data set of 132 vaccinated and 283 unvaccinated SARS-CoV-2-positive individuals by variant. The pairs of colored and black violins show non-hospitalized versus hospitalized cases per variant. Horizontal lines indicate the median and interquartile ranges of values. Breakthrough cases are shown as stars. Statistical comparisons of age were made using Kruskal-Wallis tests between variants (red brackets) and, for each variant, between non-hospitalized and hospitalized cases (black brackets). All statistically significant results are shown: * P<0.05, ** P<0.01, *** P<0.001, **** P<0.0001. **b**, Correlation analysis of clinical, demographic, and genomic data of SARS-CoV-2 infected individuals (as in A). Red and blue edges represent positive and negative correlations between connected variables, respectively, according to the scale of r values to the right. Only significant correlations (P<0.05, Spearman rank test) are displayed. Nodes are color-coded based on the grouping of variables. Node size corresponds to the strength of correlations. Ct: Cycle threshold in RT-PCR; **s**ex: male sex; time: date of sampling. **c, d**, Distribution of variants (top) and absolute variant counts (bottom), colored and separated into to the most frequently detected variants of concern (VOC) and being monitored (VBM) and all other variants combined (Other). Vaccine breakthrough infections (**c**) are shown side-by-side with unvaccinated controls (**d**) on a weekly basis starting May 1^st^ 2021 (all full weeks shown).

### Delta rapidly replaced Alpha and Iota variants in both unvaccinated and vaccine breakthrough cases recorded at NYULH

At the beginning of our study period (May 2021), several variants circulated in the New York metropolitan area, mainly Alpha, Iota and Gamma (**Fig. 1c, d**). In June, we observed a nadir of case numbers (<10 per week), followed by a rise of Delta infections starting end of June that caused a dramatic increase in total cases. This rapid shift in variants towards Delta was detected in both vaccinated and unvaccinated individuals. By the end of our study, we saw a near complete replacement of the locally-originating Iota and the imported Alpha and Gamma variants by Delta.

### Delta infections including breakthroughs exhibit a characteristic subvariant makeup

To determine the diversity and relatedness of SARS-CoV-2 genomes, we performed phylogenetic analyses including our 132 breakthrough and 283 unvaccinated cases, using US and global reference sequences (**Figs. 2a, S4** and **Table S3**). Our SARS-CoV-2 sequences covered a broad set of variants with predominance of Delta and its numerous subvariants, Alpha, Gamma, Iota, Mu, and additional B.1 lineages (**Fig. 2b**). Delta sequences appeared more densely packed and clustered, indicative of transmission chains (**Figs. 2A, S4**). One of these clusters consisted of the Delta subvariant AY.25 that emerged recently and included sequences with spike mutation S112L. We found no evidence of transmission clusters composed solely of breakthrough infections. Alpha and Iota were overall less frequently found among breakthrough infections (odds ratios and 95% confidence intervals (95%CI) of 0.35 (0.19-0.66) and 0.25 (0.09-0.69), respectively), whereas Delta counts were more frequent overthe study period, which was primarily due to B.1.617.2 and AY.25 (odds ratios and 95%CI of 2.12 (1.34-3.36) and 1.97 (1.01-3.73), respectively (**Fig. 2b**). The S112L-positive AY.25 sub-cluster contained SARS-CoV-2 sequences from one unvaccinated and five vaccinated cases (**Figs. 3, S5, S6**), with four cases occurring in New York County.

**Figure 2.**
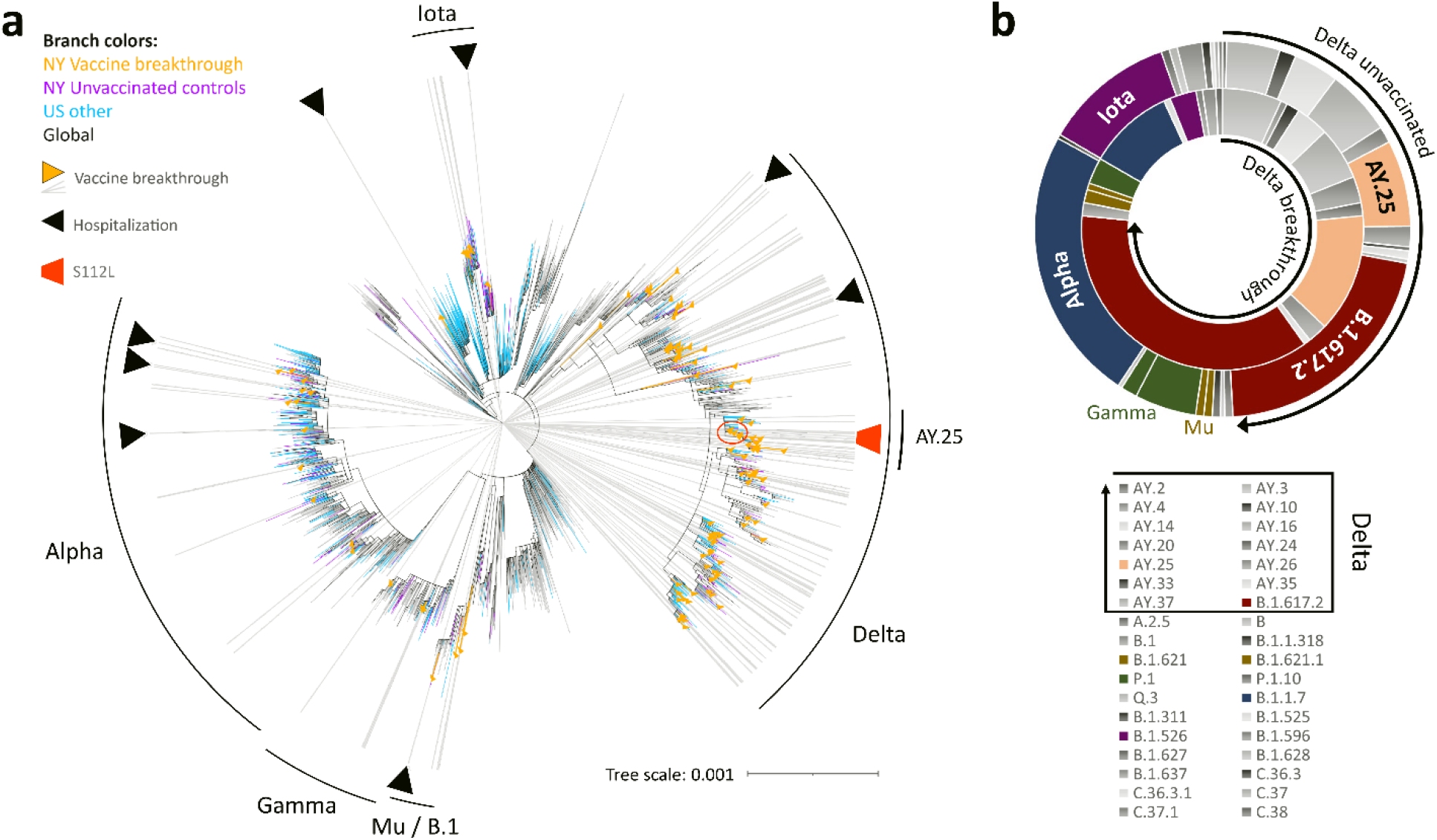
Phylogenetic analysis and variant distribution of SARS-CoV-2 vaccine breakthrough and unvaccinated control sequences. **a**, Maximum likelihood (IQ) tree of 3511 SARS-CoV-2 full genome sequences (base pairs 202-29,666 according to Wuhan-Hu-1 as reference), including 132 vaccine breakthrough (orange) and 283 unvaccinated control SARS-CoV-2 sequences from the NYU Langone Health cohort (greater NYC area) (purple) together with 920 other US (non-NYU; cyan) and 2176 global (non-US; black) reference sequences. The substitution scale of the tree, generated with 1000 bootstrap replicates and Wuhan/WH01/2019-12-26 as root, is indicated at the bottom right. Vaccine breakthrough sequences are highlighted by orange triangles (as branch symbols) and gray rays radiating from the root to the outer rim of the tree. Hospitalizations among vaccine breakthrough infections are indicated by black triangles. The variants responsible for most vaccine breakthrough infections are labeled. The Delta plus S112L sub-lineage is encircled and highlighted with a red trapezoid symbol. **b**, Double-donut plot to compare the variant distribution of breakthrough (inner ring) and unvaccinated control sequences (outer ring). The most abundant variants and Delta subvariants (highlighted by black arrows) are shown in color and labelled in the plot (outer ring only). The color code for all detected variants (Pango lineages) is provided below.

**Figure 3.**
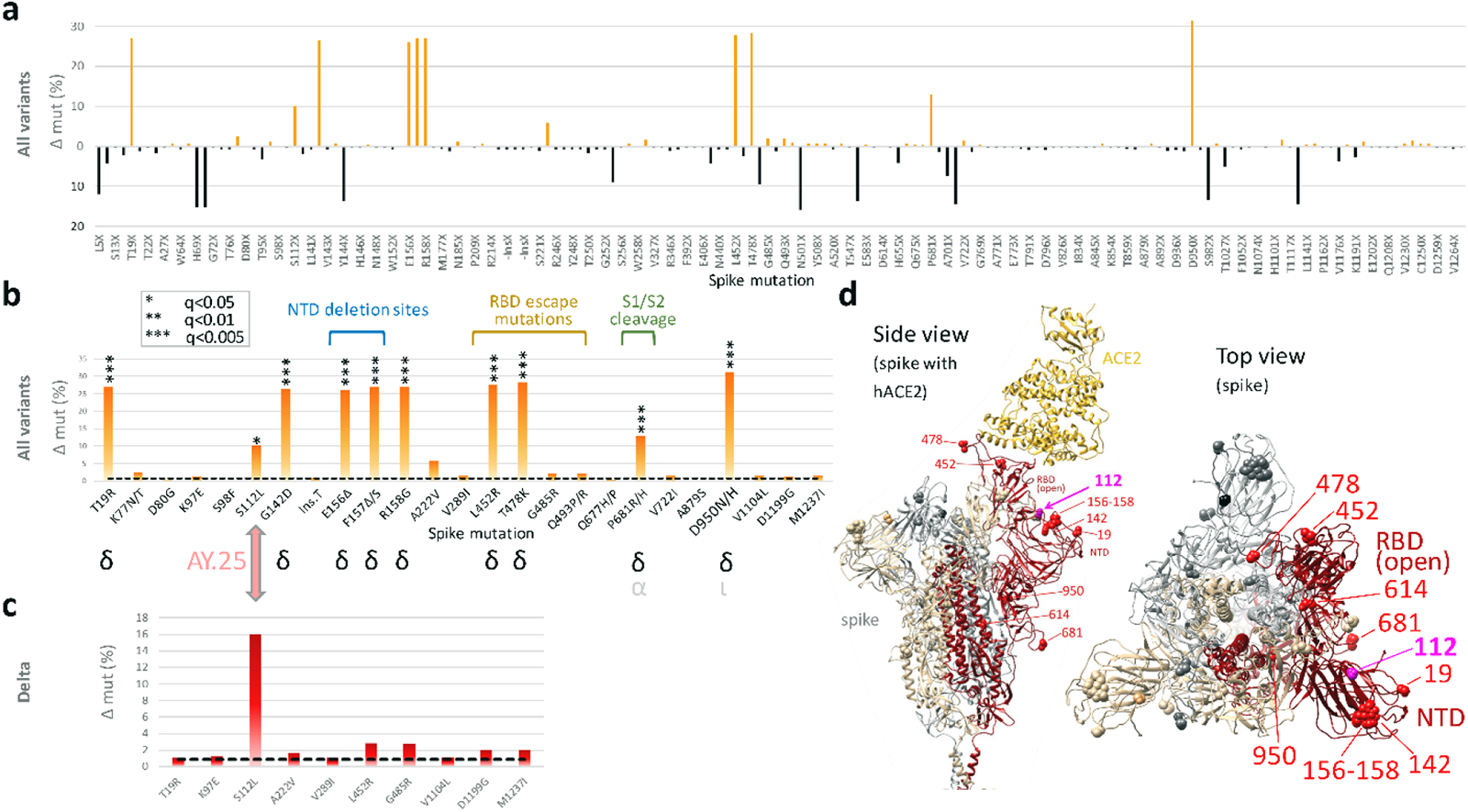
**a**, Site-specific amino acid mutation (mut) frequencies in spike in 132 vaccine breakthrough sequences compared to 283 unvaccinated controls from the same cohort. The Wuhan-Hu-1 sequence served as reference. The mirror plot displays differences of mutation frequencies per spike residue between vaccinated and unvaccinated groups, shown along the x-axis (n=168); orange (facing up) and black bars (facing down) refer to elevated mutation frequencies in vaccinated or unvaccinated individuals, respectively. **b**, Enriched spike mutations in vaccine breakthrough sequences compared to unvaccinated controls. Unique occurrences of mutations in breakthrough cases were disregarded. The dashed black line indicates the average mutation frequency across all spike residues in the unvaccinated control data set (n=283) compared to Wuhan-Hu-1. Significantly enriched mutations in Fisher exact tests are indicated by asterisks (* P<0.05, *** P<0.005) and the variants in which these mutations were found are shown below (black: main source, gray: secondary source). Mutations in the spike N-terminal domain (NTD), receptor binding domain (RBD), and near the S1/S2 interface associated with neutralization escape and/or affecting important biological functions are labeled. **c**, The same analysis as in (**b**) but focusing on Delta sequences exclusively. 101 Delta vaccine breakthrough sequences were compared to 139 Delta unvaccinated controls. The dashed black line indicates the average mutation frequency across all spike residues in the Delta unvaccinated control data set compared to Wuhan-Hu-1 as reference (n=139). **d**, Structural analysis of mutation sites on a spike trimer bound to human ACE2 (hACE2) (pdb S_ACE2). Each protomer is colored differently. The hACE2-bound protomer with the RBD in the “up” position is shown in red. Statistically enriched mutation sites in vaccine breakthroughs (according to **b**) are shown as spheres, labeled in one protomer in red (Delta) or pink (AY.25).

### Potential sites of adaptive evolution of the Delta variant under vaccine pressure

To screen for genomic signatures of vaccine breakthrough and possible adaptive selection, we assessed all base pair mutations over the full genome and also all amino acid mutations across spike, enriched in vaccinated or unvaccinated participants (**Figs. 3, 4**, **S5, S6 Tables S4-S6**). The numbers of full genome and spike mutations were comparable in viruses obtained from vaccinated and unvaccinated individuals. Breakthrough sequences had in average 35.8 base pair mutations over the full genome (range: 24-44) and 9.7 amino acid mutations per spike (range: 3-14), compared to 35.6 base pair mutations/full genome (range: 19-56) and 9.6 amino acid mutations/spike (range: 3-15) in the unvaccinated. In spike, we found a total of 168 amino acid changes compared to Wuhan-Hu-1: 115 sites were more frequently mutated in unvaccinated controls, compared to 52 sites in breakthroughs (**Fig. 3a**). To identify spike amino acid mutations that are enriched in breakthrough sequences, we performed a two-pronged approach: I) to investigate the enrichment of mutations across variants, we statistically analyzed sequences from all variants combined with a post-hoc variant assessment to identify potential bias introduced by unequal variant distribution between groups (**Fig. 3b**). II) We performed a Delta-specific mutation analysis to identify mutations enriched among Delta sequences and to confirm the results using all variants (**Fig. 3c**). Among the 52 mutations preferentially found in breakthroughs, 10 significantly differed between breakthrough and unvaccinated cases (**Fig. 3b, Table S4**). The post-hoc analysis revealed that nine of these 10 sites were Delta-defining mutations, whilst one, S112L, is not Delta-defining but contributes to the array of Delta subvariant AY.25 mutations. The comparative spike mutation analysis exclusive forthe subset of Deltasequences confirmed S112L as the only substantially enriched mutation among Delta breakthroughs (P<0.05, **Fig. S5** and **Table S5**). The S112L mutation locates at the surface-exposed top part of NTD, one of the major antigenic regions of the spike protein (**Fig. 3d**)^30^.

**Figure 4.**
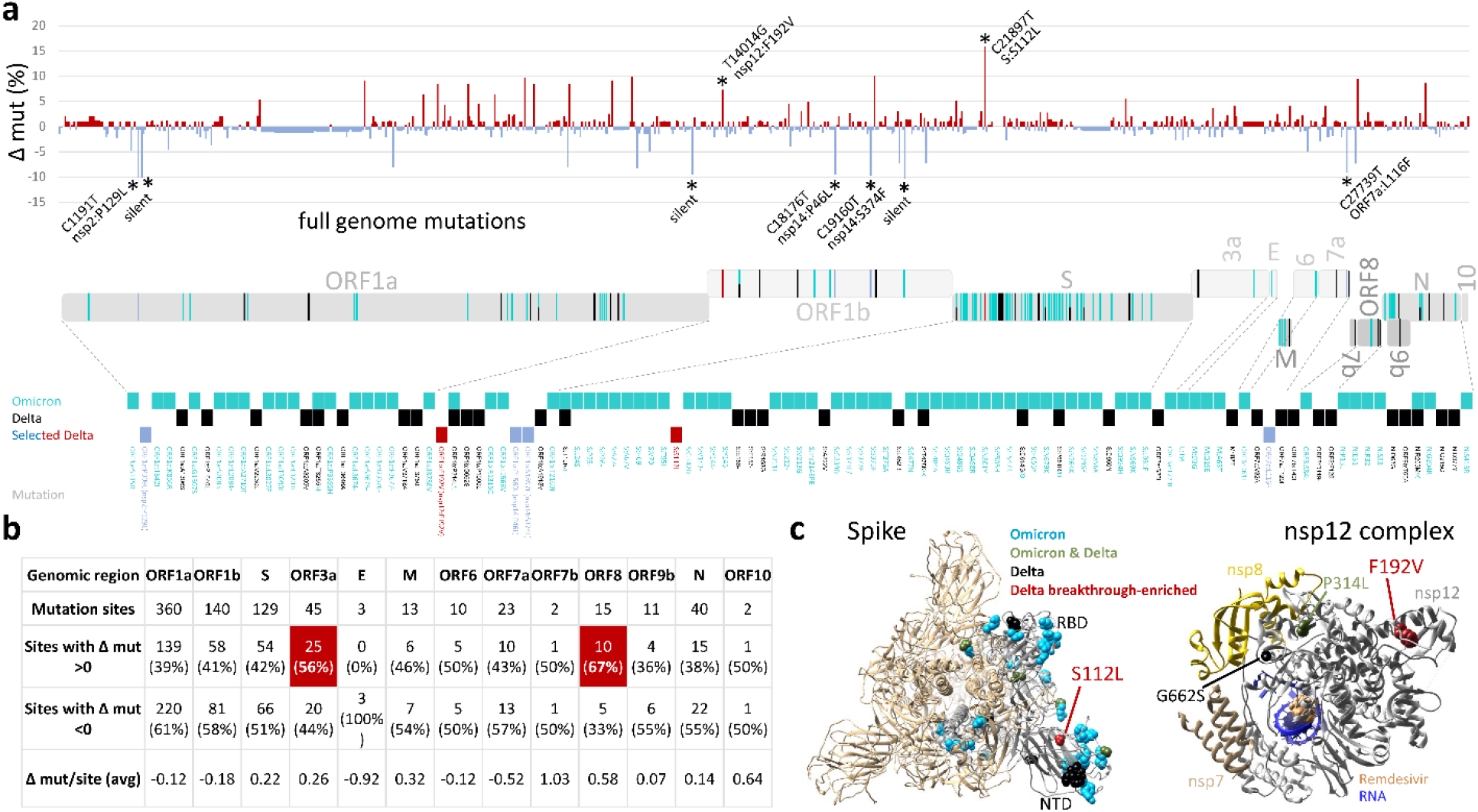
Full genome mutation analysis in SARS-CoV-2 Delta vaccine breakthrough sequences compared to Delta unvaccinated controls and in the context of Omicron mutations. a, Site-specific base pair mutation frequencies in full genomes (bp 202-29666) of 101 Delta vaccine breakthrough sequences compared to 139 Delta unvaccinated controls from the same cohort. The Wuhan-Hu-1 sequence served as reference. The mirror plot displays differences of mutation frequencies per site between vaccinated and unvaccinated groups, shown along the x-axis (n=791); red (facing up) and blue bars (facing down) refer to elevated mutation rates in vaccinated or unvaccinated individuals, respectively. Significantly enriched mutations in Fisher exact tests are indicated by asterisks (* P<0.05) and are labeled. SARS-CoV-2 coding genomic regions are shown below the plot. Non-synonymous mutations in Omicron (cyan; including B.1.1.529, BA.1, and BA.2 mutations), Delta (black), or Delta breakthrough-enriched mutations (red and blue) are shown by colored ticks. The mutation sites/names are indicated below. b, Differential full genome mutation analysis of Delta infections in vaccinated and unvaccinated individuals by SARS-CoV-2 genomic region. Regions in which mutation sites were more frequently mutated in vaccinated compared to unvaccinated individuals are highlighted in orange and percentages shown in bold. c, Structural analysis of Delta breakthrough-enriched mutations in comparison to Omicron- and Delta-defining mutations. Structures are shown for spike in the activated state with one RBD in the up position (pdb: S_ACE2; mutations only shown in the gray, activated protomer) and the nsp12 complex with bound nsp7, nsp8, template-primer RNA, and remdesivir triphosphate (pdb: 7bv2). 3a: ORF3a; 7a: ORF7a; 7b: ORF7b; 9b: ORF9b; 10: ORF10; bp: base pairs; E: envelope; mut: mutation; nsp: non-structural protein; N: nucleocapsid; NTD: N-terminal domain; ORF: open reading frame; RBD: receptor-binding domain; S: spike.

To identify sites of potential adaptive evolution in Delta beyond spike, we extended our statistical analysis to the full genome (**Fig. 4a-c**). Among 240 Delta sequences (101 breakthroughs and 139 unvaccinated controls), we observed 791 mutation sites, of which 448 were more frequently mutated in controls, 328 more frequent in breakthroughs, and 15 equally mutated in both groups. In total, we found nine sites with significantly different mutation rates between groups (**Fig. 4a, Table S6**). Two mutations were enriched in breakthrough sequences compared to unvaccinated controls, which, besides spike mutation S112L included mutation F192V in nsp12, the RNA-dependent RNA polymerase (RdRp) gene (**Fig. 4c**). In addition, seven mutations were less frequent in breakthrough sequences, which included four non-synonymous mutations in nsp2, nsp14 (2x), and ORF7a, and three synonymous mutations in nsp2, nsp9, and nsp15. A genomic region-specific mutation analysis further revealed that most coding regions had higher numbers of mutations in unvaccinated control sequences, whereas ORF3a (56%) and ORF8 (67%) had higher numbers of sites with enriched mutations in breakthrough sequences, though at low mutation rates (**Fig. 4b**).

### Mutations in Omicron and mutations enriched in Delta breakthroughs involve common genomic hot and cold spot regions

Omicron, first detected in October 2021 in Nigeria, has caused a massive SARS-CoV-2 outbreak in South Africa starting November 2021, and eventually reached the US, Europe and many countries worldwide with rising numbers as of December 2021. Omicron supposedly evolved in an immunocompromised host and acquired a broad set of mutations associated with immune escape ^4–6, 31^. Thus, both Omicron and Delta breakthrough viruses were presumably selected against immune selection pressure, which prompted us to perform a comparative analysis of their full-genome mutational landscapes. Omicron is characterized by more than 30 mutations in spike, complemented by smaller mutation hotspot regions in ORF1ab, M, and N genes (**Fig. 4a**). One of the two mutations that we found positively associated with Delta breakthrough (spike S112L) is situated in the middle of the spike NTD domain, an Omicron hotspot region that carries abundant mutations and deletions (**Fig. 4a,c**). The second breakthrough-enriched mutation in Delta (nsp12 F192V) is located in the functionally relevant nsp12 domain, coding for the RdRp polymerase, which also entails the Omicron- and/or Delta-defining mutations P314L and G662S (**Fig. 4a,c**). In contrast, the seven sites that we identified as being inversely correlated with Delta breakthrough fell into mutation cold spots of Omicron. The inverse correlations suggest that these Delta mutations had no benefit in conferring breakthrough, but instead, the wild-type residues (as found in Wuhan-Hu-1) were associated with breakthrough and possibly immune escape.

### Delta cases increased at comparable rates in vaccinated and unvaccinated individuals and Delta breakthrough is associated with a distinct set of clinical and genomic factors

We expanded our current analysis of recorded breakthrough infections in our cohort, by adding data from our previous study^17^ that covered February to April 2021, for a total of 208 breakthrough and 1329 control sequences (**Fig. 5**). The pattern of total COVID-19 case numbers in New York State overtime is comparable to our SARS-CoV-2 sequences, supporting our attempt to sequence a representative set of viruses (**Fig. 5a, b**). While increasing vaccination rates were associated with an overall decline in COVID-19 cases and SARS-CoV-2 sequences obtained, emerging Delta infections partially reverted this trend (**Fig. 5a-c**). Iota breakthrough significantly decreased and Alpha breakthrough exhibited no significant change over time since vaccination, whereas Delta breakthrough infections rose significantly in near linear fashion within the first six months post full vaccination, both in absolute and relative counts (**Fig. 5d, S7-S9**).

**Figure 5.**
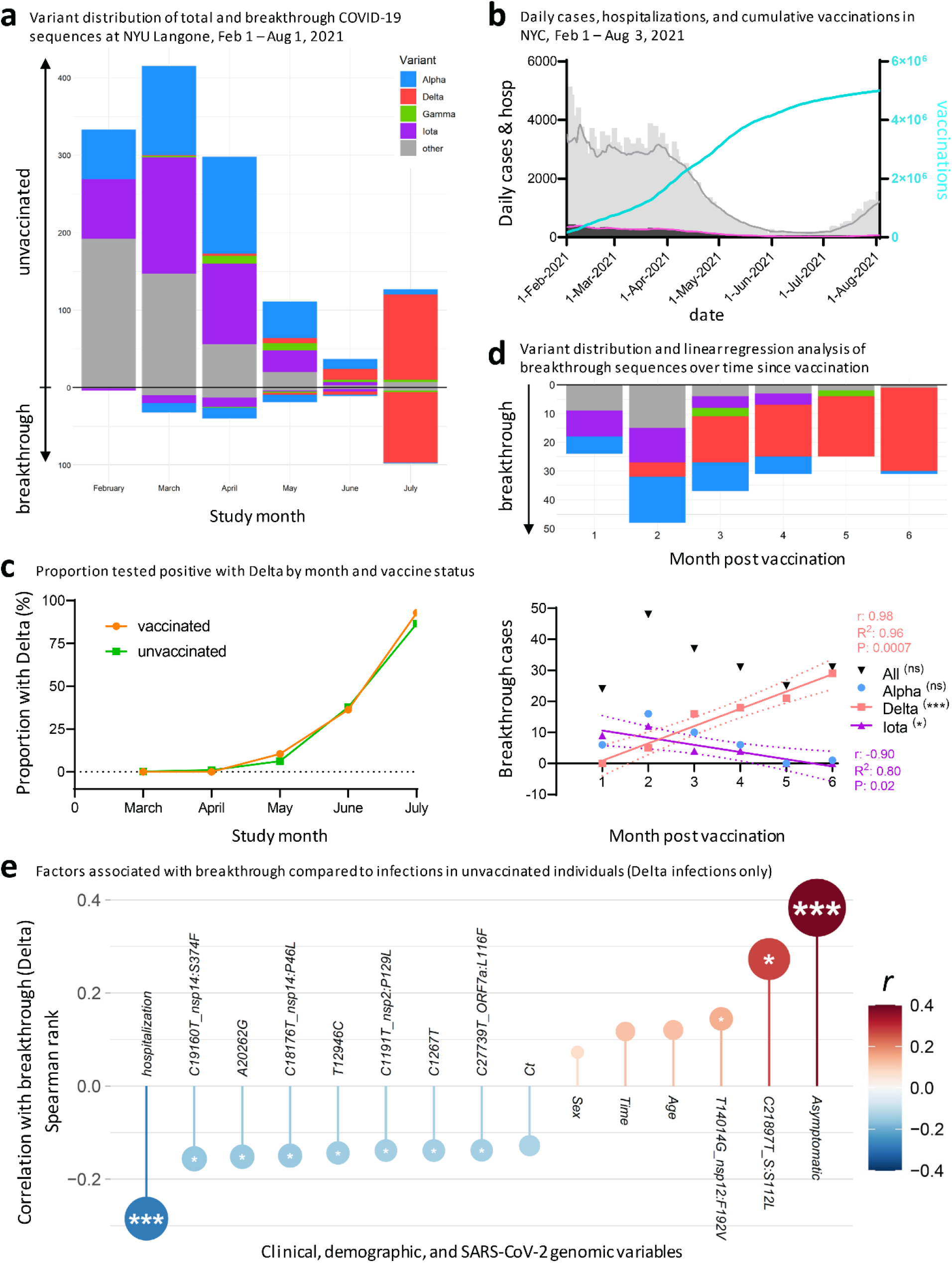
post vaccination, and clinical and genomic factors associated with Delta breakthrough. **a**, Variant distribution of SARS-CoV-2 sequences from unvaccinated (facing up) and vaccinated individuals (facing down), obtained at NYU Langone Health between February and July 2021. **b**, Daily COVID-19 cases (gray bars) and 7-day averages (gray line), daily COVID-19 deaths (black bars) and 7-day averages (pink line), and cumulative vaccination numbers (turquoise line) in New York City between February 1^st^ and August 3^rd^. Source of data: NYC Open Data and NYC Health, Citywide Immunization Registry (CIR). **c**, Probability of positive test with Delta by month in vaccinated and unvaccinated individuals, adjusted for month of test, sex, and age of participants. **d**, Variant distribution among all NYULH breakthrough sequences, displayed according to months post full vaccination (starting at day 14 after the last dose for full vaccination). The chart below shows a linear regression analysis of breakthrough infections per variant against time post vaccination. Significant results are highlighted by asterisks, labeled with the correlation coefficient (r), goodness of fit (R^2^), and P value, and the fitted line with 95% confidence intervals shown. * P<0.05, ** P<0.01, *** P<0.005. **e**, Correlation of clinical, demographic, and SARS-CoV-2 genomic factors with breakthrough by comparing Delta infections in vaccinated (n=101) and unvaccinated individuals (n=139). Spearman rank correlation is displayed on the y axis and color-coded. Multiple comparison-corrected P values (q, Benjamini-Hochberg) are indicated by asterisks within the circles (* q<0.05, ** q<0.01, *** q<0.005). Ct: Cycle threshold in RT-PCR; sex: male sex; time: date of sampling.

To assess whether Delta infections are statistically enriched among breakthrough infections compared to other variants, we performed logistic regression analyses and matched the 208 vaccinated breakthrough patients 1:1 to the 1329 unvaccinated patients, correcting for the confounding variables clinical collection date, sex, and age. While unmatched analyses suggested elevated numbers of Delta infections in vaccinated compared to unvaccinated individuals, the adjusted analysis in matched participants of the study cohort revealed no significant difference between both groups (**Fig. S8, Tables S7, S8**). Recorded Delta cases increased significantly compared to all other variants starting in May 2021 (**Table S8**). Notably, the rise of Delta occurred at comparable rates in vaccinated in unvaccinated individuals (**Fig. 5c**), indicating no major advantage of Delta *per se* in conferring breakthrough after adjustment for confounding variables.

In summary, the study of Delta infections revealed that, compared to unvaccinated controls, breakthrough infections were associated with asymptomatic disease, lower rates of hospitalization, spike mutation S112L, nsp12 mutation F192V, and the absence of seven mutations across different regions of the SARS-CoV-2 full genome (**Fig. 5e**).

## Discussion

Since emergency-use authorization of the first COVID-19 vaccine in December 2020, the spotlight has been on viral sequences from vaccinated individuals to closely monitor the potential emergence of vaccine escape mutations. While the first half of 2021 was characterized by a wealth of variants, some more prevalent than others regionally or globally, the Delta variant has rapidly replaced all other variants worldwide^32^. Our cohort in metropolitan New York is a prime example of Delta’s takeover, both in unvaccinated and vaccinated individuals with a near linear increase of Delta breakthrough infections according to elapsed time post vaccination (**Figs. 1, 5**). Breakthrough numbers in our cohort approach the absolute numbers in the unvaccinated, which is in line with other reports^33, 34^. The reasons for this change are multifactorial, including epidemiological and virological factors. First, the probability of a positive case being a breakthrough versus an unvaccinated case has risen over time due to increased vaccination rates in our area and elsewhere (**Fig. 5**)^35^. Second, Delta has been detected in increased rates in younger hosts, who tend to be vaccinated at lower rates than older individuals, or were ineligible for vaccination at the time of our study. The younger population is also associated with lower hospitalization rates (**Figs. 1, S1**)^36^. Third, Delta carries the P681R substitution in the spike furin cleavage site, which triggers enhanced S1/S2 cleavage and might explain Delta’s increased replication rate *in vitro* ^24, 37, 38^. A substitution at the same site (P681H) is found in Alpha, which evolved independently of Delta and dominated the global infection landscape between January and June 2021^39–43^. P681H/R mutations were both detected more frequently among breakthrough infections in our previous study of breakthrough infections in New York when Alpha and Iota variants were dominant regionally^17^, and in this study during the current Delta wave (**Fig. 3**). Although Delta shows compromised sensitivity to some RBD and NTD neutralizing antibodies with up to 8-fold reduced sensitivity *in vitro* to vaccine-induced antibodies compared to D614G viruses (including infectious virus assays)^24, 26, 44^, neutralization escape is substantially lower in magnitude as compared to Beta, Gamma, and Mu^45, 46^. Efficient spike cleavage (P681H/R) and replicative competence appear to be central checkpoints for SARS-CoV-2’s epidemiological success, with Delta being improved compared to Alpha^24, 37, 47–49^.

In line with data from other laboratories, the pre-Delta breakthrough infections that we studied in early 2021 displayed a variant distribution similar to infections in unvaccinated individuals^17–20^, though with signs of a starting sieve effect of neutralizing antibody escape mutations, e.g., E484K^15–17, 50^. The variant pattern in both breakthrough and unvaccinated cases has dramatically changed towards the second half of 2021 with a clear dominance of Delta-associated mutations (**Fig. 3**). In the second half of 2021, breakthrough appears to be primarily shaped by virological factors that increased transmissibility, facilitated by immunological permeability in times of waning vaccine efficacy^51^. Delta’s global rise in the second half of 2021 coincided when large parts of high-risk populations were past six months post full vaccination. The lift of US mask recommendations in May 2021, right before the ignition of the Delta wave, may have also added to its spread. Mask mandates were reinstated halfway through the Delta wave, i.e., on July 28^th^ 2021, at NYU Langone Health, whereas New York State only issued a recommendation to wear masks in early August^52^.

At a time where it is not known whether Omicron will become the predominant variant globally, Delta remains a primary source for the evolution of next generation variants, evidenced by the rapid diversification into more than 190 Delta subvariants. We observed an uptick of the subvariant AY.25 and the S112L mutation in NTD, which, while still at low numbers, preferentially spread among the vaccinated individuals in our cohort. In our phylogenetic analysis, we observed multiple Delta clusters that suggest efficient spread of distinct sublineages in the population. As shown for AY.25, they involve unvaccinated and vaccinated individuals, and their presence in clusters implies transmission chains.

High vaccine efficacy was reported against Delta infections, particularly at the beginning of the Delta wave; however, there is growing evidence that vaccine efficacy against Delta decreases over time^23, 25, 53–55^. A central question remains whether reduced vaccine efficiency is due to Delta *per se* or whether it is a matter of waning immunity and dependent on elapsed time since full vaccination. Current knowledge indicates that it may be both. Statistical analyses of retrospective cohort studies indicated that waning immunity with time, coinciding with a period of easing societal public health restrictions, played a larger role than Delta’s vaccine escape^54, 55^. However, an observational study on SARS-Cov-2-infected index cases and contacts indicated that vaccination reduced transmission more effectively in Alpha compared to Delta^25^. Our data suggest that selective adaptation processes might be in process that may eventually lead to the accumulation of mutations (spike S112L, nsp12 F192V) or new (sub)variants (AY.25) under vaccine immune pressure (**Figs. 4** and **5e**).

Of note, when we compared the selectively enriched Delta breakthrough mutations with the ones present in Omicron, a virus, which has presumably overcome immune selective pressure during its formation ^4–6, 31^, we found certain commonalities. Delta breakthrough-enriched mutations and Omicron mutations involve the NTD of spike and nsp12, whereas sites that inversely correlated with Delta breakthrough were found in cold spot regions in terms of Omicron mutations. The latter suggest, that mutations in these cold spot regions have no benefit in conferring breakthrough, whereas the wild-type residues (Wuhan-Hu-1) might be superior in conferring breakthrough and/or immune escape. Although none of the breakthrough-associated Delta mutations fell onto Omicron’s clade-defining mutations, both share comparable hot and cold spot regions, indicating common features of selective adaptation with fine specificities differing by variant, particularly for Delta and Omicron originating from different branches of the phylogenetic tree. It remains elusive at this time whether spike: S112L, located at the apical part of spike NTD, nsp12: F192V, located distant from nsp12’s active site, or the absence of mutations in the cold spot regions play an active role in breakthrough, immune evasion, enhanced transmission, or act as bystander mutations.

As with all observational genomic surveillance studies, limitations and confounding factors exist. These include demographic, temporal, and behavioral factors, diagnostic testing practices, sequencing sensitivity thresholds, and sampling, e.g., undersampling of mild or asymptomatic cases. Some but not all of these factors can be adjusted or minimized by unbiased sample collection, consistent testing procedures and guidelines, and matched data analyses. Our adjusted model indicates that the probability of Delta infection has increased at similar rates in vaccinated and unvaccinated individuals. However, Delta breakthrough cases appear to rise continuously with extended time post vaccination (**Fig. 5**), which justifies the use of booster doses ^56, 57^. Monitoring Delta, its evolving subvariants, and newly emerging variants including their early signs of adaptive evolution will be critical to adequately address upcoming SARS-CoV-2 outbreaks and the increasing numbers of vaccine breakthrough infections.

## Methods

### Study design and sample collection

This study was approved by the NYULH Institutional Review Board, protocol numbers i21-00493 and i21-00561. SARS-CoV-2 infections were studied in vaccinated and unvaccinated individuals from May 1^st^ until August 3^rd^ in the NYULH system. Cases were identified using DataCore, the system’s clinical data management and extraction resource. Vaccine breakthroughs were defined by positive real-time (RT)-PCR test for SARS-CoV-2 RNA regardless of Ct at least 14 days after the second dose of BNT162b2 (Pfizer/BioNTech) or mRNA-1273 (Moderna) vaccines, or the single-dose COVID-19 Janssen vaccine. The unvaccinated control group consisted of all SARS-CoV-2 positive cases in our healthcare system who had not received a dose of any vaccine at the time of RT-PCR positivity, collected and sequenced in the same period as the breakthrough infections. Nasopharyngeal swabs were sampled from individuals with exposure to SARS-CoV2, suspected to have an infection with SARS-CoV-2, or as part of clinical diagnostics or hospital admission.

### RNA extraction, library preparation and sequencing

Detailed methods were described recently^17^ (**Supplemental Methods**). SARS-CoV-2 sequences that were <23,000 bp or <4000x genome coverage were considered inadequate and not included in the analyses.

### Phylogenetic and mutation analyses

Methods were described recently^17^ (**Supplemental Methods**). Briefly, maximum likelihood IQ trees were produced on Mafft-aligned SARS-CoV-2 full genome sequences (1000 bootstrap replicates). Spike amino acid counts and site-specific mutation frequencies were analyzed using Fisher exact tests with multiplicity corrections (Benjamini-Hochberg)^58^.

### Statistical analysis

The comparison of 132 breakthrough and 283 unvaccinated control samples achieved 96% power in detecting a 15% difference (10% versus 25%) in mutation rates, clinical or demographic variables in a two-tailed chi squared/Fisher Exact test with a type I error of 5% (G*Power v.3.1.9.4)^59^. We evaluated the variant distribution (Alpha, Delta, Gamma, Iota, and all other variants) in fully vaccinated compared to unvaccinated individuals and addressed confounding variables arising from the use of observational data via matching and adjustment. We matched the vaccinated breakthrough cases 1:1 to the unvaccinated cases, correcting for the confounding variables: clinical collection date, sex, and age^60^. Quality controls of matching included the analysis of propensity score distributions and empirical quantile-quantile (eQQ) plots of distribution balance of co-variates using the R v.4.1.0 MatchIt package. To compare the probability of a Delta variant for the vaccinated and unvaccinated groups over time, we performed logistic regression analyses on the full data adjusting for sex, age (centered and standardized), and month of test. The relationship between variant distribution and time since vaccination was studied using linear regression analyses in Prism v.8.4.3 using two-sided Pearson tests. We compared clinical and demographic variables between groups using non-parametric Mann-Whitney tests or Kruskal-Wallis tests with Dunn’s multiplicity correction in Prism. Correlation analyses were done using two-sided Spearman rank tests in Prism. For all analyses unless otherwise stated, a two-sided Type I error rate of 0.05 was applied. Details are described in **Supplemental Methods**.

## Data availability

All sequences are publicly available in SRA (BioProject PRJNA769411) and GISAID (**Tables S1-S3**).

## Author contributions

Ralf Duerr, MD, PhD designed the study, analyzed data, created figures, and wrote the manuscript; Dacia Dimartino, PhD and Paul Zappile made libraries and sequenced the samples; Christian Marier analyzed genomic data; Samuel Levine and Fritz Francois, MD, generated total numbers of vaccinations for the entire healthcare system; Guiqing Wang, MD, PhD collected samples and performed clinical SARS-CoV-2 detection; Eduardo Iturrate, MD, and Brian Elbel, PhD monitored epidemiological data and generated lists of breakthrough infections; Meike Dittmann, PhD wrote the manuscript; Jennifer Lighter, MD reviewed clinical information; Andrea Troxel, ScD and Keith S. Goldfeld, DrPH performed statistical analyses; and Adriana Heguy, PhD designed the study, generated genomic data and wrote the manuscript. All authors reviewed and edited the manuscript.

## Supplemental Methods

RNA extraction, cDNA synthesis, library preparation and sequencing

Phylogenetic and mutation analyses

Visualization

Statistical analysis

External data sources

## Supplemental Figures

**Figure S1.** Hospitalizations and demographicdata by variant in breakthrough cases and unvaccinated controls.

**Figure S2.** Asymptomatic infections and demographic data by variant in breakthrough cases and unvaccinated controls.

**Figure S3.** Relationship of clinical, demographic, and genomic data in breakthrough cases and unvaccinated controls.

**Figure S4.** Phylogenetic analysis of SARS-CoV-2 Delta sequences.

**Figure S5.** Spike mutation patterns in vaccine breakthrough sequences.

**Figure S6.** Site-specific spike mutation analysis in SARS-CoV-2 Delta vaccine breakthrough sequences compared to Delta infections in unvaccinated controls.

**Figure S7.** Ratios of breakthrough SARS-CoV-2 infections by variant and time post vaccination

**Figure S8.** Breakthrough SARS-CoV-2 infections by variant and time post vaccination.

**Figure S9.** SARS-CoV-2 variant distribution in vaccinate breakthrough and unvaccinated cases and quality control of matching.

## Supplemental Tables

**Table S1.** Full specifications of 132 SARS-CoV-2 breakthrough infections.

**Table S2.** Full specifications of 283 SARS-CoV-2 infections in unvaccinated controls.

**Table S3.** Metadata of GISAID reference sequences used in phylogeneticanalyses.

**Table S4.** Spike mutation statistics of sites with enriched mutation rates in vaccine breakthrough infections compared to unvaccinated controls.

**Table S5.** Delta variant spike mutation statistics of sites with enriched mutation rates in Delta vaccine breakthrough infections compared to Delta infections in unvaccinated controls.

**Table S6.** Delta variant full genome mutation statistics of all sites with significantly different mutation rates in Delt avaccine breakthrough infections compared to Delta infections in unvaccinated controls.

**Table S7.** Contingency tables and chi-square tests of unmatched and 1:1 matched data from vaccinated and unvaccinated SARS-CoV-2^+^ participants.

**Table S8.** Estimated model comparing the probability of a positive Delta test for the vaccinated/unvaccinated groups, adjusted for sex, age (centered and standardized), and month of test.

## Data Availability

All sequences are publicly available in SRA (BioProject PRJNA769411) and GISAID (Tables S1-S3).

## Acknowledgements

We thank NYU Langone Health DataCore for support extracting the data for this study from clinical databases and the clinical laboratory technicians for assistance in testing, saving and retrieving specimens, especially Joanna Fung. We also thank Dr. Joan Cangiarellafor her continuous support of genomic surveillance for SARS-CoV-2 at NYULH, including providing institutional funding forthis study. The authors wish to acknowledge the software support of New York University’s Data Services, Bobst Library, and, in particular, the expertise shared by Christopher Schwarz and Senior Academic Technology Specialist Denis Rubin. We are grateful to the submitting laboratories who deposited data in GISAID, in particular to those whose sequences we used to create the phylogenetictree (**Table S3**). The NYULH Genome Technology Centeris partially supported by the Cancer Center Support Grant P30CA016087 at the Laura and Isaac Perlmutter Cancer Center.

