## Supplemental information for "Clinical and genomic signatures of rising SARS-CoV-2 Delta breakthrough infections in New York"

Submitted to Nature Medicine (2021)

### Supplemental Methods

**RNA extraction, cDNA synthesis, library preparation and sequencing**

Clinical testing was performed using various FDA emergency use authorization (EUA) assays for detection of SARS-CoV-2 RNA, i.e., the Roche Cobas 6800 SARS-CoV-2 (90% of the samples in this study), Cepheid Xpert SARS-CoV-2, or SARS-CoV-2/Flu/RSV assays.

RNA was extracted from 400 µl of each nasopharyngeal swab specimens using the MagMAX™ Viral/Pathogen Nucleic Acid Isolation Kit on the KingFisher flex system (Thermo Fisher Scientific). Eleven µl of RNA were used for first strand cDNA synthesis using the Superscript IV first-strand synthesis kit (Invitrogen, ref# 180901050). Libraries were prepared using Swift Normalase Amplicon SARS-CoV-2 Panel (SNAP) and SARS-CoV-2 additional Genome Coverage Panel (Cat# SN-5X296 core kit, 96rxn), using 10 µl of first strand cDNA, with 24 PCR cycles[1]. Libraries were pooled and run on the Illumina NovaSeq 6000 system on SP 300 cycle flow cells, as paired-end 150 cycles with dual indexing reads. Typically, two pools representing two full 96 well plates (192 samples) were sequenced on each SP300 NovaSeq flow cell. Reads were demultiplexed using the Illumina bcl2fastq2 Conversion Software v2.20, and adapters and low-quality bases were trimmed with Trimmomatic v0.36[2]. BWA v0.7.17[3] was utilized for mapping reads to the SARS-CoV-2 reference genome (NC_045512.2, wuhCor1). SNAP tiled primer sequences were removed with Primerclip v0.3.8[4]. BCFtools v1.9[5] was used to call mutations and assemble consensus sequences, which were then assigned phylogenetic lineage designations according to PANGO nomenclature, version 2021-09-28[6].

**Phylogenetic and mutation analyses**

SARS-CoV-2 full genome sequences were aligned using Mafft v.7[7]. The alignment was cropped to base pairs 202-29,666 according to the Wuhan-Hu-1 reference to remove N- and C-terminal regions with unassigned base pairs. Maximum likelihood IQ trees were performed using the IQ-TREE XSEDE tool, multicore version 2.1.2, on the Cipres Science gateway v.3.3[8]. GTR+F+I+G4 was chosen as the best-fit substitution model according to the Bayesian Information Criterion (BIC) as determined by Modelfinder[8]. Support values were generated with 1000 bootstrap replicates and the ultrafast bootstrapping method, and the oldest SARS-CoV-2 sequence (Wuhan/WH01/EPI_ISL_406798/2019-12-26) was set as the root of the tree. The tree was constructed using 132 vaccine breakthrough and 283 unvaccinated control SARS-CoV-2 sequences from our NYU cohort (greater NYC area) together with 920 US reference sequences (non-NYU) and 2176 global reference sequences (non-US) for a total of 3511 SARS-CoV-2 genomic sequences. The reference sequences were retrieved from a North America-focused Nextstrain build with global subsampling[9,10]. Phylogenetic trees were visualized in Interactive Tree Of Life (iTOL) v.6[11].

Mutations compared to Wuhan-Hu-1 as reference were determined on MAFFT-aligned SARS-CoV-2 sequences in program R v.4.1.0[12] and R Studio v.1.4.1106[13] using scripts based on the seqinr and tidyverse packages. The statistical analyses considered residues that were covered by sequencing, i.e., non-ambiguous characters and gaps, whereas ambiguous and undefined characters were excluded. Fisher exact tests and multiplicity corrections (Benjamini-Hochberg)[14] were done in Program R. Multiplicity corrected P values (q) $<0.05$ were considered significant.

**Visualization**
Mutation data tables, line charts, mirror bar graphs, and lollipop plots were created in Microsoft Excel 2016 and/or program R (ggplot2 package). Individual spike mutations of MAFFT-aligned SARS-CoV-2 vaccine breakthrough sequences in comparison to Wuhan-Hu-1 as master were visualized using the Highlighter tool provided by the Los Alamos HIV sequence database [15], focusing on the translated spike region and unambiguously defined amino acids and gaps. Circular edge bundling plots were generated in undirected mode in R using ggraph, igraph, tidyverse, and RColorBrewer packages. Edges are only shown if $P<0.05$, and nodes are sized according to the connecting edges' r values. Nodes are color-coded according to groups of variables. Stacked bar graphs and bubble plots were created using the ggplot2 package in R; box plots were made in Prism. Donut plots were created in Excel and stream graphs using RAWGraphs v.2.0[16]. Structural analyses were performed using Chimera v.1.15rc[17] and the structural models of SARS-CoV-2 spike with human ACE2, pdb S_ACE2[18] and SARS-CoV-2 polymerase RdRp (nsp12) in complex with nsp7, nsp8, template-primer RNA, and Remdesivir triphosphate, pdb 7bv2[19].

**Statistical analysis**
The comparison of 132 breakthrough and 283 unvaccinated control samples achieved 96% power in detecting a 15% difference (10% versus 25%) of a study variable, e.g., genotype or mutation rate, in a two-tailed chi squared/Fisher Exact test with an error of 5% (G*Power v.3.1.9.4)[20]. We evaluated the variant distribution in fully vaccinated compared to unvaccinated individuals, as follows. To address confounding and other sources of bias arising from the use of observational data, we performed matching and adjustment in R. We estimated a propensity score for the likelihood of full vaccination, and matched vaccinated to unvaccinated patients 1:1, including clinical collection date, age, and sex as covariates[21]. Propensity-score matching was implemented using the nearest neighbor strategy with the MatchIt algorithm in program R[22]. Matched analyses were primarily done for the accumulated data set from the moment when breakthrough cases were recorded in our cohort, i.e., in February 2021, by aggregating data from our previous (76 breakthrough and 1046 control sequences)[23] and current study for a total of 208 breakthrough and 1329 control sequences. Quality controls of matching included the analysis of propensity score distributions and empirical quantile-quantile (eQQ) plots of distribution balance of co-variates using the MatchIt package in program R. All breakthrough cases were successfully matched, reducing the standardized mean difference between the matched patients from 0.95 to 0.01. Before and after matching, we evaluated the presence of Alpha (B.1.1.7), Delta (B.1.617.2 or AY.*; * can be any number), Gamma (P.1), Iota (B.1.526), and all other variants. To compare the probability of a Delta variant for the vaccinated and unvaccinated groups over time (by study month), we performed logistic regression analyses on the full data (208 versus 1329 sequences) adjusting for sex, age (centered and standardized), and month of test. Linear regression analysis of variant distribution over time since vaccination was done in Prism v.8.4.3 using two-sided Pearson tests. Statistical comparisons of clinical and demographic variables between groups were made using non-parametric Mann-Whitney tests or Kruskal-Wallis tests with Dunn's multiplicity correction in Prism. Correlation analyses were done using two-sided Spearman rank tests in Prism and $P<0.05$ was considered significant.

**External data sources**

New York City-wide, daily COVID-19 case numbers and hospitalizations were obtained from NYC Open Data with data provided by the Department of Health and Mental Hygiene (DOHMH)[24] and NYC Health with data provided by the Citywide Immunization Registry (CIR)[25].

### Supplemental Figures

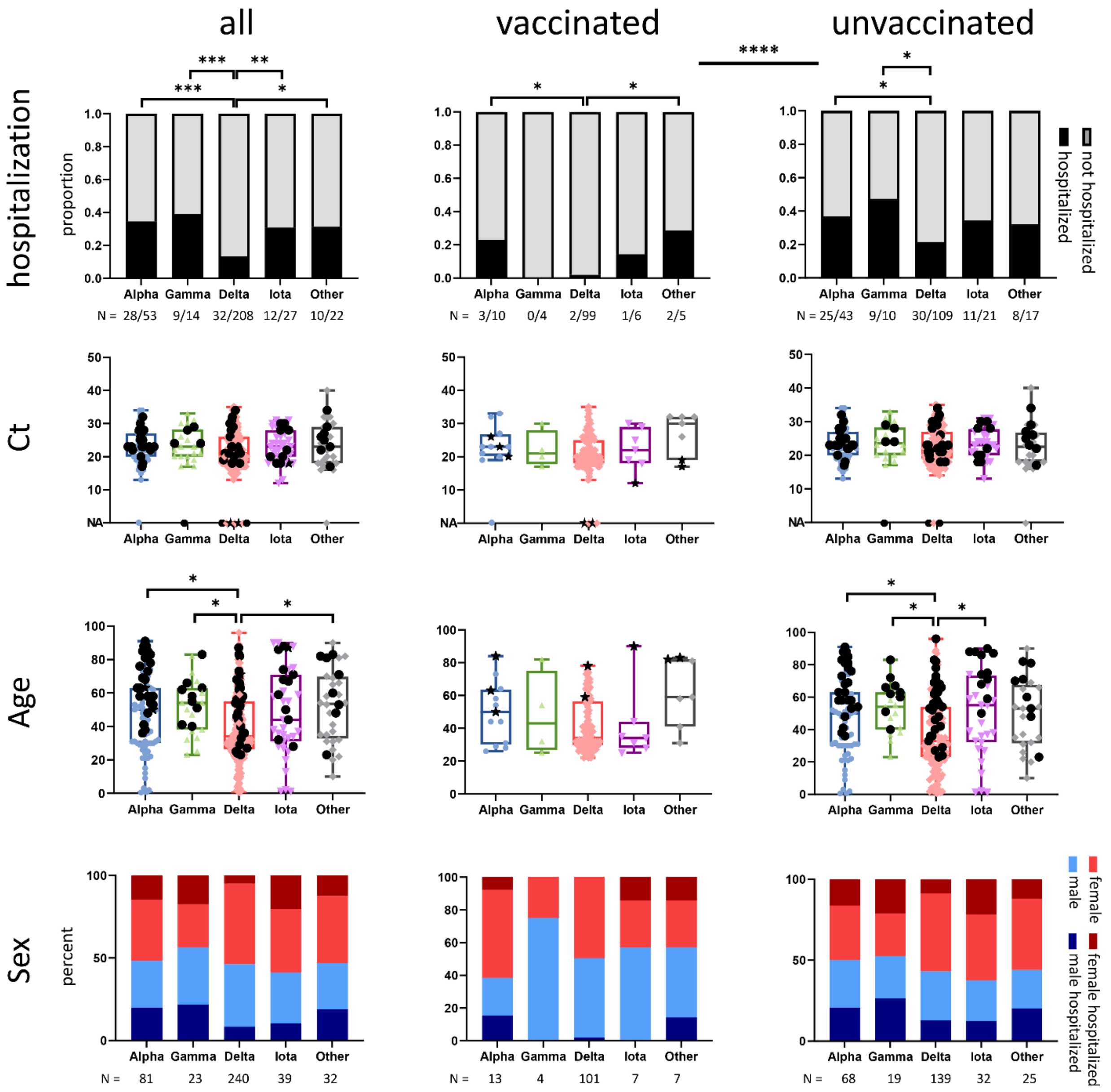

**Figure S1. Hospitalizations and demographic data by variant in breakthrough cases and unvaccinated controls.**

Bar graphs and box plots summarizing hospitalizations and different demographic variables by variant. Top row: hospitalization rates were compared using Fisher exact tests. Numbers (N) indicate hospitalizations versus non-hospitalized cases per variant. Middle two rows: hospitalized cases are shown in black circles (unvaccinated) or stars (vaccinated). Whiskers indicate the range, boxes the interquartile range, and horizontal line the median of values. Statistical analysis was done using Kruskal-Wallis test. Bottom row: sex distribution per variant where darker colors indicate hospitalizations. Total case numbers per variant are listed below (N). * P<0.05, ** P<0.01, *** P<0.005. Ct: Cycle threshold in RT-PCR; NA: not available.

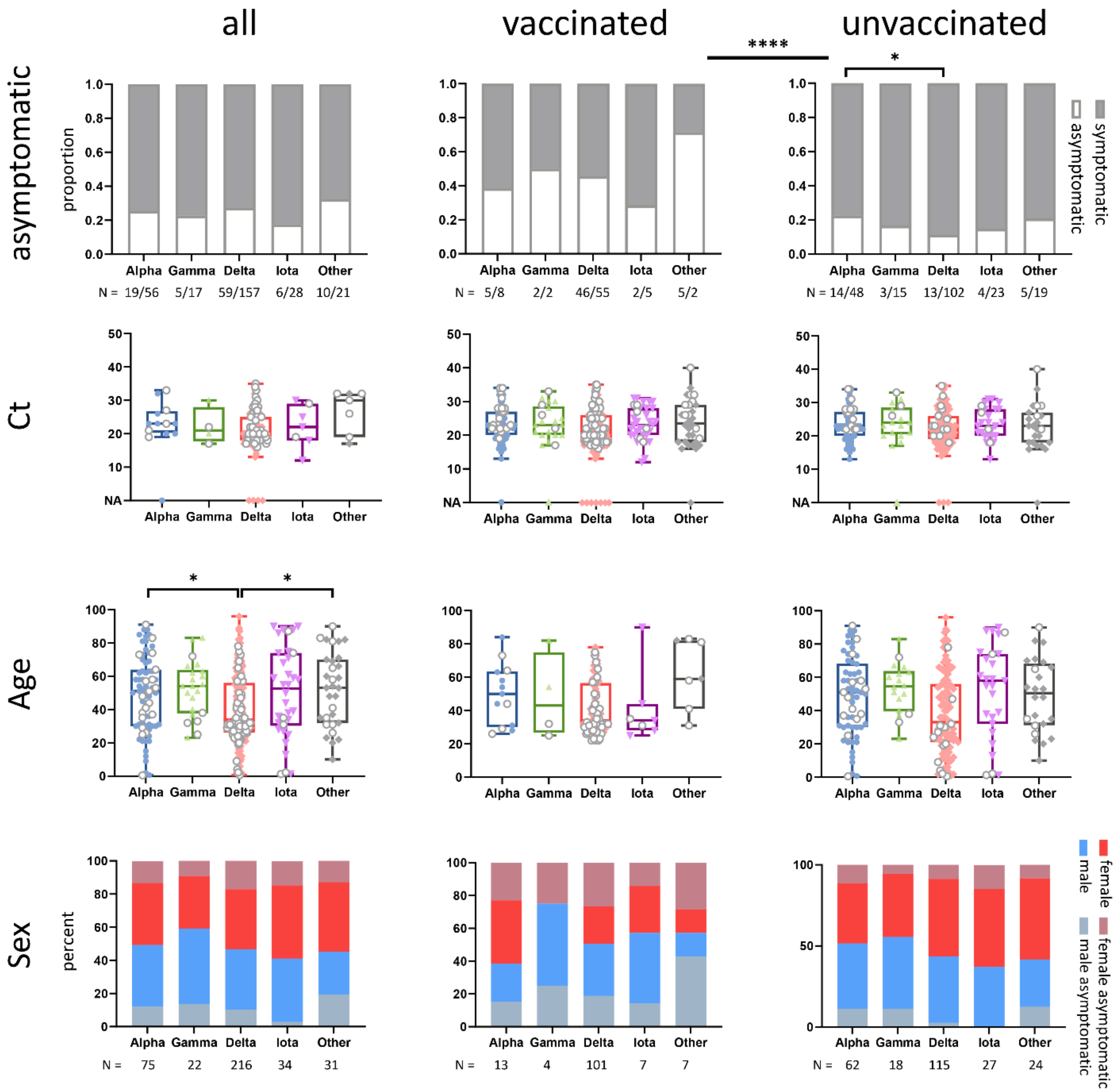

**Figure S2. Asymptomatic infections and demographic data by variant in breakthrough cases and unvaccinated controls.**

Bar graphs and box plots summarizing asymptomatic/symptomatic data and different demographic variables by variant. Top row: asymptomatic rates were compared using Fisher exact tests. Numbers (N) indicate asymptomatic versus symptomatic cases per variant. Middle two rows: asymptomatic cases are shown in gray circles with white fill. Whiskers indicate the range, boxes the interquartile range, and horizontal line the median of values. Statistical analysis was done using Kruskal-Wallis test. Bottom row: sex distribution per variant where red-gray and blue-gray colors indicate asymptomatic cases. Total case numbers per variant are listed below (N). * P<0.05, ** P<0.01, *** P<0.005. Ct: Cycle threshold in RT-PCR; NA: not available.

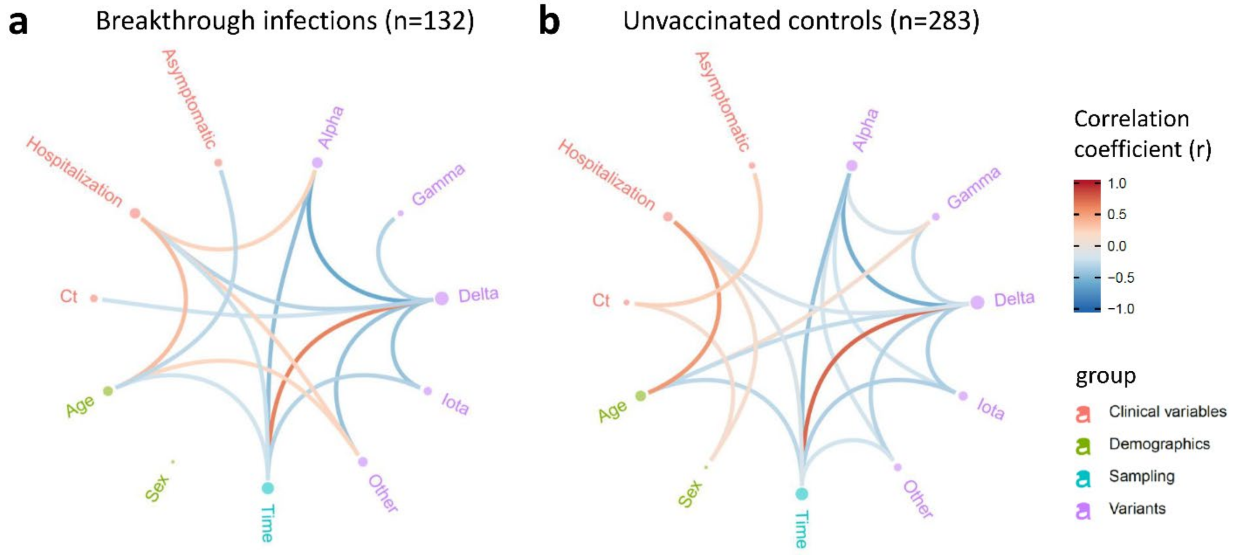

**Figure S3. Relationship of clinical, demographic, and genomic data in breakthrough cases and unvaccinated controls.**

Correlation analysis of clinical, demographic, and genomic data, shown separately for the 132 vaccinated (**a**) and 283 unvaccinated (**b**) SARS-CoV-2-positive individuals. Red and blue edges represent positive and negative correlations between connected variables, respectively, according to the scale of r values to the right. Only significant correlations (P<0.05, Spearman rank test) are displayed. Nodes are color-coded based on the grouping of variables. Node size corresponds to the strength of correlations. Ct: Cycle threshold in RT-PCR; **s**ex: male sex; time: date of sampling.

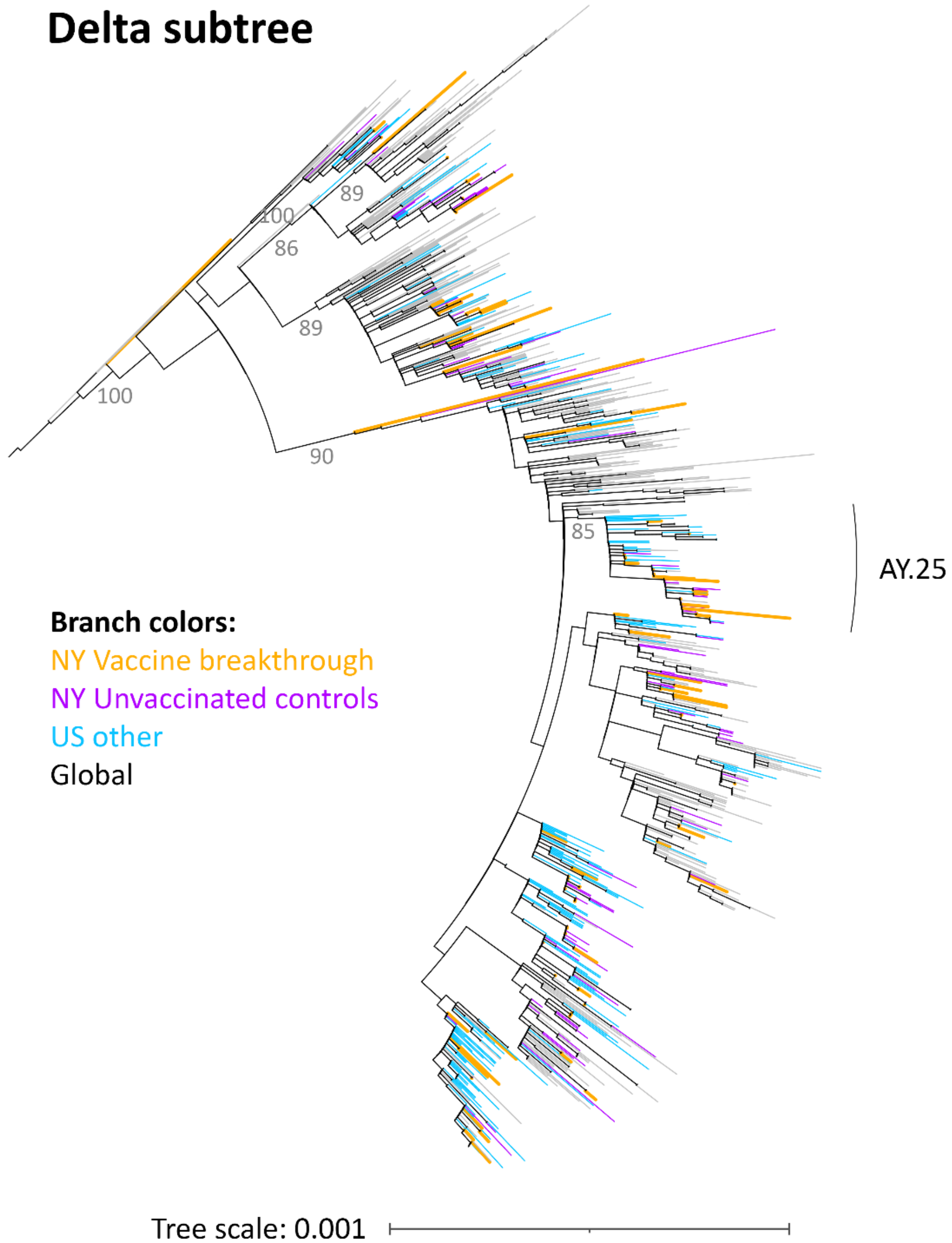

**Figure S4. Phylogenetic analysis of SARS-CoV-2 Delta sequences.**
Maximum likelihood (IQ) tree of 943 SARS-CoV-2 full genome sequences (base pairs 202-29,666 according to Wuhan-Hu-1 as reference), including 101 vaccine breakthrough (orange) and 139 unvaccinated control SARS-CoV-2 sequences from the NYU Langone Health cohort (greater NYC area) (purple) together with 700 other US (non-NYU; cyan) and global (non-US; black) Delta reference sequences and three Wuhan sequences from the beginning of the pandemic. The substitution scale of the tree, generated with 1000 bootstrap replicates and Wuhan/WH01/2019-12-26 as root, is indicated at the bottom right. Selected bootstrap values ≥85 of major branches are indicated. Sequences belonging to the Delta subvariant AY.25 are labeled.

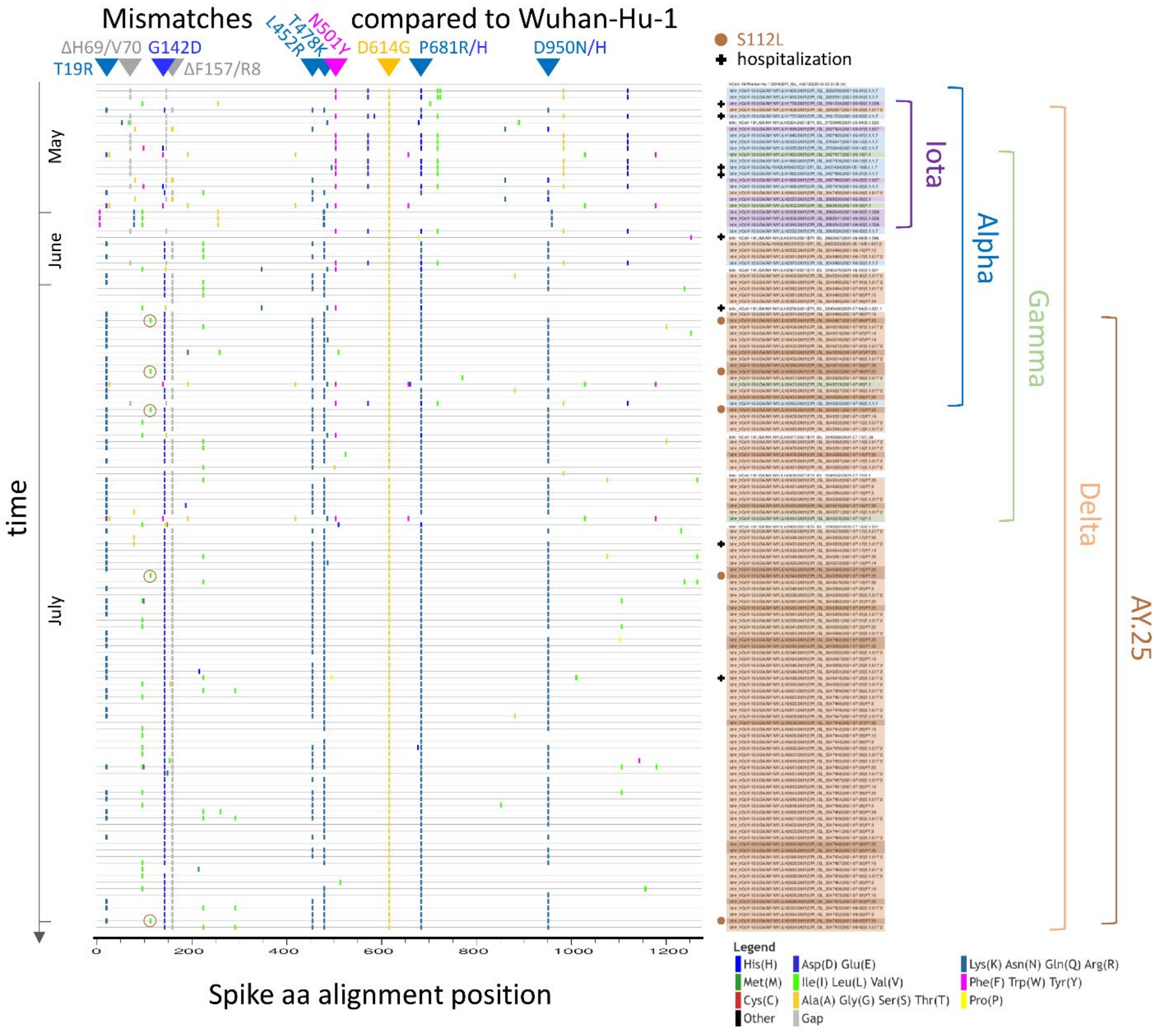

**Figure S5. Spike mutation patterns in vaccine breakthrough sequences.**

Highlighter plot showing spike amino acid mutations of 132 SARS-CoV-2 vaccine breakthrough sequences compared to the Wuhan-Hu-1 reference sequence as master (top line). Mutations are shown as ticks, color-coded according to the legend at the bottom. Frequently occurring key mutations are indicated by triangles and labeled. The presence of S112L and hospitalization of study participants are indicated by brown and black symbols, respectively. Study sequences are sorted according to time of sampling, between May 1st and August 3rd, 2021. Iota (B.1.526), Alpha (B.1.1.7), Gamma (P.1), Delta (B.1.617.2 and AY.*), and Delta subvariant AY.25 sequences are highlighted and their range of occurrences indicated.

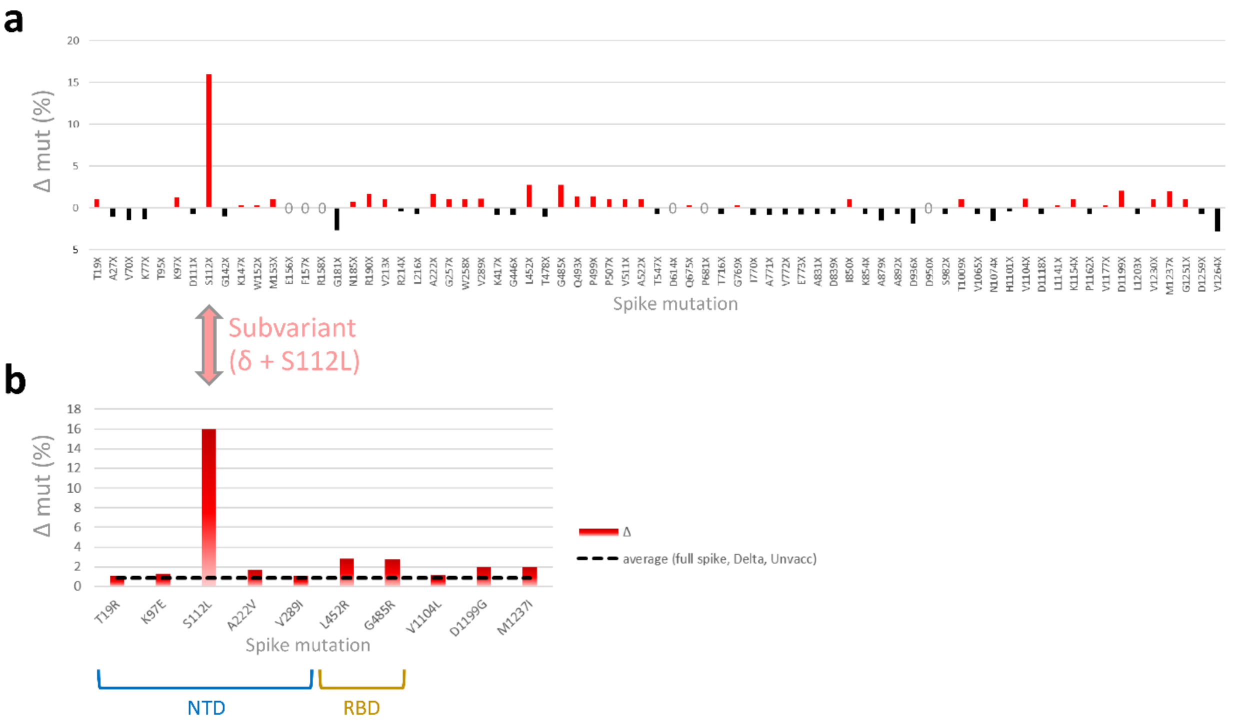

**Figure S6. Site-specific spike mutation analysis in SARS-CoV-2 Delta vaccine breakthrough sequences compared to Delta infections in unvaccinated controls.**

**a**, Comparison of site-specific amino acid mutation (mut) frequencies in spike in the subset of 101 Delta vaccine breakthrough sequences compared to 139 Delta unvaccinated controls from the same cohort. The Wuhan-Hu-1 sequence served as reference to call mutations per site, and all spike mutation sites of the study sequences are shown along the x-axis according to their spike position (n=72). The mirror plot displays differences of mutation frequencies per spike residue between vaccinated and unvaccinated groups; red bars (facing up) refer to higher mutation rates in vaccinated, whereas black bars (facing down) refer to higher mutation rates in unvaccinated controls. Mutations that are 100% conserved in both groups and thus have 0% difference are indicated by a zero on the x-axis. **b**, Enrichment of spike mutations in SARS-CoV-2 Delta vaccine breakthrough sequences. All sites with greater spike mutation rates in vaccinated compared to unvaccinated Delta infections are shown; sites with unique occurrences of mutations in Delta breakthrough cases were disregarded. Mutation sites in the spike N-terminal domain (NTD) and receptor binding domain (RBD) are highlighted. The dashed black line indicates the average mutation frequency across all spike residues in the Delta unvaccinated control data set compared to Wuhan-Hu-1 as reference (n=139).

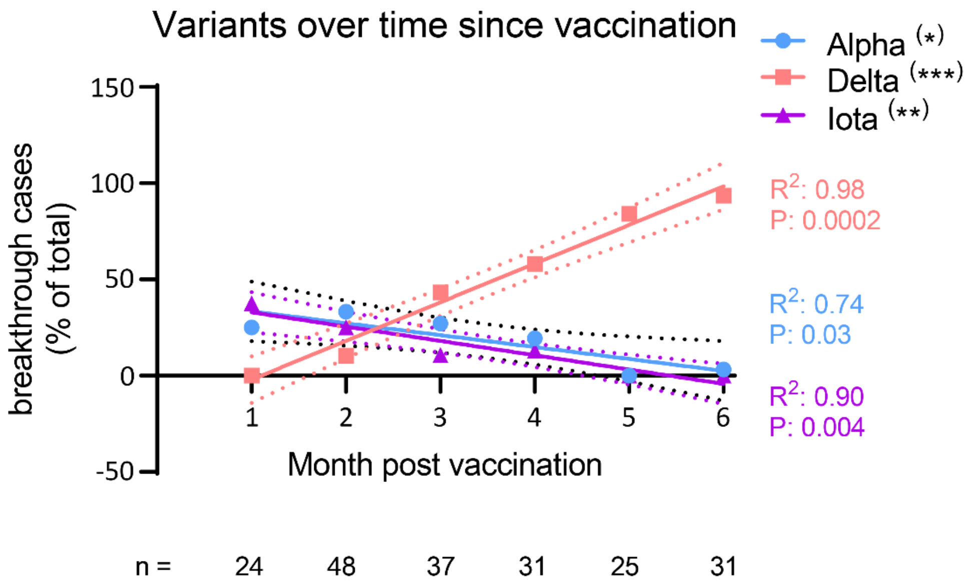

**Figure S7. Ratios of breakthrough SARS-CoV-2 infections by variant and time post vaccination.** Linear regression analysis of breakthrough infection rates per variant against time post vaccination. Significant results are highlighted by asterisks and labeled with the goodness of fit ($R^2$) and P values. Fitted lines with 95% confidence intervals are shown. * P<0.05, ** P<0.01, *** P<0.005.

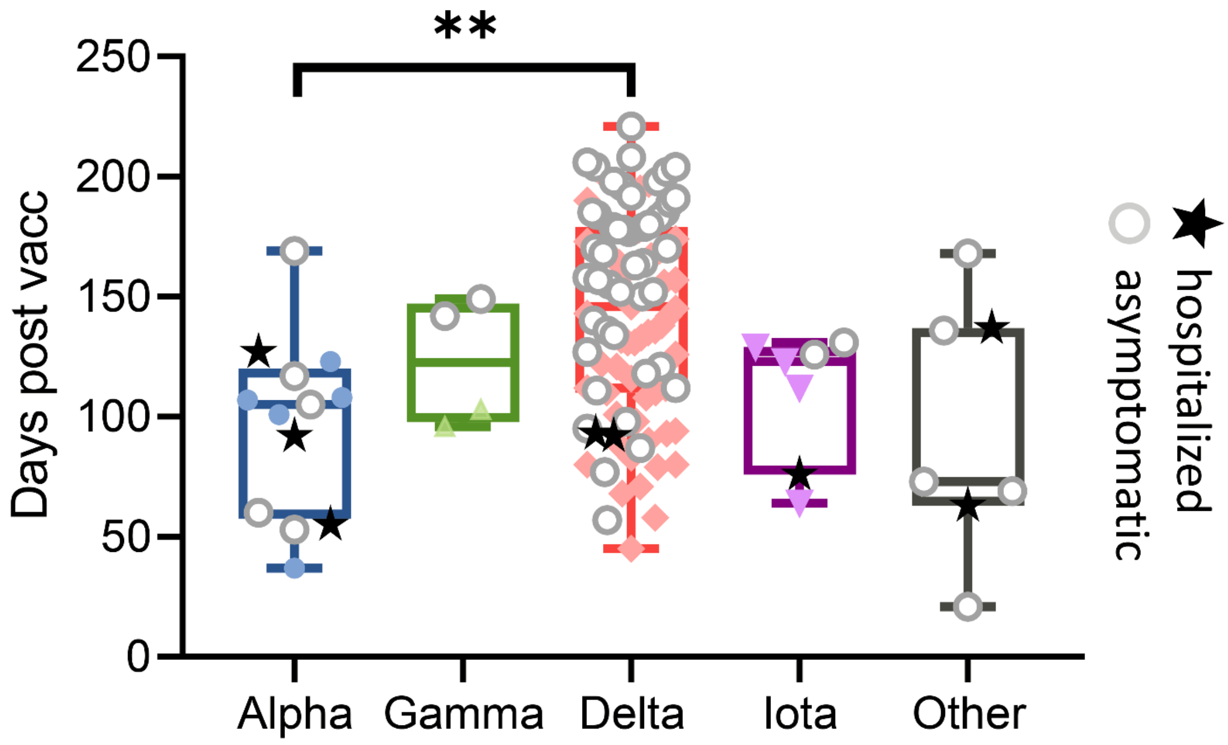

**Figure S8. Breakthrough SARS-CoV-2 infections by variant, time post vaccination, and clinical status.**

Box plots summarizing SARS-CoV-2 diagnosis in days post full vaccination (vacc) by variant. Whiskers indicate the range, boxes the interquartile range, and horizontal line the median of values. Symptomatic cases are shown as colored symbols, hospitalized cases as black stars, and asymptomatic cases as gray circles with white fill. ** $P<0.01$ (Kruskal-Wallis test).

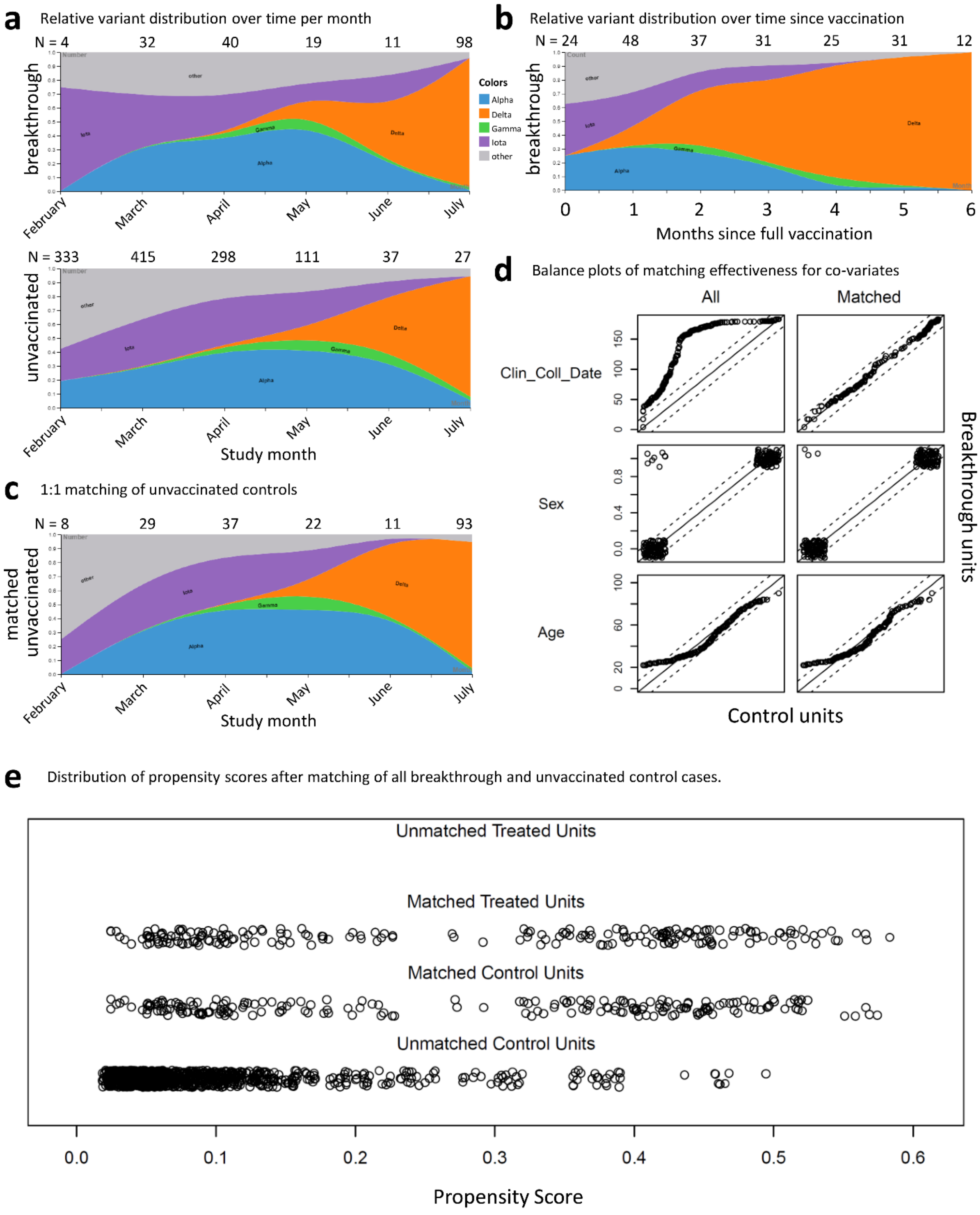

**Figure S9. SARS-CoV-2 variant distribution in vaccinate breakthrough and unvaccinated cases and quality control of matching.**

**a**, Streamgraphs of relative variant distribution over time (study months). Accumulated case numbers per month are shown on top; August was omitted because of low case numbers (<5).

**b**, Streamgraphs of relative variant distribution over time since vaccination, starting at day 14 after the last dose for full vaccination. Accumulated case numbers per thirty-day periods (months) are shown on top. **c**, Streamgraphs of relative variant distribution over time (study months) in unvaccinated controls after 1:1 matching of each breakthrough case to one unvaccinated control patient. **d**, Empirical quantile-quantile (eQQ) plots to study covariate distribution before (left) and after (right) matching. **e**, Distribution of propensity scores after matching of 208 breakthrough and 1329 unvaccinated control cases, for which we obtained full genome SARS-CoV-2 sequences between February and August 2021.

### Supplemental Tables

**Table S1. Full specifications of 132 SARS-CoV-2 breakthrough infections.**

| Sex | Age Range | State | Vaccine Type | Days post vaccination | GISAID Accession | GISAID Virus Name | GISAID Clade | Pango Lineage | Ct | Symptoms | Hospitalization | Notes for hospitalizations |
|---|---|---|---|---|---|---|---|---|---|---|---|---|
| M | 80-89 | NY | Pfizer | 55 | EPI_ISL_2161733 | NYULH1757 | GRY | B.1.1.7 | 23 | Y | Y | heart failure, cardiac amyloid, initially SARS-CoV-2 IgG negative |
| F | 90-99 | NY | Pfizer | 76 | EPI_ISL_2161734 | NYULH1758 | GH | B.1.526 | 12 | Y | Y | sarcoidosis on steroids chronically, COPD |
| M | 50-59 | NY | Pfizer | 112 | EPI_ISL_2202807 | NYULH1808 | G | B.1.617.2 | 22 | Y | N | |
| F | 20-29 | NY | Pfizer | 108 | EPI_ISL_2202790 | NYULH1809 | GRY | B.1.1.7 | 23 | Y | N | |
| M | 20-29 | NY | Pfizer | 117 | EPI_ISL_2202791 | NYULH1810 | GRY | B.1.1.7 | 27 | N | N | |
| M | 30-39 | NY | Pfizer | 64 | EPI_ISL_2427424 | NYULH1839 | GH | B.1.637 | 25 | Y | N | |
| F | 50-59 | NY | Moderna | 101 | EPI_ISL_2427425 | NYULH1840 | GRY | B.1.1.7 | 20 | Y | N | |
| F | 50-59 | NY | Pfizer | 127 | EPI_ISL_2427466 | NYULH1882 | GRY | B.1.1.7 | 20 | Y | Y | ESRD, SLE |
| F | 40-49 | NY | Pfizer | 105 | EPI_ISL_2427476 | NYULH1892 | GRY | B.1.1.7 | 19 | N | N | |
| M | 50-59 | NY | Moderna | 96 | EPI_ISL_2427477 | NYULH1893 | GR | P.1 | 20 | Y | N | |
| F | 70-79 | NY | Pfizer | 53 | EPI_ISL_2427479 | NYULH1895 | GRY | B.1.1.7 | 23 | N | N | |
| F | 40-49 | NY | Pfizer | 112 | EPI_ISL_2427480 | NYULH1896 | GH | B.1.637 | 18 | Y | N | |
| M | 60-69 | NJ | Moderna | 92 | EPI_ISL_2427484 | NYULH1900 | GRY | B.1.1.7 | 26 | Y | Y | ILD,RA on rituximab |
| M | 30-39 | NY | Pfizer | 37 | EPI_ISL_2663526 | NYULH2292 | GRY | B.1.1.7 | 23 | Y | N | |
| F | 30-39 | NY | Pfizer | 126 | EPI_ISL_2663530 | NYULH2297 | GH | B.1 | 19 | N | N | |
| M | 80-89 | NY | Moderna | 103 | EPI_ISL_2663535 | NYULH2302 | GR | P.1 | 30 | Y | N | |
| M | 20-29 | NY | Pfizer | 129 | EPI_ISL_2663540 | NYULH2308 | GH | B.1.526 | 22 | Y | N | |
| M | 20-29 | NY | Pfizer | 123 | EPI_ISL_2663541 | NYULH2309 | GH | B.1.526 | 30 | Y | N | |

| | | | | | | | | | | | | |
|---|---|---|---|---|---|---|---|---|---|---|---|---|
| M | 30-39 | NY | Pfizer | 131 | EPI_ISL_2663542 | NYULH2310 | GH | B.1.526 | 29 | N | N | |
| M | 80-89 | NY | Pfizer | 63 | EPI_ISL_2663547 | NYULH2315 | GH | B.1.596 | 17 | Y | Y (deceased) | metastatic hepatocellular carcinoma, COVID-19 and superimposed bacterial PNA |
| M | 30-39 | NJ | Pfizer | 146 | EPI_ISL_2663553 | NYULH2323 | G | B.1.617.2 | 18 | Y | N | |
| F | 70-79 | FL | Moderna | 140 | EPI_ISL_3245465 | NYULH2331 | GK | B.1.617.2 | 35 | N | N | |
| M | 70-79 | FL | Moderna | 138 | EPI_ISL_3245466 | NYULH2332 | GV | AY.10 | 13 | Y | N | |
| M | 60-69 | NY | Pfizer | 60 | EPI_ISL_2753947 | NYULH2333 | GRY | B.1.1.7 | 33 | N | N | |
| F | 80-89 | NY | Pfizer | 69 | EPI_ISL_2753948 | NYULH2334 | G | B.1.525 | 32 | N | N | |
| F | 40-49 | NY | Pfizer | 107 | EPI_ISL_2753949 | NYULH2335 | GRY | B.1.1.7 | 32 | Y | N | |
| M | 50-59 | NY | Pfizer | 21 | EPI_ISL_3245479 | NYULH2367 | GH | B.1.621 | 26 | N | N | |
| F | 20-29 | NY | Pfizer | 169 | EPI_ISL_3245485 | NYULH2373 | GR | B.1.1.7 | 21 | N | N | |
| F | 20-29 | NY | Pfizer | 173 | EPI_ISL_3245486 | NYULH2374 | GK | AY.16 | 16 | Y | N | |
| F | 20-29 | NJ | Pfizer | 164 | EPI_ISL_3245487 | NYULH2375 | GK | AY.25 | 26 | N | N | |
| F | 80-89 | NY | Pfizer | 136 | EPI_ISL_3245490 | NYULH2378 | GH | B.1.621.1 | 19 | N | Y | COPD |
| M | 60-69 | NY | Pfizer | 101 | EPI_ISL_3245493 | NYULH2381 | GV | AY.10 | 20 | Y | N | |
| F | 70-79 | NY | Pfizer | 87 | EPI_ISL_3245494 | NYULH2382 | GK | B.1.617.2 | 21 | Y | N | |
| M | 20-29 | NY | Pfizer | 161 | EPI_ISL_3245495 | NYULH2383 | G | AY.24 | 15 | Y | N | |
| M | 30-39 | NY | Pfizer | 170 | EPI_ISL_3245496 | NYULH2384 | GK | B.1.617.2 | 18 | N | N | |
| F | 50-59 | NY | Pfizer | 123 | EPI_ISL_3245505 | NYULH2393 | GR | B.1.1.7 | NA | Y | N | |
| M | 30-39 | NY | Pfizer | 168 | EPI_ISL_3245509 | NYULH2397 | GK | AY.25 | 33 | N | N | |
| F | 20-29 | NY | Pfizer | 150 | EPI_ISL_3245510 | NYULH2398 | GK | AY.25 | 19 | N | N | |
| F | 40-49 | NY | Pfizer | 77 | EPI_ISL_3245512 | NYULH2400 | GK | B.1.617.2 | 27 | N | N | |
| F | 20-29 | NY | Pfizer | 170 | EPI_ISL_3245514 | NYULH2402 | GK | B.1.617.2 | 21 | N | N | |
| M | 50-59 | NY | Moderna | 93 | EPI_ISL_3245515 | NYULH2403 | GK | AY.14 | 18 | Y | N | |
| F | 40-49 | NY | Moderna | 163 | EPI_ISL_3245516 | NYULH2404 | GK | AY.14 | 21 | N | N | |

| | | | | | | | | | | | |
|---|---|---|---|---|---|---|---|---|---|---|---|
| M | 20-29 | NY | Pfizer | 177 | EPI_ISL_3245518 | NYULH2406 | GK | B.1.617.2 | 28 | N | N |
| F | 20-29 | NY | Pfizer | 149 | EPI_ISL_3245519 | NYULH2407 | GR | P.1 | 17 | N | N |
| F | 20-29 | NY | Pfizer | 179 | EPI_ISL_3245521 | NYULH2409 | GK | AY.14 | 28 | N | N |
| M | 30-39 | NY | Pfizer | 71 | EPI_ISL_3245525 | NYULH2413 | GK | AY.25 | 19 | Y | N |
| F | 30-39 | NY | Pfizer | 58 | EPI_ISL_3245526 | NYULH2414 | GK | AY.25 | 19 | Y | N |
| F | 20-29 | NY | Pfizer | 164 | EPI_ISL_3245527 | NYULH2415 | GK | B.1.617.2 | 29 | Y | N |
| M | 30-39 | NY | Pfizer | 45 | EPI_ISL_3245529 | NYULH2417 | GK | B.1.617.2 | 17 | Y | N |
| M | 50-59 | NY | Pfizer | 110 | EPI_ISL_3245531 | NYULH2419 | GK | AY.25 | 26 | Y | N |
| F | 40-49 | NY | Pfizer | 152 | EPI_ISL_3245532 | NYULH2420 | GK | B.1.617.2 | 19 | N | N |
| F | 50-59 | NY | Pfizer | 155 | EPI_ISL_3245534 | NYULH2422 | GK | B.1.617.2 | 18 | N | N |
| M | 30-39 | NY | Pfizer | 184 | EPI_ISL_3245542 | NYULH2430 | GV | AY.26 | 18 | N | N |
| M | 30-39 | NY | Pfizer | 80 | EPI_ISL_3245544 | NYULH2432 | GK | AY.14 | 26 | Y | N |
| M | 40-49 | NY | Pfizer | 73 | EPI_ISL_3245562 | NYULH2450 | GH | B.1.621 | 30 | N | N |
| M | 70-79 | NY | Pfizer | 136 | EPI_ISL_3245563 | NYULH2451 | GK | AY.3 | 17 | N | N |
| F | 70-79 | NY | Pfizer | 136 | EPI_ISL_3245564 | NYULH2452 | GK | AY.3 | 18 | Y | N |
| F | 20-29 | NJ | Pfizer | 185 | EPI_ISL_3245568 | NYULH2456 | GK | B.1.617.2 | 31 | N | N |
| M | 30-39 | NY | Moderna | 132 | EPI_ISL_3245570 | NYULH2458 | GK | B.1.617.2 | 19 | Y | N |
| F | 50-59 | NY | Pfizer | 177 | EPI_ISL_3245571 | NYULH2459 | GK | B.1.617.2 | 18 | Y | N |
| F | 30-39 | NY | Pfizer | 178 | EPI_ISL_3245572 | NYULH2460 | GK | AY.14 | 32 | Y | N |
| M | 30-39 | NY | Pfizer | 185 | EPI_ISL_3245575 | NYULH2463 | GK | AY.25 | 25 | N | N |
| M | 30-39 | NJ | Moderna | 142 | EPI_ISL_3245576 | NYULH2464 | GR | P.1 | 22 | N | N |
| M | 30-39 | NY | Pfizer | 181 | EPI_ISL_3245579 | NYULH2467 | GK | AY.26 | 22 | Y | N |
| M | 30-39 | NY | Pfizer | 80 | EPI_ISL_3245587 | NYULH2475 | GK | B.1.617.2 | 23 | Y | N |
| M | 20-29 | NY | Pfizer | 121 | EPI_ISL_3245588 | NYULH2476 | GK | AY.25 | 21 | Y | N |
| F | 50-59 | NY | Pfizer | 137 | EPI_ISL_3245589 | NYULH2477 | GH | C.38 | 32 | Y | N |
| M | 30-39 | NJ | Pfizer | 168 | EPI_ISL_3245590 | NYULH2478 | G | B.1 | 32 | N | N |

| | | | | | | | | | | | | |
|---|---|---|---|---|---|---|---|---|---|---|---|---|
| F | 20-29 | NY | Pfizer | 178 | EPI_ISL_3245591 | NYULH2479 | GK | B.1.617.2 | 30 | N | N | |
| M | 30-39 | NY | Pfizer | 178 | EPI_ISL_3245592 | NYULH2480 | GK | B.1.617.2 | 18 | N | N | |
| F | 30-39 | NY | Pfizer | 179 | EPI_ISL_3245593 | NYULH2481 | G | B.1.617.2 | 34 | N | N | |
| F | 30-39 | NJ | Pfizer | 181 | EPI_ISL_3245594 | NYULH2482 | GK | B.1.617.2 | 20 | Y | N | |
| F | 50-59 | NY | Pfizer | 94 | EPI_ISL_3245608 | NYULH2496 | GK | AY.35 | NA | Y | N | |
| M | 50-59 | NY | Pfizer | 92 | EPI_ISL_3245609 | NYULH2497 | GK | B.1.617.2 | NA | Y | Y | IDDM |
| M | 70-79 | NY | Pfizer | 93 | EPI_ISL_3245610 | NYULH2498 | GK | B.1.617.2 | NA | Y | Y | HTN, CAD, history of NHL |
| M | 50-59 | NY | Pfizer | 157 | EPI_ISL_3245622 | NYULH2510 | GK | B.1.617.2 | 21 | Y | N | |
| F | 60-69 | NY | Pfizer | 126 | EPI_ISL_3245650 | NYULH2538 | GK | B.1.617.2 | 20 | Y | N | |
| M | 70-79 | NY | Pfizer | 142 | EPI_ISL_3245651 | NYULH2539 | GK | B.1.617.2 | 18 | Y | N | |
| F | 20-29 | NY | Pfizer | 183 | EPI_ISL_3245652 | NYULH2540 | GK | B.1.617.2 | 18 | N | N | |
| M | 20-29 | NY | Pfizer | 198 | EPI_ISL_3245653 | NYULH2541 | GK | AY.20 | 18 | N | N | |
| F | 30-39 | NY | Pfizer | 197 | EPI_ISL_3245654 | NYULH2542 | GK | B.1.617.2 | 19 | Y | N | |
| M | 30-39 | NY | Pfizer | 190 | EPI_ISL_3245655 | NYULH2543 | GK | AY.25 | 17 | Y | N | |
| M | 40-49 | NY | Pfizer | 87 | EPI_ISL_3245656 | NYULH2544 | GK | AY.25 | 29 | N | N | |
| F | 30-39 | NY | Pfizer | 174 | EPI_ISL_3245658 | NYULH2546 | GK | B.1.617.2 | 21 | Y | N | |
| F | 20-29 | NJ | Pfizer | 127 | EPI_ISL_3245659 | NYULH2547 | GK | B.1.617.2 | 25 | N | N | |
| F | 50-59 | NY | Pfizer | 90 | EPI_ISL_3245660 | NYULH2548 | GK | AY.3 | 26 | Y | N | |
| F | 30-39 | NY | Pfizer | 189 | EPI_ISL_3245661 | NYULH2549 | GK | AY.26 | 22 | N | N | |
| F | 40-49 | NY | Pfizer | 98 | EPI_ISL_3245662 | NYULH2550 | GK | AY.20 | 19 | Y | N | |
| F | 20-29 | NY | Pfizer | 180 | EPI_ISL_3245663 | NYULH2560 | GK | AY.25 | 20 | N | N | |
| M | 60-69 | NY | Pfizer | 112 | EPI_ISL_3245665 | NYULH2562 | GK | AY.25 | 17 | Y | N | |
| M | 60-69 | NY | Pfizer | 143 | EPI_ISL_3245666 | NYULH2563 | GK | B.1.617.2 | 19 | Y | N | |
| F | 50-59 | NY | Pfizer | 135 | EPI_ISL_3245667 | NYULH2564 | GK | AY.16 | 14 | Y | N | |
| M | 20-29 | NY | Pfizer | 134 | EPI_ISL_3347434 | NYULH2596 | GK | B.1.617.2 | 22 | N | N | |
| M | 60-69 | NY | Pfizer | 138 | EPI_ISL_3347435 | NYULH2597 | GK | B.1.617.2 | 31 | Y | N | |

| | | | | | | | | | | | |
|---|---|---|---|---|---|---|---|---|---|---|---|
| M | 30-39 | NY | Pfizer | 158 | EPI_ISL_3347438 | NYULH2600 | GK | AY.16 | 18 | N | N |
| F | 30-39 | NJ | Moderna | 156 | EPI_ISL_3347439 | NYULH2601 | GK | B.1.617.2 | 25 | Y | N |
| F | 30-39 | NY | Pfizer | 95 | EPI_ISL_3347440 | NYULH2602 | GK | AY.3 | 18 | N | N |
| M | 60-69 | NY | Janssen | 145 | EPI_ISL_3347441 | NYULH2603 | GK | AY.3 | 17 | Y | N |
| F | 20-29 | NY | Pfizer | 182 | EPI_ISL_3347442 | NYULH2604 | GK | AY.16 | 20 | Y | N |
| F | 30-39 | NY | Pfizer | 167 | EPI_ISL_3347443 | NYULH2605 | GK | AY.3 | 21 | Y | N |
| F | 20-29 | NY | Pfizer | 152 | EPI_ISL_3347444 | NYULH2606 | G | AY.16 | 19 | N | N |
| M | 50-59 | NY | Pfizer | 112 | EPI_ISL_3347445 | NYULH2607 | GK | B.1.617.2 | 19 | N | N |
| M | 60-69 | NY | Janssen | 68 | EPI_ISL_3347446 | NYULH2608 | GK | AY.25 | 22 | Y | N |
| F | 20-29 | NY | Pfizer | 204 | EPI_ISL_3347447 | NYULH2609 | GK | B.1.617.2 | 28 | N | N |
| M | 20-29 | NY | Pfizer | 206 | EPI_ISL_3347448 | NYULH2610 | G | AY.25 | 18 | N | N |
| M | 40-49 | NY | Moderna | 122 | EPI_ISL_3347449 | NYULH2611 | GK | B.1.617.2 | 26 | Y | N |
| F | 60-69 | NY | Moderna | 57 | EPI_ISL_3347450 | NYULH2612 | GK | B.1.617.2 | 25 | N | N |
| F | 60-69 | NY | Pfizer | 202 | EPI_ISL_3347451 | NYULH2613 | GK | AY.16 | 16 | N | N |
| M | 30-39 | NY | Pfizer | 196 | EPI_ISL_3347452 | NYULH2614 | GK | AY.20 | 21 | N | N |
| F | 20-29 | NY | Pfizer | 191 | EPI_ISL_3347453 | NYULH2615 | GK | B.1.617.2 | 20 | N | N |
| F | 30-39 | NY | Moderna | 169 | EPI_ISL_3347454 | NYULH2616 | GK | B.1.617.2 | 24 | Y | N |
| M | 50-59 | NY | Pfizer | 121 | EPI_ISL_3347455 | NYULH2617 | GK | AY.20 | 28 | N | N |
| F | 30-39 | NY | Pfizer | 79 | EPI_ISL_3347456 | NYULH2618 | GK | AY.4 | 18 | Y | N |
| M | 30-39 | NY | Janssen | 131 | EPI_ISL_3347457 | NYULH2619 | GK | B.1.617.2 | 20 | Y | N |
| F | 40-49 | NY | Pfizer | 110 | EPI_ISL_3347459 | NYULH2621 | GK | B.1.617.2 | 25 | Y | N |
| F | 20-29 | NY | Pfizer | 108 | EPI_ISL_3347460 | NYULH2622 | GK | AY.3 | 19 | Y | N |
| M | 30-39 | NY | Pfizer | 83 | EPI_ISL_3347461 | NYULH2623 | GK | B.1.617.2 | 18 | Y | N |
| M | 30-39 | NY | Pfizer | 98 | EPI_ISL_3347462 | NYULH2624 | GK | AY.25 | 27 | N | N |
| M | 30-39 | NY | Pfizer | 204 | EPI_ISL_3347463 | NYULH2625 | GK | AY.25 | 20 | N | N |
| M | 40-49 | NY | Pfizer | 200 | EPI_ISL_3347464 | NYULH2626 | G | AY.3 | 19 | Y | N |

| | | | | | | | | | | | |
|---|---|---|---|---|---|---|---|---|---|---|---|
| F | 30-39 | NY | Pfizer | 198 | EPI_ISL_3347465 | NYULH2627 | GK | B.1.617.2 | 25 | N | N |
| F | 30-39 | NY | Pfizer | 192 | EPI_ISL_3347466 | NYULH2628 | GK | AY.24 | 24 | N | N |
| F | 20-29 | TX | Pfizer | 157 | EPI_ISL_3347467 | NYULH2629 | GK | AY.16 | 20 | N | N |
| M | 50-59 | NY | Pfizer | 133 | EPI_ISL_3347468 | NYULH2630 | G | B.1.617.2 | 16 | Y | N |
| F | 20-29 | NY | Pfizer | 116 | EPI_ISL_3347469 | NYULH2631 | G | B.1.617.2 | NA | Y | N |
| M | 30-39 | NY | Pfizer | 167 | EPI_ISL_3347470 | NYULH2632 | GK | B.1.617.2 | 20 | Y | N |
| M | 40-49 | NY | Pfizer | 221 | EPI_ISL_3347473 | NYULH2635 | GK | AY.16 | 17 | N | N |
| M | 60-69 | NY | Pfizer | 131 | EPI_ISL_3347474 | NYULH2636 | GK | AY.25 | 30 | Y | N |
| F | 30-39 | NY | Pfizer | 111 | EPI_ISL_3347476 | NYULH2638 | G | B.1.617.2 | 17 | N | N |
| M | 20-29 | NY | Pfizer | 208 | EPI_ISL_3347433 | NYULH2595 | GK | AY.25 | 25 | N | N |
| M | 30-39 | NY | Pfizer | 195 | EPI_ISL_3347432 | NYULH2594 | GK | AY.3 | 20 | Y | N |
| F | 60-69 | NY | Pfizer | 118 | EPI_ISL_3347472 | NYULH2634 | GK | B.1.617.2 | 20 | N | N |

Abbreviations: CAD: coronary artery disease; COPD: chronic obstructive pulmonary disease; ESRD: end-stage renal disease; HTN: hypertension; IDDM: Insulin-dependent diabetes mellitus; ILD: interstitial lung disease; NHL: Non-Hodgkin lymphoma; PNA: pneumonia; RA: rheumatoid arthritis; SLE: systemic lupus erythematosus.

**Table S2. Full specifications of 283 SARS-CoV-2 infections in unvaccinated controls.**

| Sex | Age Range | State | GISAID Accession | GISAID Virus Name | GISAID Clade | Pango Lineage | Ct | Symptoms | Hospitalization | COVID-19 main cause of hospitalization | COVID-19 cause of death |
|---|---|---|---|---|---|---|---|---|---|---|---|
| M | 80-89 | NY | EPI_ISL_2202777 | NYULH1745 | GH | B.1.526 | 20 | Y | Y | Y | |
| M | 70-79 | NY | EPI_ISL_2161723 | NYULH1746 | GH | B.1.637 | 22 | Y | Y | Y | |
| F | 50-59 | NY | EPI_ISL_2161735 | NYULH1759 | S | A.2.5 | 16 | Y | N | | |
| M | 50-59 | NY | EPI_ISL_2161736 | NYULH1760 | GRY | B.1.1.7 | 22 | N | Y | Y | |
| F | 60-69 | NY | EPI_ISL_2161737 | NYULH1761 | GR | C.36.3.1 | 17 | Y | N | | |
| F | 60-69 | NY | EPI_ISL_2161738 | NYULH1762 | GR | C.37.1 | 23 | Y | N | | |
| M | 50-59 | NY | EPI_ISL_2161739 | NYULH1763 | GR | B.1.1.318 | 18 | na | N | | |
| F | 50-59 | NY | EPI_ISL_2161740 | NYULH1764 | GRY | B.1.1.7 | 18 | na | N | | |
| F | 40-49 | NY | EPI_ISL_2161741 | NYULH1765 | GH | B.1.526 | 18 | na | N | | |
| F | 20-29 | NY | EPI_ISL_2161742 | NYULH1766 | GRY | B.1.1.7 | 27 | N | N | | |
| M | <10 | NY | EPI_ISL_2161743 | NYULH1767 | GRY | B.1.1.7 | 23 | N | N | | |
| F | 80-89 | NY | EPI_ISL_2202768 | NYULH1768 | GRY | B.1.1.7 | 22 | Y | Y | Y | |
| F | 30-39 | NY | EPI_ISL_2161744 | NYULH1769 | GH | B.1.526 | 23 | Y | N | | |
| F | 90-99 | NY | EPI_ISL_2161745 | NYULH1770 | GRY | B.1.1.7 | 23 | N | Y | N | |
| F | 50-59 | NY | EPI_ISL_2161748 | NYULH1773 | GRY | B.1.1.7 | 23 | Y | N | | |
| F | 50-59 | NY | EPI_ISL_2161749 | NYULH1774 | GRY | B.1.1.7 | 25 | Y | N | | |
| M | 60-69 | NY | EPI_ISL_2161750 | NYULH1775 | GH | B.1.526 | 26 | na | N | | |
| F | 60-69 | NY | EPI_ISL_2161776 | NYULH1803 | S | A.2.5 | 26 | N | N | | |
| F | 20-29 | NJ | EPI_ISL_2161777 | NYULH1804 | GH | B.1.526 | 18 | na | N | | |
| F | <10 | NJ | EPI_ISL_2161778 | NYULH1805 | GH | B.1.526 | 27 | N | N | | |
| M | 50-59 | NY | EPI_ISL_2161779 | NYULH1806 | GH | B.1.526 | 21 | Y | N | | |
| F | 50-59 | NY | EPI_ISL_2161780 | NYULH1807 | GRY | B.1.1.7 | 28 | N | N | | |
| F | 50-59 | NY | EPI_ISL_2427403 | NYULH1817 | GRY | B.1.1.7 | 23 | Y | Y | Y | |

| | | | | | | | | | | | |
|---|---|---|---|---|---|---|---|---|---|---|---|
| F | 50-59 | NY | EPI_ISL_2427404 | NYULH1818 | GR | P.1 | 23 | Y | N | | |
| F | 50-59 | NY | EPI_ISL_2427405 | NYULH1819 | GH | B.1.526 | 20 | Y | N | | |
| F | 60-69 | NY | EPI_ISL_2427406 | NYULH1820 | GR | P.1 | 24 | Y | Y | Y | |
| M | 80-89 | NY | EPI_ISL_2427407 | NYULH1821 | GRY | B.1.1.7 | 28 | Y | Y | Y | Y |
| F | 50-59 | NY | EPI_ISL_2427408 | NYULH1822 | GR | P.1 | 23 | Y | N | | |
| F | 30-39 | NY | EPI_ISL_2427409 | NYULH1823 | GRY | B.1.1.7 | 25 | N | Y | N | |
| F | 20-29 | NY | EPI_ISL_2443066 | NYULH1824 | G | B.1.627 | 18 | Y | N | | |
| M | 70-79 | NY | EPI_ISL_2427410 | NYULH1825 | GR | P.1 | 29 | N | Y | Y | |
| F | 80-89 | NY | EPI_ISL_2427411 | NYULH1826 | GH | B.1.526 | 18 | N | Y | N | |
| M | 60-69 | NY | EPI_ISL_2427413 | NYULH1828 | GRY | B.1.1.7 | 27 | Y | N | | |
| F | <20 | NY | EPI_ISL_2427414 | NYULH1829 | GH | B.1.637 | 20 | Y | N | | |
| M | 40-49 | NY | EPI_ISL_2427415 | NYULH1830 | GRY | B.1.1.7 | 22 | Y | N | | |
| F | 40-49 | NY | EPI_ISL_2427416 | NYULH1831 | GR | P.1.10 | 19 | na | N | | |
| F | 20-29 | NY | EPI_ISL_2427417 | NYULH1832 | GH | B.1.526 | 26 | Y | N | | |
| M | 30-39 | NY | EPI_ISL_2427418 | NYULH1833 | GH | B.1.526 | 24 | na | N | | |
| M | 40-49 | NY | EPI_ISL_2427420 | NYULH1835 | GH | B.1.637 | 20 | Y | N | | |
| F | 50-59 | NY | EPI_ISL_2427421 | NYULH1836 | GH | B.1.526 | 20 | Y | N | | |
| F | 60-69 | NY | EPI_ISL_2427423 | NYULH1838 | GRY | B.1.1.7 | 20 | na | N | | |
| F | 50-59 | NY | EPI_ISL_2427427 | NYULH1842 | GRY | B.1.1.7 | 20 | Y | N | | |
| F | 90-99 | NY | EPI_ISL_2427428 | NYULH1843 | GH | B.1.526 | 20 | Y | Y | Y | |
| F | 80-89 | NY | EPI_ISL_2427429 | NYULH1844 | GRY | B.1.1.7 | 17 | Y | Y | Y | |
| F | 30-39 | SC | EPI_ISL_2427430 | NYULH1845 | GRY | B.1.1.7 | 28 | Y | N | | |
| M | <20 | NY | EPI_ISL_2427431 | NYULH1846 | GRY | B.1.1.7 | 16 | Y | N | | |
| F | 60-69 | NY | EPI_ISL_2427432 | NYULH1847 | GR | P.1 | 20 | Y | Y | Y | |
| M | 30-39 | NY | EPI_ISL_2427434 | NYULH1849 | GH | B.1.526 | 24 | Y | N | | |
| M | 50-59 | NY | EPI_ISL_2427435 | NYULH1850 | GRY | B.1.1.7 | 32 | Y | Y | Y | |

| | | | | | | | | | | | |
|---|---|---|---|---|---|---|---|---|---|---|---|
| F | 40-49 | NY | EPI_ISL_2427436 | NYULH1851 | GRY | B.1.1.7 | 28 | N | Y | N | |
| F | 30-39 | NY | EPI_ISL_2427437 | NYULH1852 | GRY | B.1.1.7 | 23 | Y | N | | |
| M | 80-89 | NY | EPI_ISL_2427438 | NYULH1853 | GH | B.1.637 | 27 | Y | Y | Y | |
| M | 40-49 | NY | EPI_ISL_2427439 | NYULH1854 | GR | B.1.1.7 | 18 | Y | N | | |
| M | 60-69 | NY | EPI_ISL_2427440 | NYULH1855 | GRY | B.1.1.7 | 24 | Y | Y | Y | |
| F | 20-29 | NY | EPI_ISL_2427441 | NYULH1856 | G | B.1.617.2 | 18 | Y | N | | |
| F | 20-29 | NY | EPI_ISL_2427442 | NYULH1857 | GRY | B.1.1.7 | 30 | Y | N | | |
| F | <10 | NY | EPI_ISL_2427443 | NYULH1858 | GRY | B.1.1.7 | 20 | Y | N | | |
| M | 80-89 | NY | EPI_ISL_2427444 | NYULH1859 | G | AY.26 | 20 | Y | N | | |
| F | 60-69 | NY | EPI_ISL_2427445 | NYULH1860 | GH | B.1.526 | 22 | Y | N | | |
| F | <10 | NY | EPI_ISL_2427446 | NYULH1861 | GRY | B.1.1.7 | 19 | na | N | | |
| M | 30-39 | NY | EPI_ISL_2427447 | NYULH1862 | GH | B.1.526 | 24 | Y | N | | |
| F | 30-39 | NY | EPI_ISL_2427448 | NYULH1863 | GH | B.1.526 | 18 | Y | N | | |
| F | 30-39 | NY | EPI_ISL_2427449 | NYULH1864 | GH | B.1.526 | 23 | na | N | | |
| M | <20 | NY | EPI_ISL_2427450 | NYULH1865 | GRY | B.1.1.7 | 27 | Y | N | | |
| M | 70-79 | NY | EPI_ISL_2427452 | NYULH1867 | G | B.1.617.2 | 23 | N | Y | Y | |
| F | 50-59 | NY | EPI_ISL_2427453 | NYULH1868 | GH | B.1.637 | 29 | Y | Y | Y | |
| M | 40-49 | NY | EPI_ISL_2427454 | NYULH1869 | G | B.1.617.2 | 30 | Y | Y | Y | |
| F | 80-89 | NY | EPI_ISL_2427455 | NYULH1870 | GRY | Q.3 | 25 | Y | Y | Y | Y |
| M | 60-69 | NY | EPI_ISL_2427456 | NYULH1871 | GRY | B.1.1.7 | 19 | Y | N | | |
| F | 70-79 | NY | EPI_ISL_2427457 | NYULH1872 | GH | B.1.526 | 21 | Y | N | | |
| M | 30-39 | NY | EPI_ISL_2427458 | NYULH1873 | GR | C.36.3 | 29 | N | N | | |
| F | 30-39 | NY | EPI_ISL_2427459 | NYULH1874 | GRY | B.1.1.7 | 24 | Y | N | | |
| F | 60-69 | NY | EPI_ISL_2427460 | NYULH1875 | GRY | B.1.1.7 | 23 | Y | Y | Y | |
| M | 60-69 | NY | EPI_ISL_2427461 | NYULH1876 | GRY | B.1.1.7 | 30 | Y | Y | Y | |
| M | 80-89 | NY | EPI_ISL_2427462 | NYULH1877 | GRY | B.1.1.7 | 27 | N | Y | N | |

| | | | | | | | | | | | |
|---|---|---|---|---|---|---|---|---|---|---|---|
| M | 80-89 | NY | EPI_ISL_2427463 | NYULH1878 | GR | P.1 | NA | Y | Y | Y | |
| F | 60-69 | NY | EPI_ISL_2427464 | NYULH1879 | GR | P.1 | 24 | Y | Y | Y | |
| F | 20-29 | NY | EPI_ISL_2443067 | NYULH1880 | G | B.1.627 | NA | Y | N | | |
| M | 40-49 | NY | EPI_ISL_2427465 | NYULH1881 | GRY | B.1.1.7 | 23 | Y | Y | Y | |
| M | 70-79 | NY | EPI_ISL_2427467 | NYULH1883 | G | B.1.617.2 | 18 | Y | Y | Y | |
| M | 50-59 | NY | EPI_ISL_2427468 | NYULH1884 | GR | P.1 | 28 | Y | Y | Y | |
| F | 20-29 | NY | EPI_ISL_2427469 | NYULH1885 | GRY | B.1.1.7 | 19 | Y | N | | |
| F | <10 | NY | EPI_ISL_2427470 | NYULH1886 | GH | B.1.526 | 31 | Y | N | | |
| M | 50-59 | NY | EPI_ISL_2427471 | NYULH1887 | GRY | B.1.1.7 | 20 | Y | Y | Y | |
| F | 50-59 | NY | EPI_ISL_2427472 | NYULH1888 | GR | C.36.3 | 24 | Y | N | | |
| F | 70-79 | NY | EPI_ISL_2427473 | NYULH1889 | GH | B.1.526 | 28 | Y | Y | Y | |
| M | 80-89 | NY | EPI_ISL_2427474 | NYULH1890 | GRY | B.1.1.7 | 18 | Y | Y | Y | Y |
| F | 70-79 | NY | EPI_ISL_2427475 | NYULH1891 | GRY | B.1.1.7 | 16 | Y | N | | |
| M | <20 | NY | EPI_ISL_2427478 | NYULH1894 | GH | B.1.526 | 23 | Y | N | | |
| F | 60-69 | NY | EPI_ISL_2427482 | NYULH1898 | G | B.1.617.2 | 25 | Y | Y | Y | |
| M | 70-79 | NY | EPI_ISL_2427483 | NYULH1899 | GRY | B.1.1.7 | 17 | Y | N | | |
| M | 40-49 | NY | EPI_ISL_2458185 | NYULH2265 | G | AY.16 | 23 | Y | N | | |
| F | 70-79 | NY | EPI_ISL_2458164 | NYULH2266 | GH | B.1.526 | 22 | N | Y | N | |
| F | 30-39 | NY | EPI_ISL_2458186 | NYULH2267 | GR | B.1.1.318 | 18 | Y | N | | |
| F | 60-69 | NY | EPI_ISL_2458161 | NYULH2268 | GR | P.1.10 | 21 | Y | Y | Y | Y |
| M | 60-69 | NY | EPI_ISL_2458208 | NYULH2269 | GH | B.1.526 | 28 | Y | Y | Y | |
| M | 60-69 | NY | EPI_ISL_2458177 | NYULH2270 | GRY | B.1.1.7 | 28 | Y | Y | Y | |
| M | 80-89 | NY | EPI_ISL_2458170 | NYULH2271 | GRY | B.1.1.7 | 23 | Y | Y | Y | |
| M | 50-59 | NY | EPI_ISL_2458178 | NYULH2272 | GRY | B.1.1.7 | 23 | Y | Y | Y | Y |
| F | 80-89 | NY | EPI_ISL_2458190 | NYULH2273 | GH | B.1.526 | 18 | Y | Y | Y | |
| M | 80-89 | NY | EPI_ISL_2663512 | NYULH2274 | GH | B.1.526 | 30 | Y | Y | Y | |

| | | | | | | | | | | | |
|---|---|---|---|---|---|---|---|---|---|---|---|
| F | 70-79 | NY | EPI_ISL_2663513 | NYULH2275 | GR | B.1.1.7 | 13 | Y | Y | Y | |
| F | 50-59 | NY | EPI_ISL_2663514 | NYULH2277 | GRY | B.1.1.7 | 22 | N | N | | |
| M | 40-49 | NY | EPI_ISL_2663515 | NYULH2278 | GR | P.1.10 | 30 | Y | Y | Y | |
| M | 70-79 | NY | EPI_ISL_2663516 | NYULH2279 | GH | B.1.526 | 30 | Y | Y | Y | Y |
| F | 50-59 | NY | EPI_ISL_2663517 | NYULH2280 | GH | B.1.526 | 24 | Y | Y | Y | |
| M | 70-79 | NY | EPI_ISL_2663519 | NYULH2282 | GRY | B.1.1.7 | 28 | N | N | | |
| F | 30-39 | NY | EPI_ISL_2663520 | NYULH2285 | GR | B.1.1.7 | 22 | na | N | | |
| M | 30-39 | NY | EPI_ISL_2663522 | NYULH2287 | GRY | B.1.1.7 | 31 | Y | N | | |
| M | 50-59 | NY | EPI_ISL_2663523 | NYULH2288 | GRY | B.1.1.7 | 31 | N | N | | |
| F | 30-39 | NY | EPI_ISL_2663524 | NYULH2289 | GR | B.1.1.7 | 34 | N | N | | |
| M | 20-29 | NY | EPI_ISL_2663525 | NYULH2291 | GR | C.37 | 29 | N | N | | |
| M | 40-49 | CA | EPI_ISL_2663528 | NYULH2294 | GR | P.1 | 18 | Y | N | | |
| M | 30-39 | NY | EPI_ISL_2663529 | NYULH2296 | GRY | B.1.1.7 | 22 | N | N | | |
| M | 80-89 | NY | EPI_ISL_2663531 | NYULH2298 | GH | B.1.637 | 34 | Y | Y | N | |
| F | 70-79 | NY | EPI_ISL_2663532 | NYULH2299 | GRY | B.1.1.7 | 23 | Y | Y | Y | |
| M | <10 | NY | EPI_ISL_2663533 | NYULH2300 | GRY | B.1.1.7 | 33 | Y | N | | |
| M | 20-29 | NY | EPI_ISL_2663534 | NYULH2301 | GH | B.1.526 | 31 | Y | N | | |
| M | 40-49 | CT | EPI_ISL_2663536 | NYULH2303 | GRY | B.1.1.7 | 34 | N | N | | |
| F | <10 | NY | EPI_ISL_2663537 | NYULH2304 | GH | B.1.526 | 29 | N | N | | |
| M | 40-49 | NY | EPI_ISL_2663538 | NYULH2305 | GRY | B.1.1.7 | 23 | na | N | | |
| F | 50-59 | NY | EPI_ISL_2663539 | NYULH2307 | GRY | B.1.1.7 | 22 | Y | N | | |
| M | 90-99 | NY | EPI_ISL_2663544 | NYULH2312 | G | B.1.617.2 | NA | Y | Y | Y | Y |
| M | 70-79 | NY | EPI_ISL_2663545 | NYULH2313 | GRY | B.1.1.7 | 27 | Y | Y | Y | |
| F | 20-29 | NY | EPI_ISL_2663546 | NYULH2314 | G | B.1.617.2 | 17 | Y | N | | |
| F | 50-59 | NY | EPI_ISL_2663548 | NYULH2316 | GH | B.1.526 | 13 | Y | Y | Y | |
| M | 50-59 | NY | EPI_ISL_2663550 | NYULH2318 | GH | B.1.526 | 28 | Y | N | | |

| | | | | | | | | | | |
|---|---|---|---|---|---|---|---|---|---|---|
| M | 30-39 | NY | EPI_ISL_2663551 | NYULH2319 | GRY | B.1.1.7 | 26 | Y | N | |
| M | 50-59 | NY | EPI_ISL_2753940 | NYULH2324 | G | B.1.617.2 | 22 | Y | N | |
| M | 50-59 | NY | EPI_ISL_2753941 | NYULH2325 | GR | P.1 | 21 | Y | N | |
| M | 70-79 | NY | EPI_ISL_2753942 | NYULH2326 | G | B.1.617.2 | 22 | Y | Y | Y |
| M | 60-69 | NY | EPI_ISL_2753943 | NYULH2327 | G | B.1.617.2 | 21 | Y | Y | Y |
| F | 20-29 | NY | EPI_ISL_2753946 | NYULH2330 | GR | B.1.1.7 | 15 | Y | N | |
| M | 40-49 | NY | EPI_ISL_2753951 | NYULH2339 | G | AY.26 | 21 | Y | N | |
| M | 50-59 | NY | EPI_ISL_2753952 | NYULH2342 | GR | P.1.10 | 17 | Y | N | |
| M | 30-39 | NY | EPI_ISL_3245467 | NYULH2355 | GR | P.1 | 31 | Y | N | |
| F | 60-69 | NY | EPI_ISL_3245468 | NYULH2356 | GH | B.1.311 | 17 | Y | Y | Y |
| M | 30-39 | NY | EPI_ISL_3245469 | NYULH2357 | G | AY.3 | 20 | Y | Y | Y |
| F | 20-29 | NY | EPI_ISL_3245470 | NYULH2358 | G | AY.25 | 30 | Y | N | |
| M | 30-39 | NY | EPI_ISL_3245472 | NYULH2360 | GR | B.1.1.7 | 19 | Y | Y | Y |
| F | 40-49 | NY | EPI_ISL_3245473 | NYULH2361 | GK | AY.25 | 17 | na | N | |
| M | <10 | NY | EPI_ISL_3245474 | NYULH2362 | GK | AY.25 | 21 | Y | N | |
| F | 30-39 | NY | EPI_ISL_3245475 | NYULH2363 | GR | B.1.1.7 | 26 | Y | N | |
| F | <20 | NY | EPI_ISL_3245476 | NYULH2364 | GK | B.1.617.2 | 22 | Y | N | |
| M | 40-49 | NJ | EPI_ISL_3245477 | NYULH2365 | GK | AY.2 | 22 | N | N | |
| F | 30-39 | NY | EPI_ISL_3245481 | NYULH2369 | GR | P.1 | 33 | N | N | |
| M | 50-59 | NY | EPI_ISL_3245482 | NYULH2370 | GK | AY.25 | 35 | Y | N | |
| F | 20-29 | NY | EPI_ISL_3245492 | NYULH2380 | GK | AY.25 | 26 | Y | N | |
| M | 40-49 | PR | EPI_ISL_3245497 | NYULH2385 | GK | AY.20 | 25 | Y | N | |
| M | 30-39 | NY | EPI_ISL_3245498 | NYULH2386 | GH | B.1.621 | 16 | Y | N | |
| F | 50-59 | NY | EPI_ISL_3245499 | NYULH2387 | GK | B.1.617.2 | 21 | Y | N | |
| F | <20 | NY | EPI_ISL_3245500 | NYULH2388 | GK | B.1.617.2 | 22 | N | N | |
| F | 20-29 | NY | EPI_ISL_3245502 | NYULH2390 | GR | B.1.1.7 | 16 | Y | N | |

| | | | | | | | | | | | |
|---|---|---|---|---|---|---|---|---|---|---|---|
| F | 30-39 | NY | EPI_ISL_3245503 | NYULH2391 | GR | B.1.1.7 | 18 | Y | N | | |
| M | 40-49 | NY | EPI_ISL_3245504 | NYULH2392 | GK | B.1.617.2 | 27 | Y | N | | |
| M | 40-49 | NY | EPI_ISL_3245506 | NYULH2394 | GK | AY.25 | 21 | Y | Y | N | |
| F | 20-29 | NY | EPI_ISL_3245508 | NYULH2396 | G | B.1.628 | 25 | Y | Y | Y | |
| M | 20-29 | NY | EPI_ISL_3245511 | NYULH2399 | GK | AY.16 | 17 | Y | N | | |
| M | 30-39 | NY | EPI_ISL_3245513 | NYULH2401 | GR | B.1.1.7 | 34 | Y | N | | |
| F | 20-29 | NY | EPI_ISL_3245517 | NYULH2405 | GK | B.1.617.2 | 17 | na | N | | |
| M | <20 | NY | EPI_ISL_3245520 | NYULH2408 | GK | B.1.617.2 | 17 | Y | N | | |
| F | 20-29 | NY | EPI_ISL_3245523 | NYULH2411 | GK | B.1.617.2 | 19 | Y | N | | |
| F | <10 | NY | EPI_ISL_3245524 | NYULH2412 | GK | B.1.617.2 | 21 | Y | N | | |
| F | 30-39 | NY | EPI_ISL_3245528 | NYULH2416 | GK | AY.25 | 26 | N | N | | |
| F | 40-49 | NY | EPI_ISL_3245530 | NYULH2418 | GK | B.1.617.2 | 29 | Y | N | | |
| F | 30-39 | NY | EPI_ISL_3245533 | NYULH2421 | GK | B.1.617.2 | 19 | na | N | | |
| F | 70-79 | NY | EPI_ISL_3245535 | NYULH2423 | GK | B.1.617.2 | 18 | Y | N | | |
| M | 60-69 | NY | EPI_ISL_3245536 | NYULH2424 | GK | B.1.617.2 | 23 | Y | Y | Y | |
| F | <10 | NY | EPI_ISL_3245537 | NYULH2425 | GK | B.1.617.2 | NA | Y | N | | |
| M | 60-69 | NY | EPI_ISL_3245538 | NYULH2426 | GK | AY.14 | 30 | Y | N | | |
| F | 30-39 | NY | EPI_ISL_3245539 | NYULH2427 | GK | AY.20 | 18 | Y | N | | |
| M | 80-89 | NY | EPI_ISL_3245540 | NYULH2428 | GK | B.1.617.2 | 34 | Y | Y | N | N |
| F | 20-29 | NY | EPI_ISL_3245541 | NYULH2429 | GK | B.1.617.2 | 20 | Y | N | | |
| M | 20-29 | VA | EPI_ISL_3245543 | NYULH2431 | GK | B.1.617.2 | 20 | Y | Y | Y | |
| M | <10 | NY | EPI_ISL_3245545 | NYULH2433 | GK | AY.35 | 15 | Y | N | | |
| F | <10 | NY | EPI_ISL_3245546 | NYULH2434 | GK | B.1.617.2 | 20 | N | N | | |
| F | 90-99 | NY | EPI_ISL_3245547 | NYULH2435 | G | B | 40 | N | N | | |
| M | 30-39 | NY | EPI_ISL_3245548 | NYULH2436 | GK | B.1.617.2 | 20 | Y | N | | |
| F | 80-89 | NY | EPI_ISL_3245549 | NYULH2437 | G | AY.25 | 14 | Y | N | | |

| | | | | | | | | | | |
|---|---|---|---|---|---|---|---|---|---|---|
| F | 20-29 | NY | EPI_ISL_3245551 | NYULH2439 | GK | B.1.617.2 | 22 | Y | N | |
| F | <10 | NY | EPI_ISL_3245552 | NYULH2440 | GK | AY.16 | 19 | Y | N | |
| F | 50-59 | NJ | EPI_ISL_3245554 | NYULH2442 | G | B.1.617.2 | 33 | na | N | |
| F | <10 | NY | EPI_ISL_3245555 | NYULH2443 | GK | AY.14 | 28 | N | N | |
| M | <20 | NY | EPI_ISL_3245556 | NYULH2444 | GK | AY.14 | 27 | Y | N | |
| M | 80-89 | NY | EPI_ISL_3245557 | NYULH2445 | GK | AY.25 | 28 | Y | N | |
| F | 40-49 | NY | EPI_ISL_3245558 | NYULH2446 | GK | AY.35 | 16 | na | N | |
| M | <20 | NY | EPI_ISL_3245559 | NYULH2447 | GK | B.1.617.2 | 22 | Y | N | |
| M | 30-39 | NY | EPI_ISL_3245560 | NYULH2448 | GK | B.1.617.2 | 19 | na | N | |
| F | 20-29 | NY | EPI_ISL_3245561 | NYULH2449 | GK | B.1.617.2 | 20 | Y | N | |
| M | 20-29 | NY | EPI_ISL_3245565 | NYULH2453 | GK | B.1.617.2 | 28 | na | N | |
| M | 60-69 | NY | EPI_ISL_3245566 | NYULH2454 | GK | AY.3 | 18 | Y | N | |
| F | 30-39 | NY | EPI_ISL_3245567 | NYULH2455 | GV | AY.10 | 32 | na | N | |
| F | 20-29 | NY | EPI_ISL_3245569 | NYULH2457 | GK | B.1.617.2 | 22 | na | N | |
| M | 30-39 | NY | EPI_ISL_3245573 | NYULH2461 | GR | B.1.1.7 | 19 | na | N | |
| F | 40-49 | NY | EPI_ISL_3245577 | NYULH2465 | G | AY.33 | 34 | Y | N | |
| M | <20 | NY | EPI_ISL_3245578 | NYULH2466 | GK | AY.16 | 17 | Y | N | |
| F | 30-39 | NY | EPI_ISL_3245580 | NYULH2468 | GK | B.1.617.2 | 31 | na | N | |
| M | <20 | NY | EPI_ISL_3245581 | NYULH2469 | GV | AY.10 | 18 | Y | N | |
| F | <10 | NY | EPI_ISL_3245582 | NYULH2470 | GK | AY.14 | 29 | Y | N | |
| F | <10 | NY | EPI_ISL_3245583 | NYULH2471 | GK | AY.3 | 18 | N | N | |
| F | <10 | NY | EPI_ISL_3245584 | NYULH2472 | GK | AY.14 | 19 | Y | N | |
| F | 50-59 | NY | EPI_ISL_3245596 | NYULH2484 | GK | AY.20 | 28 | Y | Y | Y |
| M | 60-69 | NY | EPI_ISL_3245597 | NYULH2485 | GK | AY.3 | 21 | Y | Y | Y |
| F | 60-69 | NY | EPI_ISL_3245598 | NYULH2486 | GK | AY.3 | 21 | Y | Y | Y |
| F | 30-39 | NY | EPI_ISL_3245599 | NYULH2487 | GH | B.1.621 | 18 | Y | N | |

| | | | | | | | | | | |
|---|---|---|---|---|---|---|---|---|---|---|
| M | 50-59 | NY | EPI_ISL_3245600 | NYULH2488 | GK | AY.14 | NA | Y | Y | N |
| F | 50-59 | NY | EPI_ISL_3245601 | NYULH2489 | GK | B.1.617.2 | 28 | Y | N | |
| F | 50-59 | NY | EPI_ISL_3245602 | NYULH2490 | GK | B.1.617.2 | 18 | Y | Y | Y |
| F | 50-59 | NY | EPI_ISL_3245603 | NYULH2491 | GR | B.1.1.7 | 20 | Y | Y | Y |
| M | 30-39 | NY | EPI_ISL_3245604 | NYULH2492 | GH | B.1.621.1 | 22 | N | N | |
| M | 20-29 | NY | EPI_ISL_3245605 | NYULH2493 | GK | AY.14 | 17 | Y | N | |
| M | <10 | NY | EPI_ISL_3245606 | NYULH2494 | GK | B.1.617.2 | 22 | Y | N | |
| F | 20-29 | NY | EPI_ISL_3245607 | NYULH2495 | GK | AY.25 | 19 | Y | N | |
| M | 20-29 | NY | EPI_ISL_3245611 | NYULH2499 | GK | AY.16 | 15 | Y | N | |
| F | 30-39 | NY | EPI_ISL_3245612 | NYULH2500 | GK | AY.25 | 23 | Y | N | |
| M | 50-59 | NY | EPI_ISL_3245613 | NYULH2501 | GK | B.1.617.2 | 32 | Y | Y | Y |
| M | 30-39 | NY | EPI_ISL_3245614 | NYULH2502 | GR | P.1 | 26 | N | N | |
| M | 30-39 | NY | EPI_ISL_3245615 | NYULH2503 | GK | B.1.617.2 | 27 | Y | N | |
| F | 20-29 | NY | EPI_ISL_3245616 | NYULH2504 | GK | AY.25 | 17 | Y | N | |
| F | 20-29 | PA | EPI_ISL_3245617 | NYULH2505 | GK | B.1.617.2 | 29 | N | N | |
| F | <20 | NY | EPI_ISL_3245618 | NYULH2506 | GK | AY.16 | 16 | Y | N | |
| M | 50-59 | NY | EPI_ISL_3245619 | NYULH2507 | GK | B.1.617.2 | 19 | Y | Y | Y |
| F | 20-29 | NY | EPI_ISL_3245620 | NYULH2508 | GK | B.1.617.2 | 22 | Y | N | |
| M | 40-49 | NY | EPI_ISL_3245621 | NYULH2509 | G | B.1.628 | 26 | Y | Y | Y |
| M | <10 | NY | EPI_ISL_3245624 | NYULH2512 | GR | B.1.1.7 | 22 | Y | N | |
| M | 60-69 | NJ | EPI_ISL_3245625 | NYULH2513 | GK | B.1.617.2 | 23 | Y | N | |
| F | <20 | NY | EPI_ISL_3245626 | NYULH2514 | GK | AY.16 | 22 | Y | N | |
| M | 20-29 | NY | EPI_ISL_3245627 | NYULH2515 | GR | B.1.1.7 | 24 | Y | N | |
| M | 30-39 | NY | EPI_ISL_3245628 | NYULH2516 | GK | AY.20 | 23 | Y | Y | Y |
| M | 60-69 | NY | EPI_ISL_3245629 | NYULH2517 | GR | P.1 | 25 | Y | Y | Y |
| M | 40-49 | NY | EPI_ISL_3245630 | NYULH2518 | GK | AY.3 | 29 | Y | Y | Y |

| | | | | | | | | | | |
|---|---|---|---|---|---|---|---|---|---|---|
| F | 80-89 | NY | EPI_ISL_3245631 | NYULH2519 | GK | AY.3 | 23 | Y | N | |
| F | 20-29 | NY | EPI_ISL_3245632 | NYULH2520 | GR | P.1 | 20 | Y | N | |
| F | 30-39 | NY | EPI_ISL_3245633 | NYULH2521 | GK | B.1.617.2 | 35 | N | N | |
| M | 30-39 | NY | EPI_ISL_3245634 | NYULH2522 | GK | AY.14 | 18 | na | N | |
| F | 50-59 | NY | EPI_ISL_3245635 | NYULH2523 | GK | AY.25 | 25 | Y | N | |
| M | <10 | NY | EPI_ISL_3245636 | NYULH2524 | GK | AY.14 | 25 | N | N | |
| F | 30-39 | NY | EPI_ISL_3245637 | NYULH2525 | GK | B.1.617.2 | 31 | na | N | |
| M | 40-49 | NY | EPI_ISL_3245638 | NYULH2526 | GK | AY.14 | 21 | na | N | |
| F | <20 | NY | EPI_ISL_3245639 | NYULH2527 | GK | AY.26 | 30 | Y | N | |
| M | <20 | NY | EPI_ISL_3245640 | NYULH2528 | GK | AY.26 | 23 | Y | N | |
| M | 20-29 | NY | EPI_ISL_3245641 | NYULH2529 | GK | AY.26 | 29 | na | N | |
| F | 30-39 | NY | EPI_ISL_3245642 | NYULH2530 | GK | B.1.617.2 | 27 | na | N | |
| M | <20 | NY | EPI_ISL_3245643 | NYULH2531 | GK | AY.3 | 26 | Y | N | |
| M | 30-39 | NY | EPI_ISL_3245644 | NYULH2532 | GK | AY.16 | 19 | na | N | |
| M | 40-49 | NY | EPI_ISL_3245645 | NYULH2533 | GK | AY.14 | 15 | na | N | |
| F | 30-39 | NY | EPI_ISL_3245646 | NYULH2534 | GK | AY.25 | 32 | N | N | |
| F | 60-69 | NY | EPI_ISL_3245647 | NYULH2535 | GK | B.1.617.2 | 31 | na | N | |
| F | 40-49 | NJ | EPI_ISL_3245648 | NYULH2536 | GK | AY.3 | 31 | Y | N | |
| M | 40-49 | NJ | EPI_ISL_3245649 | NYULH2537 | GK | B.1.617.2 | 26 | na | N | |
| M | 70-79 | NY | EPI_ISL_3245664 | NYULH2561 | GH | B.1.621.1 | 23 | Y | Y | Y |
| F | 50-59 | NY | EPI_ISL_3245668 | NYULH2565 | GK | B.1.617.2 | 16 | Y | N | |
| F | 20-29 | NY | EPI_ISL_3245669 | NYULH2566 | GK | B.1.617.2 | 20 | N | N | |
| F | 60-69 | NY | EPI_ISL_3347407 | NYULH2568 | GK | AY.3 | 31 | Y | Y | Y |
| F | 50-59 | NY | EPI_ISL_3347408 | NYULH2569 | GK | B.1.617.2 | 23 | Y | Y | Y |
| M | 60-69 | NY | EPI_ISL_3347409 | NYULH2570 | G | AY.16 | 18 | Y | N | |
| F | 20-29 | NY | EPI_ISL_3347410 | NYULH2571 | G | AY.25 | 20 | Y | N | |

| | | | | | | | | | | |
|---|---|---|---|---|---|---|---|---|---|---|
| F | <20 | NY | EPI_ISL_3347411 | NYULH2572 | GK | B.1.617.2 | 27 | Y | N | |
| F | 40-49 | NY | EPI_ISL_3347412 | NYULH2573 | GK | AY.25 | 26 | Y | Y | Y |
| F | 30-39 | NY | EPI_ISL_3347413 | NYULH2574 | GV | AY.10 | 25 | Y | Y | Y |
| F | 20-29 | NY | EPI_ISL_3347414 | NYULH2575 | GV | AY.37 | 16 | Y | N | |
| M | <10 | NY | EPI_ISL_3347415 | NYULH2576 | GK | AY.25 | 24 | Y | N | |
| F | 80-89 | NY | EPI_ISL_3347416 | NYULH2577 | G | AY.25 | 16 | Y | Y | Y |
| M | 20-29 | NY | EPI_ISL_3347417 | NYULH2578 | GK | AY.25 | 29 | Y | Y | Y |
| F | 50-59 | NY | EPI_ISL_3347418 | NYULH2579 | GK | AY.16 | 29 | Y | Y | Y |
| F | 20-29 | NJ | EPI_ISL_3352387 | NYULH2580 | G | AY.16 | 23 | N | Y | N |
| F | <20 | NY | EPI_ISL_3347419 | NYULH2581 | G | B.1.617.2 | 21 | Y | N | |
| F | 20-29 | NJ | EPI_ISL_3347420 | NYULH2582 | GK | B.1.617.2 | 26 | Y | N | |
| F | 20-29 | NY | EPI_ISL_3347421 | NYULH2583 | GK | AY.25 | 29 | Y | N | |
| F | 20-29 | NY | EPI_ISL_3347422 | NYULH2584 | G | AY.16 | 16 | Y | Y | Y |
| F | 80-89 | NY | EPI_ISL_3347423 | NYULH2585 | G | AY.25 | 15 | Y | N | |
| F | 50-59 | NY | EPI_ISL_3347424 | NYULH2586 | GK | AY.3 | 24 | Y | N | |
| M | 20-29 | NY | EPI_ISL_3347425 | NYULH2587 | G | AY.3 | 33 | na | N | |
| F | 60-69 | NY | EPI_ISL_3347426 | NYULH2588 | GK | B.1.617.2 | 20 | Y | N | |
| F | 40-49 | NY | EPI_ISL_3347427 | NYULH2589 | GK | B.1.617.2 | 25 | Y | N | |
| F | 20-29 | NY | EPI_ISL_3347428 | NYULH2590 | G | AY.16 | 19 | na | N | |
| F | 30-39 | NY | EPI_ISL_3347429 | NYULH2591 | GK | AY.16 | 20 | na | N | |
| F | 30-39 | NY | EPI_ISL_3347430 | NYULH2592 | GK | AY.16 | 17 | na | N | |
| M | 30-39 | NY | EPI_ISL_3347431 | NYULH2593 | GV | AY.10 | 20 | na | N | |
| M | 70-79 | NY | EPI_ISL_3347436 | NYULH2598 | GK | B.1.617.2 | 23 | Y | N | |
| F | 20-29 | NY | EPI_ISL_3347437 | NYULH2599 | GK | AY.16 | 18 | Y | N | |
| M | 50-59 | NY | EPI_ISL_3347471 | NYULH2633 | GK | B.1.617.2 | 18 | Y | N | |
| F | 40-49 | NY | EPI_ISL_3347475 | NYULH2637 | GK | AY.3 | 25 | Y | N | |

Abbreviations: na: not available.

**Table S3. Metadata of GISAID reference sequences used in phylogenetic analyses.**

| strain | gisaid_epi_isl | date | region | Country | division |
|---|---|---|---|---|---|
| Afghanistan/IMB07964/2020 | EPI_ISL_1001001 | 5/30/2020 | Asia | Afghanistan | Afghanistan |
| Albania/211060667/2021 | EPI_ISL_1299884 | 3/11/2021 | Europe | Albania | Albania |
| Albania/un-ChVir25862_03/2021 | EPI_ISL_3123559 | 6/16/2021 | Europe | Albania | Albania |
| Albania/un-ChVir25862_04/2021 | EPI_ISL_3123560 | 6/16/2021 | Europe | Albania | Albania |
| Albania/un-ChVir25862_07/2021 | EPI_ISL_3123562 | 7/6/2021 | Europe | Albania | Albania |
| Albania/un-ChVir25862_15/2021 | EPI_ISL_3123565 | 7/9/2021 | Europe | Albania | Albania |
| Algeria/36420/2021 | EPI_ISL_3161807 | 5/10/2021 | Africa | Algeria | Ouargla |
| Algeria/36421/2021 | EPI_ISL_3161806 | 5/10/2021 | Africa | Algeria | Ouargla |
| Algeria/51743/2021 | EPI_ISL_3375632 | 6/30/2021 | Africa | Algeria | Algiers |
| Algeria/57433/2021 | EPI_ISL_3375633 | 7/13/2021 | Africa | Algeria | Algiers |
| Algeria/57453/2021 | EPI_ISL_3375634 | 7/13/2021 | Africa | Algeria | Algiers |
| Algeria/G0638_2264/2020 | EPI_ISL_418241 | 3/2/2020 | Africa | Algeria | Boufarik |
| Algeria/SMII/2021 | EPI_ISL_3161811 | 6/23/2021 | Africa | Algeria | Algiers |
| Andorra/AND-234_2118213840_GC/2021 | EPI_ISL_3384888 | 6/25/2021 | Europe | Andorra | Andorra |
| Andorra/AND-234_2120214126_GC/2021 | EPI_ISL_3384892 | 7/12/2021 | Europe | Andorra | Andorra |
| Andorra/AND-235_2118213847_GC/2021 | EPI_ISL_3384893 | 6/7/2021 | Europe | Andorra | Andorra |
| Andorre/AND-CHU-TLS-2120214126/2021 | EPI_ISL_3267140 | 7/12/2021 | Europe | Andorra | Andorra |
| Angola/CERI-KRISP-K013510/2020 | EPI_ISL_2492827 | 9/26/2020 | Africa | Angola | Luanda |
| Angola/CERI-KRISP-K013525/2020 | EPI_ISL_2492791 | 11/24/2020 | Africa | Angola | Luanda |
| Angola/CERI-KRISP-K014920/2021 | EPI_ISL_2492837 | 4/16/2021 | Africa | Angola | Luanda |
| Angola/CERI-KRISP-K014971/2021 | EPI_ISL_2492656 | 4/16/2021 | Africa | Angola | Luanda |
| Angola/CERI-KRISP-K015455/2021 | EPI_ISL_2617182 | 5/4/2021 | Africa | Angola | Luanda |
| Angola/CERI-KRISP-K015470/2021 | EPI_ISL_2617056 | 5/14/2021 | Africa | Angola | Luanda |
| Angola/KRISP-K009709/2020 | EPI_ISL_1347898 | 6/27/2020 | Africa | Angola | Luanda |
| Angola/KRISP-K010853/2021 | EPI_ISL_1545286 | 2/22/2021 | Africa | Angola | Luanda |

| | | | | | |
|---|---|---|---|---|---|
| AntiguaandBarbuda/73642/2021 | EPI_ISL_2955588 | 3/2/2021 | North America | Antigua and Barbuda | Antigua and Barbuda |
| AntiguaandBarbuda/74113/2021 | EPI_ISL_2716570 | 5/6/2021 | North America | Antigua and Barbuda | Antigua and Barbuda |
| AntiguaandBarbuda/74114/2021 | EPI_ISL_2716571 | 5/6/2021 | North America | Antigua and Barbuda | Antigua and Barbuda |
| Argentina/INEI104260/2021 | EPI_ISL_2140069 | 4/24/2021 | South America | Argentina | Argentina |
| Argentina/INEI104478/2021 | EPI_ISL_2140145 | 4/29/2021 | South America | Argentina | Argentina |
| Argentina/INEI104770/2021 | EPI_ISL_2158829 | 5/1/2021 | South America | Argentina | Buenos Aires |
| Argentina/PAIS-A0563/2021 | EPI_ISL_2007552 | 2/23/2021 | South America | Argentina | Buenos Aires |
| Argentina/PAIS-E0178/2020 | EPI_ISL_1395866 | 9/17/2020 | South America | Argentina | Santa Fe |
| Argentina/PAIS-G0327/2021 | EPI_ISL_3230017 | 7/27/2021 | South America | Argentina | Cordoba AR |
| Argentina/PAIS-G0328/2021 | EPI_ISL_3230018 | 7/27/2021 | South America | Argentina | Cordoba AR |
| Argentina/PAIS-G0335/2021 | EPI_ISL_3230025 | 5/12/2021 | South America | Argentina | Salta |
| Argentina/PAIS-J00025/2021 | EPI_ISL_3149723 | 6/1/2021 | South America | Argentina | Chaco |
| Argentina/PAIS-J00044/2021 | EPI_ISL_3149734 | 6/9/2021 | South America | Argentina | Chaco |
| Armenia/IMB2-12/2021 | EPI_ISL_1718293 | 1/27/2021 | Asia | Armenia | Armenia |
| Armenia/IMB3-10/2021 | EPI_ISL_1718303 | 3/18/2021 | Asia | Armenia | Armenia |
| Armenia/UW-ARM28/2020 | EPI_ISL_1854622 | 8/20/2020 | Asia | Armenia | Armenia |
| Aruba/AW-RIVM-10521/2021 | EPI_ISL_905171 | 1/13/2021 | South America | Aruba | Aruba |
| Aruba/AW-RIVM-11402/2021 | EPI_ISL_1014240 | 1/28/2021 | South America | Aruba | Aruba |
| Aruba/AW-RIVM-11427/2021 | EPI_ISL_1014649 | 1/28/2021 | South America | Aruba | Aruba |
| Aruba/AW-RIVM-27788/2021 | EPI_ISL_1792912 | 4/18/2021 | South America | Aruba | Aruba |

| | | | | | |
|---|---|---|---|---|---|
| Aruba/AW-RIVM-30306/2021 | EPI_ISL_2094338 | 4/25/2021 | South America | Aruba | Aruba |
| Aruba/AW-RIVM-38105/2021 | EPI_ISL_2476283 | 5/29/2021 | South America | Aruba | Aruba |
| Aruba/AW-RIVM-38116/2021 | EPI_ISL_2476285 | 5/29/2021 | South America | Aruba | Aruba |
| Aruba/AW-RIVM-39221/2021 | EPI_ISL_2610682 | 6/6/2021 | South America | Aruba | Aruba |
| Aruba/AW-RIVM-39232/2021 | EPI_ISL_2610693 | 6/2/2021 | South America | Aruba | Aruba |
| Aruba/AW-RIVM-46310/2021 | EPI_ISL_3138385 | 7/17/2021 | South America | Aruba | Aruba |
| Aruba/AW-RIVM-46322/2021 | EPI_ISL_3138397 | 7/17/2021 | South America | Aruba | Aruba |
| Australia/NSW-1796/2021 | EPI_ISL_2828057 | 6/26/2021 | Oceania | Australia | New South Wales |
| Australia/NSW-ICPMR-3145/2021 | EPI_ISL_3426413 | 8/4/2021 | Oceania | Australia | New South Wales |
| Australia/NSW-R0282/2021 | EPI_ISL_2249258 | 5/15/2021 | Oceania | Australia | New South Wales |
| Australia/NSW-R0378/2021 | EPI_ISL_2402459 | 5/24/2021 | Oceania | Australia | New South Wales |
| Australia/NSW-RPAH-0717/2021 | EPI_ISL_3280964 | 7/18/2021 | Oceania | Australia | New South Wales |
| Australia/NSW-RPAH-0841/2021 | EPI_ISL_3374203 | 7/24/2021 | Oceania | Australia | New South Wales |
| Australia/NSW-SAVID-5029/2021 | EPI_ISL_3418687 | 8/8/2021 | Oceania | Australia | New South Wales |
| Australia/NSW1706/2021 | EPI_ISL_2812853 | 6/25/2021 | Oceania | Australia | New South Wales |
| Australia/NSW2956/2020 | EPI_ISL_707895 | 10/20/2020 | Oceania | Australia | New South Wales |
| Australia/NT164/2021 | EPI_ISL_2250197 | 4/20/2021 | Oceania | Australia | Northern Territory |
| Australia/NT167/2021 | EPI_ISL_2250200 | 4/20/2021 | Oceania | Australia | Northern Territory |
| Australia/QLD1277/2020 | EPI_ISL_693278 | 3/21/2020 | Oceania | Australia | Queensland |
| Australia/QLD1526/2021 | EPI_ISL_849762 | 1/15/2021 | Oceania | Australia | Queensland |
| Australia/SA0498/2020 | EPI_ISL_602579 | 10/12/2020 | Oceania | Australia | South Australia |
| Australia/VIC877/2020 | EPI_ISL_427132 | 4/5/2020 | Oceania | Australia | Victoria |
| Austria/CeMM0370/2020 | EPI_ISL_583572 | 3/19/2020 | Europe | Austria | Tyrol |
| Austria/CeMM11086/2021 | EPI_ISL_2617643 | 5/13/2021 | Europe | Austria | Vorarlberg |
| Austria/CeMM12039/2021 | EPI_ISL_2887430 | 6/17/2021 | Europe | Austria | Austria |

| | | | | | |
|---|---|---|---|---|---|
| Austria/CeMM12189/2021 | EPI_ISL_2887563 | 6/17/2021 | Europe | Austria | Upper Austria |
| Austria/CeMM13427/2021 | EPI_ISL_3231076 | 7/14/2021 | Europe | Austria | Austria |
| Austria/CeMM13603/2021 | EPI_ISL_3231013 | 7/13/2021 | Europe | Austria | Austria |
| Austria/CeMM2077/2020 | EPI_ISL_1008372 | 12/30/2020 | Europe | Austria | Carinthia |
| Austria/CeMM2078/2020 | EPI_ISL_1008373 | 12/30/2020 | Europe | Austria | Carinthia |
| Austria/CeMM8487/2021 | EPI_ISL_2137284 | 4/24/2021 | Europe | Austria | Lower Austria |
| Austria/CeMM9203/2021 | EPI_ISL_2382045 | 4/22/2021 | Europe | Austria | Upper Austria |
| Austria/CeMM9585/2021 | EPI_ISL_2427228 | 5/17/2021 | Europe | Austria | Vienna |
| Austria/KILM1291/2021 | EPI_ISL_3394978 | 8/1/2021 | Europe | Austria | Vienna |
| Austria/MUW_1337238/2020 | EPI_ISL_1020297 | 9/18/2020 | Europe | Austria | Austria |
| Bahamas/33384/2021 | EPI_ISL_2982832 | 3/24/2021 | North America | Bahamas | Bahamas |
| Bahamas/33424/2021 | EPI_ISL_2982859 | 4/13/2021 | North America | Bahamas | Bahamas |
| Bahrain/260104364/2020 | EPI_ISL_682301 | 10/26/2020 | Asia | Bahrain | Bahrain |
| Bahrain/341933995/2021 | EPI_ISL_2715287 | 4/25/2021 | Asia | Bahrain | Bahrain |
| Bahrain/341935037/2021 | EPI_ISL_2715285 | 4/25/2021 | Asia | Bahrain | Bahrain |
| Bahrain/342039561/2021 | EPI_ISL_2932659 | 5/11/2021 | Asia | Bahrain | Bahrain |
| Bahrain/342066574/2021 | EPI_ISL_2932663 | 5/15/2021 | Asia | Bahrain | Bahrain |
| Bahrain/342375819/2021 | EPI_ISL_3462122 | 7/16/2021 | Asia | Bahrain | Bahrain |
| Bahrain/840046569/2021 | EPI_ISL_1660424 | 4/5/2021 | Asia | Bahrain | Bahrain |
| Bahrain/920858661/2021 | EPI_ISL_1660450 | 4/10/2021 | Asia | Bahrain | Bahrain |
| Bahrain/921080211/2021 | EPI_ISL_2932653 | 6/29/2021 | Asia | Bahrain | Bahrain |
| Bahrain/921080317/2021 | EPI_ISL_2932654 | 6/29/2021 | Asia | Bahrain | Bahrain |
| Bahrain/921149464/2021 | EPI_ISL_3462160 | 7/15/2021 | Asia | Bahrain | Bahrain |
| Bahrain/BAH-20/2020 | EPI_ISL_483561 | 4/8/2020 | Asia | Bahrain | Bahrain |
| Bangladesh/BCSIR-DU-45/2021 | EPI_ISL_3447342 | 7/6/2021 | Asia | Bangladesh | Dhaka |
| Bangladesh/BSMMU_Laila_90/2021 | EPI_ISL_2968089 | 6/29/2021 | Asia | Bangladesh | Rangpur |
| Bangladesh/CHRF-0011/2020 | EPI_ISL_492028 | 3/30/2020 | Asia | Bangladesh | Bangladesh |

| | | | | | |
|---|---|---|---|---|---|
| Bangladesh/CHRF-0368/2021 | EPI_ISL_3071080 | 6/19/2021 | Asia | Bangladesh | Dhaka |
| Bangladesh/CHRF-0389/2020 | EPI_ISL_2692876 | 8/15/2020 | Asia | Bangladesh | Rangpur |
| Bangladesh/CHRF-0392/2020 | EPI_ISL_2692878 | 9/29/2020 | Asia | Bangladesh | Dhaka |
| Bangladesh/CHRF-0500/2021 | EPI_ISL_3341984 | 7/17/2021 | Asia | Bangladesh | Dhaka |
| Bangladesh/DNAS_DRCIM_RAB_isl_82_BGD/2020 | EPI_ISL_774898 | 4/29/2020 | Asia | Bangladesh | Dhaka |
| Bangladesh/G-38/2020 | EPI_ISL_600562 | 3/26/2020 | Asia | Bangladesh | Dhaka |
| Bangladesh/icddrb-1210504010/2021 | EPI_ISL_2627387 | 5/6/2021 | Asia | Bangladesh | Rajshahi |
| Bangladesh/icddrb-1210509020/2021 | EPI_ISL_2627361 | 5/29/2021 | Asia | Bangladesh | Khulna |
| Bangladesh/ideSHi-S-04/2021 | EPI_ISL_1750957 | 2/8/2021 | Asia | Bangladesh | Dhaka |
| Bangladesh/IEDCR-OIS-172/2021 | EPI_ISL_2600373 | 4/22/2021 | Asia | Bangladesh | Chattogram |
| Bangladesh/JS004/2021 | EPI_ISL_1938476 | 4/28/2021 | Asia | Bangladesh | Barishal |
| Barbados/59721/2021 | EPI_ISL_2544758 | 3/16/2021 | North America | Barbados | Barbados |
| Barbados/63454/2021 | EPI_ISL_2545191 | 4/8/2021 | North America | Barbados | Barbados |
| Barbados/63485/2021 | EPI_ISL_2545194 | 4/10/2021 | North America | Barbados | Barbados |
| Barbados/67294/2021 | EPI_ISL_2621681 | 5/4/2021 | North America | Barbados | Barbados |
| Barbados/67697/2021 | EPI_ISL_2621679 | 4/25/2021 | North America | Barbados | Barbados |
| Barbados/71159/2021 | EPI_ISL_2678169 | 5/4/2021 | North America | Barbados | Barbados |
| Barbados/76426/2021 | EPI_ISL_2967991 | 7/3/2021 | North America | Barbados | Barbados |
| Barbados/76427/2021 | EPI_ISL_2967992 | 7/3/2021 | North America | Barbados | Barbados |
| Barbados/BRB50400/2020 | EPI_ISL_977658 | 12/31/2020 | North America | Barbados | Barbados |
| Belgium/AZDelta-04536-2128R/2021 | EPI_ISL_3047403 | 7/15/2021 | Europe | Belgium | Oudenaarde |
| Belgium/Jessa_55-2132-000518/2021 | EPI_ISL_3455440 | 8/11/2021 | Europe | Belgium | Limburg BE |
| Belgium/MBLGPF351322/2021 | EPI_ISL_2885660 | 6/29/2021 | Europe | Belgium | Brussels |
| Belgium/MBLGPF837872/2021 | EPI_ISL_3133105 | 7/18/2021 | Europe | Belgium | Brussels |

| | | | | | |
|---|---|---|---|---|---|
| Belgium/rega-10475/2021 | EPI_ISL_2864752 | 5/19/2021 | Europe | Belgium | Brussels |
| Belgium/rega-10633/2021 | EPI_ISL_2877570 | 6/4/2021 | Europe | Belgium | Brussels |
| Belgium/rega-9224/2021 | EPI_ISL_2725900 | 5/3/2021 | Europe | Belgium | Hasselt |
| Belgium/UGent-205/2020 | EPI_ISL_717588 | 10/22/2020 | Europe | Belgium | Gent |
| Belgium/UGent-6399/2021 | EPI_ISL_2192525 | 4/28/2021 | Europe | Belgium | Gent |
| Belgium/ULG-10241/2020 | EPI_ISL_498147 | 6/30/2020 | Europe | Belgium | Liv®ge |
| Belgium/ULG-11528/2020 | EPI_ISL_925472 | 10/12/2020 | Europe | Belgium | Liv®ge |
| Belgium/UZA-UA-CV2313801018/2021 | EPI_ISL_3320550 | 8/3/2021 | Europe | Belgium | Mechelen |
| Belgium/UZA-UA-THX567/2021 | EPI_ISL_2016820 | 4/30/2021 | Europe | Belgium | Ieper |
| Belize/BZ-CML-TCMC-BZ032-1220/2020 | EPI_ISL_1822780 | 12/27/2020 | North America | Belize | Belize |
| Belize/CDC-6846/2020 | EPI_ISL_509712 | 3/1/2020 | North America | Belize | Belmopan |
| Benin/254242/2021 | EPI_ISL_2932558 | 2/3/2021 | Africa | Benin | Cotonou |
| Benin/260/2020 | EPI_ISL_476823 | 3/17/2020 | Africa | Benin | Cotonou |
| Bolivia/26933/2020 | EPI_ISL_837556 | 7/15/2020 | South America | Bolivia | Santa Cruz BO |
| Bolivia/26954/2020 | EPI_ISL_837572 | 3/11/2020 | South America | Bolivia | Oruro |
| Bolivia/26955/2020 | EPI_ISL_837573 | 4/20/2020 | South America | Bolivia | Tarija |
| Bolivia/LaPaz_GM3-4/2021 | EPI_ISL_2802859 | 6/2/2021 | South America | Bolivia | La Paz BO |
| Bolivia/LaPaz_GM3-5/2021 | EPI_ISL_2802860 | 6/2/2021 | South America | Bolivia | La Paz BO |
| Bolivia/LPZ-4/2021 | EPI_ISL_1363787 | 1/28/2021 | South America | Bolivia | La Paz BO |
| Bolivia/Sucre_GM2-1/2021 | EPI_ISL_2600378 | 5/1/2021 | South America | Bolivia | Chuquisaca |
| Bolivia/Sucre-GM2-4/2021 | EPI_ISL_2462063 | 5/10/2021 | South America | Bolivia | Chuquisaca |
| Bonaire/BQ-RIVM-27386/2021 | EPI_ISL_1792360 | 4/8/2021 | South America | Bonaire | Bonaire |
| Bonaire/BQ-RIVM-35139/2021 | EPI_ISL_2405596 | 5/7/2021 | South America | Bonaire | Bonaire |

| | | | | | |
|---|---|---|---|---|---|
| Bonaire/BQ-RIVM-39010/2021 | EPI_ISL_2610471 | 5/27/2021 | South America | Bonaire | Bonaire |
| Bonaire/BQ-RIVM-42355/2021 | EPI_ISL_2981896 | 6/26/2021 | South America | Bonaire | Bonaire |
| Bonaire/BQ-RIVM-42398/2021 | EPI_ISL_2981901 | 6/25/2021 | South America | Bonaire | Bonaire |
| Bonaire/BQ-RIVM-43226/2021 | EPI_ISL_3056849 | 7/1/2021 | South America | Bonaire | Bonaire |
| Bonaire/BQ-RIVM-45034/2021 | EPI_ISL_3137106 | 7/7/2021 | South America | Bonaire | Bonaire |
| BosniaandHerzegovina/04-Sarajevo/2020 | EPI_ISL_467300 | 4/8/2020 | Europe | Bosnia and Herzegovina | Sarajevo |
| BosniaandHerzegovina/AGC-MOS-002/2020 | EPI_ISL_722201 | 11/20/2020 | Europe | Bosnia and Herzegovina | Mostar |
| BosniaandHerzegovina/AGCOB6080/2021 | EPI_ISL_3184312 | 7/22/2021 | Europe | Bosnia and Herzegovina | Sarajevo |
| BosniaandHerzegovina/Alea-02/2021 | EPI_ISL_922076 | 1/10/2021 | Europe | Bosnia and Herzegovina | Sarajevo |
| BosniaandHerzegovina/ChVir7361/2020 | EPI_ISL_462464 | 3/25/2020 | Europe | Bosnia and Herzegovina | Ilidza |
| BosniaandHerzegovina/EL281/2021 | EPI_ISL_2029260 | 5/4/2021 | Europe | Bosnia and Herzegovina | Tuzla |
| BosniaandHerzegovina/KCUS70511/2021 | EPI_ISL_2938118 | 6/6/2021 | Europe | Bosnia and Herzegovina | Sarajevo |
| BosniaandHerzegovina/KCUS71429/2021 | EPI_ISL_2938102 | 6/10/2021 | Europe | Bosnia and Herzegovina | Sarajevo |
| BosniaandHerzegovina/VFS-UNSA-LMGFI006/2020 | EPI_ISL_955148 | 9/14/2020 | Europe | Bosnia and Herzegovina | Sarajevo |
| BosniaandHerzegovina/VFS-UNSA-LMGFI038/2021 | EPI_ISL_3076939 | 7/5/2021 | Europe | Bosnia and Herzegovina | Mostar |
| Botswana/BOT324710/2020 | EPI_ISL_1677719 | 11/21/2020 | Africa | Botswana | Central District BW |
| Botswana/R14B10_BHP_AAB41824/2021 | EPI_ISL_2386154 | 5/14/2021 | Africa | Botswana | Mochudi |
| Botswana/R14B25_BHP_b59_121047451/2021 | EPI_ISL_2566207 | 4/28/2021 | Africa | Botswana | Gaborone |
| Botswana/R14B27_BHP_b61_121047439/2021 | EPI_ISL_2566209 | 4/28/2021 | Africa | Botswana | Gaborone |
| Botswana/R14B3_BHP_AAB39499/2021 | EPI_ISL_2153476 | 5/10/2021 | Africa | Botswana | Gaborone |
| Botswana/R19B80_BHP_000534343/2021 | EPI_ISL_2820368 | 6/24/2021 | Africa | Botswana | Lobatse |
| Botswana/R20B62_BHP_AAB73650/2021 | EPI_ISL_2820436 | 6/26/2021 | Africa | Botswana | Lobatse |

| | | | | | |
|---|---|---|---|---|---|
| Botswana/R21B32_BHP_AAB79215/2021 | EPI_ISL_2868356 | 7/4/2021 | Africa | Botswana | Gaborone |
| Botswana/R23B42_BHP_AAB89691/2021 | EPI_ISL_3162270 | 7/13/2021 | Africa | Botswana | Gaborone |
| Botswana/R23B88_BHP_AAB117692/2021 | EPI_ISL_3453916 | 8/10/2021 | Africa | Botswana | Gaborone |
| Botswana/R23B89_BHP_AAB117724/2021 | EPI_ISL_3453917 | 8/10/2021 | Africa | Botswana | Gaborone |
| Botswana/R8B55_BHP_AAA24317/2020 | EPI_ISL_1516868 | 9/27/2020 | Africa | Botswana | Gaborone |
| Brazil/AM-FIOCRUZ-20143297CR/2020 | EPI_ISL_1068114 | 12/23/2020 | South America | Brazil | Amazonas BR |
| Brazil/PR-L118-CD7518/2020 | EPI_ISL_2344284 | 9/17/2020 | South America | Brazil | Paranv° |
| Brazil/RJ-LNN01167/2021 | EPI_ISL_2385461 | 4/28/2021 | South America | Brazil | Rio de Janeiro |
| Brazil/RJ-LNN01255/2021 | EPI_ISL_2385535 | 5/4/2021 | South America | Brazil | Rio de Janeiro |
| Brazil/SC-FIOCRUZ-31633/2021 | EPI_ISL_2983412 | 6/8/2021 | South America | Brazil | Santa Catarina |
| Brazil/SC-FIOCRUZ-34948/2021 | EPI_ISL_3190222 | 6/28/2021 | South America | Brazil | Santa Catarina |
| Brazil/SP-BT6799/2020 | EPI_ISL_875549 | 5/9/2020 | South America | Brazil | Sv£o Paulo |
| Brazil/SP-HIAE-ID363/2021 | EPI_ISL_3246175 | 7/26/2021 | South America | Brazil | Sv£o Paulo |
| Brazil/SP-HIAE-ID378/2021 | EPI_ISL_3246190 | 7/26/2021 | South America | Brazil | Sv£o Paulo |
| Brazil/SP-HIAE-ID570/2021 | EPI_ISL_3385131 | 8/3/2021 | South America | Brazil | Sv£o Paulo |
| Brazil/SP-HIAE-ID590/2021 | EPI_ISL_3385154 | 8/4/2021 | South America | Brazil | Sv£o Paulo |
| Brazil/SP-IB_100886/2021 | EPI_ISL_1966870 | 4/4/2021 | South America | Brazil | Sv£o Paulo |
| Brazil/SP-IB_101387/2021 | EPI_ISL_1967309 | 4/22/2021 | South America | Brazil | Sv£o Paulo |
| Brazil/SP-IB_102693/2021 | EPI_ISL_2345714 | 5/3/2021 | South America | Brazil | Sv£o Paulo |
| Bulgaria/21BG-EU_000677_Pl7/2021 | EPI_ISL_1401161 | 2/28/2021 | Europe | Bulgaria | Sofia |
| Bulgaria/21BG-EU_002819_Pl25/2021 | EPI_ISL_2621274 | 4/23/2021 | Europe | Bulgaria | Pernik |
| Bulgaria/21BG-EU_002893_Pl26/2021 | EPI_ISL_2621342 | 4/26/2021 | Europe | Bulgaria | Sofia |

| | | | | | |
|---|---|---|---|---|---|
| Bulgaria/21BG-EU_003022_Pl27/2021 | EPI_ISL_2648059 | 5/1/2021 | Europe | Bulgaria | Sofia |
| Bulgaria/21BG-EU_003059_Pl28/2021 | EPI_ISL_2621478 | 5/7/2021 | Europe | Bulgaria | Gabrovo |
| Bulgaria/21BG-EU_003790_Pl32/2021 | EPI_ISL_3076806 | 6/24/2021 | Europe | Bulgaria | Dobrich |
| Bulgaria/21BG-EU_003875_Pl33/2021 | EPI_ISL_3232693 | 7/12/2021 | Europe | Bulgaria | Sofia |
| Bulgaria/21BG-EU_003876_Pl33/2021 | EPI_ISL_3232694 | 7/12/2021 | Europe | Bulgaria | Sofia |
| Bulgaria/21BG-NC_003577_R14/2021 | EPI_ISL_2967173 | 6/22/2021 | Europe | Bulgaria | Sofia |
| Bulgaria/21BG-NC_A000009/2020 | EPI_ISL_2081878 | 7/8/2020 | Europe | Bulgaria | Stara Zagora |
| Bulgaria/21BG-NC_A000048/2020 | EPI_ISL_2081915 | 8/7/2020 | Europe | Bulgaria | Kyustendil |
| Bulgaria/21BG-NC_A000072/2020 | EPI_ISL_2081935 | 10/28/2020 | Europe | Bulgaria | Sofia |
| Bulgaria/21BG-NC_A000074/2020 | EPI_ISL_2081937 | 11/8/2020 | Europe | Bulgaria | Kyustendil |
| BurkinaFaso/2-052-1891/2020 | EPI_ISL_660527 | 8/20/2020 | Africa | Burkina Faso | Hauts-Bassins |
| BurkinaFaso/2550/2020 | EPI_ISL_660498 | 9/13/2020 | Africa | Burkina Faso | Hauts-Bassins |
| Burundi/UG521/2021 | EPI_ISL_2928005 | 5/30/2021 | Africa | Burundi | Burundi |
| Burundi/UG558/2021 | EPI_ISL_2928012 | 5/31/2021 | Africa | Burundi | Burundi |
| CaboVerde/Mindelo_04224/2021 | EPI_ISL_1711656 | 2/2/2021 | Africa | Cabo Verde | Nossa Senhora da Luz |
| CaboVerde/Praia_30215/2020 | EPI_ISL_1711641 | 10/4/2020 | Africa | Cabo Verde | Nossa Senhora da Gravßa |
| Cambodia/01-2106231496/2021 | EPI_ISL_2801893 | 6/23/2021 | Asia | Cambodia | Cambodia |
| Cambodia/01-2107205906/2021 | EPI_ISL_3118368 | 7/20/2021 | Asia | Cambodia | Cambodia |
| Cambodia/101372/2020 | EPI_ISL_1040028 | 10/27/2020 | Asia | Cambodia | Cambodia |
| Cambodia/22-2105242017/2021 | EPI_ISL_2406461 | 5/25/2021 | Asia | Cambodia | Cambodia |
| Cambodia/26-2106140245/2021 | EPI_ISL_3031504 | 6/27/2021 | Asia | Cambodia | Cambodia |
| Cambodia/409652/2021 | EPI_ISL_1711994 | 4/17/2021 | Asia | Cambodia | Cambodia |
| Cambodia/441923/2021 | EPI_ISL_2106244 | 4/29/2021 | Asia | Cambodia | Cambodia |
| Cambodia/465939/2021 | EPI_ISL_2106255 | 5/7/2021 | Asia | Cambodia | Cambodia |
| Cambodia/662600/2021 | EPI_ISL_3374594 | 7/25/2021 | Asia | Cambodia | Cambodia |
| Cambodia/709186/2021 | EPI_ISL_3387378 | 8/8/2021 | Asia | Cambodia | Cambodia |
| Cambodia/709189/2021 | EPI_ISL_3387379 | 8/8/2021 | Asia | Cambodia | Cambodia |

| | | | | | |
|---|---|---|---|---|---|
| Cambodia/Kunming_kms-2/2020 | EPI_ISL_682298 | 3/21/2020 | Asia | Cambodia | Phnom Penh |
| Cambodia/VIR2107864/2021 | EPI_ISL_1096139 | 1/20/2021 | Asia | Cambodia | Cambodia |
| Cameroon/Douala-20V-3045/2020 | EPI_ISL_1001051 | 3/26/2020 | Africa | Cameroon | Douala |
| Cameroon/ICGEB-CIRCB_10136/2021 | EPI_ISL_2504131 | 10/29/2020 | Africa | Cameroon | Yaoundé |
| Cameroon/ICGEB-CIRCB_26438/2021 | EPI_ISL_2504142 | 4/17/2021 | Africa | Cameroon | Yaoundé |
| Cameroon/Yaounde-20V-6189/2020 | EPI_ISL_1001043 | 4/13/2020 | Africa | Cameroon | Yaoundé |
| Canada/AB-12344/2020 | EPI_ISL_854468 | 3/26/2020 | North America | Canada | Alberta |
| Canada/AB-41922/2020 | EPI_ISL_806393 | 5/4/2020 | North America | Canada | Alberta |
| Canada/AB-ABPHL-17252/2021 | EPI_ISL_2479827 | 4/23/2021 | North America | Canada | Alberta |
| Canada/AB-ABPHL-18261/2021 | EPI_ISL_2940933 | 5/9/2021 | North America | Canada | Alberta |
| Canada/AB-ABPHL-19003/2021 | EPI_ISL_2941355 | 4/20/2021 | North America | Canada | Alberta |
| Canada/AB-ABPHL-19243/2021 | EPI_ISL_2940468 | 5/11/2021 | North America | Canada | Alberta |
| Canada/AB-ABPHL-22863/2021 | EPI_ISL_3116343 | 6/17/2021 | North America | Canada | Alberta |
| Canada/AB-ABPHL-23267/2021 | EPI_ISL_3116498 | 6/22/2021 | North America | Canada | Alberta |
| Canada/AB-ABPHL-23495/2021 | EPI_ISL_3116454 | 7/1/2021 | North America | Canada | Alberta |
| Canada/AB-ABPHL-24097/2021 | EPI_ISL_3116483 | 7/6/2021 | North America | Canada | Alberta |
| Canada/ABPHL-06226/2021 | EPI_ISL_2165370 | 10/28/2020 | North America | Canada | Alberta |
| Canada/BC-BCCDC-3575/2020 | EPI_ISL_968269 | 6/26/2020 | North America | Canada | British Columbia |
| Canada/BC-BCCDC-4764/2020 | EPI_ISL_968546 | 8/8/2020 | North America | Canada | British Columbia |
| Canada/BC-BCCDC-4844/2020 | EPI_ISL_973504 | 9/21/2020 | North America | Canada | British Columbia |
| Canada/BC-BCCDC-57660/2021 | EPI_ISL_2653437 | 4/1/2021 | North America | Canada | British Columbia |

| | | | | | |
|---|---|---|---|---|---|
| Canada/BC-BCCDC-6545/2020 | EPI_ISL_974581 | 10/22/2020 | North America | Canada | British Columbia |
| Canada/BC-BCCDC-74790/2021 | EPI_ISL_2655128 | 4/16/2021 | North America | Canada | British Columbia |
| Canada/BC-BCCDC-76765/2021 | EPI_ISL_2656351 | 4/18/2021 | North America | Canada | British Columbia |
| Canada/BC-BCCDC-78788/2021 | EPI_ISL_2720773 | 5/3/2021 | North America | Canada | British Columbia |
| Canada/BC-BCCDC-79199/2021 | EPI_ISL_2721113 | 3/30/2021 | North America | Canada | British Columbia |
| Canada/BC-BCCDC-86966/2021 | EPI_ISL_2721411 | 5/4/2021 | North America | Canada | British Columbia |
| Canada/MB-NML-1122/2020 | EPI_ISL_582457 | 8/3/2020 | North America | Canada | Manitoba |
| Canada/MB-NML-1533/2020 | EPI_ISL_935834 | 9/16/2020 | North America | Canada | Manitoba |
| Canada/MB-NML-1553/2020 | EPI_ISL_935849 | 9/20/2020 | North America | Canada | Manitoba |
| Canada/MB-NML-16315/2021 | EPI_ISL_1258359 | 1/15/2021 | North America | Canada | Manitoba |
| Canada/MB-NML-17457/2021 | EPI_ISL_1594202 | 2/12/2021 | North America | Canada | Manitoba |
| Canada/MB-NML-17747/2021 | EPI_ISL_1594107 | 2/21/2021 | North America | Canada | Manitoba |
| Canada/MB-NML-2694/2020 | EPI_ISL_1258142 | 11/6/2020 | North America | Canada | Manitoba |
| Canada/MB-NML-565/2020 | EPI_ISL_582263 | 5/3/2020 | North America | Canada | Manitoba |
| Canada/MB-NML-606/2020 | EPI_ISL_632912 | 3/28/2020 | North America | Canada | Manitoba |
| Canada/MB-NML-70495/2021 | EPI_ISL_2495667 | 4/21/2021 | North America | Canada | Manitoba |
| Canada/MB-NML-70522/2021 | EPI_ISL_2495574 | 4/21/2021 | North America | Canada | Manitoba |
| Canada/MB-NML-794/2020 | EPI_ISL_582313 | 5/30/2020 | North America | Canada | Manitoba |
| Canada/MB-NML-830/2020 | EPI_ISL_582325 | 4/21/2020 | North America | Canada | Manitoba |
| Canada/MB-NML-94818/2021 | EPI_ISL_3356838 | 6/7/2021 | North America | Canada | Manitoba |

| | | | | | |
|---|---|---|---|---|---|
| Canada/MB-NML-94863/2021 | EPI_ISL_3356854 | 6/3/2021 | North America | Canada | Manitoba |
| Canada/MB-NML-95124/2021 | EPI_ISL_3356884 | 5/24/2021 | North America | Canada | Manitoba |
| Canada/MB-NML-95142/2021 | EPI_ISL_3356900 | 5/25/2021 | North America | Canada | Manitoba |
| Canada/NB-NML-139497/2021 | EPI_ISL_3334211 | 7/21/2021 | North America | Canada | New Brunswick |
| Canada/NB-NML-139499/2021 | EPI_ISL_3334213 | 7/22/2021 | North America | Canada | New Brunswick |
| Canada/NB-NML-16626/2021 | EPI_ISL_1272271 | 2/4/2021 | North America | Canada | New Brunswick |
| Canada/NB-NML-17262/2021 | EPI_ISL_1588066 | 2/15/2021 | North America | Canada | New Brunswick |
| Canada/NB-NML-3145/2020 | EPI_ISL_961667 | 11/18/2020 | North America | Canada | New Brunswick |
| Canada/NB-NML-3272/2020 | EPI_ISL_961578 | 12/29/2020 | North America | Canada | New Brunswick |
| Canada/NB-NML-3293/2021 | EPI_ISL_961566 | 1/2/2021 | North America | Canada | New Brunswick |
| Canada/NB-NML-43559/2021 | EPI_ISL_2550593 | 4/17/2021 | North America | Canada | New Brunswick |
| Canada/NB-NML-49173/2021 | EPI_ISL_3334216 | 4/21/2021 | North America | Canada | New Brunswick |
| Canada/NL-NML-16825/2021 | EPI_ISL_1278117 | 2/12/2021 | North America | Canada | Newfoundland and Labrador |
| Canada/NL-NML-20656/2021 | EPI_ISL_1587903 | 3/7/2021 | North America | Canada | Newfoundland and Labrador |
| Canada/NL-NML-2419/2020 | EPI_ISL_1055408 | 8/27/2020 | North America | Canada | Newfoundland and Labrador |
| Canada/NL-NML-43020/2021 | EPI_ISL_2161794 | 4/16/2021 | North America | Canada | Newfoundland and Labrador |
| Canada/NL-NML-55193/2021 | EPI_ISL_2161800 | 4/19/2021 | North America | Canada | Newfoundland and Labrador |
| Canada/NL-NML-62012/2021 | EPI_ISL_2463835 | 4/27/2021 | North America | Canada | Newfoundland and Labrador |
| Canada/NL-NML-85345/2021 | EPI_ISL_2484616 | 5/14/2021 | North America | Canada | Newfoundland and Labrador |
| Canada/NL-NML-85351/2021 | EPI_ISL_2484592 | 5/17/2021 | North America | Canada | Newfoundland and Labrador |

| | | | | | |
|---|---|---|---|---|---|
| Canada/NL-NML-97345/2021 | EPI_ISL_3150969 | 6/6/2021 | North America | Canada | Newfoundland and Labrador |
| Canada/NL-NML-97351/2021 | EPI_ISL_3150975 | 6/8/2021 | North America | Canada | Newfoundland and Labrador |
| Canada/NS-NML-117307/2021 | EPI_ISL_3399741 | 5/24/2021 | North America | Canada | Nova Scotia |
| Canada/NS-NML-117312/2021 | EPI_ISL_3399729 | 6/11/2021 | North America | Canada | Nova Scotia |
| Canada/NS-NML-117332/2021 | EPI_ISL_3399727 | 6/14/2021 | North America | Canada | Nova Scotia |
| Canada/NS-NML-2075/2020 | EPI_ISL_915107 | 4/21/2020 | North America | Canada | Nova Scotia |
| Canada/NS-NML-2282/2020 | EPI_ISL_1666853 | 5/14/2020 | North America | Canada | Nova Scotia |
| Canada/NS-NML-48376/2021 | EPI_ISL_2162070 | 4/16/2021 | North America | Canada | Nova Scotia |
| Canada/NS-NML-5352/2020 | EPI_ISL_1055552 | 11/26/2020 | North America | Canada | Nova Scotia |
| Canada/NS-NML-5373/2020 | EPI_ISL_1055569 | 11/23/2020 | North America | Canada | Nova Scotia |
| Canada/NS-NML-5382/2020 | EPI_ISL_1055576 | 11/22/2020 | North America | Canada | Nova Scotia |
| Canada/NS-NML-5515/2020 | EPI_ISL_1055681 | 8/21/2020 | North America | Canada | Nova Scotia |
| Canada/NS-NML-5530/2020 | EPI_ISL_1055691 | 6/29/2020 | North America | Canada | Nova Scotia |
| Canada/NS-NML-5531/2020 | EPI_ISL_1055692 | 6/29/2020 | North America | Canada | Nova Scotia |
| Canada/NS-NML-94527/2021 | EPI_ISL_2835588 | 5/11/2021 | North America | Canada | Nova Scotia |
| Canada/NS-NML-94551/2021 | EPI_ISL_2835606 | 4/28/2021 | North America | Canada | Nova Scotia |
| Canada/ON-PPS-00393/2020 | EPI_ISL_853586 | 11/24/2020 | North America | Canada | Ontario |
| Canada/ON-S188/2020 | EPI_ISL_586375 | 3/25/2020 | North America | Canada | Ontario |
| Canada/ON-SC0238/2020 | EPI_ISL_876610 | 8/18/2020 | North America | Canada | Ontario |
| Canada/ON-SC4324/2020 | EPI_ISL_1739981 | 12/21/2020 | North America | Canada | Ontario |

| | | | | | |
|---|---|---|---|---|---|
| Canada/ON-SC5396/2021 | EPI_ISL_1740411 | 2/15/2021 | North America | Canada | Ontario |
| Canada/ON-UHTC_0528/2021 | EPI_ISL_2824507 | 3/2/2021 | North America | Canada | Ontario |
| Canada/ON-UHTC-0171/2020 | EPI_ISL_569959 | 5/18/2020 | North America | Canada | Ontario |
| Canada/QC-1nIOU-A6155288/2021 | EPI_ISL_2824104 | 2/15/2021 | North America | Canada | Quebec |
| Canada/QC-1nIPG-Q4090758R2/2020 | EPI_ISL_2386125 | 12/9/2020 | North America | Canada | Quebec |
| Canada/Qc-CHUM-2017802870a/2020 | EPI_ISL_825909 | 6/26/2020 | North America | Canada | Quebec |
| Canada/Qc-HCLM-0081235810/2020 | EPI_ISL_825753 | 8/7/2020 | North America | Canada | Quebec |
| Canada/QC-HMR-96121022/2020 | EPI_ISL_954366 | 4/12/2020 | North America | Canada | Quebec |
| Canada/Qc-HVE-Q1240849R2/2020 | EPI_ISL_1403285 | 9/24/2020 | North America | Canada | Quebec |
| Canada/Qc-L00275237/2020 | EPI_ISL_826122 | 7/22/2020 | North America | Canada | Quebec |
| Canada/Qc-L00292782/2020 | EPI_ISL_1365933 | 9/25/2020 | North America | Canada | Quebec |
| Canada/QC-L00340993001/2021 | EPI_ISL_3144143 | 3/25/2021 | North America | Canada | Quebec |
| Canada/QC-L00344463/2021 | EPI_ISL_3458348 | 4/7/2021 | North America | Canada | Quebec |
| Canada/QC-L00350254001/2021 | EPI_ISL_2970551 | 4/29/2021 | North America | Canada | Quebec |
| Canada/QC-L00352714/2021 | EPI_ISL_3458635 | 5/10/2021 | North America | Canada | Quebec |
| Canada/QC-L00352948/2021 | EPI_ISL_3458652 | 4/24/2021 | North America | Canada | Quebec |
| Canada/QC-L00357248/2021 | EPI_ISL_2989552 | 5/25/2021 | North America | Canada | Quebec |
| Canada/QC-L00358252/2021 | EPI_ISL_3458854 | 6/3/2021 | North America | Canada | Quebec |
| Canada/QC-L00360857001/2021 | EPI_ISL_3208040 | 6/14/2021 | North America | Canada | Quebec |
| Canada/QC-L00364808001/2021 | EPI_ISL_3459102 | 7/5/2021 | North America | Canada | Quebec |

| | | | | | |
|---|---|---|---|---|---|
| Canada/QC-L00366817001/2021 | EPI_ISL_3459139 | 7/11/2021 | North America | Canada | Quebec |
| Canada/Qc-LSPQ-L00210314/2020 | EPI_ISL_535716 | 2/25/2020 | North America | Canada | Quebec |
| Canada/Qc-LSPQ-L00233380/2020 | EPI_ISL_536021 | 3/23/2020 | North America | Canada | Quebec |
| Canada/Qc-LSPQ-L00233661/2020 | EPI_ISL_536031 | 3/23/2020 | North America | Canada | Quebec |
| Canada/SK-NML-109503/2021 | EPI_ISL_3255708 | 6/16/2021 | North America | Canada | Saskatchewan |
| Canada/SK-NML-17203/2020 | EPI_ISL_2883380 | 11/4/2020 | North America | Canada | Saskatchewan |
| Canada/SK-NML-17324/2020 | EPI_ISL_2883420 | 7/16/2020 | North America | Canada | Saskatchewan |
| Canada/SK-NML-2724/2020 | EPI_ISL_2883599 | 10/26/2020 | North America | Canada | Saskatchewan |
| Canada/SK-NML-3132/2020 | EPI_ISL_2883604 | 9/5/2020 | North America | Canada | Saskatchewan |
| Canada/SK-NML-492/2020 | EPI_ISL_2562954 | 4/28/2020 | North America | Canada | Saskatchewan |
| Canada/SK-NML-55354/2021 | EPI_ISL_2564719 | 4/19/2021 | North America | Canada | Saskatchewan |
| Canada/SK-NML-715/2020 | EPI_ISL_2883720 | 6/16/2020 | North America | Canada | Saskatchewan |
| Canada/SK-NML-72068/2021 | EPI_ISL_2586892 | 4/29/2021 | North America | Canada | Saskatchewan |
| Canada/SK-NML-74442/2021 | EPI_ISL_2587810 | 5/11/2021 | North America | Canada | Saskatchewan |
| Canada/SK-NML-94253/2021 | EPI_ISL_2889505 | 5/30/2021 | North America | Canada | Saskatchewan |
| Canada/SK-NML-94678/2021 | EPI_ISL_2889758 | 6/7/2021 | North America | Canada | Saskatchewan |
| Canada/UN-CRCHUM-PreFreeze-9/2020 | EPI_ISL_667766 | 9/4/2020 | North America | Canada | Canada |
| Canada/un-RIM-1/2020 | EPI_ISL_2932447 | 3/26/2020 | North America | Canada | Canada |
| CentralAfricanRepublic/301/2021 | EPI_ISL_2365416 | 4/22/2021 | Africa | Central African Republic | Bayanga |
| CentralAfricanRepublic/326/2021 | EPI_ISL_2365417 | 4/26/2021 | Africa | Central African Republic | Bayanga |

| | | | | | |
|---|---|---|---|---|---|
| Chile/AN-119268/2021 | EPI_ISL_3132252 | 6/30/2021 | South America | Chile | Antofagasta |
| Chile/LR-89690/2021 | EPI_ISL_2391238 | 5/3/2021 | South America | Chile | Los Rios CL |
| Chile/MA-212217/2020 | EPI_ISL_1300477 | 10/7/2020 | South America | Chile | Magallanes |
| Chile/MA-CADIUMAG-35/2020 | EPI_ISL_681700 | 8/15/2020 | South America | Chile | Magallanes |
| Chile/RM-100571/2021 | EPI_ISL_2557300 | 5/27/2021 | South America | Chile | Region Metropolitana de Santiago |
| Chile/RM-105574/2021 | EPI_ISL_2659196 | 6/4/2021 | South America | Chile | Region Metropolitana de Santiago |
| Chile/RM-119999/2021 | EPI_ISL_3132258 | 7/3/2021 | South America | Chile | Region Metropolitana de Santiago |
| Chile/RM-120001/2021 | EPI_ISL_3132260 | 7/3/2021 | South America | Chile | Region Metropolitana de Santiago |
| Chile/RM-128078/2020 | EPI_ISL_746497 | 7/18/2020 | South America | Chile | Region Metropolitana de Santiago |
| Chile/RM-222868/2020 | EPI_ISL_1167696 | 4/11/2020 | South America | Chile | Region Metropolitana de Santiago |
| Chile/RM-266423/2020 | EPI_ISL_1167925 | 12/30/2020 | South America | Chile | Region Metropolitana de Santiago |
| Chile/RM-34060/2021 | EPI_ISL_1321587 | 2/14/2021 | South America | Chile | Region Metropolitana de Santiago |
| Chile/RM-55193/2021 | EPI_ISL_1633487 | 3/16/2021 | South America | Chile | Region Metropolitana de Santiago |
| Chile/RM-72428/2020 | EPI_ISL_746775 | 5/22/2020 | South America | Chile | Region Metropolitana de Santiago |
| Chile/RM-73255/2020 | EPI_ISL_746791 | 5/22/2020 | South America | Chile | Region Metropolitana de Santiago |
| Chile/RM-77288/2021 | EPI_ISL_2597232 | 4/13/2021 | South America | Chile | Region Metropolitana de Santiago |
| Chile/RM-83100/2021 | EPI_ISL_2597283 | 4/21/2021 | South America | Chile | Region Metropolitana de Santiago |
| Chile/RM-85695/2020 | EPI_ISL_746814 | 5/31/2020 | South America | Chile | Region Metropolitana de Santiago |
| Chile/RM-86374/2021 | EPI_ISL_2391078 | 4/29/2021 | South America | Chile | Region Metropolitana de Santiago |
| Chongqing/210052/2021 | EPI_ISL_1970349 | 4/21/2021 | Asia | China | Chongqing |

| | | | | | |
|---|---|---|---|---|---|
| Colombia/ANT-CWOHC-VG-SEC00744P/2021 | EPI_ISL_2621839 | 4/27/2021 | South America | Colombia | Antioquia |
| Colombia/ANT-PRO-LDSP40/2021 | EPI_ISL_3347200 | 4/27/2021 | South America | Colombia | Antioquia |
| Colombia/BOL-INS-VG-3060/2021 | EPI_ISL_3385778 | 6/1/2021 | South America | Colombia | Bolivar CO |
| Colombia/COV_24770/2021 | EPI_ISL_3216908 | 5/19/2021 | South America | Colombia | Valle del Cauca |
| Colombia/CUN-INS-VG-5033/2021 | EPI_ISL_3459403 | 7/8/2021 | South America | Colombia | Cundinamarca |
| Colombia/DC-INS-05-07/2020 | EPI_ISL_526974 | 6/17/2020 | South America | Colombia | Bogota |
| Colombia/DC-INSV-H-6758/2021 | EPI_ISL_3459414 | 8/4/2021 | South America | Colombia | Bogota |
| Colombia/GUR-37097/2020 | EPI_ISL_941995 | 6/23/2020 | South America | Colombia | Atlantico |
| Colombia/GVI-133304/2020 | EPI_ISL_941986 | 5/29/2020 | South America | Colombia | Norte de Santander |
| Colombia/HUI-INS-VG-4805/2021 | EPI_ISL_3456029 | 7/16/2021 | South America | Colombia | Huila CO |
| Colombia/MET-INS-VG-3994/2021 | EPI_ISL_3007486 | 6/8/2021 | South America | Colombia | Meta |
| Colombia/VAC-INS-VG-3536/2021 | EPI_ISL_2657871 | 5/4/2021 | South America | Colombia | Valle del Cauca |
| Congo/FCRM-16-8378/2021 | EPI_ISL_2889848 | 6/19/2021 | Africa | Republic of the Congo | Brazzaville |
| Congo/FCRM-43-A29-03-07/2021 | EPI_ISL_3040130 | 7/3/2021 | Africa | Republic of the Congo | Brazzaville |
| Congo/FCRM-47-A16-03-07/2021 | EPI_ISL_3040134 | 7/3/2021 | Africa | Republic of the Congo | Brazzaville |
| Congo/FCRM-57-A24-19-04/2021 | EPI_ISL_3342566 | 4/19/2021 | Africa | Republic of the Congo | Brazzaville |
| Congo/FCRM-59-A9-26-04/2021 | EPI_ISL_3342568 | 4/26/2021 | Africa | Republic of the Congo | Brazzaville |
| CostaRica/CRV-0023/2020 | EPI_ISL_480327 | 4/4/2020 | North America | Costa Rica | San Jose |
| CostaRica/HNN-0356/2020 | EPI_ISL_1517400 | 9/1/2020 | North America | Costa Rica | Costa Rica |
| CostaRica/HNN-0359/2020 | EPI_ISL_1517403 | 5/31/2020 | North America | Costa Rica | Heredia |

| | | | | | |
|---|---|---|---|---|---|
| CostaRica/HNN-0361/2020 | EPI_ISL_1517405 | 6/2/2020 | North America | Costa Rica | Limon |
| CostaRica/HNN-0374/2020 | EPI_ISL_1517418 | 12/30/2020 | North America | Costa Rica | Limon |
| CostaRica/HNN-0392/2021 | EPI_ISL_1527012 | 2/17/2021 | North America | Costa Rica | Limon |
| CostaRica/HNN-0398/2021 | EPI_ISL_1527018 | 2/22/2021 | North America | Costa Rica | Heredia |
| CostaRica/HNN-0457/2021 | EPI_ISL_1811227 | 4/5/2021 | North America | Costa Rica | Costa Rica |
| CostaRica/HNN-0466/2021 | EPI_ISL_1811236 | 4/17/2021 | North America | Costa Rica | Costa Rica |
| CostaRica/HNN-0567/2021 | EPI_ISL_2103368 | 4/1/2021 | North America | Costa Rica | Costa Rica |
| CostaRica/INC-0039/2020 | EPI_ISL_491450 | 3/18/2020 | North America | Costa Rica | Guanacaste |
| CostaRica/INC-0052/2020 | EPI_ISL_512658 | 3/21/2020 | North America | Costa Rica | Puntarenas |
| CostaRica/INC-0053/2020 | EPI_ISL_512659 | 6/8/2020 | North America | Costa Rica | Alajuela |
| CostaRica/INC-0062/2020 | EPI_ISL_512668 | 6/19/2020 | North America | Costa Rica | Puntarenas |
| CostaRica/INC-0068/2020 | EPI_ISL_512673 | 7/2/2020 | North America | Costa Rica | Limon |
| CostaRica/INC-0069/2020 | EPI_ISL_512674 | 7/2/2020 | North America | Costa Rica | Costa Rica |
| CostaRica/INC-0071/2020 | EPI_ISL_527739 | 3/6/2020 | North America | Costa Rica | Costa Rica |
| CostaRica/INC-0079/2020 | EPI_ISL_527747 | 7/3/2020 | North America | Costa Rica | Guanacaste |
| CostaRica/INC-0080/2020 | EPI_ISL_527748 | 7/5/2020 | North America | Costa Rica | Limon |
| CostaRica/INC-0096/2020 | EPI_ISL_682238 | 7/3/2020 | North America | Costa Rica | Guanacaste |
| CostaRica/INC-0127/2020 | EPI_ISL_682266 | 8/6/2020 | North America | Costa Rica | San Jose |
| CostaRica/INC-0143/2020 | EPI_ISL_769993 | 9/5/2020 | North America | Costa Rica | Cartago CR |
| CostaRica/INC-0145/2020 | EPI_ISL_770011 | 9/7/2020 | North America | Costa Rica | San Jose |

| | | | | | |
|---|---|---|---|---|---|
| CostaRica/INC-0148/2020 | EPI_ISL_769991 | 9/15/2020 | North America | Costa Rica | Alajuela |
| CostaRica/INC-0155/2020 | EPI_ISL_769992 | 9/30/2020 | North America | Costa Rica | Alajuela |
| CostaRica/INC-0161/2020 | EPI_ISL_770002 | 10/9/2020 | North America | Costa Rica | Guanacaste |
| CostaRica/INC-0179/2020 | EPI_ISL_770024 | 11/16/2020 | North America | Costa Rica | Alajuela |
| CostaRica/INC-0185/2020 | EPI_ISL_770005 | 11/25/2020 | North America | Costa Rica | Heredia |
| CostaRica/INC-0201/2020 | EPI_ISL_914807 | 12/21/2020 | North America | Costa Rica | Alajuela |
| CostaRica/INC-0211/2020 | EPI_ISL_914817 | 12/31/2020 | North America | Costa Rica | Heredia |
| CostaRica/INC-0215/2020 | EPI_ISL_914821 | 12/23/2020 | North America | Costa Rica | Guanacaste |
| CostaRica/INC-0294/2021 | EPI_ISL_1196437 | 2/15/2021 | North America | Costa Rica | San Jose |
| CostaRica/INC-0323/2021 | EPI_ISL_1379416 | 2/22/2021 | North America | Costa Rica | Cartago CR |
| CostaRica/INC-0342/2021 | EPI_ISL_1379434 | 2/27/2021 | North America | Costa Rica | Puntarenas |
| CostaRica/INC-0413/2021 | EPI_ISL_1712382 | 3/3/2021 | North America | Costa Rica | Alajuela |
| CostaRica/INC-0423/2021 | EPI_ISL_1712392 | 3/1/2021 | North America | Costa Rica | Limon |
| CostaRica/INC-0433/2021 | EPI_ISL_1712402 | 3/1/2021 | North America | Costa Rica | Guanacaste |
| CostaRica/INC-0451/2021 | EPI_ISL_1712419 | 3/9/2021 | North America | Costa Rica | Heredia |
| CostaRica/INC-0492/2021 | EPI_ISL_1827521 | 3/15/2021 | North America | Costa Rica | San Jose |
| CostaRica/INC-0524/2021 | EPI_ISL_2103379 | 3/24/2021 | North America | Costa Rica | San Jose |
| CostaRica/INC-0535/2021 | EPI_ISL_2103389 | 3/26/2021 | North America | Costa Rica | Alajuela |
| CostaRica/INC-0558/2021 | EPI_ISL_2103409 | 4/9/2021 | North America | Costa Rica | Alajuela |
| CostaRica/INC-0564/2021 | EPI_ISL_2103415 | 4/14/2021 | North America | Costa Rica | Limon |

| | | | | | |
|---|---|---|---|---|---|
| CostaRica/INC-0591/2021 | EPI_ISL_2272977 | 4/16/2021 | North America | Costa Rica | Limon |
| CostaRica/INC-0592/2021 | EPI_ISL_2272978 | 4/19/2021 | North America | Costa Rica | Cartago CR |
| CostaRica/INC-0598/2021 | EPI_ISL_2272984 | 4/19/2021 | North America | Costa Rica | San Jose |
| CostaRica/INC-0599/2021 | EPI_ISL_2272985 | 4/20/2021 | North America | Costa Rica | Puntarenas |
| CostaRica/INC-0601/2021 | EPI_ISL_2272987 | 4/16/2021 | North America | Costa Rica | Puntarenas |
| CostaRica/INC-0605/2021 | EPI_ISL_2272991 | 4/19/2021 | North America | Costa Rica | Alajuela |
| CostaRica/INC-0618/2021 | EPI_ISL_2273004 | 4/27/2021 | North America | Costa Rica | Cartago CR |
| CostaRica/INC-0620/2021 | EPI_ISL_2502719 | 4/22/2021 | North America | Costa Rica | Heredia |
| CostaRica/INC-0621/2021 | EPI_ISL_2657686 | 4/23/2021 | North America | Costa Rica | Alajuela |
| CostaRica/INC-0624/2021 | EPI_ISL_2502722 | 4/29/2021 | North America | Costa Rica | San Jose |
| CostaRica/INC-0630/2021 | EPI_ISL_2502728 | 4/26/2021 | North America | Costa Rica | Guanacaste |
| CostaRica/INC-0631/2021 | EPI_ISL_2502729 | 5/5/2021 | North America | Costa Rica | Guanacaste |
| CostaRica/INC-0633/2021 | EPI_ISL_2502731 | 5/4/2021 | North America | Costa Rica | Heredia |
| CostaRica/INC-0634/2021 | EPI_ISL_2502732 | 5/5/2021 | North America | Costa Rica | Cartago CR |
| CostaRica/INC-0635/2021 | EPI_ISL_2502733 | 5/3/2021 | North America | Costa Rica | Limon |
| CostaRica/INC-0639/2021 | EPI_ISL_2502737 | 4/30/2021 | North America | Costa Rica | Limon |
| CostaRica/INC-0640/2021 | EPI_ISL_2502738 | 5/11/2021 | North America | Costa Rica | Puntarenas |
| CostaRica/INC-0643/2021 | EPI_ISL_2502741 | 5/9/2021 | North America | Costa Rica | Alajuela |
| CostaRica/INC-0646/2021 | EPI_ISL_2502744 | 5/11/2021 | North America | Costa Rica | Limon |
| CostaRica/INC-0647/2021 | EPI_ISL_2502745 | 5/10/2021 | North America | Costa Rica | Cartago CR |

| | | | | | |
|---|---|---|---|---|---|
| CostaRica/INC-0648/2021 | EPI_ISL_2502746 | 5/14/2021 | North America | Costa Rica | Costa Rica |
| CostaRica/INC-0656/2021 | EPI_ISL_2502752 | 5/21/2021 | North America | Costa Rica | San Jose |
| CostaRica/INC-0658/2021 | EPI_ISL_2502754 | 5/18/2021 | North America | Costa Rica | San Jose |
| CostaRica/INC-0664/2021 | EPI_ISL_2502760 | 5/20/2021 | North America | Costa Rica | Heredia |
| CostaRica/INC-0666/2021 | EPI_ISL_2502762 | 5/21/2021 | North America | Costa Rica | Costa Rica |
| CostaRica/INC-0668/2021 | EPI_ISL_2658265 | 4/23/2021 | North America | Costa Rica | Guanacaste |
| CostaRica/INC-0680/2021 | EPI_ISL_2658277 | 4/29/2021 | North America | Costa Rica | Heredia |
| CostaRica/INC-0681/2021 | EPI_ISL_2658278 | 5/2/2021 | North America | Costa Rica | Alajuela |
| CostaRica/INC-0682/2021 | EPI_ISL_3274346 | 5/5/2021 | North America | Costa Rica | Guanacaste |
| CostaRica/INC-0712/2021 | EPI_ISL_2827982 | 5/25/2021 | North America | Costa Rica | Puntarenas |
| CostaRica/INC-0747/2021 | EPI_ISL_3026022 | 6/24/2021 | North America | Costa Rica | Costa Rica |
| CostaRica/INC-0750/2021 | EPI_ISL_3026025 | 6/22/2021 | North America | Costa Rica | Heredia |
| CostaRica/INC-0751/2021 | EPI_ISL_3026026 | 6/30/2021 | North America | Costa Rica | Puntarenas |
| CostaRica/INC-0753/2021 | EPI_ISL_3026028 | 6/29/2021 | North America | Costa Rica | Alajuela |
| CostaRica/INC-0754/2021 | EPI_ISL_3026029 | 7/1/2021 | North America | Costa Rica | San Jose |
| CostaRica/INC-0757/2021 | EPI_ISL_3026030 | 7/7/2021 | North America | Costa Rica | Heredia |
| CostaRica/INC-0759/2021 | EPI_ISL_3026032 | 7/7/2021 | North America | Costa Rica | Limon |
| CostaRica/INC-0768/2021 | EPI_ISL_3037802 | 6/17/2021 | North America | Costa Rica | Cartago CR |
| CostaRica/INC-0775/2021 | EPI_ISL_3037810 | 6/18/2021 | North America | Costa Rica | Limon |
| CostaRica/INC-0778/2021 | EPI_ISL_3037812 | 6/17/2021 | North America | Costa Rica | Guanacaste |

| | | | | | |
|---|---|---|---|---|---|
| CostaRica/INC-0779/2021 | EPI_ISL_3037814 | 6/16/2021 | North America | Costa Rica | Puntarenas |
| CostaRica/INC-0790/2021 | EPI_ISL_3037824 | 6/22/2021 | North America | Costa Rica | Cartago CR |
| CostaRica/INC-0793/2021 | EPI_ISL_3274359 | 6/24/2021 | North America | Costa Rica | Alajuela |
| CostaRica/INC-0795/2021 | EPI_ISL_3037826 | 6/29/2021 | North America | Costa Rica | San Jose |
| CostaRica/INC-0796/2021 | EPI_ISL_3037828 | 6/28/2021 | North America | Costa Rica | Heredia |
| CostaRica/INC-0800/2021 | EPI_ISL_3037834 | 6/29/2021 | North America | Costa Rica | Limon |
| CostaRica/INC-0806/2021 | EPI_ISL_3274364 | 6/30/2021 | North America | Costa Rica | San Jose |
| CostaRica/INC-0814/2021 | EPI_ISL_3274367 | 7/1/2021 | North America | Costa Rica | San Jose |
| CostaRica/INC-0815/2021 | EPI_ISL_3274368 | 7/7/2021 | North America | Costa Rica | Cartago CR |
| CostaRica/INC-0874/2021 | EPI_ISL_3298362 | 7/19/2021 | North America | Costa Rica | Guanacaste |
| CostaRica/INC-0894/2021 | EPI_ISL_3464520 | 7/9/2021 | North America | Costa Rica | Puntarenas |
| CostaRica/INC-0895/2021 | EPI_ISL_3464521 | 7/21/2021 | North America | Costa Rica | Heredia |
| CostaRica/INC-0898/2021 | EPI_ISL_3464523 | 7/19/2021 | North America | Costa Rica | Limon |
| CostaRica/INC-0906/2021 | EPI_ISL_3464529 | 7/23/2021 | North America | Costa Rica | Alajuela |
| CostaRica/INC-0917/2021 | EPI_ISL_3464514 | 7/18/2021 | North America | Costa Rica | Puntarenas |
| CostaRica/INC-0919/2021 | EPI_ISL_3464538 | 7/20/2021 | North America | Costa Rica | Guanacaste |
| CostaRica/INC-0929/2021 | EPI_ISL_3464546 | 7/24/2021 | North America | Costa Rica | Alajuela |
| Crimea/CR-CRIE-L189Z0141ubh/2021 | EPI_ISL_3101330 | 5/29/2021 | Europe | Ukraine | Crimea |
| Crimea/RII-MH29254S/2021 | EPI_ISL_3123000 | 7/1/2021 | Europe | Ukraine | Crimea |
| Crimea/RII-MH29259S/2021 | EPI_ISL_3123004 | 7/1/2021 | Europe | Ukraine | Crimea |
| Crimea/RII-MH2937S/2020 | EPI_ISL_733078 | 9/21/2020 | Europe | Ukraine | Crimea |

| | | | | | |
|---|---|---|---|---|---|
| Crimea/RII-MH9353S/2020 | EPI_ISL_1400527 | 10/11/2020 | Europe | Ukraine | Crimea |
| Croatia/3997/2021 | EPI_ISL_2657628 | 4/20/2021 | Europe | Croatia | Zagreb |
| Croatia/4799/2021 | EPI_ISL_2674469 | 4/20/2021 | Europe | Croatia | Zagreb |
| Croatia/5478/2021 | EPI_ISL_2674135 | 5/16/2021 | Europe | Croatia | Zagreb |
| Croatia/5571/2021 | EPI_ISL_2673910 | 5/20/2021 | Europe | Croatia | Dubrovnik-Neretva County |
| Croatia/6759/2021 | EPI_ISL_3061341 | 6/9/2021 | Europe | Croatia | Pozega-Slavonia County |
| Croatia/6860/2021 | EPI_ISL_3061828 | 7/1/2021 | Europe | Croatia | Primorje-Gorski Kotar County |
| Croatia/6959/2021 | EPI_ISL_3151164 | 6/29/2021 | Europe | Croatia | Split-Dalmatia County |
| Croatia/7100/2021 | EPI_ISL_3151788 | 7/8/2021 | Europe | Croatia | Dubrovnik-Neretva County |
| Croatia/Zagreb-27/2020 | EPI_ISL_710547 | 9/3/2020 | Europe | Croatia | Zagreb |
| Cuba/USAFSAM-S030/2020 | EPI_ISL_513312 | 3/19/2020 | North America | Cuba | Cuba |
| Cuba/USAFSAM-S031/2020 | EPI_ISL_513313 | 3/24/2020 | North America | Cuba | Cuba |
| Curacao/CW-RIVM-10364/2020 | EPI_ISL_636519 | 8/11/2020 | South America | Curacao | Curacao |
| Curacao/CW-RIVM-10365/2020 | EPI_ISL_636520 | 8/11/2020 | South America | Curacao | Curacao |
| Curacao/CW-RIVM-10366/2020 | EPI_ISL_636521 | 8/11/2020 | South America | Curacao | Curacao |
| Curacao/CW-RIVM-13012/2021 | EPI_ISL_1035770 | 1/27/2021 | South America | Curacao | Curacao |
| Curacao/CW-RIVM-21708/2020 | EPI_ISL_1014574 | 11/7/2020 | South America | Curacao | Curacao |
| Curacao/CW-RIVM-21718/2020 | EPI_ISL_1014564 | 10/31/2020 | South America | Curacao | Curacao |
| Curacao/CW-RIVM-21719/2020 | EPI_ISL_1014563 | 10/31/2020 | South America | Curacao | Curacao |
| Curacao/CW-RIVM-21720/2020 | EPI_ISL_1014562 | 10/30/2020 | South America | Curacao | Curacao |
| Curacao/CW-RIVM-30491/2021 | EPI_ISL_2094367 | 4/26/2021 | South America | Curacao | Curacao |
| Curacao/CW-RIVM-31692/2021 | EPI_ISL_2220456 | 4/28/2021 | South America | Curacao | Curacao |

| | | | | | |
|---|---|---|---|---|---|
| Curacao/CW-RIVM-36939/2021 | EPI_ISL_2475681 | 5/19/2021 | South America | Curacao | Curacao |
| Curacao/CW-RIVM-36968/2021 | EPI_ISL_2475682 | 5/22/2021 | South America | Curacao | Curacao |
| Curacao/CW-RIVM-43118/2021 | EPI_ISL_3056740 | 6/22/2021 | South America | Curacao | Curacao |
| Curacao/CW-RIVM-43128/2021 | EPI_ISL_3056750 | 6/23/2021 | South America | Curacao | Curacao |
| Curacao/CW-RIVM-44509/2021 | EPI_ISL_3136581 | 7/10/2021 | South America | Curacao | Curacao |
| Curacao/CW-RIVM-48240/2021 | EPI_ISL_3259188 | 7/22/2021 | South America | Curacao | Curacao |
| Curacao/CW-RIVM-50160/2021 | EPI_ISL_3390661 | 8/1/2021 | South America | Curacao | Curacao |
| Curacao/CW-RIVM-50177/2021 | EPI_ISL_3389278 | 8/1/2021 | South America | Curacao | Curacao |
| Cyprus/008/2020 | EPI_ISL_463748 | 4/27/2020 | Europe | Cyprus | Cyprus |
| Cyprus/Cy038919/2020 | EPI_ISL_1164629 | 7/11/2020 | Europe | Cyprus | Cyprus |
| Cyprus/Cy167120/2021 | EPI_ISL_1164746 | 1/26/2021 | Europe | Cyprus | Cyprus |
| CzechRepublic/46/2021 | EPI_ISL_3308764 | 7/11/2021 | Europe | Czech Republic | Czech Republic |
| CzechRepublic/CSQ0325/2021 | EPI_ISL_3396497 | 8/2/2021 | Europe | Czech Republic | Vysocina Region |
| CzechRepublic/CSQ0397/2021 | EPI_ISL_3446646 | 8/2/2021 | Europe | Czech Republic | Vysocina Region |
| CzechRepublic/FNB31310/2021 | EPI_ISL_2545562 | 4/21/2021 | Europe | Czech Republic | South Moravian Region |
| CzechRepublic/IAB20_016_036/2020 | EPI_ISL_889361 | 7/22/2020 | Europe | Czech Republic | Prague |
| CzechRepublic/NRL_10092/2021 | EPI_ISL_3318987 | 2/12/2021 | Europe | Czech Republic | Southern Bohemia Region |
| CzechRepublic/NRL_10749/2020 | EPI_ISL_737014 | 11/2/2020 | Europe | Czech Republic | Usti nad Labem |
| CzechRepublic/NRL_4711-2/2021 | EPI_ISL_1828701 | 3/30/2021 | Europe | Czech Republic | Vysocina Region |
| CzechRepublic/NRL_5428_21/2021 | EPI_ISL_2002105 | 3/22/2021 | Europe | Czech Republic | Plze≈à Region |
| CzechRepublic/NRL_7027/2021 | EPI_ISL_2380424 | 4/22/2021 | Europe | Czech Republic | Prague |
| CzechRepublic/NRL_7439_21/2021 | EPI_ISL_2324946 | 5/4/2021 | Europe | Czech Republic | Czech Republic |
| CzechRepublic/NRL_7779/2021 | EPI_ISL_2466508 | 5/17/2021 | Europe | Czech Republic | Prague |
| CzechRepublic/NRL_9348/2021 | EPI_ISL_3063136 | 6/22/2021 | Europe | Czech Republic | Prague |

| | | | | | |
|---|---|---|---|---|---|
| CzechRepublic/NRL_9560/2021 | EPI_ISL_3229664 | 6/22/2021 | Europe | Czech Republic | Pardubice Region |
| CzechRepublic/NRL_9882/2021 | EPI_ISL_3318886 | 7/9/2021 | Europe | Czech Republic | Prague |
| CzechRepublic/NRL-10288/2020 | EPI_ISL_660557 | 10/22/2020 | Europe | Czech Republic | Prague |
| CzechRepublic/NRL-6834/2020 | EPI_ISL_541333 | 5/23/2020 | Europe | Czech Republic | Moravian-Silesian Region |
| CzechRepublic/NRL-7963/2020 | EPI_ISL_584078 | 7/29/2020 | Europe | Czech Republic | Prague |
| Denmark/ALAB-SSI-634/2020 | EPI_ISL_437010 | 3/26/2020 | Europe | Denmark | Denmark |
| Denmark/DCGC-109873/2021 | EPI_ISL_2680848 | 5/24/2021 | Europe | Denmark | Hovedstaden |
| Denmark/DCGC-115059/2021 | EPI_ISL_2668530 | 5/24/2021 | Europe | Denmark | Hovedstaden |
| Denmark/DCGC-116366/2021 | EPI_ISL_2669458 | 6/7/2021 | Europe | Denmark | Hovedstaden |
| Denmark/DCGC-118902/2021 | EPI_ISL_2671258 | 6/14/2021 | Europe | Denmark | Hovedstaden |
| Denmark/DCGC-11998/2020 | EPI_ISL_671245 | 7/27/2020 | Europe | Denmark | Syddanmark |
| Denmark/DCGC-142030/2021 | EPI_ISL_3240890 | 7/31/2021 | Europe | Denmark | Hovedstaden |
| Denmark/DCGC-142511/2021 | EPI_ISL_3241336 | 7/31/2021 | Europe | Denmark | Hovedstaden |
| Denmark/DCGC-144363/2021 | EPI_ISL_3252793 | 8/2/2021 | Europe | Denmark | Hovedstaden |
| Denmark/DCGC-150161/2021 | EPI_ISL_3418236 | 8/9/2021 | Europe | Denmark | Hovedstaden |
| Denmark/DCGC-1817/2020 | EPI_ISL_617940 | 5/11/2020 | Europe | Denmark | Hovedstaden |
| Denmark/DCGC-32696/2021 | EPI_ISL_869700 | 1/11/2021 | Europe | Denmark | Hovedstaden |
| Denmark/DCGC-67988/2021 | EPI_ISL_1876559 | 2/22/2021 | Europe | Denmark | Hovedstaden |
| Denmark/DCGC-73800/2021 | EPI_ISL_1882371 | 4/5/2021 | Europe | Denmark | Hovedstaden |
| Denmark/DCGC-74590/2021 | EPI_ISL_1883161 | 4/19/2021 | Europe | Denmark | Sjaelland |
| Denmark/DCGC-82058/2021 | EPI_ISL_1890724 | 3/22/2021 | Europe | Denmark | Midtjylland |
| Denmark/DCGC-92006/2021 | EPI_ISL_2028301 | 4/19/2021 | Europe | Denmark | Hovedstaden |
| Denmark/SSI-01/2020 | EPI_ISL_416142 | 2/26/2020 | Europe | Denmark | Hovedstaden |
| Djibouti/NAMRU3_A104/2021 | EPI_ISL_2858619 | 5/3/2021 | Africa | Djibouti | Djibouti |
| Djibouti/NAMRU3_C58/2021 | EPI_ISL_2858683 | 1/20/2021 | Africa | Djibouti | Djibouti |
| DominicanRepublic/CDC-9KZX-8454/2020 | EPI_ISL_906851 | 12/27/2020 | North America | Dominican Republic | Dominican Republic |
| DominicanRepublic/UNIBE001S/2021 | EPI_ISL_2601034 | 6/10/2021 | North America | Dominican Republic | Dominican Republic |

| | | | | | |
|---|---|---|---|---|---|
| DominicanRepublic/UNIBE002/2021 | EPI_ISL_2601035 | 6/10/2021 | North America | Dominican Republic | Dominican Republic |
| DominicanRepublic/Yale-2290/2020 | EPI_ISL_1378844 | 11/18/2020 | North America | Dominican Republic | Dominican Republic |
| DominicanRepublic/Yale-5479/2021 | EPI_ISL_2776175 | 4/14/2021 | North America | Dominican Republic | Santo Domingo |
| DominicanRepublic/Yale-5487/2021 | EPI_ISL_2776177 | 4/23/2021 | North America | Dominican Republic | Santo Domingo |
| DominicanRepublic/Yale-5491/2021 | EPI_ISL_2776180 | 4/7/2021 | North America | Dominican Republic | Santo Domingo |
| DominicanRepublic/Yale-6241/2021 | EPI_ISL_3104762 | 5/11/2021 | North America | Dominican Republic | Dominican Republic |
| DominicanRepublic/Yale-6265/2021 | EPI_ISL_3188565 | 5/12/2021 | North America | Dominican Republic | Dominican Republic |
| DominicanRepublic/Yale-6278/2021 | EPI_ISL_3236435 | 5/18/2021 | North America | Dominican Republic | Santo Domingo |
| DominicanRepublic/Yale-6280/2021 | EPI_ISL_3236437 | 5/18/2021 | North America | Dominican Republic | Santo Domingo |
| DominicanRepublic/Yale-6288/2021 | EPI_ISL_3236445 | 6/1/2021 | North America | Dominican Republic | Santo Domingo |
| DominicanRepublic/Yale-6298/2021 | EPI_ISL_3236451 | 6/29/2021 | North America | Dominican Republic | Santo Domingo |
| DRC/146998/2021 | EPI_ISL_965122 | 1/28/2021 | Africa | Democratic Republic of the Congo | Kinshasa |
| DRC/2723/2020 | EPI_ISL_3133658 | 4/16/2020 | Africa | Democratic Republic of the Congo | Kinshasa |
| DRC/298978/2021 | EPI_ISL_2135841 | 4/20/2021 | Africa | Democratic Republic of the Congo | Kinshasa |
| DRC/299174/2021 | EPI_ISL_2135844 | 4/20/2021 | Africa | Democratic Republic of the Congo | Kinshasa |
| DRC/INRB-RDC-534/2021 | EPI_ISL_3086912 | 7/12/2021 | Africa | Democratic Republic of the Congo | Kinshasa |
| DRC/INRB-RDC-548/2021 | EPI_ISL_3086923 | 7/12/2021 | Africa | Democratic Republic of the Congo | Kinshasa |
| DRC/KCTCOV006/2021 | EPI_ISL_1785567 | 3/23/2021 | Africa | Democratic Republic of the Congo | Kongo Central |
| DRC/RDC-129/2021 | EPI_ISL_2968721 | 5/24/2021 | Africa | Democratic Republic of the Congo | Kinshasa |
| DRC/RDC-142/2021 | EPI_ISL_2968730 | 5/25/2021 | Africa | Democratic Republic of the Congo | Kinshasa |

| | | | | | |
|---|---|---|---|---|---|
| DRC/RDC-368/2021 | EPI_ISL_2966639 | 6/18/2021 | Africa | Democratic Republic of the Congo | Haut-U√©l√© |
| DRC/RDC-380/2021 | EPI_ISL_2966646 | 6/14/2021 | Africa | Democratic Republic of the Congo | Haut-U√©l√© |
| Ecuador/52255/2020 | EPI_ISL_491952 | 5/29/2020 | South America | Ecuador | Guayas |
| Ecuador/EC-M-48776/2020 | EPI_ISL_826820 | 5/24/2020 | South America | Ecuador | Manabi |
| Ecuador/EC-R-65804/2020 | EPI_ISL_826823 | 6/25/2020 | South America | Ecuador | Los Rios EC |
| Ecuador/NIC-INSPI-268325/2021 | EPI_ISL_2348792 | 3/23/2021 | South America | Ecuador | El Oro |
| Ecuador/NIC-INSPI-333303/2021 | EPI_ISL_2895671 | 6/22/2021 | South America | Ecuador | El Oro |
| Ecuador/NIC-INSPI-334444/2021 | EPI_ISL_2895674 | 6/24/2021 | South America | Ecuador | El Oro |
| Ecuador/NIC-INSPI-336275/2021 | EPI_ISL_2895673 | 7/1/2021 | South America | Ecuador | Guayas |
| Ecuador/NIC-INSPI-34277/2021 | EPI_ISL_2348789 | 5/23/2021 | South America | Ecuador | Pastaza |
| Ecuador/UEES-ECU420/2020 | EPI_ISL_1443642 | 9/6/2020 | South America | Ecuador | Imbabura |
| Ecuador/UEES-ECU539/2020 | EPI_ISL_1443651 | 10/29/2020 | South America | Ecuador | Imbabura |
| Ecuador/UEES-ECU633/2020 | EPI_ISL_1443656 | 12/10/2020 | South America | Ecuador | Imbabura |
| Ecuador/UEES-INTERLAB-6849811/2021 | EPI_ISL_3088329 | 7/20/2021 | South America | Ecuador | Guayas |
| Ecuador/USFQ-1097/2021 | EPI_ISL_1805714 | 4/8/2021 | South America | Ecuador | Pichincha |
| Ecuador/USFQ-1181/2021 | EPI_ISL_1896697 | 4/19/2021 | South America | Ecuador | Santo Domingo de los Tsachilas |
| Ecuador/USFQ-1205-UTPL060/2021 | EPI_ISL_2004107 | 4/21/2021 | South America | Ecuador | Loja |
| Ecuador/USFQ-1304/2021 | EPI_ISL_2361456 | 3/31/2021 | South America | Ecuador | Orellana |
| Ecuador/USFQ-1416/2021 | EPI_ISL_2361471 | 5/10/2021 | South America | Ecuador | Galapagos |
| Ecuador/USFQ-185/2020 | EPI_ISL_697784 | 8/12/2020 | South America | Ecuador | Imbabura |

| | | | | | |
|---|---|---|---|---|---|
| Ecuador/USFQ-1865/2021 | EPI_ISL_3260710 | 8/1/2021 | South America | Ecuador | Imbabura |
| Ecuador/USFQ-561/2020 | EPI_ISL_824290 | 12/29/2020 | South America | Ecuador | Imbabura |
| Ecuador/ZZ-SARS-2/2020 | EPI_ISL_681703 | 4/30/2020 | South America | Ecuador | Pichincha |
| Egypt/ARMY-306/2021 | EPI_ISL_1936245 | 4/22/2021 | Africa | Egypt | Cairo |
| Egypt/ARMY-307/2021 | EPI_ISL_1936246 | 4/22/2021 | Africa | Egypt | Cairo |
| Egypt/CCHE57357_Wave_3_A019/2021 | EPI_ISL_2566479 | 5/8/2021 | Africa | Egypt | Egypt |
| Egypt/CCHE57357_Wave_3_A070/2021 | EPI_ISL_2566512 | 5/10/2021 | Africa | Egypt | Egypt |
| Egypt/CPHL-A3/2021 | EPI_ISL_3274159 | 8/1/2021 | Africa | Egypt | Cairo |
| Egypt/CPHL-NRC-20/2020 | EPI_ISL_794593 | 3/13/2020 | Africa | Egypt | Egypt |
| Egypt/CPHL-NRC-23/2020 | EPI_ISL_794602 | 3/13/2020 | Africa | Egypt | Egypt |
| Egypt/CPHL-S4/2021 | EPI_ISL_3274151 | 6/16/2021 | Africa | Egypt | Cairo |
| Egypt/CPHL-S9/2021 | EPI_ISL_3274156 | 7/8/2021 | Africa | Egypt | Sohag |
| Egypt/NRC-6888/2020 | EPI_ISL_2232330 | 10/14/2020 | Africa | Egypt | Egypt |
| ElSalvador/Gorgas-27/2021 | EPI_ISL_3275314 | 4/29/2021 | North America | El Salvador | El Salvador |
| ElSalvador/Gorgas-92/2021 | EPI_ISL_3275343 | 4/30/2021 | North America | El Salvador | El Salvador |
| ElSalvador/INS-01/2020 | EPI_ISL_671974 | 9/16/2020 | North America | El Salvador | San Salvador |
| England/ALDP-148AB33/2021 | EPI_ISL_1487669 | 3/26/2021 | Europe | United Kingdom | England |
| England/EXET-139146/2020 | EPI_ISL_664612 | 10/27/2020 | Europe | United Kingdom | England |
| England/HSLL-15BC311/2021 | EPI_ISL_2436959 | 5/25/2021 | Europe | United Kingdom | England |
| England/HSLL-161984F/2021 | EPI_ISL_2516071 | 6/3/2021 | Europe | United Kingdom | England |
| England/MILK-151B3D4/2021 | EPI_ISL_1759200 | 4/19/2021 | Europe | United Kingdom | England |
| England/MILK-1550661/2021 | EPI_ISL_1985736 | 4/29/2021 | Europe | United Kingdom | England |
| England/MILK-15AD60B/2021 | EPI_ISL_2346547 | 5/22/2021 | Europe | United Kingdom | England |
| England/MILK-B93B8E/2020 | EPI_ISL_676278 | 11/13/2020 | Europe | United Kingdom | England |
| England/PHEC-P307PCA8/2021 | EPI_ISL_3179613 | 7/30/2021 | Europe | United Kingdom | England |

| | | | | | |
|---|---|---|---|---|---|
| England/PHEC-R304R968/2021 | EPI_ISL_3292547 | 8/6/2021 | Europe | United Kingdom | England |
| env/Liaoning/Dalian-IVDC-Pollock-30-05/2020 | EPI_ISL_2170893 | 5/30/2020 | Asia | China | Liaoning |
| EquatorialGuinea/23265/2020 | EPI_ISL_648368 | 6/22/2020 | Africa | Equatorial Guinea | Bioko Norte |
| EquatorialGuinea/47001/2020 | EPI_ISL_953411 | 8/4/2020 | Africa | Equatorial Guinea | Bioko Norte |
| EquatorialGuinea/6777/2020 | EPI_ISL_953403 | 5/8/2020 | Africa | Equatorial Guinea | Litoral |
| EquatorialGuinea/89984/2021 | EPI_ISL_1673322 | 2/3/2021 | Africa | Equatorial Guinea | Equatorial Guinea |
| EquatorialGuinea/98273/2021 | EPI_ISL_2002688 | 3/1/2021 | Africa | Equatorial Guinea | Equatorial Guinea |
| EquatorialGuinea/98283/2021 | EPI_ISL_1700687 | 3/1/2021 | Africa | Equatorial Guinea | Equatorial Guinea |
| Estonia/15572186/2021 | EPI_ISL_2545909 | 4/23/2021 | Europe | Estonia | Estonia |
| Estonia/16043642/2021 | EPI_ISL_2545924 | 4/23/2021 | Europe | Estonia | Estonia |
| Estonia/16733150/2021 | EPI_ISL_2788837 | 5/6/2021 | Europe | Estonia | Estonia |
| Estonia/16739141/2021 | EPI_ISL_3087615 | 6/3/2021 | Europe | Estonia | Estonia |
| Estonia/17319563/2021 | EPI_ISL_3087639 | 6/3/2021 | Europe | Estonia | Estonia |
| Estonia/C145366/2021 | EPI_ISL_2788800 | 5/8/2021 | Europe | Estonia | Estonia |
| Estonia/Cov11025/2021 | EPI_ISL_2642237 | 3/27/2021 | Europe | Estonia | Harjumaa |
| Estonia/Cov4512/2020 | EPI_ISL_2642816 | 11/16/2020 | Europe | Estonia | Estonia |
| Estonia/Cov4823/2020 | EPI_ISL_2642810 | 11/15/2020 | Europe | Estonia | Estonia |
| Estonia/Cov7703/2020 | EPI_ISL_2642761 | 12/26/2020 | Europe | Estonia | Estonia |
| Eswatini/N4796/2021 | EPI_ISL_1827699 | 2/23/2021 | Africa | Eswatini | Hhohho |
| Ethiopia/AHRI-002/2020 | EPI_ISL_3071136 | 12/18/2020 | Africa | Ethiopia | Addis Ababa |
| Ethiopia/ICGEB-S11/2021 | EPI_ISL_2241493 | 1/28/2021 | Africa | Ethiopia | Addis Ababa |
| Ethiopia/ICGEB-S15/2021 | EPI_ISL_2241623 | 5/1/2021 | Africa | Ethiopia | Addis Ababa |
| Finland/1/2020 | EPI_ISL_407079 | 1/29/2020 | Europe | Finland | Finland |
| Finland/10098/2021 | EPI_ISL_3135404 | 7/11/2021 | Europe | Finland | Uusimaa |
| Finland/17MR10cH11/2020 | EPI_ISL_757302 | 11/17/2020 | Europe | Finland | Uusimaa |
| Finland/22TouS1G7/2020 | EPI_ISL_1233188 | 5/22/2020 | Europe | Finland | Uusimaa |
| Finland/29M70S2/2020 | EPI_ISL_1240167 | 3/29/2020 | Europe | Finland | Uusimaa |

| | | | | | |
|---|---|---|---|---|---|
| Finland/7842/2021 | EPI_ISL_3014589 | 5/25/2021 | Europe | Finland | Uusimaa |
| Finland/8780/2021 | EPI_ISL_3135635 | 6/17/2021 | Europe | Finland | Uusimaa |
| Finland/8849/2021 | EPI_ISL_3135664 | 6/20/2021 | Europe | Finland | Uusimaa |
| Finland/9714/2021 | EPI_ISL_3135147 | 7/3/2021 | Europe | Finland | Uusimaa |
| Finland/THL-202112375/2021 | EPI_ISL_2363788 | 4/27/2021 | Europe | Finland | Finland |
| Finland/THL-202112512/2021 | EPI_ISL_2508226 | 4/27/2021 | Europe | Finland | Finland |
| Finland/THL-202118021/2021 | EPI_ISL_2644929 | 5/7/2021 | Europe | Finland | Finland |
| France/ARA-HCL021085298401/2021 | EPI_ISL_2373664 | 4/27/2021 | Europe | France | Auvergne-Rhv¥ne-Alpes |
| France/ARA-HCL021138335402/2021 | EPI_ISL_3472380 | 8/2/2021 | Europe | France | Auvergne-Rhv¥ne-Alpes |
| France/GES-8536/2020 | EPI_ISL_560587 | 5/26/2020 | Europe | France | Grand Est |
| France/GP-HMN-21052200198/2021 | EPI_ISL_2464487 | 5/10/2021 | North America | Guadeloupe | Guadeloupe |
| France/GP-HMN-21052200199/2021 | EPI_ISL_2464488 | 5/11/2021 | North America | Guadeloupe | Guadeloupe |
| France/GP-HMN-21072050400/2021 | EPI_ISL_2983904 | 6/29/2021 | North America | Guadeloupe | Guadeloupe |
| France/GP-HMN-21072050411/2021 | EPI_ISL_2983829 | 6/29/2021 | North America | Guadeloupe | Guadeloupe |
| France/HDF-IPP12232/2021 | EPI_ISL_2293459 | 5/11/2021 | Europe | France | Hauts de France |
| France/IDF_HB_112001106076/2020 | EPI_ISL_940552 | 1/29/2020 | Europe | France | Ile de France |
| France/IDF-0515/2020 | EPI_ISL_408430 | 1/29/2020 | Europe | France | Ile de France |
| France/IDF-HMN-21042290740/2021 | EPI_ISL_2151655 | 4/27/2021 | Europe | France | Ile de France |
| France/IDF-HMN-21052270313/2021 | EPI_ISL_2464863 | 5/25/2021 | Europe | France | Ile de France |
| France/IDF-HMN-21082050241/2021 | EPI_ISL_3477489 | 8/2/2021 | Europe | France | Ile de France |
| France/IDF-IPP14150/2021 | EPI_ISL_2709263 | 6/7/2021 | Europe | France | Ile de France |
| France/PAC-HCL021127175801/2021 | EPI_ISL_3130669 | 7/6/2021 | Europe | France | Provence-Alpes-Cv¥te d'Azur |
| France/PAC-HCL021140989701/2021 | EPI_ISL_3473250 | 7/23/2021 | Europe | France | Provence-Alpes-Cv¥te d'Azur |
| France/PAC-IHU-4048-N1/2021 | EPI_ISL_1694791 | 4/2/2021 | Europe | France | Provence-Alpes-Cv¥te d'Azur |

| | | | | | |
|---|---|---|---|---|---|
| France/PAC-IHUCOVID-0244/2020 | EPI_ISL_900153 | 3/30/2020 | Europe | France | Provence-Alpes-C√¥te d'Azur |
| France/PAC-IHUCOVID-2709/2020 | EPI_ISL_900129 | 10/28/2020 | Europe | France | Provence-Alpes-C√¥te d'Azur |
| France/PDL-IPP14812/2021 | EPI_ISL_2835886 | 6/22/2021 | Europe | France | Pays de la Loire |
| Fujian/2021BD-VIR-SPE-0408/2021 | EPI_ISL_3006798 | 5/21/2021 | Asia | China | Fujian |
| Fujian/2021BD-VIR-SPE-0410/2021 | EPI_ISL_3006799 | 5/21/2021 | Asia | China | Fujian |
| Gabon/AP360/2020 | EPI_ISL_1760554 | 12/28/2020 | Africa | Gabon | Libreville |
| Gabon/CERMEL-CC0433/2021 | EPI_ISL_2442383 | 4/20/2021 | Africa | Gabon | Tchibanga |
| Gabon/CERMEL-DD0321/2021 | EPI_ISL_2442279 | 5/7/2021 | Africa | Gabon | Lambarene |
| Gabon/CERMEL-DD0590/2021 | EPI_ISL_2424160 | 5/12/2021 | Africa | Gabon | Lambarene |
| Gabon/ITM-K028/2020 | EPI_ISL_539576 | 3/27/2020 | Africa | Gabon | Libreville |
| Gabon/LPDG0405/2021 | EPI_ISL_3129580 | 3/29/2021 | Africa | Gabon | Libreville |
| Gabon/LPDG662/2021 | EPI_ISL_3461136 | 4/20/2021 | Africa | Gabon | Libreville |
| Gambia/0214/2020 | EPI_ISL_471158 | 3/29/2020 | Africa | Gambia | Kombo |
| Gambia/23736/2020 | EPI_ISL_1234531 | 12/22/2020 | Africa | Gambia | Kombo |
| Gambia/31887/2021 | EPI_ISL_2958626 | 4/21/2021 | Africa | Gambia | Kombo |
| Gambia/33059/2021 | EPI_ISL_2958632 | 4/27/2021 | Africa | Gambia | Kombo |
| Gambia/36747/2021 | EPI_ISL_2958645 | 5/19/2021 | Africa | Gambia | Kombo |
| Gambia/37853/2021 | EPI_ISL_2958636 | 5/23/2021 | Africa | Gambia | Kombo |
| Gambia/46581/2021 | EPI_ISL_3150943 | 6/29/2021 | Africa | Gambia | Basse |
| Gambia/46728/2021 | EPI_ISL_3150942 | 6/30/2021 | Africa | Gambia | Basse |
| Gambia/47701/2021 | EPI_ISL_3132302 | 7/4/2021 | Africa | Gambia | Kombo |
| Gambia/48862/2021 | EPI_ISL_3150936 | 7/9/2021 | Africa | Gambia | Kombo |
| Gambia/NPHL-3281/2020 | EPI_ISL_561316 | 7/28/2020 | Africa | Gambia | Kombo |
| Georgia/Tb-86952/2020 | EPI_ISL_754180 | 9/17/2020 | Asia | Georgia | Tbilisi |
| Georgia/Tb-RNGS003/2021 | EPI_ISL_1921859 | 10/10/2020 | Asia | Georgia | Tbilisi |
| Georgia/Tb-RNGS040/2021 | EPI_ISL_3098702 | 4/26/2021 | Asia | Georgia | Tbilisi |
| Georgia/Tb-SNGS031/2021 | EPI_ISL_1791042 | 2/22/2021 | Asia | Georgia | Tbilisi |

| | | | | | |
|---|---|---|---|---|---|
| Georgia/Tb-SNGS067/2021 | EPI_ISL_1914670 | 3/17/2021 | Asia | Georgia | Tbilisi |
| Georgia/Tb-SNGS168/2021 | EPI_ISL_3204260 | 5/23/2021 | Asia | Georgia | Tbilisi |
| Georgia/Tb-SNGS243/2021 | EPI_ISL_3204247 | 5/5/2021 | Asia | Georgia | Tbilisi |
| Georgia/Tb-SNGS260/2021 | EPI_ISL_3204237 | 6/4/2021 | Asia | Georgia | Tbilisi |
| Georgia/Tb-SNGS269/2021 | EPI_ISL_3204251 | 6/9/2021 | Asia | Georgia | Tbilisi |
| Georgia/Tb-SNGS372/2021 | EPI_ISL_3208351 | 7/20/2021 | Asia | Georgia | Tbilisi |
| Georgia/Tb-SNGS374/2021 | EPI_ISL_3208445 | 7/17/2021 | Asia | Georgia | Tbilisi |
| Germany/BE-ChVir-LB-210806-5843/2021 | EPI_ISL_3447115 | 8/3/2021 | Europe | Germany | Berlin |
| Germany/BE-ChVir-LB-210813-4575/2021 | EPI_ISL_3447217 | 8/4/2021 | Europe | Germany | Berlin |
| Germany/BE-RKI-I-106660/2021 | EPI_ISL_1850943 | 4/18/2021 | Europe | Germany | Berlin |
| Germany/BY-RKI-I-137265/2021 | EPI_ISL_2120884 | 4/29/2021 | Europe | Germany | Bavaria |
| Germany/BY-RKI-I-185285/2021 | EPI_ISL_2845698 | 5/30/2021 | Europe | Germany | Bavaria |
| Germany/BY-RKI-I-185287/2021 | EPI_ISL_2845700 | 5/25/2021 | Europe | Germany | Bavaria |
| Germany/BY-RKI-I-185553/2021 | EPI_ISL_2845803 | 6/1/2021 | Europe | Germany | Bavaria |
| Germany/NW-HHU-7863/2021 | EPI_ISL_2993641 | 7/9/2021 | Europe | Germany | North Rhine Westphalia |
| Germany/NW-RKI-I-187850/2021 | EPI_ISL_3044294 | 7/10/2021 | Europe | Germany | North Rhine Westphalia |
| Germany/NW-RKI-I-189360/2021 | EPI_ISL_3120799 | 6/5/2021 | Europe | Germany | North Rhine Westphalia |
| Germany/SL-SU-20239546/2020 | EPI_ISL_708013 | 10/14/2020 | Europe | Germany | Saarland |
| Germany/SL-SU-20242379/2020 | EPI_ISL_708015 | 10/21/2020 | Europe | Germany | Saarland |
| Germany/un-RKI-I-081827/2021 | EPI_ISL_1646872 | 3/24/2021 | Europe | Germany | Germany |
| Germany/un-RKI-I-096081/2021 | EPI_ISL_1843038 | 4/7/2021 | Europe | Germany | Germany |
| Ghana/1622_S2/2020 | EPI_ISL_422384 | 3/24/2020 | Africa | Ghana | Greater Accra |
| Ghana/nCoV-TRA-735/2021 | EPI_ISL_2873872 | 4/20/2021 | Africa | Ghana | Greater Accra |
| Ghana/NMIMR-NTRA-21-25429/2021 | EPI_ISL_2508390 | 2/9/2021 | Africa | Ghana | Ghana |
| Ghana/WACCBIP-GS1106/2021 | EPI_ISL_3268148 | 7/5/2021 | Africa | Ghana | Greater Accra |
| Ghana/WACCBIP-GS1125/2021 | EPI_ISL_3268026 | 7/1/2021 | Africa | Ghana | Greater Accra |
| Ghana/WACCBIP-GS1163/2021 | EPI_ISL_3268156 | 6/29/2021 | Africa | Ghana | Greater Accra |

| | | | | | |
|---|---|---|---|---|---|
| Ghana/WACCBIP-GS479/2020 | EPI_ISL_2422527 | 9/3/2020 | Africa | Ghana | Eastern Region GH |
| Ghana/WACCBIP-GS485/2020 | EPI_ISL_2422532 | 9/3/2020 | Africa | Ghana | Greater Accra |
| Ghana/WACCBIP-GS905/2021 | EPI_ISL_2836945 | 6/18/2021 | Africa | Ghana | Greater Accra |
| Ghana/WACCBIP-GS911/2021 | EPI_ISL_2836950 | 5/21/2021 | Africa | Ghana | Greater Accra |
| Ghana/WACCBIP-GS917/2021 | EPI_ISL_2836955 | 5/24/2021 | Africa | Ghana | Greater Accra |
| Ghana/WACCBIP-TRA744/2020 | EPI_ISL_2285867 | 4/22/2021 | Africa | Ghana | Greater Accra |
| Greece/16348/2021 | EPI_ISL_3045183 | 6/11/2021 | Europe | Greece | Greece |
| Greece/17276/2021 | EPI_ISL_3045257 | 6/17/2021 | Europe | Greece | Greece |
| Greece/180471/2021 | EPI_ISL_2343929 | 4/8/2021 | Europe | Greece | Western Greece |
| Greece/218163/2021 | EPI_ISL_2617296 | 5/23/2021 | Europe | Greece | Peloponnese |
| Greece/218321/2021 | EPI_ISL_2617369 | 5/26/2021 | Europe | Greece | Peloponnese |
| Greece/92010/2021 | EPI_ISL_1503229 | 3/13/2021 | Europe | Greece | Greece |
| Greece/V874/2021 | EPI_ISL_1964145 | 4/23/2021 | Europe | Greece | Greece |
| Greece/V955/2021 | EPI_ISL_2233869 | 4/30/2021 | Europe | Greece | Thessaloniki |
| Grenada/63766/2021 | EPI_ISL_2545198 | 4/8/2021 | North America | Grenada | Grenada |
| Grenada/75608/2021 | EPI_ISL_2967993 | 6/24/2021 | North America | Grenada | Grenada |
| Guadeloupe/GP-HMN-21072130432/2021 | EPI_ISL_3390946 | 7/6/2021 | North America | Guadeloupe | Guadeloupe |
| Guadeloupe/GP-HMN-21072130434/2021 | EPI_ISL_3390947 | 7/6/2021 | North America | Guadeloupe | Guadeloupe |
| Guadeloupe/IPG-7320/2020 | EPI_ISL_613420 | 3/14/2020 | North America | Guadeloupe | Les Abymes |
| Guadeloupe/IPG-7330/2020 | EPI_ISL_613430 | 3/16/2020 | North America | Guadeloupe | Lamentin |
| Guadeloupe/IPG-7334/2020 | EPI_ISL_613433 | 3/23/2020 | North America | Guadeloupe | Guadeloupe |
| Guadeloupe/IPG-7339/2020 | EPI_ISL_613438 | 3/22/2020 | North America | Guadeloupe | Le Moule |
| Guadeloupe/IPG-7340/2020 | EPI_ISL_613439 | 3/24/2020 | North America | Guadeloupe | Trois-RiviV®res |
| Guadeloupe/IPG-7346/2020 | EPI_ISL_613443 | 3/28/2020 | North America | Guadeloupe | Les Abymes |

| | | | | | |
|---|---|---|---|---|---|
| Guadeloupe/IPG-7353/2020 | EPI_ISL_613445 | 3/31/2020 | North America | Guadeloupe | M√©tropole |
| Guadeloupe/IPP05357/2021 | EPI_ISL_1381163 | 3/2/2021 | North America | Guadeloupe | Pointe-A-Pitre |
| Guadeloupe/IPP06253/2021 | EPI_ISL_1416981 | 2/26/2021 | North America | Guadeloupe | Les Abymes |
| Guadeloupe/IPP11460/2021 | EPI_ISL_2188319 | 4/27/2021 | North America | Guadeloupe | Pointe-A-Pitre |
| Guadeloupe/IPP11462/2021 | EPI_ISL_2188321 | 4/27/2021 | North America | Guadeloupe | Pointe-A-Pitre |
| Guadeloupe/IPP12789/2021 | EPI_ISL_2462927 | 5/11/2021 | North America | Guadeloupe | Basse-Terre |
| Guadeloupe/IPP12864/2021 | EPI_ISL_2462934 | 5/11/2021 | North America | Guadeloupe | Pointe-A-Pitre |
| Guadeloupe/IPP12865/2021 | EPI_ISL_2462935 | 5/11/2021 | North America | Guadeloupe | Pointe-A-Pitre |
| Guadeloupe/IPP12983/2021 | EPI_ISL_2462904 | 5/10/2021 | North America | Guadeloupe | Les Abymes |
| Guadeloupe/IPP13824/2021 | EPI_ISL_2531889 | 5/24/2021 | North America | Guadeloupe | Les Abymes |
| Guadeloupe/IPP14050/2021 | EPI_ISL_2628275 | 6/8/2021 | North America | Guadeloupe | Pointe-A-Pitre |
| Guadeloupe/IPP14148/2021 | EPI_ISL_2709261 | 5/25/2021 | North America | Guadeloupe | Baie-Mahault |
| Guadeloupe/IPP14836/2021 | EPI_ISL_2835906 | 6/22/2021 | North America | Guadeloupe | Basse-Terre |
| Guadeloupe/IPP14837/2021 | EPI_ISL_2835907 | 6/22/2021 | North America | Guadeloupe | Basse-Terre |
| Guadeloupe/IPP15246/2021 | EPI_ISL_3031653 | 6/29/2021 | North America | Guadeloupe | Pointe-A-Pitre |
| Guadeloupe/IPP15819/2021 | EPI_ISL_3058248 | 7/6/2021 | North America | Guadeloupe | Pointe-A-Pitre |
| Guadeloupe/IPP18572/2021 | EPI_ISL_3355749 | 7/20/2021 | North America | Guadeloupe | Les Abymes |
| Guadeloupe/IPP18586/2021 | EPI_ISL_3355762 | 7/20/2021 | North America | Guadeloupe | Les Abymes |
| Guadeloupe/IPP18932/2021 | EPI_ISL_3392683 | 7/19/2021 | North America | Guadeloupe | Pointe-A-Pitre |
| Guam/GU-CDC-2-3847017/2020 | EPI_ISL_1273073 | 11/27/2020 | North America | USA | Guam |

| | | | | | |
|---|---|---|---|---|---|
| Guatemala/1915/2021 | EPI_ISL_3275191 | 4/30/2021 | North America | Guatemala | Guatemala |
| Guatemala/19335/2021 | EPI_ISL_3275183 | 3/26/2021 | North America | Guatemala | Guatemala |
| Guatemala/2009/2021 | EPI_ISL_3275222 | 5/11/2021 | North America | Guatemala | Guatemala |
| Guatemala/2294/2021 | EPI_ISL_3319047 | 6/15/2021 | North America | Guatemala | Guatemala |
| Guatemala/2434/2021 | EPI_ISL_3275242 | 6/19/2021 | North America | Guatemala | Guatemala |
| Guatemala/34860/2021 | EPI_ISL_3275248 | 5/24/2021 | North America | Guatemala | Guatemala |
| Guatemala/730/2021 | EPI_ISL_3275203 | 4/7/2021 | North America | Guatemala | Guatemala |
| Guatemala/839/2021 | EPI_ISL_3275216 | 4/26/2021 | North America | Guatemala | Guatemala |
| Guatemala/ASI01001/2020 | EPI_ISL_3446721 | 6/24/2020 | North America | Guatemala | Totonicapan |
| Guatemala/ASI01026/2020 | EPI_ISL_3446724 | 6/8/2020 | North America | Guatemala | El Progreso |
| Guatemala/ASI02009/2021 | EPI_ISL_3446677 | 3/2/2021 | North America | Guatemala | Guatemala City |
| Guatemala/ASI02020/2021 | EPI_ISL_3446698 | 3/1/2021 | North America | Guatemala | San Marcos |
| Guatemala/ASI02052/2021 | EPI_ISL_3446713 | 3/19/2021 | North America | Guatemala | Jalapa |
| Guatemala/ASI03005/2021 | EPI_ISL_3347863 | 7/7/2021 | North America | Guatemala | Guatemala |
| Guatemala/ASI03007/2021 | EPI_ISL_3347954 | 7/21/2021 | North America | Guatemala | Guatemala |
| Guatemala/GUA-LNS-1661/2021 | EPI_ISL_2825081 | 1/8/2021 | North America | Guatemala | Guatemala |
| Guatemala/GUA-LNS-2064/2021 | EPI_ISL_2825088 | 1/10/2021 | North America | Guatemala | Guatemala |
| Guatemala/GUA-LNS-3394/2021 | EPI_ISL_2826930 | 1/12/2021 | North America | Guatemala | Quiche |
| Guatemala/GUA-LNS-3619/2021 | EPI_ISL_2826932 | 1/14/2021 | North America | Guatemala | El Progreso |
| Guatemala/GUA-LNS-47753/2020 | EPI_ISL_2827829 | 10/2/2020 | North America | Guatemala | Quetzaltenango |

| | | | | | |
|---|---|---|---|---|---|
| Guatemala/GUA-LNS-48359/2020 | EPI_ISL_2827835 | 10/7/2020 | North America | Guatemala | Jalapa |
| Guatemala/GUA-LNS-48504/2020 | EPI_ISL_2825117 | 10/8/2020 | North America | Guatemala | Zacapa |
| Guatemala/GUA-LNS-49688/2020 | EPI_ISL_2827848 | 10/19/2020 | North America | Guatemala | Huehuetenango |
| Guinea/C4/2021 | EPI_ISL_3281521 | 6/25/2021 | Africa | Guinea | Boke |
| Guinea/Conakry_IPG002/2020 | EPI_ISL_2245780 | 3/31/2020 | Africa | Guinea | Conakry |
| Guinea/Conakry_IPG11718/2020 | EPI_ISL_2245960 | 9/16/2020 | Africa | Guinea | Dabola |
| Guinea/Conakry_IPG17343/2021 | EPI_ISL_2245877 | 1/9/2021 | Africa | Guinea | Conakry |
| Guinea/Conakry_IPG1998/2020 | EPI_ISL_2245796 | 5/3/2020 | Africa | Guinea | Conakry |
| Guinea/Conakry_IPG20021/2021 | EPI_ISL_2245922 | 3/30/2021 | Africa | Guinea | Conakry |
| Guinea/Conakry_IPG2287/2020 | EPI_ISL_2245798 | 5/9/2020 | Africa | Guinea | Conakry |
| Guyana/50817/2020 | EPI_ISL_2230692 | 3/27/2020 | South America | Guyana | Guyana |
| Haiti/18256/2021 | EPI_ISL_2157559 | 2/1/2021 | North America | Haiti | Haiti |
| Haiti/27703/2021 | EPI_ISL_2492524 | 5/18/2021 | North America | Haiti | Haiti |
| Haiti/27716/2021 | EPI_ISL_2492536 | 5/20/2021 | North America | Haiti | Haiti |
| Haiti/33970/2020 | EPI_ISL_2157560 | 3/22/2020 | North America | Haiti | Haiti |
| Honduras/316254/2021 | EPI_ISL_2650523 | 4/15/2021 | North America | Honduras | Olancho |
| Honduras/318102/2021 | EPI_ISL_2648242 | 4/16/2021 | North America | Honduras | La Paz HN |
| Honduras/321522/2021 | EPI_ISL_2650524 | 4/21/2021 | North America | Honduras | Comayagua |
| Honduras/321542/2021 | EPI_ISL_2648244 | 4/22/2021 | North America | Honduras | Comayagua |
| Honduras/324506/2021 | EPI_ISL_2648246 | 4/21/2021 | North America | Honduras | El Parav≠so |
| Honduras/326764/2021 | EPI_ISL_2650529 | 4/23/2021 | North America | Honduras | El Parav≠so |
| Honduras/327005/2021 | EPI_ISL_2648252 | 4/25/2021 | North America | Honduras | Valle |

| | | | | | |
|---|---|---|---|---|---|
| Honduras/327008/2021 | EPI_ISL_2648253 | 4/25/2021 | North America | Honduras | Valle |
| Honduras/327331/2021 | EPI_ISL_2650530 | 4/26/2021 | North America | Honduras | Francisco Morazv°n |
| Honduras/327649/2021 | EPI_ISL_2648254 | 4/26/2021 | North America | Honduras | Francisco Morazv°n |
| Honduras/327951/2021 | EPI_ISL_2648256 | 4/16/2021 | North America | Honduras | Choluteca |
| Honduras/328015/2021 | EPI_ISL_2650533 | 4/24/2021 | North America | Honduras | La Paz HN |
| Honduras/336319/2021 | EPI_ISL_2650534 | 5/4/2021 | North America | Honduras | Santa Barbara |
| Honduras/38712/2021 | EPI_ISL_2648260 | 4/27/2021 | North America | Honduras | Cortv©s |
| Honduras/38845/2021 | EPI_ISL_2650537 | 4/27/2021 | North America | Honduras | Cortv©s |
| Honduras/5413/2021 | EPI_ISL_2650535 | 4/24/2021 | North America | Honduras | Departamento Colon |
| Honduras/5748/2021 | EPI_ISL_2648259 | 4/27/2021 | North America | Honduras | Atlv°ntida |
| Honduras/CM-USAFSAM-S4219/2021 | EPI_ISL_3375855 | 5/27/2021 | North America | Honduras | Comayagua |
| Honduras/CM-USAFSAM-S4221/2021 | EPI_ISL_3375857 | 5/27/2021 | North America | Honduras | Comayagua |
| HongKong/CM21000337/2021 | EPI_ISL_2931338 | 4/18/2021 | Asia | Hong Kong | Hong Kong |
| HongKong/CM21000372/2021 | EPI_ISL_2611661 | 5/24/2021 | Asia | Hong Kong | Hong Kong |
| HongKong/CM21000386/2021 | EPI_ISL_2651119 | 6/6/2021 | Asia | Hong Kong | Hong Kong |
| HongKong/CM21000387/2021 | EPI_ISL_2651120 | 6/6/2021 | Asia | Hong Kong | Hong Kong |
| HongKong/CM21000437/2021 | EPI_ISL_3219440 | 7/18/2021 | Asia | Hong Kong | Hong Kong |
| HongKong/HKPU-11837/2021 | EPI_ISL_2713263 | 5/26/2021 | Asia | Hong Kong | Hong Kong |
| HongKong/HKU-201216-084/2020 | EPI_ISL_1034433 | 7/24/2020 | Asia | Hong Kong | Hong Kong |
| HongKong/HKU-201216-145/2020 | EPI_ISL_1034438 | 7/27/2020 | Asia | Hong Kong | Hong Kong |
| HongKong/HKU-201216-167/2020 | EPI_ISL_1034460 | 8/4/2020 | Asia | Hong Kong | Hong Kong |
| HongKong/VM21021001/2021 | EPI_ISL_2931339 | 4/18/2021 | Asia | Hong Kong | Hong Kong |
| HongKong/VM21032402/2021 | EPI_ISL_3019491 | 7/1/2021 | Asia | Hong Kong | Hong Kong |

| | | | | | |
|---|---|---|---|---|---|
| Hungary/HM-0121-26/2021 | EPI_ISL_1041201 | 1/21/2021 | Europe | Hungary | B√°cs-Kiskun County |
| Hungary/HM-0201-71/2021 | EPI_ISL_1041211 | 2/1/2021 | Europe | Hungary | B√°cs-Kiskun County |
| Hungary/SRC-00817/2020 | EPI_ISL_435413 | 3/30/2020 | Europe | Hungary | Baranya County |
| Hungary/US-57575w/2020 | EPI_ISL_677798 | 9/28/2020 | Europe | Hungary | B√©k√©s County |
| Iceland/13/2020 | EPI_ISL_417765 | 2/27/2020 | Europe | Iceland | Reykjavik |
| Iceland/5456/2020 | EPI_ISL_829138 | 11/21/2020 | Europe | Iceland | Reykjavik |
| India/AP-CCMB-CIA5896/2021 | EPI_ISL_3060713 | 6/19/2021 | Asia | India | Andhra Pradesh |
| India/AP-CCMB-CIA6346/2021 | EPI_ISL_3453444 | 7/2/2021 | Asia | India | Andhra Pradesh |
| India/AP-CCMB-CIA6411/2021 | EPI_ISL_3453474 | 7/5/2021 | Asia | India | Andhra Pradesh |
| India/DL-ILBS-19969/2020 | EPI_ISL_3276914 | 6/9/2020 | Asia | India | Delhi |
| India/DL-ILBS-20926/2020 | EPI_ISL_3276927 | 6/11/2020 | Asia | India | Delhi |
| India/MH-NIV-10435/2020 | EPI_ISL_541683 | 4/16/2020 | Asia | India | Maharashtra |
| India/MH-NIV-4271/2020 | EPI_ISL_454530 | 3/22/2020 | Asia | India | Maharashtra |
| India/MH-NIV-5104/2020 | EPI_ISL_454537 | 3/26/2020 | Asia | India | Maharashtra |
| India/PY-JsCOVb-41026_S173_R1_001/2021 | EPI_ISL_3265125 | 5/18/2021 | Asia | India | Puducherry |
| India/TG-CCMB-CIA2007/2021 | EPI_ISL_2373395 | 5/6/2021 | Asia | India | Telangana |
| India/TG-CCMB-CIA4834/2021 | EPI_ISL_2774958 | 6/3/2021 | Asia | India | Andhra Pradesh |
| India/TN-C_100_S15_R1_001/2021 | EPI_ISL_2379462 | 2/5/2021 | Asia | India | Tamil Nadu |
| India/TN-C_117_S56_R1_001/2021 | EPI_ISL_2379479 | 4/28/2021 | Asia | India | Tamil Nadu |
| India/TN-C_217_S184_R1_001/2021 | EPI_ISL_2379575 | 4/22/2021 | Asia | India | Tamil Nadu |
| Indonesia/JB-GSILab-532100/2021 | EPI_ISL_2824991 | 6/28/2021 | Asia | Indonesia | West Java |
| Indonesia/JB-KWG-LIPI-NIHRD-0023/2021 | EPI_ISL_2631464 | 5/26/2021 | Asia | Indonesia | West Java |
| Indonesia/JI-ITD-3590NT/2020 | EPI_ISL_437188 | 4/14/2020 | Asia | Indonesia | East Java |
| Indonesia/JK-GSILab-579423/2021 | EPI_ISL_3230181 | 7/12/2021 | Asia | Indonesia | Jakarta |
| Indonesia/JK-GSILab-643704/2021 | EPI_ISL_3401855 | 8/3/2021 | Asia | Indonesia | Jakarta |
| Indonesia/JK-GSILab-646024/2021 | EPI_ISL_3404358 | 8/3/2021 | Asia | Indonesia | Jakarta |
| Indonesia/JK-NIHRD-MI2102800/2021 | EPI_ISL_1824601 | 2/25/2021 | Asia | Indonesia | Jakarta |

| | | | | | |
|---|---|---|---|---|---|
| Indonesia/JK-NIHRD-WGS02010/2021 | EPI_ISL_2382405 | 3/11/2021 | Asia | Indonesia | Jakarta |
| Indonesia/JK-NIHRD-WGS02966/2021 | EPI_ISL_2262260 | 4/30/2021 | Asia | Indonesia | Jakarta |
| Indonesia/JK-NIHRD-WGS03209/2021 | EPI_ISL_2854698 | 5/3/2021 | Asia | Indonesia | Jakarta |
| Indonesia/KI-NIHRD-WGS02871/2021 | EPI_ISL_2233090 | 4/20/2021 | Asia | Indonesia | East Kalimantan |
| Indonesia/KS-NIHRD-WGS00908/2020 | EPI_ISL_1257851 | 12/15/2020 | Asia | Indonesia | South Kalimantan |
| Indonesia/SB-GSILab-638526/2021 | EPI_ISL_3383410 | 6/29/2021 | Asia | Indonesia | West Sumatra |
| Indonesia/SB-NIHRD-WGS09040/2021 | EPI_ISL_3375342 | 7/12/2021 | Asia | Indonesia | West Sumatra |
| Iran/GRC-9673/2020 | EPI_ISL_596452 | 5/18/2020 | Asia | Iran | Tehran |
| Iran/GRC-Ahv-12-123/2021 | EPI_ISL_2547429 | 3/1/2021 | Asia | Iran | Ahvaz |
| Iran/GRC-S1213/2021 | EPI_ISL_2227272 | 4/28/2021 | Asia | Iran | Yazd |
| Iran/GRC-S1216/2021 | EPI_ISL_2227271 | 4/28/2021 | Asia | Iran | Yazd |
| Iran/GRC-Sh-11-110/2021 | EPI_ISL_2254718 | 1/26/2021 | Asia | Iran | Shiraz |
| Iran/GRC-T11763/2021 | EPI_ISL_2365354 | 2/15/2021 | Asia | Iran | Tehran |
| Iran/Kerman-NIC-K1/2021 | EPI_ISL_2360250 | 5/8/2021 | Asia | Iran | Kerman |
| Iran/Kerman-NIC-K2/2021 | EPI_ISL_2360251 | 5/8/2021 | Asia | Iran | Kerman |
| Iran/PN2179/2020 | EPI_ISL_463749 | 3/26/2020 | Asia | Iran | Iran |
| Iran/Qom-629/2020 | EPI_ISL_1014676 | 9/19/2020 | Asia | Iran | Qom |
| Iran/Tehran-055M/2020 | EPI_ISL_1014687 | 8/17/2020 | Asia | Iran | Tehran |
| Iraq/Erbil4/2021 | EPI_ISL_2234383 | 3/9/2021 | Asia | Iraq | Iraq |
| Iraq/Hula-02/2020 | EPI_ISL_3402237 | 12/14/2020 | Asia | Iraq | Al-Najaf-Al-Ashraf |
| Iraq/Samawa-73/2021 | EPI_ISL_2467924 | 2/11/2021 | Asia | Iraq | Al-Muthanna Province |
| Iraq/Thi-Qar-4/2021 | EPI_ISL_2931134 | 4/27/2021 | Asia | Iraq | Dhi Qar Province |
| Iraq/Thi-Qar-5/2021 | EPI_ISL_2931135 | 4/27/2021 | Asia | Iraq | Dhi Qar Province |
| Iraq/USAFSAM-S072/2020 | EPI_ISL_812267 | 6/7/2020 | Asia | Iraq | Iraq |
| Ireland/CO-NVRL-72IRL13311/2020 | EPI_ISL_528470 | 8/12/2020 | Europe | Ireland | Cork |
| Ireland/CO-NVRL-M35IRL07724/2021 | EPI_ISL_3467377 | 8/7/2021 | Europe | Ireland | Cork |
| Ireland/D-NVRL-20IRL27755/2020 | EPI_ISL_501261 | 6/12/2020 | Europe | Ireland | Dublin |

| | | | | | |
|---|---|---|---|---|---|
| Ireland/D-NVRL-21IRL58947/2021 | EPI_ISL_2240835 | 5/7/2021 | Europe | Ireland | Dublin |
| Ireland/D-NVRL-g22IRL36774/2021 | EPI_ISL_1972662 | 4/21/2021 | Europe | Ireland | Dublin |
| Ireland/D-NVRL-Q23IRL75498/2021 | EPI_ISL_2932234 | 6/10/2021 | Europe | Ireland | Dublin |
| Ireland/D-NVRL-Q23IRL80081/2021 | EPI_ISL_2932231 | 6/11/2021 | Europe | Ireland | Dublin |
| Ireland/D-NVRL-S21IRL00130262/2021 | EPI_ISL_3356242 | 7/25/2021 | Europe | Ireland | Dublin |
| Ireland/D-NVRL-t90IRL00397/2021 | EPI_ISL_2284223 | 4/27/2021 | Europe | Ireland | Dublin |
| Ireland/G-NVRL-M34IRL66540/2021 | EPI_ISL_3467814 | 8/2/2021 | Europe | Ireland | Galway |
| Ireland/LD-NVRL-20G48734/2020 | EPI_ISL_708522 | 4/3/2020 | Europe | Ireland | Longford |
| Ireland/LH-NVRL-20IRL26172/2020 | EPI_ISL_501260 | 6/4/2020 | Europe | Ireland | Louth |
| Ireland/LK-NVRL-30IRL39018/2021 | EPI_ISL_2454884 | 5/19/2021 | Europe | Ireland | Limerick |
| Ireland/U-NVRL-G21IRL70282/2021 | EPI_ISL_3011512 | 7/5/2021 | Europe | Ireland | Dublin |
| Israel/CVL_S7-3/2020 | EPI_ISL_2811910 | 4/18/2020 | Asia | Israel | Israel |
| Israel/CVL-13279/2021 | EPI_ISL_2183479 | 4/29/2021 | Asia | Israel | Central District |
| Israel/CVL-13562/2021 | EPI_ISL_2183806 | 5/10/2021 | Asia | Israel | Jerusalem District |
| Israel/CVL-14052/2021 | EPI_ISL_3278325 | 6/28/2021 | Asia | Israel | Israel |
| Israel/CVL-14436/2021 | EPI_ISL_3278340 | 7/4/2021 | Asia | Israel | Israel |
| Israel/CVL-14584/2021 | EPI_ISL_3278455 | 6/30/2021 | Asia | Israel | Tel Aviv District |
| Israel/CVL-246ngs/2020 | EPI_ISL_889051 | 12/27/2020 | Asia | Palestine | West Bank |
| Israel/CVL-928ngs/2020 | EPI_ISL_889141 | 12/31/2020 | Asia | Palestine | West Bank |
| Israel/HMO-VL-2497871/2020 | EPI_ISL_2096776 | 10/13/2020 | Asia | Israel | Israel |
| Israel/HMO-VL-2674042/2021 | EPI_ISL_2096927 | 4/25/2021 | Asia | Israel | Israel |
| Israel/SMC-7003318/2021 | EPI_ISL_2567034 | 5/19/2021 | Asia | Israel | Israel |
| Israel/SMC-7005693/2021 | EPI_ISL_3022017 | 7/3/2021 | Asia | Israel | Israel |
| Italy/CAM-TIGEM-79/2020 | EPI_ISL_1086105 | 10/26/2020 | Europe | Italy | Campania |
| Italy/EMR-C046_21_30/2021 | EPI_ISL_3266550 | 7/5/2021 | Europe | Italy | Emilia-Romagna |
| Italy/EMR-C046_21_32/2021 | EPI_ISL_3266551 | 7/5/2021 | Europe | Italy | Emilia-Romagna |
| Italy/EMR-C058-21-07_full_variant_seq/2021 | EPI_ISL_3291310 | 8/2/2021 | Europe | Italy | Emilia-Romagna |

| | | | | | |
|---|---|---|---|---|---|
| Italy/EMR-C058-21-35_full_variant_seq/2021 | EPI_ISL_3291260 | 8/2/2021 | Europe | Italy | Emilia-Romagna |
| Italy/LAZ-INMI1-cs/2020 | EPI_ISL_410546 | 1/29/2020 | Europe | Italy | Lazio |
| Italy/LOM-Pavia-38390/2020 | EPI_ISL_1166098 | 4/7/2020 | Europe | Italy | Lombardy |
| Italy/LOM-Sacco_Var_T12802/2021 | EPI_ISL_3120108 | 5/27/2021 | Europe | Italy | Lombardy |
| Italy/LOM-UniMI-L310/2020 | EPI_ISL_542263 | 2/29/2020 | Europe | Italy | Lombardy |
| Italy/SAR-Cagliari259/2021 | EPI_ISL_2361500 | 5/25/2021 | Europe | Italy | Sardinia |
| Italy/TAA-1900571751/2021 | EPI_ISL_1923584 | 4/22/2021 | Europe | Italy | Trentino-Alto Adige |
| Italy/VEN-IZSVe-20RS2155-23_VI/2020 | EPI_ISL_766571 | 11/25/2020 | Europe | Italy | Veneto |
| Italy/VEN-IZSVe-21RS1020-7_VR/2021 | EPI_ISL_2392075 | 4/20/2021 | Europe | Italy | Veneto |
| Italy/VEN-IZSVe-21RS1729-1_PD/2021 | EPI_ISL_2975142 | 6/28/2021 | Europe | Italy | Veneto |
| Italy/VEN-IZSVe-21RS1736-6_PD/2021 | EPI_ISL_2975350 | 6/29/2021 | Europe | Italy | Veneto |
| Italy/VEN-IZSVe-21RS606-6_PD/2021 | EPI_ISL_1524641 | 3/7/2021 | Europe | Italy | Veneto |
| Italy/VEN-UniVR-28/2020 | EPI_ISL_751325 | 8/21/2020 | Europe | Italy | Veneto |
| Jamaica/34728/2021 | EPI_ISL_3235111 | 5/3/2021 | North America | Jamaica | Jamaica |
| Jamaica/34730/2021 | EPI_ISL_3235144 | 5/3/2021 | North America | Jamaica | Jamaica |
| Jamaica/JM-CDC-0078/2020 | EPI_ISL_450792 | 3/11/2020 | North America | Jamaica | Jamaica |
| Jamaica/JM-CDC-5836/2020 | EPI_ISL_450795 | 3/14/2020 | North America | Jamaica | Jamaica |
| Japan/IC-0102/2020 | EPI_ISL_591443 | 7/20/2020 | Asia | Japan | Japan |
| Japan/IC-0583/2020 | EPI_ISL_779669 | 10/28/2020 | Asia | Japan | Japan |
| Japan/IC-0606/2021 | EPI_ISL_779692 | 1/4/2021 | Asia | Japan | Japan |
| Japan/IC-0729/2021 | EPI_ISL_860106 | 1/18/2021 | Asia | Japan | Japan |
| Japan/IC-0951/2021 | EPI_ISL_1927127 | 4/1/2021 | Asia | Japan | Japan |
| Japan/IC-1117/2021 | EPI_ISL_2131643 | 4/25/2021 | Asia | Japan | Japan |
| Japan/IC-1381/2021 | EPI_ISL_2828257 | 6/21/2021 | Asia | Japan | Japan |
| Japan/IC-1427/2021 | EPI_ISL_2828303 | 6/25/2021 | Asia | Japan | Japan |
| Japan/IC-1551/2021 | EPI_ISL_3076649 | 7/12/2021 | Asia | Japan | Japan |

| | | | | | |
|---|---|---|---|---|---|
| Japan/IC-1622/2021 | EPI_ISL_3076720 | 7/21/2021 | Asia | Japan | Japan |
| Japan/IC-1709/2021 | EPI_ISL_3392348 | 8/1/2021 | Asia | Japan | Japan |
| Japan/IC-1711/2021 | EPI_ISL_3392350 | 8/2/2021 | Asia | Japan | Japan |
| Japan/PG-24254/2021 | EPI_ISL_1429597 | 2/12/2021 | Asia | Japan | Mie |
| Japan/PG-45101/2021 | EPI_ISL_2338548 | 4/16/2021 | Asia | Japan | Osaka |
| Japan/PG-48059/2021 | EPI_ISL_2329932 | 5/6/2021 | Asia | Japan | Hiroshima |
| Japan/PG-48060/2021 | EPI_ISL_2329933 | 5/6/2021 | Asia | Japan | Hiroshima |
| Japan/PG-50013/2021 | EPI_ISL_2763105 | 4/27/2021 | Asia | Japan | Fukuoka |
| Japan/PG-6298/2020 | EPI_ISL_689409 | 8/1/2020 | Asia | Japan | Ibaraki |
| Jordan/AM-Biolab021/2021 | EPI_ISL_1824717 | 4/18/2021 | Asia | Jordan | Amman |
| Jordan/AM-Biolab042/2021 | EPI_ISL_2105674 | 5/4/2021 | Asia | Jordan | Amman |
| Jordan/Biolab0079/2021 | EPI_ISL_2658760 | 5/24/2021 | Asia | Jordan | Amman |
| Jordan/Biolab0123/2021 | EPI_ISL_2868422 | 7/3/2021 | Asia | Jordan | Amman |
| Jordan/Biolab020/2021 | EPI_ISL_1823202 | 4/16/2021 | Asia | Jordan | Amman |
| Jordan/MA-ALSR-2583/2020 | EPI_ISL_635780 | 6/4/2020 | Asia | Jordan | Mafraq |
| Jordan/PHBC10/2021 | EPI_ISL_2932618 | 6/1/2021 | Asia | Jordan | Amman |
| Jordan/PHBC8/2021 | EPI_ISL_2932611 | 6/1/2021 | Asia | Jordan | Amman |
| Jordan/SEARCH-5444/2020 | EPI_ISL_755124 | 8/24/2020 | Asia | Jordan | Amman |
| Jordan/SR-055/2020 | EPI_ISL_430013 | 4/2/2020 | Asia | Jordan | Amman |
| Kazakhstan/3352/2021 | EPI_ISL_1341503 | 1/1/2021 | Asia | Kazakhstan | Nur-Sultan |
| Kazakhstan/63610/2020 | EPI_ISL_1365697 | 12/15/2020 | Asia | Kazakhstan | West-Kazakhstan Region |
| Kazakhstan/9008/2021 | EPI_ISL_1448020 | 2/26/2021 | Asia | Kazakhstan | Karaganda Region |
| Kazakhstan/KAR-NRL-565S/2021 | EPI_ISL_2438635 | 4/25/2021 | Asia | Kazakhstan | Karaganda Region |
| Kazakhstan/KZY-NRL-473S/2021 | EPI_ISL_2438606 | 4/18/2021 | Asia | Kazakhstan | Kyzylorda Region |
| Kenya/AFI-KOM-1492/2021 | EPI_ISL_3031402 | 5/31/2021 | Africa | Kenya | Kisumu |
| Kenya/C102434/2021 | EPI_ISL_3049776 | 7/4/2021 | Africa | Kenya | Kenya |
| Kenya/C103187/2021 | EPI_ISL_3049826 | 7/7/2021 | Africa | Kenya | Kenya |

| | | | | | |
|---|---|---|---|---|---|
| Kenya/C76749/2020 | EPI_ISL_855544 | 12/4/2020 | Africa | Kenya | Kwale |
| Kenya/C85270/2021 | EPI_ISL_1440122 | 2/26/2021 | Africa | Kenya | Kilifi |
| Kenya/C93292/2021 | EPI_ISL_2602597 | 4/20/2021 | Africa | Kenya | Taita Taveta |
| Kenya/C99508/2021 | EPI_ISL_3049432 | 6/7/2021 | Africa | Kenya | Kenya |
| Kenya/CBRD344/2021 | EPI_ISL_2602988 | 4/16/2021 | Africa | Kenya | Nairobi |
| Kenya/K99536/2020 | EPI_ISL_2602824 | 11/9/2020 | Africa | Kenya | Homa Bay |
| Kenya/MAL-561-A/2020 | EPI_ISL_2779506 | 5/30/2020 | Africa | Kenya | Mombasa |
| Kenya/RUS-COV-2687/2021 | EPI_ISL_3031392 | 5/13/2021 | Africa | Kenya | Kisumu |
| Kenya/SS930/2021 | EPI_ISL_3049740 | 6/25/2021 | Africa | Kenya | Kenya |
| Kosovo/ChVir25780_12/2021 | EPI_ISL_2833659 | 6/5/2021 | Europe | Kosovo | Kosovo |
| Kosovo/ChVir25780_24/2021 | EPI_ISL_2833665 | 4/30/2021 | Europe | Kosovo | Kosovo |
| Kosovo/ChVir25780_25/2021 | EPI_ISL_2833646 | 5/2/2021 | Europe | Kosovo | Kosovo |
| Kosovo/ChVir25780_27/2021 | EPI_ISL_2833648 | 6/2/2021 | Europe | Kosovo | Kosovo |
| Kosovo/ChVir25780_6/2021 | EPI_ISL_2833655 | 5/7/2021 | Europe | Kosovo | Kosovo |
| Kuwait/JA-USAFSAM-S4145/2021 | EPI_ISL_3048169 | 6/23/2021 | Asia | Kuwait | Jahra |
| Kuwait/JA-USAFSAM-S4146/2021 | EPI_ISL_3048170 | 6/23/2021 | Asia | Kuwait | Jahra |
| Kuwait/Jaber2120282021/2021 | EPI_ISL_2958695 | 7/7/2021 | Asia | Kuwait | Hawalli |
| Kuwait/JAH3090859/2021 | EPI_ISL_2557251 | 5/30/2021 | Asia | Kuwait | Kuwait |
| Kuwait/JAH3101604/2021 | EPI_ISL_2556117 | 5/31/2021 | Asia | Kuwait | Kuwait |
| Kuwait/KCCC-2301-3615832/2021 | EPI_ISL_2984242 | 7/8/2021 | Asia | Kuwait | Jahra |
| Kuwait/KU-5667/2021 | EPI_ISL_2429129 | 2/1/2021 | Asia | Kuwait | Hawalli |
| Kuwait/KU-5858/2021 | EPI_ISL_2429131 | 2/2/2021 | Asia | Kuwait | Hawalli |
| Kuwait/KU09/2020 | EPI_ISL_416541 | 3/2/2020 | Asia | Kuwait | Kuwait |
| Latvia/029/2020 | EPI_ISL_486412 | 4/29/2020 | Europe | Latvia | Latvia |
| Latvia/046/2020 | EPI_ISL_486430 | 3/22/2020 | Europe | Latvia | Latvia |
| Latvia/1169/2020 | EPI_ISL_1312680 | 10/7/2020 | Europe | Latvia | Latvia |
| Latvia/1172/2020 | EPI_ISL_1312683 | 11/19/2020 | Europe | Latvia | Latvia |

| | | | | | |
|---|---|---|---|---|---|
| Latvia/2104079572/2021 | EPI_ISL_2800190 | 4/21/2021 | Europe | Latvia | Latvia |
| Latvia/2105034949/2021 | EPI_ISL_2798633 | 5/9/2021 | Europe | Latvia | Latvia |
| Latvia/2105069145/2021 | EPI_ISL_2798827 | 5/18/2021 | Europe | Latvia | Latvia |
| Latvia/2120/2021 | EPI_ISL_2141697 | 4/7/2021 | Europe | Latvia | Latvia |
| Latvia/2403/2021 | EPI_ISL_2141965 | 4/26/2021 | Europe | Latvia | Latvia |
| Lebanon/BI_MEIH1/2021 | EPI_ISL_2348487 | 5/3/2021 | Asia | Lebanon | Beirut |
| Lebanon/LAU-CVD-28/2021 | EPI_ISL_2893218 | 7/1/2021 | Asia | Lebanon | Mount Lebanon |
| Lebanon/LAU-CVD-44/2021 | EPI_ISL_3062345 | 6/29/2021 | Asia | Lebanon | South |
| Lebanon/LAU-CVD-45/2021 | EPI_ISL_3062346 | 6/29/2021 | Asia | Lebanon | Mount Lebanon |
| Lebanon/LAU-CVD-88/2021 | EPI_ISL_3233213 | 7/12/2021 | Asia | Lebanon | Lebanon |
| Lebanon/LAU4-53460/2020 | EPI_ISL_637113 | 8/14/2020 | Asia | Lebanon | Beirut |
| Lebanon/LEB-UK-B/2020 | EPI_ISL_1072985 | 12/30/2020 | Asia | Lebanon | Beirut |
| Lebanon/MDC_LAU_16/2021 | EPI_ISL_1910617 | 4/19/2021 | Asia | Lebanon | Tyr |
| Lebanon/QIB-144/2021 | EPI_ISL_3343356 | 1/27/2021 | Asia | Lebanon | Lebanon |
| Lebanon/QIB-786/2021 | EPI_ISL_3343967 | 4/12/2021 | Asia | Lebanon | Lebanon |
| Lebanon/QIB-898/2021 | EPI_ISL_3344067 | 5/4/2021 | Asia | Lebanon | Lebanon |
| Lebanon/QIB-904/2021 | EPI_ISL_3344056 | 4/29/2021 | Asia | Lebanon | Lebanon |
| Liaoning/IVDC-02/2020 | EPI_ISL_498694 | 7/22/2020 | Asia | China | Liaoning |
| Libya/EMC-5/2021 | EPI_ISL_2860640 | 5/30/2021 | Africa | Libya | Libya |
| Libya/EMC-8/2021 | EPI_ISL_2860643 | 6/5/2021 | Africa | Libya | Libya |
| Libya/EMC-9/2021 | EPI_ISL_2860644 | 6/7/2021 | Africa | Libya | Libya |
| Liechtenstein/FL-Risch-2171601300/2021 | EPI_ISL_3388900 | 7/6/2021 | Europe | Liechtenstein | Liechtenstein |
| Liechtenstein/FL-Risch-2171902223/2021 | EPI_ISL_3389002 | 7/15/2021 | Europe | Liechtenstein | Liechtenstein |
| Liechtenstein/FL-UHB-4198371701/2021 | EPI_ISL_1973556 | 4/21/2021 | Europe | Liechtenstein | Liechtenstein |
| Liechtenstein/FL-UHB-4202201501/2021 | EPI_ISL_3344073 | 5/8/2021 | Europe | Liechtenstein | Liechtenstein |
| Liechtenstein/UHB-4168696501/2020 | EPI_ISL_1233663 | 11/16/2020 | Europe | Liechtenstein | Mauren |
| Liechtenstein/UHB-761057800/2021 | EPI_ISL_2768045 | 5/1/2021 | Europe | Liechtenstein | Liechtenstein |

| | | | | | |
|---|---|---|---|---|---|
| Lithuania/IBT-LSC-VU_r15_13/2021 | EPI_ISL_2885903 | 6/13/2021 | Europe | Lithuania | Alytaus Apskritis |
| Lithuania/ID3369/2020 | EPI_ISL_934070 | 12/10/2020 | Europe | Lithuania | Vilniaus Apskritis |
| Lithuania/LSMULKKGMMK14C63/2021 | EPI_ISL_3481787 | 7/12/2021 | Europe | Lithuania | Kauno Apskritis |
| Lithuania/LSMULKKGMMK17C124/2021 | EPI_ISL_3464873 | 8/5/2021 | Europe | Lithuania | Marijampoles Apskritis |
| Lithuania/LSMULKKGMMK17C166/2021 | EPI_ISL_3464851 | 8/6/2021 | Europe | Lithuania | Kauno Apskritis |
| Lithuania/LSMULKKGMMK4C33/2021 | EPI_ISL_1357643 | 2/9/2021 | Europe | Lithuania | Kauno Apskritis |
| Lithuania/LTU000_KUL_005136212/2021 | EPI_ISL_2082466 | 4/21/2021 | Europe | Lithuania | Klaipedos Apskritis |
| Lithuania/LTU000_NMRVI_123978/2021 | EPI_ISL_2693996 | 6/2/2021 | Europe | Lithuania | Vilniaus Apskritis |
| Lithuania/MR-LUHS-Eilnr192/2020 | EPI_ISL_636842 | 7/28/2020 | Europe | Lithuania | Kauno Apskritis |
| Lithuania/S21C40/2021 | EPI_ISL_1324593 | 2/15/2021 | Europe | Lithuania | Kauno Apskritis |
| Lithuania/S21E1157/2021 | EPI_ISL_2495920 | 5/19/2021 | Europe | Lithuania | Siauliu Apskritis |
| Lithuania/S21E651/2021 | EPI_ISL_2428591 | 4/25/2021 | Europe | Lithuania | Vilniaus Apskritis |
| Lithuania/S21E856/2021 | EPI_ISL_2428908 | 5/10/2021 | Europe | Lithuania | Vilniaus Apskritis |
| Lithuania/S21G130/2021 | EPI_ISL_3060068 | 7/3/2021 | Europe | Lithuania | Vilnius |
| Lithuania/T_15/2020 | EPI_ISL_904938 | 9/26/2020 | Europe | Lithuania | Vilnius |
| Luxembourg/LNS0489651/2021 | EPI_ISL_3334749 | 7/28/2021 | Europe | Luxembourg | Luxembourg |
| Luxembourg/LNS1903466/2021 | EPI_ISL_3334842 | 8/2/2021 | Europe | Luxembourg | Luxembourg |
| Luxembourg/LNS1956201/2020 | EPI_ISL_740028 | 8/3/2020 | Europe | Luxembourg | Luxembourg |
| Luxembourg/LNS3227270/2021 | EPI_ISL_2401686 | 5/18/2021 | Europe | Luxembourg | Luxembourg |
| Luxembourg/LNS3411274/2021 | EPI_ISL_3147200 | 7/2/2021 | Europe | Luxembourg | Luxembourg |
| Luxembourg/LNS4779946/2021 | EPI_ISL_2401288 | 4/26/2021 | Europe | Luxembourg | Luxembourg |
| Luxembourg/LNS4978842/2021 | EPI_ISL_1916588 | 4/22/2021 | Europe | Luxembourg | Luxembourg |
| Luxembourg/LNS5100176/2021 | EPI_ISL_2401150 | 5/14/2021 | Europe | Luxembourg | Luxembourg |
| Luxembourg/LNS6279788/2021 | EPI_ISL_3145700 | 6/1/2021 | Europe | Luxembourg | Luxembourg |
| Luxembourg/LNS6542216/2021 | EPI_ISL_3147550 | 6/30/2021 | Europe | Luxembourg | Luxembourg |
| Luxembourg/LNS6961222/2021 | EPI_ISL_3334633 | 8/2/2021 | Europe | Luxembourg | Luxembourg |
| Luxembourg/LNS7441642/2021 | EPI_ISL_1383384 | 2/1/2021 | Europe | Luxembourg | Luxembourg |

| | | | | | |
|---|---|---|---|---|---|
| Luxembourg/LNS9643402/2020 | EPI_ISL_744958 | 7/30/2020 | Europe | Luxembourg | Luxembourg |
| Madagascar/IPM-01116/2021 | EPI_ISL_1660308 | 1/23/2021 | Africa | Madagascar | Analamanga |
| Madagascar/IPM-01917/2021 | EPI_ISL_1660273 | 2/15/2021 | Africa | Madagascar | Diana |
| Madagascar/IPM-38708/2020 | EPI_ISL_1660315 | 11/12/2020 | Africa | Madagascar | Boeny |
| Madagascar/IPM-40419/2020 | EPI_ISL_1660234 | 12/26/2020 | Africa | Madagascar | Boeny |
| Madagascar/VIRO-2722/2020 | EPI_ISL_677635 | 3/26/2020 | Africa | Madagascar | Toamasina |
| Malawi/CERI-KRISP-K015146/2021 | EPI_ISL_2686020 | 2/22/2021 | Africa | Malawi | Malawi |
| Malawi/CERI-KRISP-K015281/2021 | EPI_ISL_2494966 | 4/30/2021 | Africa | Malawi | Malawi |
| Malawi/CERI-KRISP-K015290/2021 | EPI_ISL_2494972 | 4/30/2021 | Africa | Malawi | Malawi |
| Malaysia/5760/2020 | EPI_ISL_506999 | 3/26/2020 | Asia | Malaysia | Kuala Lumpur |
| Malaysia/6359/2020 | EPI_ISL_501220 | 2/25/2020 | Asia | Malaysia | Kuala Lumpur |
| Malaysia/7924/2020 | EPI_ISL_507000 | 4/10/2020 | Asia | Malaysia | Kuala Lumpur |
| Malaysia/IIUM101/2021 | EPI_ISL_3241561 | 7/1/2021 | Asia | Malaysia | Pahang |
| Malaysia/IIUM5755/2021 | EPI_ISL_2622047 | 4/24/2021 | Asia | Malaysia | Kelantan |
| Malaysia/IIUM5770/2021 | EPI_ISL_2622088 | 4/28/2021 | Asia | Malaysia | Kelantan |
| Malaysia/IMR_025404/2021 | EPI_ISL_2812576 | 6/8/2021 | Asia | Malaysia | Malaysia |
| Malaysia/IMR_065549/2021 | EPI_ISL_2811998 | 6/3/2021 | Asia | Malaysia | Malaysia |
| Malaysia/IMR_75201/2021 | EPI_ISL_2649998 | 5/17/2021 | Asia | Malaysia | Malaysia |
| Malaysia/IMR_G194145/2021 | EPI_ISL_2549562 | 5/25/2021 | Asia | Malaysia | Malaysia |
| Malaysia/IMR_WC451949/2021 | EPI_ISL_3446659 | 7/26/2021 | Asia | Malaysia | Malaysia |
| Malaysia/MGI_DNALAB-DM210400130/2021 | EPI_ISL_2342548 | 4/1/2021 | Asia | Malaysia | Selangor |
| Maldives/MAV00499/2020 | EPI_ISL_3275380 | 4/11/2020 | Asia | Maldives | Maldives |
| Maldives/MAV01979/2020 | EPI_ISL_3275385 | 6/11/2020 | Asia | Maldives | Maldives |
| Maldives/MAV09481/2020 | EPI_ISL_3275392 | 9/16/2020 | Asia | Maldives | Maldives |
| Maldives/MAV14010/2021 | EPI_ISL_3452719 | 1/7/2021 | Asia | Maldives | Maldives |
| Maldives/MAV17718/2021 | EPI_ISL_3452720 | 2/12/2021 | Asia | Maldives | Maldives |
| Maldives/MAV22405/2021 | EPI_ISL_3275175 | 3/20/2021 | Asia | Maldives | MalV© |

| | | | | | |
|---|---|---|---|---|---|
| Maldives/MAV28931/2021 | EPI_ISL_3275166 | 4/27/2021 | Asia | Maldives | Rasdhoo |
| Maldives/MAV29502/2021 | EPI_ISL_3275174 | 4/29/2021 | Asia | Maldives | Mal√© |
| Maldives/MAV31878/2021 | EPI_ISL_3275167 | 5/3/2021 | Asia | Maldives | Himmafushi |
| Maldives/MAV36627/2021 | EPI_ISL_3275170 | 5/8/2021 | Asia | Maldives | Maafushi |
| Mali/UCRC-31/2020 | EPI_ISL_812967 | 3/31/2020 | Africa | Mali | Mali |
| Mali/UCRC-68/2020 | EPI_ISL_812966 | 3/18/2020 | Africa | Mali | Mali |
| Malta/BAL-Sliema-7/2020 | EPI_ISL_576123 | 9/5/2020 | Europe | Malta | Malta |
| Malta/BAL-Sliema-8/2020 | EPI_ISL_576124 | 9/7/2020 | Europe | Malta | Malta |
| Malta/MDxMDH371/2021 | EPI_ISL_2101358 | 4/18/2021 | Europe | Malta | Malta |
| Malta/MDxMDH409/2021 | EPI_ISL_2105939 | 4/27/2021 | Europe | Malta | Malta |
| Malta/MDxMDH418/2021 | EPI_ISL_2105948 | 5/1/2021 | Europe | Malta | Malta |
| Malta/MDxMDH489/2021 | EPI_ISL_2718542 | 6/21/2021 | Europe | Malta | Malta |
| Malta/MDxMDH494/2021 | EPI_ISL_2854655 | 5/27/2021 | Europe | Malta | Malta |
| Malta/MDxMDH506/2021 | EPI_ISL_2970376 | 6/30/2021 | Europe | Malta | Malta |
| Malta/MDxMDH527/2021 | EPI_ISL_2988655 | 7/7/2021 | Europe | Malta | Malta |
| Malta/MDxMDH528/2021 | EPI_ISL_2988656 | 7/7/2021 | Europe | Malta | Malta |
| Mauritius/104633/2021 | EPI_ISL_2657333 | 4/28/2021 | Africa | Mauritius | Savanne |
| Mauritius/104657/2021 | EPI_ISL_2657337 | 5/4/2021 | Africa | Mauritius | Grand Port |
| Mauritius/125759/2021 | EPI_ISL_2657341 | 4/29/2021 | Africa | Mauritius | Savanne |
| Mauritius/156377/2021 | EPI_ISL_3231396 | 5/9/2021 | Africa | Mauritius | Plaine-Wilhems |
| Mauritius/248302/2021 | EPI_ISL_2834926 | 6/17/2021 | Africa | Mauritius | Mauritius |
| Mauritius/248310/2021 | EPI_ISL_2834928 | 6/17/2021 | Africa | Mauritius | Mauritius |
| Mauritius/N4272/2021 | EPI_ISL_2285196 | 3/11/2021 | Africa | Mauritius | Plaine-Wilhems |
| Mauritius/P-001880/2020 | EPI_ISL_2499903 | 10/23/2020 | Africa | Mauritius | Grand Port |
| Mauritius/P-014863/2020 | EPI_ISL_2499905 | 11/9/2020 | Africa | Mauritius | Grand Port |
| Mexico/AGU_IBT_IMSS_1490/2021 | EPI_ISL_2681293 | 6/1/2021 | North America | Mexico | Aguascalientes |
| Mexico/AGU_LANGEBIO_IMSS_0658/2021 | EPI_ISL_2402074 | 4/19/2021 | North America | Mexico | Aguascalientes |

| | | | | | |
|---|---|---|---|---|---|
| Mexico/AGU_LANGEBIO_IMSS_0711/2021 | EPI_ISL_2402125 | 4/29/2021 | North America | Mexico | Aguascalientes |
| Mexico/AGU-IBT-IMSS-446/2020 | EPI_ISL_1301631 | 4/8/2020 | North America | Mexico | Aguascalientes |
| Mexico/AGU-InDRE_FB16010_S2656/2021 | EPI_ISL_2495924 | 5/3/2021 | North America | Mexico | Aguascalientes |
| Mexico/AGU-InDRE_FB18596_S4465/2021 | EPI_ISL_2937902 | 6/25/2021 | North America | Mexico | Aguascalientes |
| Mexico/AGU-InDRE_FB19720_S4911/2021 | EPI_ISL_3265407 | 7/1/2021 | North America | Mexico | Aguascalientes |
| Mexico/AGU-InDRE_FB22306_S5488/2021 | EPI_ISL_3460204 | 7/19/2021 | North America | Mexico | Aguascalientes |
| Mexico/AGU-InDRE-55/2020 | EPI_ISL_576258 | 6/15/2020 | North America | Mexico | Aguascalientes |
| Mexico/AGU-InDRE-58/2020 | EPI_ISL_576261 | 8/7/2020 | North America | Mexico | Aguascalientes |
| Mexico/AGU-InDRE-IBT-46/2020 | EPI_ISL_1301685 | 4/13/2020 | North America | Mexico | Aguascalientes |
| Mexico/AGU-LANGEBIO_IMSS_0995/2021 | EPI_ISL_2671483 | 5/17/2021 | North America | Mexico | Aguascalientes |
| Mexico/BCN-ALSR-105346/2021 | EPI_ISL_3374007 | 6/29/2021 | North America | Mexico | Baja California |
| Mexico/BCN-ALSR-2457/2020 | EPI_ISL_635485 | 5/16/2020 | North America | Mexico | Baja California |
| Mexico/BCN-ALSR-4896/2020 | EPI_ISL_730200 | 7/7/2020 | North America | Mexico | Baja California |
| Mexico/BCN-ALSR-6612/2020 | EPI_ISL_1081451 | 12/23/2020 | North America | Mexico | Baja California |
| Mexico/BCN-ALSR-8461/2020 | EPI_ISL_1531924 | 10/2/2020 | North America | Mexico | Baja California |
| Mexico/BCN-InDRE_FB13740_S1819/2021 | EPI_ISL_2139914 | 4/20/2021 | North America | Mexico | Baja California |
| Mexico/BCN-LANGEBIO_IMSS_1802/2021 | EPI_ISL_2942877 | 6/29/2021 | North America | Mexico | Baja California |
| Mexico/BCN-SEARCH-104087/2021 | EPI_ISL_3215964 | 4/20/2021 | North America | Mexico | Baja California |
| Mexico/BCN-SEARCH-104122/2021 | EPI_ISL_3215989 | 2/16/2021 | North America | Mexico | Baja California |
| Mexico/BCN-SEARCH-104127/2021 | EPI_ISL_3215993 | 7/6/2021 | North America | Mexico | Baja California |

| | | | | | |
|---|---|---|---|---|---|
| Mexico/BCN-SEARCH-104138/2021 | EPI_ISL_3216000 | 7/6/2021 | North America | Mexico | Baja California |
| Mexico/BCN-SEARCH-7391/2021 | EPI_ISL_1295691 | 2/12/2021 | North America | Mexico | Baja California |
| Mexico/BCN-SEARCH-7577/2021 | EPI_ISL_1295837 | 2/2/2021 | North America | Mexico | Baja California |
| Mexico/BCS_IBT_IMSS_1026/2021 | EPI_ISL_1811531 | 4/10/2021 | North America | Mexico | Baja California Sur |
| Mexico/BCS_IBT_IMSS_1574/2021 | EPI_ISL_2681297 | 5/24/2021 | North America | Mexico | Baja California Sur |
| Mexico/BCS-IBT-IMSS-20/2020 | EPI_ISL_955255 | 4/6/2020 | North America | Mexico | Baja California Sur |
| Mexico/BCS-IBT-IMSS-252/2021 | EPI_ISL_1288440 | 2/12/2021 | North America | Mexico | Baja California Sur |
| Mexico/BCS-InDRE_326/2020 | EPI_ISL_1060747 | 7/31/2020 | North America | Mexico | Baja California Sur |
| Mexico/BCS-InDRE_337/2020 | EPI_ISL_1060748 | 7/31/2020 | North America | Mexico | Baja California Sur |
| Mexico/BCS-InDRE_470/2021 | EPI_ISL_1168540 | 1/17/2021 | North America | Mexico | Baja California Sur |
| Mexico/BCS-InDRE_F11109_S606/2021 | EPI_ISL_1337392 | 2/27/2021 | North America | Mexico | Baja California Sur |
| Mexico/BCS-InDRE_FB13293_S1612/2021 | EPI_ISL_1821184 | 4/10/2021 | North America | Mexico | Baja California Sur |
| Mexico/BCS-InDRE_FB13624_S1818/2021 | EPI_ISL_2139913 | 4/17/2021 | North America | Mexico | Baja California Sur |
| Mexico/BCS-InDRE_FB14099_S1990/2021 | EPI_ISL_2158231 | 4/22/2021 | North America | Mexico | Baja California Sur |
| Mexico/BCS-InDRE_FB16340_S2822/2021 | EPI_ISL_2545730 | 5/22/2021 | North America | Mexico | Baja California Sur |
| Mexico/BCS-InDRE_FB19255_S4502/2021 | EPI_ISL_3033356 | 6/23/2021 | North America | Mexico | Baja California Sur |
| Mexico/BCS-InDRE_FB19273_S4573/2021 | EPI_ISL_3033426 | 6/27/2021 | North America | Mexico | Baja California Sur |
| Mexico/BCS-InDRE_FB21427_S5030/2021 | EPI_ISL_3265526 | 7/9/2021 | North America | Mexico | Baja California Sur |
| Mexico/BCS-INER_IMSS_1585/2021 | EPI_ISL_3155595 | 7/5/2021 | North America | Mexico | Baja California Sur |
| Mexico/CAM_IBT_IMSS_2153/2021 | EPI_ISL_3347538 | 7/13/2021 | North America | Mexico | Campeche |

| | | | | | |
|---|---|---|---|---|---|
| Mexico/CAM_INER_IMSS_1107/2021 | EPI_ISL_2490468 | 4/27/2021 | North America | Mexico | Campeche |
| Mexico/CAM_INER_IMSS_1119/2021 | EPI_ISL_2490418 | 4/29/2021 | North America | Mexico | Campeche |
| Mexico/CAM_INER_IMSS_1150/2021 | EPI_ISL_2490485 | 5/10/2021 | North America | Mexico | Campeche |
| Mexico/CAM-InDRE_544/2021 | EPI_ISL_1168614 | 1/17/2021 | North America | Mexico | Campeche |
| Mexico/CAM-InDRE_F11511_S956/2021 | EPI_ISL_1483066 | 3/8/2021 | North America | Mexico | Campeche |
| Mexico/CAM-InDRE_FB14881_S2388/2021 | EPI_ISL_2455913 | 5/10/2021 | North America | Mexico | Campeche |
| Mexico/CAM-InDRE_FB18130_S4024/2021 | EPI_ISL_2859038 | 6/17/2021 | North America | Mexico | Campeche |
| Mexico/CAM-InDRE_FB21561_S5066/2021 | EPI_ISL_3265562 | 7/5/2021 | North America | Mexico | Campeche |
| Mexico/CAM-InDRE-IBT-48/2020 | EPI_ISL_1301687 | 5/8/2020 | North America | Mexico | Campeche |
| Mexico/CAM-LANGEBIO_IMSS_1518/2021 | EPI_ISL_2942631 | 6/23/2021 | North America | Mexico | Campeche |
| Mexico/CHH_IBT_IMSS_1406/2021 | EPI_ISL_2681228 | 5/27/2021 | North America | Mexico | Chihuahua |
| Mexico/CHH_IBT_IMSS_1430/2021 | EPI_ISL_2681289 | 6/2/2021 | North America | Mexico | Chihuahua |
| Mexico/CHH_IBT_IMSS_2311/2021 | EPI_ISL_3347627 | 7/12/2021 | North America | Mexico | Chihuahua |
| Mexico/CHH_IBT_IMSS_2328/2021 | EPI_ISL_3347640 | 7/13/2021 | North America | Mexico | Chihuahua |
| Mexico/CHH_LANGEBIO_IMSS_0578/2021 | EPI_ISL_2401998 | 4/25/2021 | North America | Mexico | Chihuahua |
| Mexico/CHH-IBT-IMSS-439/2020 | EPI_ISL_1301552 | 4/6/2020 | North America | Mexico | Chihuahua |
| Mexico/CHH-InDRE_366/2020 | EPI_ISL_1060678 | 12/16/2020 | North America | Mexico | Chihuahua |
| Mexico/CHH-InDRE_F11009_S866/2021 | EPI_ISL_1399271 | 2/22/2021 | North America | Mexico | Chihuahua |
| Mexico/CHH-InDRE_F11012_S819/2021 | EPI_ISL_1424021 | 2/23/2021 | North America | Mexico | Chiapas |
| Mexico/CHH-InDRE_FB13831_S1951/2021 | EPI_ISL_2158193 | 4/21/2021 | North America | Mexico | Chihuahua |

| | | | | | |
|---|---|---|---|---|---|
| Mexico/CHH-InDRE_FB14901_S2402/2021 | EPI_ISL_2455927 | 5/5/2021 | North America | Mexico | Chihuahua |
| Mexico/CHH-LANGEBIO_IMSS_1693/2021 | EPI_ISL_2942782 | 6/29/2021 | North America | Mexico | Chihuahua |
| Mexico/CHP_IBT_IMSS_2456/2021 | EPI_ISL_3347548 | 7/19/2021 | North America | Mexico | Chiapas |
| Mexico/CHP_IBT_IMSS_2530/2021 | EPI_ISL_3347549 | 7/19/2021 | North America | Mexico | Chiapas |
| Mexico/CHP_INER_IMSS_00947/2021 | EPI_ISL_2091391 | 4/13/2021 | North America | Mexico | Chiapas |
| Mexico/CHP_LANGEBIO_IMSS_0816/2021 | EPI_ISL_2402226 | 4/27/2021 | North America | Mexico | Chiapas |
| Mexico/CHP-IBT-IMSS-119/2021 | EPI_ISL_1288327 | 2/11/2021 | North America | Mexico | Chiapas |
| Mexico/CHP-IBT-IMSS-825/2021 | EPI_ISL_1416631 | 3/6/2021 | North America | Mexico | Chiapas |
| Mexico/CHP-InDRE_132/2020 | EPI_ISL_913935 | 11/20/2020 | North America | Mexico | Chiapas |
| Mexico/CHP-InDRE_443/2021 | EPI_ISL_1168521 | 1/18/2021 | North America | Mexico | Chiapas |
| Mexico/CHP-InDRE_FB14079_S2113/2021 | EPI_ISL_2296085 | 4/22/2021 | North America | Mexico | Chiapas |
| Mexico/CHP-InDRE_FB14861_S2552/2021 | EPI_ISL_2479885 | 5/3/2021 | North America | Mexico | Chiapas |
| Mexico/CHP-InDRE_FB15965_S2603/2021 | EPI_ISL_2492494 | 5/16/2021 | North America | Mexico | Chiapas |
| Mexico/CHP-InDRE_FB18388_S4345/2021 | EPI_ISL_2937807 | 6/23/2021 | North America | Mexico | Chiapas |
| Mexico/CHP-InDRE_FB19059_S4788/2021 | EPI_ISL_3033610 | 6/28/2021 | North America | Mexico | Chiapas |
| Mexico/CHP-InDRE-IBT-196/2020 | EPI_ISL_1302310 | 12/21/2020 | North America | Mexico | Oaxaca |
| Mexico/CMX-IMSS_01/2020 | EPI_ISL_424731 | 3/13/2020 | North America | Mexico | Chihuahua |
| Mexico/CMX-InDRE_FB14732_S2314/2021 | EPI_ISL_2342990 | 4/29/2021 | North America | Mexico | Mexico City |
| Mexico/CMX-InDRE_FB17931_S4374/2021 | EPI_ISL_2937840 | 6/9/2021 | North America | Mexico | Mexico City |
| Mexico/CMX-InDRE-06/2020 | EPI_ISL_424673 | 3/12/2020 | North America | Mexico | Mexico City |

| | | | | | |
|---|---|---|---|---|---|
| Mexico/CMX-InDRE-IBT-115/2020 | EPI_ISL_1301583 | 5/4/2020 | North America | Mexico | Mexico City |
| Mexico/CMX-INER-0177/2020 | EPI_ISL_837773 | 7/17/2020 | North America | Mexico | Mexico City |
| Mexico/CMX-INER-04/2020 | EPI_ISL_424626 | 3/15/2020 | North America | Mexico | Mexico City |
| Mexico/CMX-INER-IBT-0333/2020 | EPI_ISL_2839340 | 10/1/2020 | North America | Mexico | Mexico City |
| Mexico/CMX-INER-IBT-110/2020 | EPI_ISL_1302334 | 12/31/2020 | North America | Mexico | Mexico City |
| Mexico/CMX-INER-IBT-26/2020 | EPI_ISL_1301522 | 5/3/2020 | North America | Mexico | Mexico City |
| Mexico/CMX-INER-IBT-34/NC/2020 | EPI_ISL_3463587 | 3/29/2020 | North America | Mexico | Mexico City |
| Mexico/CMX-INER-IBT-70/2020 | EPI_ISL_1302369 | 11/18/2020 | North America | Mexico | Mexico City |
| Mexico/CMX-INER-INMEGEN-00072/2021 | EPI_ISL_1824426 | 2/19/2021 | North America | Mexico | Mexico City |
| Mexico/CMX-INMEGEN-02-01-15/2021 | EPI_ISL_1040607 | 1/27/2021 | North America | Mexico | Mexico City |
| Mexico/CMX-INMEGEN-02-03-18/2021 | EPI_ISL_1040643 | 1/27/2021 | North America | Mexico | Mexico City |
| Mexico/CMX-INMEGEN-03-08-49/2021 | EPI_ISL_1315524 | 3/4/2021 | North America | Mexico | Mexico City |
| Mexico/CMX-INMEGEN-03-11-326/2021 | EPI_ISL_1591667 | 3/29/2021 | North America | Mexico | Mexico City |
| Mexico/CMX-INMEGEN-04-04-63/2021 | EPI_ISL_1628571 | 4/7/2021 | North America | Mexico | Mexico City |
| Mexico/CMX-INMEGEN-05-01-3/2021 | EPI_ISL_2230926 | 4/29/2021 | North America | Mexico | Mexico City |
| Mexico/CMX-INMEGEN-05-06-182/2021 | EPI_ISL_2603673 | 5/25/2021 | North America | Mexico | Mexico City |
| Mexico/CMX-INMEGEN-05-06-31/2021 | EPI_ISL_2603618 | 5/26/2021 | North America | Mexico | Mexico City |
| Mexico/CMX-INMEGEN-16-225/2021 | EPI_ISL_2978545 | 7/4/2021 | North America | Mexico | Mexico City |
| Mexico/CMX-INMEGEN-17-282/2021 | EPI_ISL_3067732 | 7/15/2021 | North America | Mexico | Mexico City |
| Mexico/CMX-LANGEBIO_IMSS_1719/2021 | EPI_ISL_2942804 | 6/18/2021 | North America | Mexico | Mexico City |

| | | | | | |
|---|---|---|---|---|---|
| Mexico/COA_IBT_IMSS_2340/2021 | EPI_ISL_3347650 | 7/14/2021 | North America | Mexico | Coahuila |
| Mexico/COA_LANGEBIO_IMSS_0269/2021 | EPI_ISL_1662042 | 3/30/2021 | North America | Mexico | Coahuila |
| Mexico/COA_LANGEBIO_IMSS_0284/2021 | EPI_ISL_1662048 | 4/1/2021 | North America | Mexico | Coahuila |
| Mexico/COA_LANGEBIO_IMSS_0514/2021 | EPI_ISL_2401941 | 4/19/2021 | North America | Mexico | Coahuila |
| Mexico/COA_LANGEBIO_IMSS_0592/2021 | EPI_ISL_2402011 | 4/17/2021 | North America | Mexico | Coahuila |
| Mexico/COA-InDRE_40/2020 | EPI_ISL_516620 | 6/19/2020 | North America | Mexico | Coahuila |
| Mexico/COA-InDRE_FB16761_S3202/2021 | EPI_ISL_2674689 | 6/9/2021 | North America | Mexico | Coahuila |
| Mexico/COA-InDRE_FB16772_S2777/2021 | EPI_ISL_2533803 | 5/24/2021 | North America | Mexico | Coahuila |
| Mexico/COA-InDRE_FB17711_S4112/2021 | EPI_ISL_2920683 | 6/8/2021 | North America | Mexico | Coahuila |
| Mexico/COA-InDRE-36/2020 | EPI_ISL_516618 | 6/13/2020 | North America | Mexico | Coahuila |
| Mexico/COA-InDRE-IBT-28845/NC/2020 | EPI_ISL_3463608 | 5/31/2020 | North America | Mexico | Coahuila |
| Mexico/COA-INER-IMSS-00244/2021 | EPI_ISL_1279507 | 2/15/2021 | North America | Mexico | Coahuila |
| Mexico/COA-LANGEBIO_IMSS_0873/2021 | EPI_ISL_2671545 | 5/23/2021 | North America | Mexico | Coahuila |
| Mexico/COA-LANGEBIO_IMSS_1691/2021 | EPI_ISL_2942780 | 7/1/2021 | North America | Mexico | Coahuila |
| Mexico/COL_IBT_IMSS_2189/2021 | EPI_ISL_3347813 | 7/12/2021 | North America | Mexico | Colima |
| Mexico/COL_INER_IMSS_00519/2021 | EPI_ISL_1585522 | 3/15/2021 | North America | Mexico | Colima |
| Mexico/COL_LANGEBIO_IMSS_0738/2021 | EPI_ISL_2402149 | 4/20/2021 | North America | Mexico | Colima |
| Mexico/COL_LANGEBIO_IMSS_0754/2021 | EPI_ISL_2402165 | 4/27/2021 | North America | Mexico | Colima |
| Mexico/COL-IBT_IMSS_1847/2021 | EPI_ISL_2801720 | 6/2/2021 | North America | Mexico | Colima |
| Mexico/COL-IBT-IMSS-461/2020 | EPI_ISL_1301644 | 4/24/2020 | North America | Mexico | Colima |

| | | | | | |
|---|---|---|---|---|---|
| Mexico/COL-InDRE_FB11127_S1721/2021 | EPI_ISL_1857276 | 4/5/2021 | North America | Mexico | Colima |
| Mexico/COL-InDRE_FB19288_S4580/2021 | EPI_ISL_3033433 | 6/25/2021 | North America | Mexico | Colima |
| Mexico/COL-InDRE_FB23257_S5392/2021 | EPI_ISL_3460007 | 7/20/2021 | North America | Mexico | Colima |
| Mexico/COL-InDRE-IBT-137/2020 | EPI_ISL_1301482 | 7/8/2020 | North America | Mexico | Colima |
| Mexico/COL-InDRE-IBT-20796/NC/2020 | EPI_ISL_3463595 | 6/1/2020 | North America | Mexico | Colima |
| Mexico/COL-LANGEBIO_IMSS_1020/2021 | EPI_ISL_2671552 | 5/22/2021 | North America | Mexico | Colima |
| Mexico/COL-LANGEBIO_IMSS_1045/2021 | EPI_ISL_2671556 | 5/29/2021 | North America | Mexico | Colima |
| Mexico/DUR_IBT_IMSS_2318/2021 | EPI_ISL_3347632 | 7/14/2021 | North America | Mexico | Durango |
| Mexico/DUR_IBT_IMSS_2339/2021 | EPI_ISL_3347649 | 7/16/2021 | North America | Mexico | Durango |
| Mexico/DUR_INER_IMSS_00408/2021 | EPI_ISL_1585417 | 3/16/2021 | North America | Mexico | Durango |
| Mexico/DUR_LANGEBIO_IMSS_0522/2021 | EPI_ISL_2401949 | 4/20/2021 | North America | Mexico | Durango |
| Mexico/DUR_LANGEBIO_IMSS_0551/2021 | EPI_ISL_2401974 | 4/24/2021 | North America | Mexico | Durango |
| Mexico/DUR_LANGEBIO_IMSS_61232-NC/2021 | EPI_ISL_2969933 | 5/18/2021 | North America | Mexico | Durango |
| Mexico/DUR-InDRE_FB16735_S2904/2021 | EPI_ISL_2545805 | 5/26/2021 | North America | Mexico | Durango |
| Mexico/DUR-InDRE_FB17384_S3445/2021 | EPI_ISL_2800934 | 6/9/2021 | North America | Mexico | Durango |
| Mexico/DUR-InDRE-09/2020 | EPI_ISL_455432 | 3/11/2020 | North America | Mexico | Durango |
| Mexico/DUR-INER-IMSS-00228/2021 | EPI_ISL_1279493 | 2/15/2021 | North America | Mexico | Durango |
| Mexico/DUR-LANGEBIO_IMSS_1684/2021 | EPI_ISL_2942775 | 6/30/2021 | North America | Mexico | Durango |
| Mexico/GRO_IBT_IMSS_1017/2021 | EPI_ISL_1811523 | 4/9/2021 | North America | Mexico | Guerrero |
| Mexico/GRO_IBT_IMSS_2549/2021 | EPI_ISL_3347807 | 7/23/2021 | North America | Mexico | Guerrero |

| | | | | | |
|---|---|---|---|---|---|
| Mexico/GRO_LANGEBIO_IMSS_03966-NC/2021 | EPI_ISL_2969934 | 4/21/2021 | North America | Mexico | Guerrero |
| Mexico/GRO_LANGEBIO_IMSS_0806/2021 | EPI_ISL_2402216 | 4/22/2021 | North America | Mexico | Guerrero |
| Mexico/GRO-IBT-IMSS-10/2020 | EPI_ISL_955245 | 4/8/2020 | North America | Mexico | Guerrero |
| Mexico/GRO-IBT-IMSS-26/2020 | EPI_ISL_955236 | 4/17/2020 | North America | Mexico | Guerrero |
| Mexico/GRO-IBT-IMSS-824/2021 | EPI_ISL_1416630 | 3/6/2021 | North America | Mexico | Guerrero |
| Mexico/GRO-InDRE_136/2020 | EPI_ISL_913939 | 11/26/2020 | North America | Mexico | Guerrero |
| Mexico/GRO-InDRE_284/2020 | EPI_ISL_1054946 | 8/15/2020 | North America | Mexico | Guerrero |
| Mexico/GRO-InDRE_384/2020 | EPI_ISL_1054991 | 7/26/2020 | North America | Mexico | Guerrero |
| Mexico/GRO-InDRE_FB16449_S3711/2021 | EPI_ISL_2779214 | 5/23/2021 | North America | Mexico | Guerrero |
| Mexico/GRO-InDRE_FB17786_S4179/2021 | EPI_ISL_2920750 | 6/8/2021 | North America | Mexico | Guerrero |
| Mexico/GRO-InDRE-10/2020 | EPI_ISL_455434 | 3/13/2020 | North America | Mexico | Guerrero |
| Mexico/GRO-InDRE-93/2020 | EPI_ISL_658901 | 9/7/2020 | North America | Mexico | Guerrero |
| Mexico/GRO-InDRE-IBT-132/2020 | EPI_ISL_1301523 | 6/10/2020 | North America | Mexico | Guerrero |
| Mexico/GRO-LANGEBIO_IMSS_1051/2021 | EPI_ISL_2671561 | 5/13/2021 | North America | Mexico | Guerrero |
| Mexico/GRO-LANGEBIO_IMSS_1838/2021 | EPI_ISL_2942906 | 6/30/2021 | North America | Mexico | Guerrero |
| Mexico/GRO-LANGEBIO_IMSS_1872/2021 | EPI_ISL_2942933 | 7/1/2021 | North America | Mexico | Guerrero |
| Mexico/GUA_INER_IMSS_00860/2021 | EPI_ISL_2091311 | 4/13/2021 | North America | Mexico | Guanajuato |
| Mexico/GUA-InDRE_306/2020 | EPI_ISL_1060741 | 8/4/2020 | North America | Mexico | Guanajuato |
| Mexico/GUA-InDRE_F10376_S662/2021 | EPI_ISL_1359077 | 2/15/2021 | North America | Mexico | Guanajuato |
| Mexico/GUA-InDRE_F10379_S638/2021 | EPI_ISL_1340647 | 2/16/2021 | North America | Mexico | Guanajuato |

| | | | | | |
|---|---|---|---|---|---|
| Mexico/GUA-InDRE_FB13899_S1978/2021 | EPI_ISL_2158220 | 4/21/2021 | North America | Mexico | Guanajuato |
| Mexico/GUA-InDRE_FB13922_S2263/2021 | EPI_ISL_2362691 | 4/22/2021 | North America | Mexico | Guanajuato |
| Mexico/GUA-InDRE_FB16465_S3477/2021 | EPI_ISL_2778992 | 5/26/2021 | North America | Mexico | Guanajuato |
| Mexico/GUA-InDRE_FB17346_S3533/2021 | EPI_ISL_2779048 | 6/11/2021 | North America | Mexico | Guanajuato |
| Mexico/GUA-InDRE_FB17350_S3979/2021 | EPI_ISL_2858993 | 6/11/2021 | North America | Mexico | Guanajuato |
| Mexico/GUA-InDRE_FB24015_S5479/2021 | EPI_ISL_3460196 | 7/19/2021 | North America | Mexico | Guanajuato |
| Mexico/GUA-InDRE-FB14809-S2124/2021 | EPI_ISL_2361362 | 5/4/2021 | North America | Mexico | Guanajuato |
| Mexico/GUA-INER_IMSS_1474/2021 | EPI_ISL_3155484 | 7/8/2021 | North America | Mexico | Guanajuato |
| Mexico/HID-21-053-007-41160/2020 | EPI_ISL_2795658 | 11/24/2020 | North America | Mexico | Hidalgo |
| Mexico/HID-IBT_IMSS_2047/2021 | EPI_ISL_2801891 | 6/11/2021 | North America | Mexico | Hidalgo |
| Mexico/HID-IBT-IMSS-506/2020 | EPI_ISL_1301463 | 5/16/2020 | North America | Mexico | Hidalgo |
| Mexico/HID-IBT-IMSS-508/2020 | EPI_ISL_1301674 | 5/18/2020 | North America | Mexico | Hidalgo |
| Mexico/HID-InDRE_309/2020 | EPI_ISL_1060700 | 7/23/2020 | North America | Mexico | Hidalgo |
| Mexico/HID-InDRE_FB13033_S1670/2021 | EPI_ISL_1857233 | 4/6/2021 | North America | Mexico | Hidalgo |
| Mexico/HID-InDRE_FB13988_S1985/2021 | EPI_ISL_2158227 | 4/20/2021 | North America | Mexico | Hidalgo |
| Mexico/HID-InDRE_FB13990_S2230/2021 | EPI_ISL_2340957 | 4/23/2021 | North America | Mexico | Hidalgo |
| Mexico/HID-InDRE_FB17207_S3093/2021 | EPI_ISL_2663360 | 5/21/2021 | North America | Mexico | Hidalgo |
| Mexico/HID-InDRE_FB17215_S3175/2021 | EPI_ISL_2674662 | 5/26/2021 | North America | Mexico | Hidalgo |
| Mexico/HID-InDRE_FB17234_S3102/2021 | EPI_ISL_2663369 | 6/5/2021 | North America | Mexico | Hidalgo |
| Mexico/HID-InDRE_FB20305_S5083/2021 | EPI_ISL_3265579 | 7/5/2021 | North America | Mexico | Hidalgo |

| | | | | | |
|---|---|---|---|---|---|
| Mexico/HID-InDRE-54/2020 | EPI_ISL_576257 | 7/22/2020 | North America | Mexico | Hidalgo |
| Mexico/HID-INMEGEN-16-270/2021 | EPI_ISL_2978583 | 7/5/2021 | North America | Mexico | Hidalgo |
| Mexico/JAL_IBT_IMSS_2288/2021 | EPI_ISL_3347893 | 7/24/2021 | North America | Mexico | Jalisco |
| Mexico/JAL_LANGEBIO_IMSS_0720/2021 | EPI_ISL_2402133 | 4/16/2021 | North America | Mexico | Jalisco |
| Mexico/JAL_LANGEBIO_IMSS_0735/2021 | EPI_ISL_2402146 | 4/22/2021 | North America | Mexico | Jalisco |
| Mexico/JAL-IBT_IMSS_1852/2021 | EPI_ISL_2801725 | 6/3/2021 | North America | Mexico | Jalisco |
| Mexico/JAL-LANGEBIO_IMSS_1022/2021 | EPI_ISL_2671575 | 5/24/2021 | North America | Mexico | Jalisco |
| Mexico/JAL-LANGEBIO_IMSS_1030/2021 | EPI_ISL_2671580 | 5/26/2021 | North America | Mexico | Jalisco |
| Mexico/JAL-LANGEBIO_IMSS_1256/2021 | EPI_ISL_2942415 | 6/14/2021 | North America | Mexico | Jalisco |
| Mexico/JAL-LANGEBIO_IMSS_1626/2021 | EPI_ISL_2942730 | 7/1/2021 | North America | Mexico | Jalisco |
| Mexico/MEX_IBT_IMSS_1329/2021 | EPI_ISL_2391571 | 4/22/2021 | North America | Mexico | Estado de Mexico |
| Mexico/MEX_INER_IMSS_00990/2021 | EPI_ISL_2091429 | 4/10/2021 | North America | Mexico | Estado de Mexico |
| Mexico/MEX_LANGEBIO_IMSS_0496/2021 | EPI_ISL_1662121 | 4/2/2021 | North America | Mexico | Estado de Mexico |
| Mexico/MEX-IBT-IMSS-48313/NC/2021 | EPI_ISL_3463388 | 1/30/2021 | North America | Mexico | Estado de Mexico |
| Mexico/MEX-IBT-IMSS-551/2021 | EPI_ISL_1302252 | 1/31/2021 | North America | Mexico | Estado de Mexico |
| Mexico/MEX-InDRE_324/2020 | EPI_ISL_1060752 | 7/28/2020 | North America | Mexico | Estado de Mexico |
| Mexico/MEX-InDRE_467/2021 | EPI_ISL_1168537 | 1/16/2021 | North America | Mexico | Estado de Mexico |
| Mexico/MEX-InDRE_FB20474_S5141/2021 | EPI_ISL_3265630 | 7/7/2021 | North America | Mexico | Estado de Mexico |
| Mexico/MEX-InDRE-IBT-120/2020 | EPI_ISL_1301576 | 4/28/2020 | North America | Mexico | Estado de Mexico |
| Mexico/MEX-InDRE-IBT-122/2020 | EPI_ISL_1301578 | 4/28/2020 | North America | Mexico | Estado de Mexico |

| | | | | | |
|---|---|---|---|---|---|
| Mexico/MEX-INER-IMSS-00310/2021 | EPI_ISL_1279563 | 2/22/2021 | North America | Mexico | Estado de Mexico |
| Mexico/MEX-INMEGEN-05-01-295/2021 | EPI_ISL_2105805 | 4/29/2021 | North America | Mexico | Estado de Mexico |
| Mexico/MEX-INMEGEN-05-05-148/2021 | EPI_ISL_2444489 | 5/21/2021 | North America | Mexico | Estado de Mexico |
| Mexico/MEX-INMEGEN-16-17/2021 | EPI_ISL_3062841 | 6/26/2021 | North America | Mexico | Estado de Mexico |
| Mexico/MEX-INMEGEN-16-5/2021 | EPI_ISL_3062837 | 6/23/2021 | North America | Mexico | Estado de Mexico |
| Mexico/MEX-INMEGEN-17-75/2021 | EPI_ISL_3067758 | 7/3/2021 | North America | Mexico | Estado de Mexico |
| Mexico/MEX-LANGEBIO_IMSS_1062/2021 | EPI_ISL_2671589 | 5/17/2021 | North America | Mexico | Estado de Mexico |
| Mexico/MIC_IBT_IMSS_2221/2021 | EPI_ISL_3347840 | 7/16/2021 | North America | Mexico | Michoacan |
| Mexico/MIC_LANGEBIO_IMSS_31907-NC/2021 | EPI_ISL_2969984 | 3/23/2021 | North America | Mexico | Michoacan |
| Mexico/MIC-IBT-IMSS-459/2020 | EPI_ISL_1301642 | 4/20/2020 | North America | Mexico | Michoacan |
| Mexico/MIC-InDRE_191/2020 | EPI_ISL_933668 | 11/21/2020 | North America | Mexico | Michoacan |
| Mexico/MIC-InDRE_F10346_S853/2021 | EPI_ISL_1399259 | 2/17/2021 | North America | Mexico | Michoacan |
| Mexico/MIC-InDRE_FB15012_S2160/2021 | EPI_ISL_2340897 | 4/21/2021 | North America | Mexico | Michoacan |
| Mexico/MIC-InDRE_FB15017_S2161/2021 | EPI_ISL_2340898 | 4/22/2021 | North America | Mexico | Michoacan |
| Mexico/MIC-InDRE_FB15029_S2166/2021 | EPI_ISL_2340870 | 5/1/2021 | North America | Mexico | Michoacan |
| Mexico/MIC-InDRE_FB17632_S3453/2021 | EPI_ISL_2736849 | 6/10/2021 | North America | Mexico | Michoacan |
| Mexico/MIC-InDRE_FB18219_S4182/2021 | EPI_ISL_2920753 | 6/14/2021 | North America | Mexico | Michoacan |
| Mexico/MIC-InDRE_FB19328_S4590/2021 | EPI_ISL_3033443 | 7/1/2021 | North America | Mexico | Michoacan |
| Mexico/MIC-LANGEBIO_IMSS_1028/2021 | EPI_ISL_2671610 | 5/21/2021 | North America | Mexico | Michoacan |
| Mexico/MOR_IBT_IMSS_1569/2021 | EPI_ISL_2681334 | 5/24/2021 | North America | Mexico | Morelos |

| | | | | | |
|---|---|---|---|---|---|
| Mexico/MOR_IBT_SSMor_27/2021 | EPI_ISL_3342677 | 7/24/2021 | North America | Mexico | Morelos |
| Mexico/MOR_LANGEBIO_IMSS_06222-NC/2021 | EPI_ISL_2969985 | 4/29/2021 | North America | Mexico | Morelos |
| Mexico/MOR-IBT_IMSS_15905-NC/2021 | EPI_ISL_2801791 | 6/1/2021 | North America | Mexico | Morelos |
| Mexico/MOR-IBT_IMSS_1927/2021 | EPI_ISL_2801793 | 6/1/2021 | North America | Mexico | Morelos |
| Mexico/MOR-IBT-IMSS-173/2021 | EPI_ISL_1288372 | 2/12/2021 | North America | Mexico | Morelos |
| Mexico/MOR-IBT-IMSS-467/2020 | EPI_ISL_1301650 | 4/27/2020 | North America | Mexico | Morelos |
| Mexico/MOR-InDRE_F12307_S1305/2021 | EPI_ISL_1651906 | 3/23/2021 | North America | Mexico | Morelos |
| Mexico/MOR-InDRE_FB13805_S1938/2021 | EPI_ISL_2158180 | 4/21/2021 | North America | Mexico | Morelos |
| Mexico/MOR-InDRE_FB21916_S5233/2021 | EPI_ISL_3459943 | 7/11/2021 | North America | Mexico | Morelos |
| Mexico/MOR-LANGEBIO_IMSS_1149/2021 | EPI_ISL_2671615 | 5/24/2021 | North America | Mexico | Morelos |
| Mexico/NAY_IBT_IMSS_2206/2021 | EPI_ISL_3347828 | 7/13/2021 | North America | Mexico | Nayarit |
| Mexico/NAY_IBT_IMSS_2280/2021 | EPI_ISL_3347886 | 7/21/2021 | North America | Mexico | Nayarit |
| Mexico/NAY_INER_IMSS_1166/2021 | EPI_ISL_2490395 | 5/3/2021 | North America | Mexico | Nayarit |
| Mexico/NAY_LANGEBIO_IMSS_0740/2021 | EPI_ISL_2402151 | 4/22/2021 | North America | Mexico | Nayarit |
| Mexico/NAY-IBT_IMSS_1851/2021 | EPI_ISL_2801724 | 6/2/2021 | North America | Mexico | Nayarit |
| Mexico/NAY-InDRE_FB13205_S1589/2021 | EPI_ISL_1821161 | 4/10/2021 | North America | Mexico | Nayarit |
| Mexico/NAY-InDRE_FB14020_S1962/2021 | EPI_ISL_2158204 | 4/20/2021 | North America | Mexico | Nayarit |
| Mexico/NAY-LANGEBIO_IMSS_1027/2021 | EPI_ISL_2671617 | 5/24/2021 | North America | Mexico | Nayarit |
| Mexico/NAY-LANGEBIO_IMSS_1286/2021 | EPI_ISL_2942440 | 6/21/2021 | North America | Mexico | Nayarit |
| Mexico/NLE_IBT_IMSS_1402/2021 | EPI_ISL_2681260 | 6/1/2021 | North America | Mexico | Nuevo Leon |

| | | | | | |
|---|---|---|---|---|---|
| Mexico/NLE_IBT_IMSS_2361/2021 | EPI_ISL_3347666 | 7/18/2021 | North America | Mexico | Nuevo Leon |
| Mexico/NLE_INER_IMSS_1023/2021 | EPI_ISL_2490441 | 5/3/2021 | North America | Mexico | Nuevo Leon |
| Mexico/NLE_LANGEBIO_IMSS_0554/2021 | EPI_ISL_2401977 | 4/27/2021 | North America | Mexico | Nuevo Leon |
| Mexico/NLE-InDRE_FB17360_S3921/2021 | EPI_ISL_2858935 | 6/7/2021 | North America | Mexico | Nuevo Leon |
| Mexico/NLE-INER_IMSS_1428/2021 | EPI_ISL_3155438 | 7/9/2021 | North America | Mexico | Nuevo Leon |
| Mexico/NLE-LANGEBIO_IMSS_0820/2021 | EPI_ISL_2671621 | 5/17/2021 | North America | Mexico | Nuevo Leon |
| Mexico/NLE-LESPNL-20210512-420/2021 | EPI_ISL_2102615 | 4/29/2021 | North America | Mexico | Nuevo Leon |
| Mexico/NLE-UANL-041/2020 | EPI_ISL_979333 | 12/26/2020 | North America | Mexico | Nuevo Leon |
| Mexico/OAX_IBT_IMSS_2506/2021 | EPI_ISL_3347563 | 7/21/2021 | North America | Mexico | Oaxaca |
| Mexico/OAX_LANGEBIO_IMSS_0461/2021 | EPI_ISL_1662147 | 3/22/2021 | North America | Mexico | Oaxaca |
| Mexico/OAX_LANGEBIO_IMSS_0787/2021 | EPI_ISL_2402198 | 4/21/2021 | North America | Mexico | Oaxaca |
| Mexico/OAX_LANGEBIO_IMSS_0788/2021 | EPI_ISL_2402199 | 4/22/2021 | North America | Mexico | Oaxaca |
| Mexico/OAX-InDRE_FB21361_S5253/2021 | EPI_ISL_3460066 | 7/4/2021 | North America | Mexico | Oaxaca |
| Mexico/OAX-InDRE-IBT-19/2020 | EPI_ISL_1301698 | 6/4/2020 | North America | Mexico | Oaxaca |
| Mexico/OAX-InDRE-IBT-27468/NC/2020 | EPI_ISL_3463472 | 6/3/2020 | North America | Mexico | Oaxaca |
| Mexico/OAX-LANGEBIO_IMSS_1080/2021 | EPI_ISL_2671638 | 5/16/2021 | North America | Mexico | Oaxaca |
| Mexico/OAX-LANGEBIO_IMSS_1127/2021 | EPI_ISL_2671640 | 5/18/2021 | North America | Mexico | Oaxaca |
| Mexico/OAX-LANGEBIO_IMSS_1396/2021 | EPI_ISL_2942522 | 6/17/2021 | North America | Mexico | Oaxaca |
| Mexico/OAX-LANGEBIO_IMSS_1825/2021 | EPI_ISL_2942898 | 6/27/2021 | North America | Mexico | Oaxaca |
| Mexico/PUE_IBT_IMSS_1169/2021 | EPI_ISL_2391670 | 5/1/2021 | North America | Mexico | Puebla |

| | | | | | |
|---|---|---|---|---|---|
| Mexico/PUE_IBT_IMSS_1251/2021 | EPI_ISL_2391678 | 5/6/2021 | North America | Mexico | Puebla |
| Mexico/PUE_IBT_IMSS_1310/2021 | EPI_ISL_2391532 | 4/28/2021 | North America | Mexico | Puebla |
| Mexico/PUE_LANGEBIO_IMSS_02371-NC/2021 | EPI_ISL_2970012 | 4/19/2021 | North America | Mexico | Puebla |
| Mexico/PUE-IBT-IMSS-450/2020 | EPI_ISL_1301633 | 4/18/2020 | North America | Mexico | Puebla |
| Mexico/PUE-InDRE_190/2020 | EPI_ISL_933667 | 10/21/2020 | North America | Mexico | Puebla |
| Mexico/PUE-InDRE_381/2020 | EPI_ISL_1054968 | 7/30/2020 | North America | Mexico | Puebla |
| Mexico/PUE-InDRE_FB17746_S3755/2021 | EPI_ISL_2779257 | 6/11/2021 | North America | Mexico | Puebla |
| Mexico/PUE-InDRE_FB19630_S4515/2021 | EPI_ISL_3033368 | 6/22/2021 | North America | Mexico | Puebla |
| Mexico/PUE-InDRE-05/2020 | EPI_ISL_424672 | 3/11/2020 | North America | Mexico | Puebla |
| Mexico/PUE-InDRE-17/2020 | EPI_ISL_455455 | 3/14/2020 | North America | Mexico | Puebla |
| Mexico/PUE-LABOPAT-69_26794/2021 | EPI_ISL_3055559 | 7/6/2021 | North America | Mexico | Puebla |
| Mexico/PUE-LABOPAT-91_27256/2021 | EPI_ISL_3055552 | 7/9/2021 | North America | Mexico | Puebla |
| Mexico/QRO-InDRE-88/2020 | EPI_ISL_658888 | 11/3/2020 | North America | Mexico | Queretaro |
| Mexico/QRO-InDRE-90/2020 | EPI_ISL_658893 | 11/3/2020 | North America | Mexico | Queretaro |
| Mexico/QUE_IBT_IMSS_1168/2021 | EPI_ISL_2391715 | 4/29/2021 | North America | Mexico | Queretaro |
| Mexico/QUE-InDRE_378/2021 | EPI_ISL_1168456 | 1/24/2021 | North America | Mexico | Queretaro |
| Mexico/QUE-InDRE_380/2021 | EPI_ISL_1168458 | 1/25/2021 | North America | Mexico | Queretaro |
| Mexico/QUE-InDRE_401/2020 | EPI_ISL_1054971 | 7/30/2020 | North America | Mexico | Queretaro |
| Mexico/QUE-InDRE_402/2020 | EPI_ISL_1054972 | 7/30/2020 | North America | Mexico | Queretaro |
| Mexico/QUE-InDRE_F9841_S619/2021 | EPI_ISL_1340657 | 2/15/2021 | North America | Mexico | Queretaro |

| | | | | | |
|---|---|---|---|---|---|
| Mexico/QUE-InDRE_FB13501_S1907/2021 | EPI_ISL_2158150 | 4/19/2021 | North America | Mexico | Queretaro |
| Mexico/QUE-InDRE_FB15742_S2982/2021 | EPI_ISL_2559323 | 5/3/2021 | North America | Mexico | Queretaro |
| Mexico/QUE-InDRE_FB15778_S2635/2021 | EPI_ISL_2495946 | 5/5/2021 | North America | Mexico | Queretaro |
| Mexico/QUE-InDRE_FB18359_S4142/2021 | EPI_ISL_2920713 | 6/25/2021 | North America | Mexico | Queretaro |
| Mexico/QUE-InDRE_FB19414_S4615/2021 | EPI_ISL_3046094 | 6/30/2021 | North America | Mexico | Queretaro |
| Mexico/QUE-InDRE_FB23918_S5471/2021 | EPI_ISL_3460189 | 7/23/2021 | North America | Mexico | Queretaro |
| Mexico/QUE-InDRE_FB23938_S5460/2021 | EPI_ISL_3459936 | 7/23/2021 | North America | Mexico | Queretaro |
| Mexico/QUE-InDRE-IBT-21067/NC/2020 | EPI_ISL_3463413 | 5/26/2020 | North America | Mexico | Queretaro |
| Mexico/QUE-InDRE-IBT-53/2020 | EPI_ISL_1301541 | 6/2/2020 | North America | Mexico | Queretaro |
| Mexico/ROO_LANGEBIO_IMSS_07604-NC/2021 | EPI_ISL_2970024 | 4/17/2021 | North America | Mexico | Quintana Roo |
| Mexico/ROO-IBT-IMSS-427/2020 | EPI_ISL_1301498 | 3/25/2020 | North America | Mexico | Quintana Roo |
| Mexico/ROO-InDRE_FB13728_S1840/2021 | EPI_ISL_2157320 | 4/17/2021 | North America | Mexico | Quintana Roo |
| Mexico/ROO-InDRE_FB16788_S2908/2021 | EPI_ISL_2545808 | 5/25/2021 | North America | Mexico | Quintana Roo |
| Mexico/ROO-INMEGEN-05-06-213/2021 | EPI_ISL_2603701 | 5/31/2021 | North America | Mexico | Quintana Roo |
| Mexico/ROO-INMEGEN-06-02-69/2021 | EPI_ISL_2692601 | 6/7/2021 | North America | Mexico | Quintana Roo |
| Mexico/ROO-INMEGEN-18-79/2021 | EPI_ISL_3160709 | 7/13/2021 | North America | Mexico | Quintana Roo |
| Mexico/ROO-INMEGEN-19-170/2021 | EPI_ISL_3277691 | 7/20/2021 | North America | Mexico | Quintana Roo |
| Mexico/ROO-LANGEBIO_IMSS_1185/2021 | EPI_ISL_2942346 | 6/17/2021 | North America | Mexico | Quintana Roo |
| Mexico/SEARCH-100421/2020 | EPI_ISL_2231405 | 12/3/2020 | North America | Mexico | Baja California |
| Mexico/SEARCH-101440/2021 | EPI_ISL_2422832 | 5/11/2021 | North America | Mexico | Baja California |

| | | | | | |
|---|---|---|---|---|---|
| Mexico/SEARCH-102112/2021 | EPI_ISL_2628540 | 5/29/2021 | North America | Mexico | Baja California |
| Mexico/SIN_CIAD_S6757/2021 | EPI_ISL_2249253 | 5/9/2021 | North America | Mexico | Sinaloa |
| Mexico/SIN_LANGEBIO_IMSS_0723/2021 | EPI_ISL_2402136 | 4/16/2021 | North America | Mexico | Sinaloa |
| Mexico/SIN-IBT_IMSS_1832/2021 | EPI_ISL_2801707 | 5/29/2021 | North America | Mexico | Sinaloa |
| Mexico/SIN-InDRE_FB13789_S1931/2021 | EPI_ISL_2158173 | 4/16/2021 | North America | Mexico | Sinaloa |
| Mexico/SIN-InDRE_FB14665_S2140/2021 | EPI_ISL_2340884 | 4/27/2021 | North America | Mexico | Sinaloa |
| Mexico/SIN-InDRE_FB22149_S5272/2021 | EPI_ISL_3460078 | 7/14/2021 | North America | Mexico | Sinaloa |
| Mexico/SIN-INER_IMSS_1451/2021 | EPI_ISL_3155461 | 7/3/2021 | North America | Mexico | Sinaloa |
| Mexico/SIN-INMEGEN-18-203/2021 | EPI_ISL_3160827 | 6/29/2021 | North America | Mexico | Sinaloa |
| Mexico/SIN-INMEGEN-18-210/2021 | EPI_ISL_3160833 | 6/29/2021 | North America | Mexico | Sinaloa |
| Mexico/SLP_IBT_IMSS_1474/2021 | EPI_ISL_2681212 | 5/27/2021 | North America | Mexico | San Luis Potosi |
| Mexico/SLP_INER_IMSS_00934/2021 | EPI_ISL_2091379 | 4/12/2021 | North America | Mexico | San Luis Potosi |
| Mexico/SLP_LANGEBIO_IMSS_0671/2021 | EPI_ISL_2402086 | 4/21/2021 | North America | Mexico | San Luis Potosi |
| Mexico/SLP_LANGEBIO_IMSS_0695/2021 | EPI_ISL_2402109 | 4/23/2021 | North America | Mexico | San Luis Potosi |
| Mexico/SLP_UASLP_A007/2021 | EPI_ISL_1469117 | 1/20/2021 | North America | Mexico | San Luis Potosi |
| Mexico/SLP_UASLP_A020/2021 | EPI_ISL_1469111 | 1/21/2021 | North America | Mexico | San Luis Potosi |
| Mexico/SLP-IBT_IMSS_1823/2021 | EPI_ISL_2801699 | 6/6/2021 | North America | Mexico | San Luis Potosi |
| Mexico/SLP-IBT-IMSS-289/2021 | EPI_ISL_1288475 | 2/12/2021 | North America | Mexico | San Luis Potosi |
| Mexico/SLP-IBT-IMSS-562/2021 | EPI_ISL_1302352 | 1/31/2021 | North America | Mexico | San Luis Potosi |
| Mexico/SLP-InDRE_FB16835B_S2931/2021 | EPI_ISL_2545819 | 5/12/2021 | North America | Mexico | San Luis Potosi |

| | | | | | |
|---|---|---|---|---|---|
| Mexico/SLP-InDRE_FB20630_S4974/2021 | EPI_ISL_3265470 | 7/1/2021 | North America | Mexico | San Luis Potosi |
| Mexico/SLP-INER_IMSS_1437/2021 | EPI_ISL_3155447 | 7/7/2021 | North America | Mexico | San Luis Potosi |
| Mexico/SLP-INER-IMSS-00168/2021 | EPI_ISL_1279444 | 2/21/2021 | North America | Mexico | San Luis Potosi |
| Mexico/SLP-LANGEBIO_IMSS_1333/2021 | EPI_ISL_2942476 | 6/24/2021 | North America | Mexico | San Luis Potosi |
| Mexico/SLP-UASLP-AH1COV2SS027_S16/2020 | EPI_ISL_1494725 | 5/22/2020 | North America | Mexico | San Luis Potosi |
| Mexico/SO-SEARCH-102895/2020 | EPI_ISL_2835375 | 5/12/2020 | North America | Mexico | Sonora |
| Mexico/SON-InDRE_336/2020 | EPI_ISL_1060680 | 12/8/2020 | North America | Mexico | Sonora |
| Mexico/SON-InDRE_FB14082_S2255/2021 | EPI_ISL_2340978 | 4/19/2021 | North America | Mexico | Sonora |
| Mexico/SON-InDRE_FB14085_S1758/2021 | EPI_ISL_2101897 | 4/25/2021 | North America | Mexico | Sonora |
| Mexico/SON-InDRE_FB16353_S3442/2021 | EPI_ISL_2800931 | 5/24/2021 | North America | Mexico | Sonora |
| Mexico/SON-InDRE_FB16807_S2912/2021 | EPI_ISL_2545812 | 5/21/2021 | North America | Mexico | Sonora |
| Mexico/SON-InDRE_FB17812_S4305/2021 | EPI_ISL_2937767 | 6/10/2021 | North America | Mexico | Sonora |
| Mexico/SON-InDRE_FB21626_S5054/2021 | EPI_ISL_3265550 | 7/7/2021 | North America | Mexico | Sonora |
| Mexico/SON-InDRE-IBT-29413/NC/2020 | EPI_ISL_3463394 | 6/9/2020 | North America | Mexico | Sonora |
| Mexico/SON-INER_IMSS_1490/2021 | EPI_ISL_3155500 | 7/8/2021 | North America | Mexico | Sonora |
| Mexico/SON-LANGEBIO_IMSS_1313/2021 | EPI_ISL_2942460 | 6/23/2021 | North America | Mexico | Sonora |
| Mexico/SON-SEARCH-102633/2020 | EPI_ISL_2712699 | 5/5/2020 | North America | Mexico | Sonora |
| Mexico/SON-SEARCH-102658/2020 | EPI_ISL_2712724 | 5/9/2020 | North America | Mexico | Sonora |
| Mexico/TAB_IBT_IMSS_2172/2021 | EPI_ISL_3347918 | 7/17/2021 | North America | Mexico | Tabasco |
| Mexico/TAB_LANGEBIO_IMSS_0627/2021 | EPI_ISL_2402044 | 4/21/2021 | North America | Mexico | Tabasco |

| | | | | | |
|---|---|---|---|---|---|
| Mexico/TAB_LANGEBIO_IMSS_0639/2021 | EPI_ISL_2402056 | 4/24/2021 | North America | Mexico | Tabasco |
| Mexico/TAB-IBT_IMSS_1887/2021 | EPI_ISL_2801585 | 6/3/2021 | North America | Mexico | Tabasco |
| Mexico/TAB-InDRE_FB17650_S3732/2021 | EPI_ISL_2779235 | 6/11/2021 | North America | Mexico | Tabasco |
| Mexico/TAB-InDRE_FB20838_S5335/2021 | EPI_ISL_3459948 | 7/10/2021 | North America | Mexico | Tabasco |
| Mexico/TAB-InDRE-IBT-31/2020 | EPI_ISL_1301537 | 5/29/2020 | North America | Mexico | Tabasco |
| Mexico/TAB-LANGEBIO_IMSS_0947/2021 | EPI_ISL_2671712 | 5/22/2021 | North America | Mexico | Tabasco |
| Mexico/TAB-LANGEBIO_IMSS_0948/2021 | EPI_ISL_2671713 | 5/22/2021 | North America | Mexico | Tabasco |
| Mexico/TAM_LANGEBIO_IMSS_0034/2021 | EPI_ISL_1351498 | 2/22/2021 | North America | Mexico | Tabasco |
| Mexico/TAM_LANGEBIO_IMSS_0053/2021 | EPI_ISL_1351552 | 2/27/2021 | North America | Mexico | Sonora |
| Mexico/TAM_LANGEBIO_IMSS_0098/2021 | EPI_ISL_1351688 | 3/7/2021 | North America | Mexico | Nayarit |
| Mexico/TAM_LANGEBIO_IMSS_0570/2021 | EPI_ISL_2401991 | 4/28/2021 | North America | Mexico | Tamaulipas |
| Mexico/TAM-InDRE_FB14047_S2279/2021 | EPI_ISL_2340997 | 4/13/2021 | North America | Mexico | Tamaulipas |
| Mexico/TAM-InDRE_FB14065_S1964/2021 | EPI_ISL_2158206 | 4/24/2021 | North America | Mexico | Tamaulipas |
| Mexico/TAM-InDRE_FB16027_S2426/2021 | EPI_ISL_2455950 | 5/13/2021 | North America | Mexico | Tamaulipas |
| Mexico/TAM-InDRE_FB16729_S2941/2021 | EPI_ISL_2545689 | 5/21/2021 | North America | Mexico | Tamaulipas |
| Mexico/TAM-InDRE_FB18874_S4154/2021 | EPI_ISL_2920725 | 6/16/2021 | North America | Mexico | Tamaulipas |
| Mexico/TAM-InDRE_FB18919_S4639/2021 | EPI_ISL_3033469 | 6/21/2021 | North America | Mexico | Tamaulipas |
| Mexico/TAM-InDRE_FB21167_S5080/2021 | EPI_ISL_3265576 | 7/1/2021 | North America | Mexico | Tamaulipas |
| Mexico/TAM-InDRE_FB22607_S5496/2021 | EPI_ISL_3460002 | 7/12/2021 | North America | Mexico | Tamaulipas |
| Mexico/TAM-InDRE-94/2020 | EPI_ISL_794592 | 12/31/2020 | North America | Mexico | Tamaulipas |

| | | | | | |
|---|---|---|---|---|---|
| Mexico/TAM-UANL-013/2020 | EPI_ISL_1091272 | 6/27/2020 | North America | Mexico | Tamaulipas |
| Mexico/TLA_IBT_IMSS_1022/2021 | EPI_ISL_1811528 | 4/8/2021 | North America | Mexico | Tlaxcala |
| Mexico/TLA_IBT_IMSS_1163/2021 | EPI_ISL_2391606 | 4/30/2021 | North America | Mexico | Tlaxcala |
| Mexico/TLA_IBT_IMSS_17405/NC/2021 | EPI_ISL_3133787 | 6/7/2021 | North America | Mexico | Tlaxcala |
| Mexico/TLA_IBT_IMSS_2393/2021 | EPI_ISL_3347698 | 7/16/2021 | North America | Mexico | Tlaxcala |
| Mexico/TLA_IBT_IMSS_2526/2021 | EPI_ISL_3347794 | 7/21/2021 | North America | Mexico | Tlaxcala |
| Mexico/TLA_INER_IMSS_00603/2021 | EPI_ISL_1585600 | 3/17/2021 | North America | Mexico | Tlaxcala |
| Mexico/TLA_INER_IMSS_1325/2021 | EPI_ISL_2490582 | 5/13/2021 | North America | Mexico | Tlaxcala |
| Mexico/TLA-IBT-IMSS-429/2020 | EPI_ISL_1301622 | 3/27/2020 | North America | Mexico | Tlaxcala |
| Mexico/TLA-InDRE_147/2020 | EPI_ISL_913950 | 12/14/2020 | North America | Mexico | Tlaxcala |
| Mexico/TLA-InDRE_174/2020 | EPI_ISL_913975 | 12/4/2020 | North America | Mexico | Tlaxcala |
| Mexico/TLA-InDRE_281/2020 | EPI_ISL_1054948 | 8/13/2020 | North America | Mexico | Tlaxcala |
| Mexico/TLA-InDRE_FB13373_S1888/2021 | EPI_ISL_2158131 | 4/17/2021 | North America | Mexico | Tlaxcala |
| Mexico/TLA-InDRE_FB18148_S4379/2021 | EPI_ISL_2937845 | 6/20/2021 | North America | Mexico | Tlaxcala |
| Mexico/TLA-InDRE-IBT-16/2020 | EPI_ISL_1301540 | 5/6/2020 | North America | Mexico | Tlaxcala |
| Mexico/TLA-InDRE-IBT-73/2020 | EPI_ISL_1301692 | 6/6/2020 | North America | Mexico | Tlaxcala |
| Mexico/TLA-InDRE-IBT-76/2020 | EPI_ISL_1301693 | 6/8/2020 | North America | Mexico | Tlaxcala |
| Mexico/TLA-INER-IMSS-00283/2021 | EPI_ISL_1287775 | 2/22/2021 | North America | Mexico | Tlaxcala |
| Mexico/TLA-LANGEBIO_IMSS_1148/2021 | EPI_ISL_2671724 | 5/23/2021 | North America | Mexico | Tlaxcala |
| Mexico/VER_IBT_IMSS_1013/2021 | EPI_ISL_1811520 | 4/5/2021 | North America | Mexico | Veracruz |

| | | | | | |
|---|---|---|---|---|---|
| Mexico/VER_IBT_IMSS_1123/2021 | EPI_ISL_1811463 | 4/5/2021 | North America | Mexico | Veracruz |
| Mexico/VER_IBT_IMSS_1331/2021 | EPI_ISL_2391555 | 4/29/2021 | North America | Mexico | Veracruz |
| Mexico/VER-IBT-IMSS-269/2021 | EPI_ISL_1288457 | 2/11/2021 | North America | Mexico | Veracruz |
| Mexico/VER-IBT-IMSS-512/2020 | EPI_ISL_1301677 | 5/17/2020 | North America | Mexico | Veracruz |
| Mexico/VER-InDRE_152/2020 | EPI_ISL_913955 | 12/17/2020 | North America | Mexico | Veracruz |
| Mexico/VER-InDRE_169/2020 | EPI_ISL_913970 | 11/21/2020 | North America | Mexico | Veracruz |
| Mexico/VER-InDRE_FB14022_S2244/2021 | EPI_ISL_2340969 | 4/20/2021 | North America | Mexico | Veracruz |
| Mexico/VER-InDRE_FB20977_S5150/2021 | EPI_ISL_3265639 | 7/7/2021 | North America | Mexico | Veracruz |
| Mexico/VER-INMEGEN-06-04-288/2021 | EPI_ISL_2894335 | 6/12/2021 | North America | Mexico | Veracruz |
| Mexico/VER-INMEGEN-16-354/2021 | EPI_ISL_2978658 | 6/14/2021 | North America | Mexico | Veracruz |
| Mexico/VER-INMEGEN-18-329/2021 | EPI_ISL_3160939 | 7/15/2021 | North America | Mexico | Veracruz |
| Mexico/VER-LANGEBIO_IMSS_1107/2021 | EPI_ISL_2671741 | 5/18/2021 | North America | Mexico | Veracruz |
| Mexico/VER-LANGEBIO_IMSS_1133/2021 | EPI_ISL_2671743 | 5/20/2021 | North America | Mexico | Veracruz |
| Mexico/YUC_LANGEBIO_IMSS_0628/2021 | EPI_ISL_2402045 | 4/23/2021 | North America | Mexico | Yucatan |
| Mexico/YUC-InDRE_FB15156_S2200/2021 | EPI_ISL_2340935 | 3/23/2021 | North America | Mexico | Yucatan |
| Mexico/YUC-InDRE_FB16573_S2744/2021 | EPI_ISL_2533770 | 5/17/2021 | North America | Mexico | Yucatan |
| Mexico/YUC-InDRE_FB16644_S2874/2021 | EPI_ISL_2545776 | 5/21/2021 | North America | Mexico | Yucatan |
| Mexico/YUC-InDRE-IBT-135/2020 | EPI_ISL_1301591 | 4/28/2020 | North America | Mexico | Yucatan |
| Mexico/YUC-INMEGEN-06-01-306/2021 | EPI_ISL_2617025 | 6/3/2021 | North America | Mexico | Yucatan |
| Mexico/YUC-INMEGEN-06-04-76/2021 | EPI_ISL_2894220 | 6/9/2021 | North America | Mexico | Yucatan |

| | | | | | |
|---|---|---|---|---|---|
| Mexico/YUC-INMEGEN-18-15/2021 | EPI_ISL_3160648 | 7/13/2021 | North America | Mexico | Yucatan |
| Mexico/YUC-INMEGEN-19-164/2021 | EPI_ISL_3277687 | 7/20/2021 | North America | Mexico | Yucatan |
| Mexico/YUC-NYGC-1156-SM/2021 | EPI_ISL_3023810 | 4/13/2021 | North America | Mexico | Yucatan |
| Mexico/YUC-NYGC-1186-SM/2021 | EPI_ISL_3023945 | 4/21/2021 | North America | Mexico | Yucatan |
| Mexico/YUC-NYGC-4170-21/2021 | EPI_ISL_3023824 | 1/15/2021 | North America | Mexico | Yucatan |
| Mexico/YUC-NYGC-58303-20/2020 | EPI_ISL_3024025 | 11/10/2020 | North America | Mexico | Yucatan |
| Mexico/YUC-NYGC-63648-20/2020 | EPI_ISL_3023959 | 11/30/2020 | North America | Mexico | Yucatan |
| Mexico/YUC-NYGC-65280-20/2020 | EPI_ISL_3023846 | 12/7/2020 | North America | Mexico | Yucatan |
| Mexico/YUC-NYGC-8334-20/2020 | EPI_ISL_3023837 | 5/13/2020 | North America | Mexico | Yucatan |
| Mexico/ZAC_IBT_IMSS_1458/2021 | EPI_ISL_2681207 | 5/24/2021 | North America | Mexico | Zacatecas |
| Mexico/ZAC_IBT_IMSS_2112/2021 | EPI_ISL_3347896 | 7/14/2021 | North America | Mexico | Zacatecas |
| Mexico/ZAC_INER_IMSS_1244/2021 | EPI_ISL_2835945 | 5/15/2021 | North America | Mexico | Zacatecas |
| Mexico/ZAC_LANGEBIO_IMSS_06651-NC/2021 | EPI_ISL_2970094 | 4/19/2021 | North America | Mexico | Zacatecas |
| Mexico/ZAC_LANGEBIO_IMSS_0715/2021 | EPI_ISL_2402128 | 4/30/2021 | North America | Mexico | Zacatecas |
| Mexico/ZAC-IBT_IMSS_1827/2021 | EPI_ISL_2801702 | 6/8/2021 | North America | Mexico | Zacatecas |
| Mexico/ZAC-InDRE-IBT-49/2020 | EPI_ISL_1301519 | 4/30/2020 | North America | Mexico | Zacatecas |
| Mexico/ZAC-InDRE-IBT-57/2020 | EPI_ISL_1301690 | 5/2/2020 | North America | Mexico | Zacatecas |
| Mexico/ZAC-InDRE-IBT-59/2020 | EPI_ISL_1301483 | 6/1/2020 | North America | Mexico | Zacatecas |
| Mexico/ZAC-INER_IMSS_1433/2021 | EPI_ISL_3155443 | 7/6/2021 | North America | Mexico | Zacatecas |
| Mexico/ZAC-LANGEBIO_IMSS_1321/2021 | EPI_ISL_2942468 | 6/14/2021 | North America | Mexico | Zacatecas |

| | | | | | |
|---|---|---|---|---|---|
| mink/Netherlands/NB-EMC-32-6/2020 | EPI_ISL_577762 | 8/15/2020 | Europe | Netherlands | Limburg |
| Moldova/ICGEB_MD6/2020 | EPI_ISL_516938 | 6/17/2020 | Europe | Moldova | Rezina |
| Moldova/ICGEB_MD7/2020 | EPI_ISL_516936 | 6/17/2020 | Europe | Moldova | Straseni |
| Moldova/un-ChVir25728/2021 | EPI_ISL_2894995 | 5/10/2021 | Europe | Moldova | Moldova |
| Moldova/un-ChVir25732/2021 | EPI_ISL_2894996 | 5/17/2021 | Europe | Moldova | Moldova |
| Moldova/un-ChVir25847/2021 | EPI_ISL_3064708 | 7/6/2021 | Europe | Moldova | Moldova |
| Moldova/un-ChVir25851/2021 | EPI_ISL_3064712 | 6/28/2021 | Europe | Moldova | Moldova |
| Moldova/un-ChVir25852/2021 | EPI_ISL_3064713 | 6/28/2021 | Europe | Moldova | Moldova |
| Monaco/CERBAHC-HTNG3AFX2-S17/2021 | EPI_ISL_2562031 | 4/17/2021 | Europe | Monaco | Monaco |
| Monaco/CERBAHC-HTNG3AFX2-S63/2021 | EPI_ISL_2562045 | 4/17/2021 | Europe | Monaco | Monaco |
| Monaco/IPP04454/2021 | EPI_ISL_1219949 | 2/22/2021 | Europe | Monaco | Monaco |
| Monaco/IPP14376/2021 | EPI_ISL_2757668 | 6/12/2021 | Europe | Monaco | Monaco |
| Monaco/IPP14377/2021 | EPI_ISL_2757669 | 6/14/2021 | Europe | Monaco | Monaco |
| Mongolia/30/2020 | EPI_ISL_1805697 | 10/9/2020 | Asia | Mongolia | Mongolia |
| Mongolia/3631/2020 | EPI_ISL_1805651 | 3/18/2020 | Asia | Mongolia | Mongolia |
| Mongolia/62497/2020 | EPI_ISL_1805717 | 11/4/2020 | Asia | Mongolia | Mongolia |
| Morocco/CNRST-IND01/2021 | EPI_ISL_2110643 | 4/22/2021 | Africa | Morocco | Casablanca |
| Morocco/CNRST-IND02/2021 | EPI_ISL_2966236 | 4/22/2021 | Africa | Morocco | Casablanca |
| Morocco/FMP-169/2021 | EPI_ISL_1913014 | 2/24/2021 | Africa | Morocco | Casablanca |
| Morocco/FMP-183/2020 | EPI_ISL_2318037 | 8/8/2020 | Africa | Morocco | Sidi Lahcen |
| Morocco/FMP-288/2021 | EPI_ISL_1905079 | 4/9/2021 | Africa | Morocco | Morocco |
| Morocco/FMP-45/2020 | EPI_ISL_775221 | 12/15/2020 | Africa | Morocco | Temara |
| Morocco/FMP-57/2020 | EPI_ISL_775256 | 12/21/2020 | Africa | Morocco | Temara |
| Morocco/HMIMV-121300/2020 | EPI_ISL_2968041 | 7/18/2020 | Africa | Morocco | Rabat |
| Morocco/HMIMV-14N/2020 | EPI_ISL_2968038 | 7/18/2020 | Africa | Morocco | Rabat |
| Morocco/ION_CODE_4/2021 | EPI_ISL_3155339 | 6/17/2021 | Africa | Morocco | Casablanca |
| Morocco/ION-CODE-101/2021 | EPI_ISL_3071138 | 7/8/2021 | Africa | Morocco | Casablanca |

| | | | | | |
|---|---|---|---|---|---|
| Morocco/RA156-1C/2021 | EPI_ISL_3239896 | 6/20/2021 | Africa | Morocco | Morocco |
| Mozambique/CERI-KRISP-K015020/2021 | EPI_ISL_2617109 | 3/11/2021 | Africa | Mozambique | Maputo |
| Mozambique/CERI-KRISP-K015053/2021 | EPI_ISL_2617101 | 3/17/2021 | Africa | Mozambique | Maputo |
| Mozambique/CERI-KRISP-K016042/2021 | EPI_ISL_3447990 | 4/13/2021 | Africa | Mozambique | Maputo |
| Mozambique/CERI-KRISP-K016059/2021 | EPI_ISL_3447979 | 4/19/2021 | Africa | Mozambique | Maputo |
| Mozambique/CERI-KRISP-K016082/2021 | EPI_ISL_3447977 | 4/20/2021 | Africa | Mozambique | Maputo |
| Mozambique/INS-K007913/2020 | EPI_ISL_887428 | 12/26/2020 | Africa | Mozambique | Maputo |
| Mozambique/INS-K011810/2020 | EPI_ISL_2396915 | 10/10/2020 | Africa | Mozambique | Maputo |
| Myanmar/98826-20/2020 | EPI_ISL_3252976 | 8/25/2020 | Asia | Myanmar | Myanmar |
| Myanmar/DSMRC-006/2021 | EPI_ISL_849739 | 1/9/2021 | Asia | Myanmar | Myanmar |
| Myanmar/DSMRC011/2021 | EPI_ISL_2612300 | 5/28/2021 | Asia | Myanmar | Sagaing |
| Myanmar/DSMRC015/2021 | EPI_ISL_2595726 | 6/1/2021 | Asia | Myanmar | Mandalay |
| Myanmar/DSMRC017/2021 | EPI_ISL_2596341 | 6/1/2021 | Asia | Myanmar | Mandalay |
| Myanmar/DSMRC021/2021 | EPI_ISL_2597312 | 5/26/2021 | Asia | Myanmar | Yangon |
| Myanmar/MMC_137/2020 | EPI_ISL_512844 | 4/22/2020 | Asia | Myanmar | Yangon |
| Nepal/2097/2020 | EPI_ISL_754068 | 7/30/2020 | Asia | Nepal | Bagmati |
| Nepal/61/2020 | EPI_ISL_410301 | 1/13/2020 | Asia | Nepal | Bagmati |
| Nepal/S-104/2021 | EPI_ISL_3184525 | 5/28/2021 | Asia | Nepal | Sudurpaschim |
| Nepal/S-115/2021 | EPI_ISL_3184536 | 6/8/2021 | Asia | Nepal | Karnali |
| Nepal/S-126/2021 | EPI_ISL_3184555 | 7/1/2021 | Asia | Nepal | Bagmati |
| Nepal/S-26/2021 | EPI_ISL_2479975 | 4/29/2021 | Asia | Nepal | Bagmati |
| Nepal/S-35/2021 | EPI_ISL_2479966 | 4/27/2021 | Asia | Nepal | Bagmati |
| Nepal/S-54/2021 | EPI_ISL_2674083 | 5/29/2021 | Asia | Nepal | Bagmati |
| Nepal/S-56/2021 | EPI_ISL_2674085 | 6/2/2021 | Asia | Nepal | Province 2 |
| Nepal/S-97/2021 | EPI_ISL_3184592 | 7/7/2021 | Asia | Nepal | Bagmati |
| Netherlands/Diemen_1363454/2020 | EPI_ISL_413570 | 2/28/2020 | Europe | Netherlands | North Holland |
| Netherlands/DR-RIVM-29546/2021 | EPI_ISL_1961180 | 4/26/2021 | Europe | Netherlands | Drenthe |

| | | | | | |
|---|---|---|---|---|---|
| Netherlands/GE-RIVM-25173/2021 | EPI_ISL_1703430 | 4/2/2021 | Europe | Netherlands | Gelderland |
| Netherlands/GR-UMCG-MMB_1930/2021 | EPI_ISL_2285972 | 5/11/2021 | Europe | Netherlands | Groningen |
| Netherlands/LI-MUMC-1594/2021 | EPI_ISL_1509712 | 3/18/2021 | Europe | Netherlands | Limburg |
| Netherlands/LI-MUMC-3225/2021 | EPI_ISL_3474654 | 8/2/2021 | Europe | Netherlands | Limburg |
| Netherlands/LI-RIVM-49521/2021 | EPI_ISL_3390123 | 7/25/2021 | Europe | Netherlands | Limburg |
| Netherlands/NB-EMC-689/2020 | EPI_ISL_1311245 | 10/16/2020 | Europe | Netherlands | North Brabant |
| Netherlands/NB-RIVM-30855/2021 | EPI_ISL_2093278 | 4/20/2021 | Europe | Netherlands | North Brabant |
| Netherlands/NH-RIVM-21096/2020 | EPI_ISL_823991 | 12/31/2020 | Europe | Netherlands | North Holland |
| Netherlands/OV-RIVM-41605/2021 | EPI_ISL_2787578 | 6/17/2021 | Europe | Netherlands | Overijssel |
| Netherlands/UT-UMCU-2407718/2021 | EPI_ISL_2887983 | 6/16/2021 | Europe | Netherlands | Utrecht |
| Netherlands/UT-UMCU-3106539/2021 | EPI_ISL_3428445 | 8/4/2021 | Europe | Netherlands | Utrecht |
| Netherlands/ZH-EMC-2661/2021 | EPI_ISL_2145773 | 5/6/2021 | Europe | Netherlands | South Holland |
| Netherlands/ZH-EMC-687/2020 | EPI_ISL_632321 | 8/20/2020 | Europe | Netherlands | South Holland |
| Netherlands/ZH-RIVM-49575/2021 | EPI_ISL_3390171 | 7/23/2021 | Europe | Netherlands | South Holland |
| NewZealand/20CV0065/2020 | EPI_ISL_548105 | 6/19/2020 | Oceania | New Zealand | Auckland |
| NewZealand/20CV0131/2020 | EPI_ISL_579059 | 8/19/2020 | Oceania | New Zealand | Auckland |
| NewZealand/20CV0209/2020 | EPI_ISL_548114 | 8/29/2020 | Oceania | New Zealand | Auckland |
| NewZealand/20VR3038/2020 | EPI_ISL_456400 | 4/25/2020 | Oceania | New Zealand | Wellington |
| NewZealand/21CH0033/2021 | EPI_ISL_2811951 | 6/2/2021 | Oceania | New Zealand | Canterbury |
| NewZealand/21CH0034/2021 | EPI_ISL_2811952 | 6/7/2021 | Oceania | New Zealand | Canterbury |
| NewZealand/21MV0077/2021 | EPI_ISL_1016878 | 2/11/2021 | Oceania | New Zealand | Wellington |
| NewZealand/21MV0116/2021 | EPI_ISL_1172031 | 2/22/2021 | Oceania | New Zealand | Auckland |
| NewZealand/21MV0352/2021 | EPI_ISL_1904857 | 4/18/2021 | Oceania | New Zealand | Auckland |
| NewZealand/21MV0363/2021 | EPI_ISL_1904853 | 4/20/2021 | Oceania | New Zealand | Auckland |
| NewZealand/21MV0416/2021 | EPI_ISL_2103200 | 5/6/2021 | Oceania | New Zealand | Auckland |
| NewZealand/21MV0485/2021 | EPI_ISL_2650019 | 5/28/2021 | Oceania | New Zealand | Auckland |
| NewZealand/21MV0602/2021 | EPI_ISL_3164090 | 7/15/2021 | Oceania | New Zealand | Auckland |

| | | | | | |
|---|---|---|---|---|---|
| NewZealand/21MV0631/2021 | EPI_ISL_3164081 | 7/22/2021 | Oceania | New Zealand | Auckland |
| NewZealand/21MV0676/2021 | EPI_ISL_3477077 | 8/6/2021 | Oceania | New Zealand | Auckland |
| Niger/CE4783/2021 | EPI_ISL_2835624 | 1/9/2021 | Africa | Niger | Niamey |
| Niger/M37/2020 | EPI_ISL_2835634 | 12/4/2020 | Africa | Niger | Agadez |
| Niger/MI-36/2020 | EPI_ISL_2835640 | 5/8/2020 | Africa | Niger | Maradi |
| Nigeria/BCVL-19053/2021 | EPI_ISL_985072 | 1/11/2021 | Africa | Nigeria | Oyo State |
| Nigeria/CV1322/2021 | EPI_ISL_2240771 | 2/16/2021 | Africa | Nigeria | Rivers |
| Nigeria/Lagos01/2020 | EPI_ISL_413550 | 2/27/2020 | Africa | Nigeria | Lagos |
| Nigeria/NCDC-FCT-10T/2021 | EPI_ISL_2432953 | 5/17/2021 | Africa | Nigeria | Abuja |
| Nigeria/NCDC-FCT-8T/2021 | EPI_ISL_2432922 | 4/19/2021 | Africa | Nigeria | Abuja |
| Nigeria/NCDC-NR01/2021 | EPI_ISL_2987211 | 6/29/2021 | Africa | Nigeria | Cross River State |
| Nigeria/NCDC-NR15/2021 | EPI_ISL_2983224 | 6/20/2021 | Africa | Nigeria | Lagos |
| Nigeria/NCDC-NR158/2021 | EPI_ISL_3256005 | 7/21/2021 | Africa | Nigeria | Akwa-Ibom |
| Nigeria/NCDC-NR192/2021 | EPI_ISL_3430318 | 8/3/2021 | Africa | Nigeria | Kaduna State |
| Nigeria/NCDC-NR98/2021 | EPI_ISL_3314959 | 7/19/2021 | Africa | Nigeria | Akwa-Ibom |
| Nigeria/OS-CV301/2020 | EPI_ISL_729979 | 9/1/2020 | Africa | Nigeria | Osun State |
| Nigeria/OS-CV306/2020 | EPI_ISL_729984 | 10/9/2020 | Africa | Nigeria | Osun State |
| NorthernIreland/PHEC-M304M072/2021 | EPI_ISL_2904345 | 6/28/2021 | Europe | United Kingdom | Northern Ireland |
| NorthernMarianaIslands/CDC-2-3831048/2020 | EPI_ISL_1169504 | 7/2/2020 | North America | USA | Northern Mariana Islands |
| NorthernMarianaIslands/CDC-2-3831114/2020 | EPI_ISL_1169527 | 10/17/2020 | North America | USA | Northern Mariana Islands |
| NorthernMarianaIslands/MP-CDC-2-4392999/2021 | EPI_ISL_2383896 | 4/29/2021 | North America | USA | Northern Mariana Islands |
| NorthernMarianaIslands/MP-CDC-2-4634175/2021 | EPI_ISL_3303926 | 6/28/2021 | North America | USA | Northern Mariana Islands |
| NorthernMarianaIslands/MP-CDC-2-4634281/2021 | EPI_ISL_3127898 | 7/7/2021 | North America | USA | Northern Mariana Islands |
| NorthMacedonia/15411-k/2021 | EPI_ISL_2681439 | 4/22/2021 | Europe | North Macedonia | Skopje |
| NorthMacedonia/15496-k/2021 | EPI_ISL_2987572 | 5/27/2021 | Europe | North Macedonia | Skopje |
| NorthMacedonia/20481-k/2021 | EPI_ISL_3020343 | 7/11/2021 | Europe | North Macedonia | Tetovo |

| | | | | | |
|---|---|---|---|---|---|
| NorthMacedonia/3515/2020 | EPI_ISL_677707 | 6/15/2020 | Europe | North Macedonia | Northeastern Region MK |
| NorthMacedonia/36464/2021 | EPI_ISL_2001038 | 4/9/2021 | Europe | North Macedonia | Southwestern Region |
| NorthMacedonia/38620/2021 | EPI_ISL_2839168 | 4/28/2021 | Europe | North Macedonia | Southwestern Region |
| NorthMacedonia/38916/2021 | EPI_ISL_2987571 | 5/8/2021 | Europe | North Macedonia | Southeastern Region |
| NorthMacedonia/4637/2020 | EPI_ISL_677709 | 6/22/2020 | Europe | North Macedonia | Southwestern Region |
| NorthMacedonia/6752/2020 | EPI_ISL_677715 | 7/10/2020 | Europe | North Macedonia | Pelagonia |
| NorthMacedonia/883/2020 | EPI_ISL_677674 | 5/20/2020 | Europe | North Macedonia | Pelagonia |
| NorthMacedonia/ZMC6203/2020 | EPI_ISL_735392 | 6/5/2020 | Europe | North Macedonia | Skopje |
| NorthMacedonia/ZMC63707/2021 | EPI_ISL_2832105 | 7/1/2021 | Europe | North Macedonia | Tetovo |
| Norway/10263/2021 | EPI_ISL_1828905 | 4/18/2021 | Europe | Norway | Viken |
| Norway/11529/2021 | EPI_ISL_2357823 | 5/3/2021 | Europe | Norway | Innlandet |
| Norway/12383/2021 | EPI_ISL_2483204 | 5/12/2021 | Europe | Norway | Innlandet |
| Norway/14443/2021 | EPI_ISL_2788475 | 6/14/2021 | Europe | Norway | Viken |
| Norway/15257/2021 | EPI_ISL_3133807 | 7/5/2021 | Europe | Norway | Oslo |
| Norway/16039/2021 | EPI_ISL_3266534 | 6/30/2021 | Europe | Norway | Viken |
| Norway/16421/2021 | EPI_ISL_3426913 | 7/30/2021 | Europe | Norway | Viken |
| Norway/1728/2020 | EPI_ISL_2226155 | 2/29/2020 | Europe | Norway | Vestland |
| Norway/2093/2020 | EPI_ISL_420310 | 3/16/2020 | Europe | Norway | Norway |
| Norway/2113/2020 | EPI_ISL_420311 | 3/18/2020 | Europe | Norway | Norway |
| Norway/2244/2020 | EPI_ISL_447838 | 3/23/2020 | Europe | Norway | Norway |
| Norway/2499/2020 | EPI_ISL_481211 | 4/28/2020 | Europe | Norway | Norway |
| Norway/2832/2020 | EPI_ISL_493385 | 6/30/2020 | Europe | Norway | Norway |
| Norway/2858/2020 | EPI_ISL_500781 | 7/2/2020 | Europe | Norway | Norway |
| Norway/5786/2021 | EPI_ISL_1314516 | 3/5/2021 | Europe | Norway | Viken |
| Norway/Ahus-1205/2021 | EPI_ISL_3374556 | 8/4/2021 | Europe | Norway | Viken |
| Norway/Ahus-1257/2021 | EPI_ISL_3394940 | 8/6/2021 | Europe | Norway | Viken |
| Norway/Ahus-353/2021 | EPI_ISL_1731388 | 4/18/2021 | Europe | Norway | Viken |

| | | | | | |
|---|---|---|---|---|---|
| Oman/182621/2020 | EPI_ISL_766569 | 12/22/2020 | Asia | Oman | Muscat |
| Oman/205027055/2020 | EPI_ISL_458123 | 4/6/2020 | Asia | Oman | Muscat |
| Oman/205038157/2020 | EPI_ISL_491993 | 5/22/2020 | Asia | Oman | Muscat |
| Oman/205041374/2020 | EPI_ISL_492016 | 6/4/2020 | Asia | Oman | Muscat |
| Oman/520257977/2020 | EPI_ISL_1532295 | 9/14/2020 | Asia | Oman | Dakhiliyah |
| Oman/520261221/2020 | EPI_ISL_1532299 | 9/26/2020 | Asia | Oman | South Batinah |
| Oman/52110333/2021 | EPI_ISL_2921200 | 5/17/2021 | Asia | Oman | Muscat |
| Oman/52110370/2021 | EPI_ISL_2921201 | 5/13/2021 | Asia | Oman | Muscat |
| Oman/52111724/2021 | EPI_ISL_2921181 | 4/20/2021 | Asia | Oman | Muscat |
| Oman/52111727/2021 | EPI_ISL_2921183 | 4/20/2021 | Asia | Oman | Muscat |
| Oman/5213230/2021 | EPI_ISL_2921198 | 2/8/2021 | Asia | Oman | Muscat |
| Oman/72110566/2021 | EPI_ISL_2921174 | 4/8/2021 | Asia | Oman | Al Wusta |
| Oman/RESP-20-6701/2020 | EPI_ISL_457998 | 3/28/2020 | Asia | Oman | South Batinah |
| Oman/RESP-20-797/2020 | EPI_ISL_457701 | 2/23/2020 | Asia | Oman | Muscat |
| Oman/RESP-20-837/2020 | EPI_ISL_457704 | 2/24/2020 | Asia | Oman | Muscat |
| Pakistan/KPK-KUST-SJTU/2020 | EPI_ISL_513925 | 5/15/2020 | Asia | Pakistan | Pakistan |
| Pakistan/NIH-B10-S1/2021 | EPI_ISL_2894974 | 5/17/2021 | Asia | Pakistan | Pakistan |
| Pakistan/NIH-B12-S13/2021 | EPI_ISL_2757746 | 5/31/2021 | Asia | Pakistan | Pakistan |
| Pakistan/NIH-B12-S27/2021 | EPI_ISL_2757758 | 6/8/2021 | Asia | Pakistan | Pakistan |
| Pakistan/NIH-B12-S6/2021 | EPI_ISL_2757739 | 6/3/2021 | Asia | Pakistan | Rawalpindi |
| Pakistan/NIH-B13-S27/2021 | EPI_ISL_3462523 | 8/5/2021 | Asia | Pakistan | Karachi |
| Pakistan/NIH-B13-S3/2021 | EPI_ISL_3462500 | 8/5/2021 | Asia | Pakistan | Karachi |
| Pakistan/NIH-B9-S11/2021 | EPI_ISL_2894973 | 4/27/2021 | Asia | Pakistan | Islamabad |
| Pakistan/NIH-S12/2021 | EPI_ISL_1969995 | 4/26/2021 | Asia | Pakistan | Islamabad |
| Pakistan/PPHRL-01/2021 | EPI_ISL_3371848 | 7/31/2021 | Asia | Pakistan | Punjab PK |
| Pakistan/PPHRL-AH002/2021 | EPI_ISL_2544506 | 1/15/2021 | Asia | Pakistan | Punjab PK |
| Pakistan/PPHRL-AH073//2021 | EPI_ISL_3372650 | 7/30/2021 | Asia | Pakistan | Punjab PK |

| | | | | | |
|---|---|---|---|---|---|
| Palestine/23/2020 | EPI_ISL_596510 | 3/22/2020 | Asia | Palestine | Ramallah and al-Bireh Governorate |
| Palestine/34/2020 | EPI_ISL_596519 | 3/4/2020 | Asia | Palestine | West Bank |
| Palestine/88/2020 | EPI_ISL_596561 | 6/22/2020 | Asia | Palestine | Hebron |
| Palestine/AAS44/2021 | EPI_ISL_1273102 | 1/4/2021 | Asia | Palestine | Ramallah and al-Bireh Governorate |
| Panama/332238/2020 | EPI_ISL_496705 | 3/14/2020 | North America | Panama | Cocle |
| Panama/332311/2020 | EPI_ISL_496709 | 3/22/2020 | North America | Panama | Panama Oeste |
| Panama/333568/2020 | EPI_ISL_496748 | 3/16/2020 | North America | Panama | Cocle |
| Panama/334280/2020 | EPI_ISL_496772 | 3/28/2020 | North America | Panama | Herrera |
| Panama/334655/2020 | EPI_ISL_496790 | 3/31/2020 | North America | Panama | Cocle |
| Panama/335862/2020 | EPI_ISL_496822 | 4/3/2020 | North America | Panama | Panama Oeste |
| Panama/335941/2020 | EPI_ISL_496829 | 4/3/2020 | North America | Panama | Panama Oeste |
| Panama/336320/2020 | EPI_ISL_496841 | 4/4/2020 | North America | Panama | Panama Center |
| Panama/GMI-PA314/2020 | EPI_ISL_1502831 | 4/11/2020 | North America | Panama | Veraguas |
| Panama/GMI-PA329551/2020 | EPI_ISL_1502818 | 3/14/2020 | North America | Panama | Veraguas |
| Panama/GMI-PA338765/2020 | EPI_ISL_1502828 | 4/9/2020 | North America | Panama | Bocas del Toro |
| Panama/GMI-PA340450/2020 | EPI_ISL_1502837 | 4/13/2020 | North America | Panama | Darien |
| Panama/GMI-PA341363/2020 | EPI_ISL_1225330 | 4/14/2020 | North America | Panama | Darien |
| Panama/GMI-PA344037/2020 | EPI_ISL_1225346 | 4/19/2020 | North America | Panama | Herrera |
| Panama/GMI-PA344204/2020 | EPI_ISL_1502841 | 4/20/2020 | North America | Panama | Bocas del Toro |
| Panama/GMI-PA345048/2020 | EPI_ISL_1225352 | 4/20/2020 | North America | Panama | Cocle |

| | | | | | |
|---|---|---|---|---|---|
| Panama/GMI-PA345365/2020 | EPI_ISL_1225355 | 4/21/2020 | North America | Panama | Los Santos |
| Panama/GMI-PA347337/2020 | EPI_ISL_1225364 | 4/23/2020 | North America | Panama | Herrera |
| Panama/GMI-PA362699/2020 | EPI_ISL_1225418 | 5/16/2020 | North America | Panama | Cocle |
| Panama/GMI-PA369149/2020 | EPI_ISL_1225444 | 5/28/2020 | North America | Panama | Panama Center |
| Panama/GMI-PA369162/2020 | EPI_ISL_1225450 | 5/28/2020 | North America | Panama | Bocas del Toro |
| Panama/GMI-PA369167/2020 | EPI_ISL_1225453 | 5/28/2020 | North America | Panama | Bocas del Toro |
| Panama/GMI-PA369170/2020 | EPI_ISL_1225455 | 5/28/2020 | North America | Panama | Herrera |
| Panama/GMI-PA376271/2020 | EPI_ISL_1001457 | 6/7/2020 | North America | Panama | Panama City |
| Panama/GMI-PA380508/2020 | EPI_ISL_1225466 | 6/11/2020 | North America | Panama | Darien |
| Panama/GMI-PA387533/2020 | EPI_ISL_1225472 | 6/17/2020 | North America | Panama | Chiriqui |
| Panama/GMI-PA389033/2020 | EPI_ISL_1225475 | 6/18/2020 | North America | Panama | Chiriqui |
| Panama/GMI-PA392814/2020 | EPI_ISL_1225483 | 6/22/2020 | North America | Panama | Veraguas |
| Panama/GMI-PA404218/2020 | EPI_ISL_1225500 | 6/30/2020 | North America | Panama | Cocle |
| Panama/GMI-PA425903/2020 | EPI_ISL_1225513 | 7/17/2020 | North America | Panama | Bocas del Toro |
| Panama/GMI-PA436432/2020 | EPI_ISL_1225524 | 7/28/2020 | North America | Panama | Colon Province |
| Panama/GMI-PA437737/2020 | EPI_ISL_1225529 | 7/29/2020 | North America | Panama | Bocas del Toro |
| Panama/GMI-PA438718/2020 | EPI_ISL_1225531 | 7/30/2020 | North America | Panama | Herrera |
| Panama/GMI-PA447387/2020 | EPI_ISL_1225539 | 8/4/2020 | North America | Panama | Los Santos |
| Panama/GMI-PA450871/2020 | EPI_ISL_1225542 | 7/31/2020 | North America | Panama | Darien |
| Panama/GMI-PA451586/2020 | EPI_ISL_1225543 | 8/8/2020 | North America | Panama | Los Santos |

| | | | | | |
|---|---|---|---|---|---|
| Panama/GMI-PA461381/2020 | EPI_ISL_1225549 | 8/16/2020 | North America | Panama | Los Santos |
| Panama/GMI-PA463358/2020 | EPI_ISL_1225556 | 8/18/2020 | North America | Panama | Herrera |
| Panama/GMI-PA571783/2020 | EPI_ISL_1225574 | 11/29/2020 | North America | Panama | Panama Center |
| Panama/GMI-PA627940/2020 | EPI_ISL_1502937 | 12/29/2020 | North America | Panama | Herrera |
| Panama/GMI-PA650555/2021 | EPI_ISL_1502978 | 1/8/2021 | North America | Panama | Panama Oeste |
| Panama/GMI-PA670198/2021 | EPI_ISL_1503122 | 1/16/2021 | North America | Panama | Colon Province |
| Panama/GMI-PA705926/2021 | EPI_ISL_1503104 | 2/17/2021 | North America | Panama | Panama Oeste |
| Panama/GMI-PA707161/2021 | EPI_ISL_1503114 | 2/20/2021 | North America | Panama | San Miguelito |
| PapuaNewGuinea/13/2020 | EPI_ISL_693483 | 10/6/2020 | Oceania | Papua New Guinea | Papua New Guinea |
| PapuaNewGuinea/3/2020 | EPI_ISL_693473 | 8/21/2020 | Oceania | Papua New Guinea | Papua New Guinea |
| PapuaNewGuinea/7/2020 | EPI_ISL_693477 | 8/29/2020 | Oceania | Papua New Guinea | Papua New Guinea |
| PapuaNewGuinea/8/2020 | EPI_ISL_693478 | 9/1/2020 | Oceania | Papua New Guinea | Papua New Guinea |
| PapuaNewGuinea/PNG183/2021 | EPI_ISL_1424591 | 2/26/2021 | Oceania | Papua New Guinea | Papua New Guinea |
| PapuaNewGuinea/PNG293/2021 | EPI_ISL_1424680 | 1/1/2021 | Oceania | Papua New Guinea | Papua New Guinea |
| Paraguay/33615/2020 | EPI_ISL_1340764 | 5/5/2020 | South America | Paraguay | Paraguay |
| Paraguay/cyr_7382/2021 | EPI_ISL_2234907 | 3/2/2021 | South America | Paraguay | Central Paraguay |
| Paraguay/iics_12288/2021 | EPI_ISL_2444830 | 4/26/2021 | South America | Paraguay | Central Paraguay |
| Paraguay/iics_12458/2021 | EPI_ISL_2444823 | 4/28/2021 | South America | Paraguay | Central Paraguay |
| Paraguay/iics_12537/2021 | EPI_ISL_2444819 | 5/3/2021 | South America | Paraguay | Central Paraguay |
| Paraguay/iics_12700/2021 | EPI_ISL_2444814 | 5/1/2021 | South America | Paraguay | Central Paraguay |
| Paraguay/lcsp_185951/2021 | EPI_ISL_3132169 | 7/7/2021 | South America | Paraguay | Central Paraguay |

| | | | | | |
|---|---|---|---|---|---|
| Paraguay/lcsp_187766/2021 | EPI_ISL_3132170 | 7/8/2021 | South America | Paraguay | Central Paraguay |
| Peru/AMA-UPCH-0272/2020 | EPI_ISL_729916 | 9/21/2020 | South America | Peru | Amazonas PE |
| Peru/C02m16/2021 | EPI_ISL_3050705 | 6/8/2021 | South America | Peru | Callao |
| Peru/C03m30/2021 | EPI_ISL_3372591 | 7/12/2021 | South America | Peru | Callao |
| Peru/C03m42/2021 | EPI_ISL_3372601 | 7/14/2021 | South America | Peru | Callao |
| Peru/CAL-INS-654/2020 | EPI_ISL_1111114 | 4/1/2020 | South America | Peru | Callao |
| Peru/CUS-INS-879/2020 | EPI_ISL_1534653 | 7/24/2020 | South America | Peru | Cuzco |
| Peru/CUS-UPCH-0820/2021 | EPI_ISL_3020268 | 6/1/2021 | South America | Peru | Cuzco |
| Peru/LAM-INS-278/2020 | EPI_ISL_1093172 | 8/25/2020 | South America | Peru | Lambayeque |
| Peru/LAM-INS-722/2020 | EPI_ISL_1111182 | 3/29/2020 | South America | Peru | Lambayeque |
| Peru/LIM-INS-1543/2021 | EPI_ISL_2921475 | 4/29/2021 | South America | Peru | Lima |
| Peru/LIM-INS-329/2020 | EPI_ISL_1111227 | 4/1/2020 | South America | Peru | Lima |
| Peru/LIM-INS-869/2020 | EPI_ISL_1534645 | 11/30/2020 | South America | Peru | Lima |
| Peru/LIM-UPCH-0164/2020 | EPI_ISL_729872 | 10/14/2020 | South America | Peru | Lima |
| Peru/LIM-UPCH-0449/2021 | EPI_ISL_1629795 | 3/1/2021 | South America | Peru | Lima |
| Peru/PAS-INS-982/2021 | EPI_ISL_2536755 | 4/16/2021 | South America | Peru | Pasco |
| Peru/TUM-INS-2305/2021 | EPI_ISL_3401492 | 5/14/2021 | South America | Peru | Tumbes |
| Peru/TUM-INS-2309/2021 | EPI_ISL_3401495 | 5/14/2021 | South America | Peru | Tumbes |
| Philippines/PH-PGC-00213/2020 | EPI_ISL_2153998 | 10/31/2020 | Asia | Philippines | Manila |
| Philippines/PH-PGC-00225/2020 | EPI_ISL_2153883 | 12/27/2020 | Asia | Philippines | Manila |

| | | | | | |
|---|---|---|---|---|---|
| Philippines/PH-PGC-03442/2020 | EPI_ISL_2156710 | 11/23/2020 | Asia | Philippines | Manila |
| Philippines/PH-PGC-21512/2021 | EPI_ISL_2859774 | 3/28/2021 | Asia | Philippines | Central Visayas |
| Philippines/PH-PGC-21513/2021 | EPI_ISL_2859728 | 3/28/2021 | Asia | Philippines | Central Visayas |
| Philippines/PH-PGC-44655/2021 | EPI_ISL_2859234 | 4/18/2021 | Asia | Philippines | Central Luzon |
| Philippines/PH-PGC-44766/2021 | EPI_ISL_2859180 | 4/23/2021 | Asia | Philippines | Manila |
| Philippines/PH-PGC-45991/2021 | EPI_ISL_2859179 | 5/5/2021 | Asia | Philippines | Manila |
| Philippines/PH-PGC-46015/2021 | EPI_ISL_2859237 | 5/8/2021 | Asia | Philippines | Manila |
| Philippines/PH-RITM-0020/2020 | EPI_ISL_491474 | 6/22/2020 | Asia | Philippines | Manila |
| Philippines/PH-RITM-0028/2020 | EPI_ISL_833333 | 9/26/2020 | Asia | Philippines | Calabarzon |
| Philippines/PH-RITM-0062/2020 | EPI_ISL_3105871 | 4/9/2020 | Asia | Philippines | Manila |
| Poland/15SNR21_wsserze/2021 | EPI_ISL_3251450 | 7/23/2021 | Europe | Poland | Warminsko-Mazurskie |
| Poland/2108-050_wsselodz/2021 | EPI_ISL_3332847 | 7/28/2021 | Europe | Poland | Lubelskie |
| Poland/2108-136_wsselodz/2021 | EPI_ISL_3375157 | 8/1/2021 | Europe | Poland | Zachodniopomorskie |
| Poland/2108-144_wsselodz/2021 | EPI_ISL_3375163 | 8/1/2021 | Europe | Poland | Zachodniopomorskie |
| Poland/IHG_PAS_4_67/2020 | EPI_ISL_485399 | 6/1/2020 | Europe | Poland | Opolskie |
| Poland/Pomorskie_MWB_60/2020 | EPI_ISL_906752 | 8/30/2020 | Europe | Poland | Pomorskie |
| Poland/Pomorskie_MWB_82/2021 | EPI_ISL_1040999 | 1/27/2021 | Europe | Poland | Pomorskie |
| Poland/PZH-GUM-0609/2021 | EPI_ISL_2140250 | 4/26/2021 | Europe | Poland | Zachodniopomorskie |
| Poland/PZH-GUM-0634/2021 | EPI_ISL_2140273 | 4/26/2021 | Europe | Poland | Zachodniopomorskie |
| Poland/PZH-GUM-1802/2021 | EPI_ISL_2897097 | 6/30/2021 | Europe | Poland | Lubelskie |
| Poland/PZH-GUM-1805/2021 | EPI_ISL_2897099 | 6/30/2021 | Europe | Poland | Lubelskie |
| Poland/PZH-LUM-1373/2021 | EPI_ISL_2543743 | 5/28/2021 | Europe | Poland | Wielkopolskie |
| Poland/WGS-CoV_PZH-LUM-0333/2021 | EPI_ISL_1908394 | 4/13/2021 | Europe | Poland | Dolno≈õlƒÖskie |
| Poland/WGS-CoV_PZH-LUM-1043/2021 | EPI_ISL_2365753 | 5/13/2021 | Europe | Poland | Malopolskie |
| Portugal/PT0141/2020 | EPI_ISL_453857 | 3/27/2020 | Europe | Portugal | Portugal |
| Portugal/PT10530/2021 | EPI_ISL_2810460 | 6/22/2021 | Europe | Portugal | Portugal |
| Portugal/PT11379/2021 | EPI_ISL_2989015 | 7/5/2021 | Europe | Portugal | Portugal |

| | | | | | |
|---|---|---|---|---|---|
| Portugal/PT11380/2021 | EPI_ISL_2989302 | 7/6/2021 | Europe | Portugal | Portugal |
| Portugal/PT1160/2020 | EPI_ISL_511707 | 4/1/2020 | Europe | Portugal | Portugal |
| Portugal/PT13793/2021 | EPI_ISL_3432077 | 8/1/2021 | Europe | Portugal | Portugal |
| Portugal/PT13795/2021 | EPI_ISL_3432079 | 8/1/2021 | Europe | Portugal | Portugal |
| Portugal/PT7187/2021 | EPI_ISL_2004323 | 4/19/2021 | Europe | Portugal | Portugal |
| Portugal/PT7465/2021 | EPI_ISL_2249047 | 5/11/2021 | Europe | Portugal | Portugal |
| Portugal/PT8394/2021 | EPI_ISL_2536041 | 4/27/2021 | Europe | Portugal | Portugal |
| Portugal/PT8622/2021 | EPI_ISL_2628805 | 6/4/2021 | Europe | Portugal | Portugal |
| Portugal/PT9806/2021 | EPI_ISL_2796193 | 5/31/2021 | Europe | Portugal | Portugal |
| Qatar/DA-USAFSAM-S3135/2021 | EPI_ISL_2661492 | 5/23/2021 | Asia | Qatar | Ad-Dawhah |
| Qatar/DA-USAFSAM-S3140/2021 | EPI_ISL_2661497 | 5/23/2021 | Asia | Qatar | Ad-Dawhah |
| Qatar/DA-USAFSAM-S4134/2021 | EPI_ISL_3048165 | 6/6/2021 | Asia | Qatar | Ad-Dawhah |
| Qatar/QA-QU_09-1-A5/2020 | EPI_ISL_1713076 | 4/29/2020 | Asia | Qatar | Doha |
| Qatar/QA-QU_12-10-A11/2020 | EPI_ISL_1713195 | 10/26/2020 | Asia | Qatar | Doha |
| Qatar/QA-QU_12-10-B8/2020 | EPI_ISL_1713207 | 10/25/2020 | Asia | Qatar | Doha |
| Qatar/QA-QU_12-11-B2/2020 | EPI_ISL_1713249 | 11/9/2020 | Asia | Qatar | Doha |
| Qatar/QA-QU_12-11-B5/2020 | EPI_ISL_1713252 | 11/24/2020 | Asia | Qatar | Doha |
| Qatar/QA-QU_12-11-F9/2020 | EPI_ISL_1713274 | 11/9/2020 | Asia | Qatar | Doha |
| Qatar/QA-QU_12-8-F6/2020 | EPI_ISL_1713352 | 8/19/2020 | Asia | Qatar | Doha |
| Qatar/QA-QU_18-E6/2021 | EPI_ISL_1713902 | 2/2/2021 | Asia | Qatar | Doha |
| Qatar/QA-WCMQ_FD17571328/2020 | EPI_ISL_1714322 | 12/29/2020 | Asia | Qatar | Doha |
| Qatar/QA.QU_18.12.C3/2021 | EPI_ISL_2274381 | 4/17/2021 | Asia | Qatar | Doha |
| Qatar/QA.QU_18.15.E5/2021 | EPI_ISL_2842975 | 4/29/2021 | Asia | Qatar | Doha |
| Qatar/QA.QU_18.19.C1/2021 | EPI_ISL_2843263 | 6/7/2021 | Asia | Qatar | Doha |
| RepublicoftheCongo/RC-016/2021 | EPI_ISL_2978201 | 6/4/2021 | Africa | Republic of the Congo | Republic of the Congo |
| RepublicoftheCongo/RC-021/2021 | EPI_ISL_2978206 | 5/19/2021 | Africa | Republic of the Congo | Republic of the Congo |

| | | | | | |
|---|---|---|---|---|---|
| RepublicoftheCongo/RC-051/2021 | EPI_ISL_2978221 | 5/18/2021 | Africa | Republic of the Congo | Republic of the Congo |
| Romania/284468/2020 | EPI_ISL_455479 | 5/14/2020 | Europe | Romania | Bucharest |
| Romania/B-18321/2021 | EPI_ISL_3259624 | 7/27/2021 | Europe | Romania | Bucharest |
| Romania/B-18336/2021 | EPI_ISL_3341994 | 7/27/2021 | Europe | Romania | Bucharest |
| Romania/Bucuresti_450700/2021 | EPI_ISL_3021349 | 5/9/2021 | Europe | Romania | Bucharest |
| Romania/CT-16216/2021 | EPI_ISL_2687992 | 6/2/2021 | Europe | Romania | Constanta |
| Romania/Dolj_455008/2021 | EPI_ISL_3021370 | 5/18/2021 | Europe | Romania | Dolj |
| Romania/IF-12634/2021 | EPI_ISL_2099853 | 4/29/2021 | Europe | Romania | Ilfov |
| Romania/IS-16650/2021 | EPI_ISL_2932516 | 6/12/2021 | Europe | Romania | Iasi |
| Romania/IS-18860/2021 | EPI_ISL_3342029 | 8/3/2021 | Europe | Romania | Iasi |
| Romania/MS-11418/2021 | EPI_ISL_1969968 | 4/21/2021 | Europe | Romania | Mures |
| Romania/ROSV_12723/2020 | EPI_ISL_491048 | 6/10/2020 | Europe | Romania | Suceava |
| Romania/TL-18771/2021 | EPI_ISL_3342014 | 8/2/2021 | Europe | Romania | Tulcea |
| Russia/MOS-CRIE-L106E0365b/2021 | EPI_ISL_3101257 | 4/9/2021 | Europe | Russia | Moscow Oblast |
| Russia/MOW-CRIE-D186B0049b/2021 | EPI_ISL_2626431 | 1/2/2021 | Europe | Russia | Moscow Oblast |
| Russia/MOW-CRIE-L188M0809/2021 | EPI_ISL_3101105 | 5/17/2021 | Europe | Russia | Moscow Oblast |
| Russia/MOW-RII-MH22307S/2021 | EPI_ISL_2816258 | 6/12/2021 | Europe | Russia | Moscow Oblast |
| Russia/MOW-RII-MH29548S/2021 | EPI_ISL_3086656 | 7/6/2021 | Europe | Russia | Moscow Oblast |
| Russia/MOW-RII-MH30207S/2021 | EPI_ISL_3123078 | 7/13/2021 | Europe | Russia | Moscow Oblast |
| Russia/PSK-RII-MH15748S/2021 | EPI_ISL_1652623 | 3/1/2021 | Europe | Russia | Pskov Oblast |
| Russia/SAR-RII-MH25570S/2021 | EPI_ISL_3122890 | 6/11/2021 | Europe | Russia | Saratov |
| Russia/SPE-RII-45232S/2021 | EPI_ISL_3454793 | 8/3/2021 | Europe | Russia | Saint-Petersburg |
| Russia/SPE-RII-45517S/2021 | EPI_ISL_3454804 | 8/8/2021 | Europe | Russia | Saint-Petersburg |
| Russia/SPE-RII-MH17259S/2021 | EPI_ISL_2385244 | 4/29/2021 | Europe | Russia | Saint-Petersburg |
| Russia/SPE-RII-MH17266S/2021 | EPI_ISL_2385247 | 4/26/2021 | Europe | Russia | Saint-Petersburg |
| Russia/SPE-RII-MH17295S/2021 | EPI_ISL_2385271 | 4/13/2021 | Europe | Russia | Saint-Petersburg |
| Russia/SPE-RII-MH18491S/2021 | EPI_ISL_2523605 | 5/24/2021 | Europe | Russia | Saint-Petersburg |

| | | | | | |
|---|---|---|---|---|---|
| Russia/TA-KFU_169/2020 | EPI_ISL_1859503 | 8/23/2020 | Europe | Russia | Tatarstan |
| Russia/TA-KFU_29/2020 | EPI_ISL_1859579 | 7/27/2020 | Europe | Russia | Tatarstan |
| Russia/TA-KFU_35/2020 | EPI_ISL_1859633 | 7/24/2020 | Europe | Russia | Tatarstan |
| Rwanda/NRLNAT1068/2020 | EPI_ISL_925900 | 11/25/2020 | Africa | Rwanda | Kigali |
| Rwanda/NRLNAT2023/2020 | EPI_ISL_960278 | 8/21/2020 | Africa | Rwanda | Kigali |
| Rwanda/NRLNAT3049/2021 | EPI_ISL_1301735 | 1/20/2021 | Africa | Rwanda | Kigali |
| Rwanda/NRLNAT4029/2021 | EPI_ISL_2362516 | 3/23/2021 | Africa | Rwanda | Kigali |
| Rwanda/NRLNAT4031/2021 | EPI_ISL_2362518 | 3/23/2021 | Africa | Rwanda | Kigali |
| Rwanda/NRLNAT4038/2021 | EPI_ISL_2362525 | 4/18/2021 | Africa | Rwanda | Kigali |
| Rwanda/NRLNAT4056/2021 | EPI_ISL_2521993 | 5/2/2021 | Africa | Rwanda | Kigali |
| Rwanda/NRLNAT4060/2021 | EPI_ISL_2521997 | 5/21/2021 | Africa | Rwanda | Kigali |
| Rwanda/NRLNAT4066/2021 | EPI_ISL_2522096 | 4/25/2021 | Africa | Rwanda | Kigali |
| Rwanda/NRLNAT4096/2021 | EPI_ISL_2828480 | 6/20/2021 | Africa | Rwanda | Rwanda |
| Rwanda/NRLNAT4157/2021 | EPI_ISL_3012193 | 7/7/2021 | Africa | Rwanda | Kigali |
| Rwanda/NRLNAT4169/2021 | EPI_ISL_3012204 | 7/5/2021 | Africa | Rwanda | Kigali |
| Rwanda/NRLNAT4202/2021 | EPI_ISL_3012235 | 6/25/2021 | Africa | Rwanda | Kigali |
| SaintKittsandNevis/72432/2021 | EPI_ISL_2756582 | 5/31/2021 | North America | Saint Kitts and Nevis | Saint Kitts and Nevis |
| SaintKittsandNevis/73322/2021 | EPI_ISL_2716602 | 6/5/2021 | North America | Saint Kitts and Nevis | Saint Kitts and Nevis |
| SaintLucia/59975/2021 | EPI_ISL_2478952 | 3/13/2021 | North America | Saint Lucia | Saint Lucia |
| SaintLucia/59980/2021 | EPI_ISL_2478956 | 3/16/2021 | North America | Saint Lucia | Saint Lucia |
| SaintLucia/59981/2021 | EPI_ISL_2478957 | 3/15/2021 | North America | Saint Lucia | Saint Lucia |
| SaintLucia/66447/2021 | EPI_ISL_2626661 | 4/27/2021 | North America | Saint Lucia | Saint Lucia |
| SaintLucia/66451/2021 | EPI_ISL_2626663 | 4/29/2021 | North America | Saint Lucia | Saint Lucia |
| SaintLucia/74394/2021 | EPI_ISL_2967979 | 6/12/2021 | North America | Saint Lucia | Saint Lucia |

| | | | | | |
|---|---|---|---|---|---|
| SaintLucia/74401/2021 | EPI_ISL_2967984 | 5/16/2021 | North America | Saint Lucia | Saint Lucia |
| SaintLucia/74403/2021 | EPI_ISL_2967986 | 5/13/2021 | North America | Saint Lucia | Saint Lucia |
| SaintVincentandtheGrenadines/71763/2021 | EPI_ISL_2678171 | 5/26/2021 | North America | Saint Vincent and the Grenadines | Saint Vincent and the Grenadines |
| SaintVincentandtheGrenadines/71767/2021 | EPI_ISL_2678165 | 5/26/2021 | North America | Saint Vincent and the Grenadines | Saint Vincent and the Grenadines |
| SaudiArabia/DM-21-102044A/2021 | EPI_ISL_3236928 | 4/14/2021 | Asia | Saudi Arabia | Jeddah |
| SaudiArabia/DM-21-110634A/2021 | EPI_ISL_3237047 | 4/25/2021 | Asia | Saudi Arabia | Jeddah |
| SaudiArabia/DM-21-111027A/2021 | EPI_ISL_3237049 | 4/26/2021 | Asia | Saudi Arabia | Jeddah |
| SaudiArabia/KAUST-RIYADH1454/2020 | EPI_ISL_678172 | 6/27/2020 | Asia | Saudi Arabia | Riyadh |
| Scotland/QEUH-1B1E03E/2021 | EPI_ISL_3444896 | 8/6/2021 | Europe | United Kingdom | Scotland |
| Senegal/SC20-262/2021 | EPI_ISL_3152071 | 1/27/2021 | Africa | Senegal | Dakar |
| Senegal/SC20-347/2020 | EPI_ISL_3152075 | 12/23/2020 | Africa | Senegal | Dakar |
| Senegal/SC20-5112/2020 | EPI_ISL_1697387 | 9/3/2020 | Africa | Senegal | Dakar |
| Senegal/SN-IR1-38865/2021 | EPI_ISL_2887851 | 6/1/2021 | Africa | Senegal | Dakar |
| Senegal/SN-IR1-40292/2021 | EPI_ISL_2887852 | 5/31/2021 | Africa | Senegal | Dakar |
| Senegal/SN-IR2-0019063/2021 | EPI_ISL_2620889 | 5/6/2021 | Africa | Senegal | Dakar |
| Senegal/SN-IR8-1183/2021 | EPI_ISL_2873829 | 6/5/2021 | Africa | Senegal | Dakar |
| Serbia/Belgrade-C329421811/2020 | EPI_ISL_955166 | 11/18/2020 | Europe | Serbia | Belgrade |
| Serbia/C17700/2020 | EPI_ISL_1056959 | 8/1/2020 | Europe | Serbia | Serbia |
| Serbia/Cacak-3301880905/2021 | EPI_ISL_3163954 | 5/9/2021 | Europe | Serbia | Moravica District |
| Serbia/Cacak-3444740207/2021 | EPI_ISL_3163962 | 7/2/2021 | Europe | Serbia | Moravica District |
| Serbia/Kraljevo-ZJZ00012906/2021 | EPI_ISL_2932530 | 6/29/2021 | Europe | Serbia | Ra≈°ka District |
| Serbia/L5565/2020 | EPI_ISL_1057040 | 9/29/2020 | Europe | Serbia | Serbia |
| Serbia/Smederevo-3322461405/2021 | EPI_ISL_3163943 | 5/14/2021 | Europe | Serbia | Podunavlje District |
| Serbia/ZJZKV0002-29-06/2021 | EPI_ISL_3102051 | 6/29/2021 | Europe | Serbia | Ra≈°ka District |
| Serbia/ZJZKV00030607/2021 | EPI_ISL_2986987 | 7/6/2021 | Europe | Serbia | Ra≈°ka District |
| Shanghai/SH0085/2020 | EPI_ISL_416385 | 1/30/2020 | Asia | China | Shanghai |

| | | | | | |
|---|---|---|---|---|---|
| SierraLeone/NMIMR-SLE-SLSEQ21/2021 | EPI_ISL_2001068 | 1/14/2021 | Africa | Sierra Leone | Sierra Leone |
| SierraLeone/SL15/2021 | EPI_ISL_2716630 | 1/27/2021 | Africa | Sierra Leone | Sierra Leone |
| Singapore/1063/2021 | EPI_ISL_2508714 | 5/17/2021 | Asia | Singapore | Singapore |
| Singapore/125/2021 | EPI_ISL_937515 | 1/31/2021 | Asia | Singapore | Singapore |
| Singapore/1318/2020 | EPI_ISL_648806 | 5/21/2020 | Asia | Singapore | Singapore |
| Singapore/1367/2021 | EPI_ISL_2509022 | 5/29/2021 | Asia | Singapore | Singapore |
| Singapore/1503/2021 | EPI_ISL_2621920 | 6/7/2021 | Asia | Singapore | Singapore |
| Singapore/1512/2020 | EPI_ISL_768628 | 12/24/2020 | Asia | Singapore | Singapore |
| Singapore/1543/2021 | EPI_ISL_2621960 | 6/11/2021 | Asia | Singapore | Singapore |
| Singapore/2023/2021 | EPI_ISL_3052751 | 7/15/2021 | Asia | Singapore | Singapore |
| Singapore/234/2021 | EPI_ISL_1164355 | 2/26/2021 | Asia | Singapore | Singapore |
| Singapore/2655/2021 | EPI_ISL_3188683 | 7/28/2021 | Asia | Singapore | Singapore |
| Singapore/3360/2021 | EPI_ISL_3387230 | 8/8/2021 | Asia | Singapore | Singapore |
| Singapore/3463/2021 | EPI_ISL_3393652 | 8/6/2021 | Asia | Singapore | Singapore |
| Singapore/683/2021 | EPI_ISL_1816928 | 4/22/2021 | Asia | Singapore | Singapore |
| Singapore/685/2021 | EPI_ISL_1816930 | 4/22/2021 | Asia | Singapore | Singapore |
| Singapore/815/2020 | EPI_ISL_516815 | 8/11/2020 | Asia | Singapore | Singapore |
| Singapore/894/2020 | EPI_ISL_536435 | 2/2/2020 | Asia | Singapore | Singapore |
| Singapore/913/2020 | EPI_ISL_538436 | 2/20/2020 | Asia | Singapore | Singapore |
| SintMaarten/SX-RIVM-29848/2021 | EPI_ISL_2093157 | 4/23/2021 | North America | Sint Maarten | Sint Maarten |
| SintMaarten/SX-RIVM-29902/2021 | EPI_ISL_2093182 | 4/23/2021 | North America | Sint Maarten | Sint Maarten |
| SintMaarten/SX-RIVM-34863/2021 | EPI_ISL_2474205 | 5/14/2021 | North America | Sint Maarten | Sint Maarten |
| SintMaarten/SX-RIVM-36317/2021 | EPI_ISL_2474452 | 5/20/2021 | North America | Sint Maarten | Sint Maarten |
| SintMaarten/SX-RIVM-39248/2021 | EPI_ISL_2610709 | 6/9/2021 | North America | Sint Maarten | Sint Maarten |
| SintMaarten/SX-RIVM-40555/2021 | EPI_ISL_2673280 | 6/16/2021 | North America | Sint Maarten | Sint Maarten |

| | | | | | |
|---|---|---|---|---|---|
| SintMaarten/SX-RIVM-42371/2021 | EPI_ISL_2981854 | 7/5/2021 | North America | Sint Maarten | Sint Maarten |
| SintMaarten/SX-RIVM-50077/2021 | EPI_ISL_3390590 | 7/26/2021 | North America | Sint Maarten | Sint Maarten |
| Slovakia/020282837/2021 | EPI_ISL_1280095 | 2/2/2021 | Europe | Slovakia | Trnava |
| Slovakia/113060242E/2020 | EPI_ISL_1280140 | 11/30/2020 | Europe | Slovakia | Bratislava |
| Slovakia/CeMM4299/2021 | EPI_ISL_1495211 | 1/7/2021 | Europe | Slovakia | Kosice |
| Slovakia/UKBA-1653/2021 | EPI_ISL_1239415 | 2/20/2021 | Europe | Slovakia | Zilina |
| Slovakia/UKBA-1822/2021 | EPI_ISL_1234401 | 2/28/2021 | Europe | Slovakia | Slovakia |
| Slovakia/UKBA-3043/2021 | EPI_ISL_2151034 | 4/30/2021 | Europe | Slovakia | Nitra |
| Slovakia/UKBA-316/2020 | EPI_ISL_583484 | 9/10/2020 | Europe | Slovakia | Bratislava |
| Slovakia/UKBA-3309/2021 | EPI_ISL_2562149 | 5/24/2021 | Europe | Slovakia | Trnava |
| Slovakia/UKBA-3663/2021 | EPI_ISL_2894467 | 6/25/2021 | Europe | Slovakia | Trenƒç√≠n |
| Slovakia/UKBA-3937/2021 | EPI_ISL_3277429 | 7/15/2021 | Europe | Slovakia | Trnava |
| Slovakia/UVZ_PL14_D12_9153/2021 | EPI_ISL_1749451 | 4/17/2021 | Europe | Slovakia | Bratislava |
| Slovakia/UVZ_PL17_E5_11318/2021 | EPI_ISL_2611881 | 5/19/2021 | Europe | Slovakia | Trnava |
| Slovakia/UVZ_PL25_E8_14133/2021 | EPI_ISL_3100334 | 6/21/2021 | Europe | Slovakia | Zilina |
| Slovakia/UVZ_PL27_E5_14393/2021 | EPI_ISL_3117850 | 7/8/2021 | Europe | Slovakia | Bratislava |
| Slovakia/UVZ_PL30_C8_15560/2021 | EPI_ISL_3452600 | 8/2/2021 | Europe | Slovakia | Bratislava |
| Slovakia/UVZ_PL30_F10_15631/2021 | EPI_ISL_3452617 | 8/3/2021 | Europe | Slovakia | Kosice |
| Slovenia/08-148288-MB/2021 | EPI_ISL_3316710 | 8/1/2021 | Europe | Slovenia | Podravska |
| Slovenia/08-148292-MB/2021 | EPI_ISL_3316713 | 8/1/2021 | Europe | Slovenia | Podravska |
| Slovenia/104383/2020 | EPI_ISL_2644389 | 9/23/2020 | Europe | Slovenia | Slovenia |
| Slovenia/127424/2020 | EPI_ISL_2886956 | 10/10/2020 | Europe | Slovenia | Slovenia |
| Slovenia/156810/2020 | EPI_ISL_1098776 | 10/24/2020 | Europe | Slovenia | Slovenia |
| Slovenia/17-039988-KR/2021 | EPI_ISL_2982948 | 6/24/2021 | Europe | Slovenia | Osrednjeslovenska |
| Slovenia/184809/2020 | EPI_ISL_1964494 | 11/4/2020 | Europe | Slovenia | Slovenia |
| Slovenia/208131/2021 | EPI_ISL_2030439 | 4/17/2021 | Europe | Slovenia | Slovenia |
| Slovenia/230478/2021 | EPI_ISL_2322341 | 5/2/2021 | Europe | Slovenia | Slovenia |

| | | | | | |
|---|---|---|---|---|---|
| Slovenia/296835/2021 | EPI_ISL_3039364 | 6/21/2021 | Europe | Slovenia | Slovenia |
| Slovenia/44883/2020 | EPI_ISL_1511033 | 6/26/2020 | Europe | Slovenia | Slovenia |
| Slovenia/5C-012189-KP/2021 | EPI_ISL_1240247 | 2/24/2021 | Europe | Slovenia | Obalnokraska |
| Slovenia/90-023454-NM/2021 | EPI_ISL_2789139 | 5/7/2021 | Europe | Slovenia | Jugovzhodna |
| Slovenia/90-032115-NM/2021 | EPI_ISL_3317138 | 7/19/2021 | Europe | Slovenia | Jugovzhodna |
| Slovenia/90-032182-NM/2021 | EPI_ISL_3317146 | 7/20/2021 | Europe | Slovenia | Jugovzhodna |
| Slovenia/Golnik-12115-04/2021 | EPI_ISL_1939832 | 4/20/2021 | Europe | Slovenia | Gorenjska |
| Slovenia/P21-30946/2021 | EPI_ISL_979251 | 1/16/2021 | Europe | Slovenia | Slovenia |
| snowleopard/USA/KY-20-035685-003/2020 | EPI_ISL_2928447 | 12/4/2020 | North America | USA | Kentucky |
| Somalia/CV1182/2020 | EPI_ISL_2107100 | 4/21/2020 | Africa | Somalia | Benadir |
| Somalia/CV1232/2020 | EPI_ISL_2308255 | 6/8/2020 | Africa | Somalia | Awdal |
| SouthAfrica/NHLS-UCT-GP-6550/2021 | EPI_ISL_2802127 | 5/24/2021 | Africa | South Africa | Western Cape Province |
| SouthAfrica/NICD-DQ53569/2021 | EPI_ISL_2463302 | 5/11/2021 | Africa | South Africa | Gauteng |
| SouthAfrica/NICD-N00396/2020 | EPI_ISL_2447802 | 10/13/2020 | Africa | South Africa | Eastern Cape |
| SouthAfrica/NICD-N00737/2020 | EPI_ISL_1239483 | 12/22/2020 | Africa | South Africa | Northern Cape |
| SouthAfrica/NICD-R01865/2021 | EPI_ISL_2178786 | 4/23/2021 | Africa | South Africa | Mpumalanga |
| SouthAfrica/NICD-R01871/2021 | EPI_ISL_2178792 | 4/23/2021 | Africa | South Africa | Gauteng |
| SouthAfrica/NICD-R10575/2021 | EPI_ISL_3219880 | 7/13/2021 | Africa | South Africa | Gauteng |
| SouthAfrica/NICD-R11525/2021 | EPI_ISL_3451601 | 8/2/2021 | Africa | South Africa | KwaZulu-Natal |
| SouthAfrica/NICD-R11603/2021 | EPI_ISL_3451622 | 8/2/2021 | Africa | South Africa | North-West |
| SouthAfrica/Tygerberg_1180/2021 | EPI_ISL_2876367 | 6/24/2021 | Africa | South Africa | Western Cape Province |
| SouthAfrica/Tygerberg_1191/2021 | EPI_ISL_2876310 | 6/24/2021 | Africa | South Africa | Western Cape Province |
| SouthAfrica/Tygerberg_1510/2021 | EPI_ISL_3247125 | 7/19/2021 | Africa | South Africa | Western Cape Province |
| SouthKorea/KCDC2014/2020 | EPI_ISL_426187 | 2/12/2020 | Asia | South Korea | South Korea |
| SouthKorea/KDCA0488/2020 | EPI_ISL_850200 | 6/8/2020 | Asia | South Korea | South Korea |
| SouthKorea/KDCA0545/2020 | EPI_ISL_850237 | 7/22/2020 | Asia | South Korea | South Korea |
| SouthKorea/KDCA1305/2021 | EPI_ISL_1007645 | 1/15/2021 | Asia | South Korea | South Korea |

| | | | | | |
|---|---|---|---|---|---|
| SouthKorea/KDCA1718/2021 | EPI_ISL_1165006 | 1/11/2021 | Asia | South Korea | South Korea |
| SouthKorea/KDCA3458/2021 | EPI_ISL_1936637 | 4/16/2021 | Asia | South Korea | South Korea |
| SouthKorea/KDCA3640/2020 | EPI_ISL_2161133 | 12/19/2020 | Asia | South Korea | South Korea |
| SouthKorea/KDCA3770/2021 | EPI_ISL_2284595 | 5/2/2021 | Asia | South Korea | South Korea |
| SouthKorea/KDCA4118/2021 | EPI_ISL_2332427 | 4/19/2021 | Asia | South Korea | South Korea |
| SouthKorea/KDCA4165/2021 | EPI_ISL_2332460 | 5/4/2021 | Asia | South Korea | South Korea |
| SouthKorea/KDCA5646/2021 | EPI_ISL_2967363 | 6/18/2021 | Asia | South Korea | South Korea |
| SouthKorea/KDCA5647/2021 | EPI_ISL_2967364 | 6/18/2021 | Asia | South Korea | South Korea |
| SouthKorea/KDCA7208/2021 | EPI_ISL_3369120 | 7/18/2021 | Asia | South Korea | South Korea |
| SouthKorea/KDCA7461/2021 | EPI_ISL_3451927 | 7/20/2021 | Asia | South Korea | South Korea |
| SouthSudan/UG406/2021 | EPI_ISL_2450785 | 1/28/2021 | Africa | South Sudan | South Sudan |
| SouthSudan/UG433/2021 | EPI_ISL_2450805 | 2/6/2021 | Africa | South Sudan | South Sudan |
| SouthSudan/UG570/2021 | EPI_ISL_2928017 | 4/29/2021 | Africa | South Sudan | South Sudan |
| SouthSudan/UG571/2021 | EPI_ISL_2928018 | 4/29/2021 | Africa | South Sudan | South Sudan |
| SouthSudan/UG573/2021 | EPI_ISL_2928020 | 5/2/2021 | Africa | South Sudan | South Sudan |
| SouthSudan/UG576/2021 | EPI_ISL_2928023 | 6/7/2021 | Africa | South Sudan | South Sudan |
| SouthSudan/UG578/2020 | EPI_ISL_2928025 | 8/5/2020 | Africa | South Sudan | South Sudan |
| SouthSudan/UG579/2021 | EPI_ISL_2928026 | 6/2/2021 | Africa | South Sudan | South Sudan |
| SouthSudan/UG580/2021 | EPI_ISL_2928027 | 5/26/2021 | Africa | South Sudan | South Sudan |
| Spain/AN-IBV-003945/2020 | EPI_ISL_500168 | 4/7/2020 | Europe | Spain | Andalusia |
| Spain/AN-IBV-98001124/2020 | EPI_ISL_538179 | 3/27/2020 | Europe | Spain | Andalusia |
| Spain/CT-HUGTiPR013JG9H12/2021 | EPI_ISL_1719835 | 4/8/2021 | Europe | Spain | Catalunya |
| Spain/CT-HUVH-E22308/2021 | EPI_ISL_3326276 | 7/29/2021 | Europe | Spain | Catalunya |
| Spain/CT-IBV-98006533/2020 | EPI_ISL_541069 | 5/1/2020 | Europe | Spain | Catalunya |
| Spain/CT-IBV-98006562/2020 | EPI_ISL_541075 | 6/25/2020 | Europe | Spain | Catalunya |
| Spain/IB-IBV-99010803/2020 | EPI_ISL_691621 | 6/30/2020 | Europe | Spain | Balear Islands |
| Spain/MD-14272345/2021 | EPI_ISL_2179677 | 4/16/2021 | Europe | Spain | Madrid |

| | | | | | |
|---|---|---|---|---|---|
| Spain/MD-14272346/2021 | EPI_ISL_2179678 | 4/16/2021 | Europe | Spain | Madrid |
| Spain/MD-HGUGM-5770060/2021 | EPI_ISL_3386942 | 8/5/2021 | Europe | Spain | Madrid |
| Spain/MD-HGUGM-676416/2021 | EPI_ISL_2402506 | 5/25/2021 | Europe | Spain | Madrid |
| Spain/MD-HGUGM-682839/2021 | EPI_ISL_2601719 | 6/6/2021 | Europe | Spain | Madrid |
| Spain/MD-HGUGM-690511/2021 | EPI_ISL_2601703 | 6/11/2021 | Europe | Spain | Madrid |
| Spain/MD-HRYC-11968912/2021 | EPI_ISL_3432386 | 8/1/2021 | Europe | Spain | Madrid |
| Spain/MD-HRYC-71436788/2021 | EPI_ISL_3432377 | 7/29/2021 | Europe | Spain | Madrid |
| Spain/MD-IBV-99020675/2020 | EPI_ISL_1195537 | 9/10/2020 | Europe | Spain | Madrid |
| Spain/NC-CHN-01003170/2021 | EPI_ISL_2510594 | 5/22/2021 | Europe | Spain | Navarra |
| Spain/VC-IBV-98003578/2020 | EPI_ISL_537992 | 3/30/2020 | Europe | Spain | Comunitat Valenciana |
| SriLanka/aicbu124/2021 | EPI_ISL_3070833 | 7/8/2021 | Asia | Sri Lanka | Central Province |
| SriLanka/aicbu582/2021 | EPI_ISL_3070850 | 6/29/2021 | Asia | Sri Lanka | Northern Province |
| SriLanka/aicbu718/2021 | EPI_ISL_3275394 | 7/29/2021 | Asia | Sri Lanka | Colombo |
| SriLanka/aicbu743/2021 | EPI_ISL_3275465 | 8/1/2021 | Asia | Sri Lanka | Western Province |
| SriLanka/CDR100/2021 | EPI_ISL_1970350 | 4/28/2021 | Asia | Sri Lanka | North Western Province |
| SriLanka/CDR112/2021 | EPI_ISL_2481342 | 5/7/2021 | Asia | Sri Lanka | North Western Province |
| SriLanka/CDR151/2021 | EPI_ISL_2803228 | 6/16/2021 | Asia | Sri Lanka | Western Province |
| SriLanka/CDR503/2020 | EPI_ISL_525479 | 3/31/2020 | Asia | Sri Lanka | Kalutara |
| SriLanka/CDR84/2021 | EPI_ISL_1970397 | 4/28/2021 | Asia | Sri Lanka | Western Province |
| SriLanka/CMC107488/2021 | EPI_ISL_2481411 | 5/24/2021 | Asia | Sri Lanka | Colombo |
| SriLanka/NR125/2021 | EPI_ISL_1233116 | 2/3/2021 | Asia | Sri Lanka | Sri Lanka |
| SriLanka/NR23/2021 | EPI_ISL_1533847 | 3/12/2021 | Asia | Sri Lanka | Sri Lanka |
| SriLanka/NR24/2021 | EPI_ISL_1533848 | 3/12/2021 | Asia | Sri Lanka | Sri Lanka |
| Suriname/SR-124/2021 | EPI_ISL_2502440 | 1/22/2021 | South America | Suriname | Suriname |
| Suriname/SR-185/2021 | EPI_ISL_2502517 | 4/8/2021 | South America | Suriname | Suriname |
| Suriname/SR-21/2020 | EPI_ISL_517632 | 6/25/2020 | South America | Suriname | Suriname |

| | | | | | |
|---|---|---|---|---|---|
| Suriname/SR-216/2021 | EPI_ISL_2502518 | 4/17/2021 | South America | Suriname | Suriname |
| Suriname/SR-219/2021 | EPI_ISL_2502520 | 4/17/2021 | South America | Suriname | Suriname |
| Suriname/SR-318/2021 | EPI_ISL_3462737 | 5/24/2021 | South America | Suriname | Suriname |
| Suriname/SR-330/2021 | EPI_ISL_3462732 | 5/27/2021 | South America | Suriname | Suriname |
| Suriname/SR-369/2021 | EPI_ISL_3462733 | 6/10/2021 | South America | Suriname | Suriname |
| Suriname/SR-374/2021 | EPI_ISL_3462734 | 6/13/2021 | South America | Suriname | Suriname |
| Suriname/SR-470/2021 | EPI_ISL_3464726 | 7/12/2021 | South America | Suriname | Suriname |
| Suriname/SR-495/2021 | EPI_ISL_3462564 | 7/23/2021 | South America | Suriname | Suriname |
| Suriname/SR-522/2021 | EPI_ISL_3462741 | 8/1/2021 | South America | Suriname | Suriname |
| Suriname/SR-523/2021 | EPI_ISL_3462723 | 8/2/2021 | South America | Suriname | Suriname |
| Suriname/SR-RIVM-22503/2021 | EPI_ISL_1521325 | 3/5/2021 | South America | Suriname | Suriname |
| Sweden/01_SE100_test663TEST/2020 | EPI_ISL_2365964 | 11/12/2020 | Europe | Sweden | Stockholm |
| Sweden/10095305/2021 | EPI_ISL_2980226 | 6/30/2021 | Europe | Sweden | Stockholm |
| Sweden/1912611964/2021 | EPI_ISL_2210009 | 2/11/2021 | Europe | Sweden | Vastra Gotaland |
| Sweden/20-04631/2020 | EPI_ISL_430847 | 2/27/2020 | Europe | Sweden | Stockholm |
| Sweden/20244757P/2021 | EPI_ISL_2417804 | 5/14/2021 | Europe | Sweden | Blekinge |
| Sweden/21-52884/2021 | EPI_ISL_2613182 | 4/20/2021 | Europe | Sweden | Vastra Gotaland |
| Sweden/2A41382730099/2021 | EPI_ISL_2034478 | 4/20/2021 | Europe | Sweden | Norrbotten |
| Sweden/5465721776/2021 | EPI_ISL_1808139 | 3/5/2021 | Europe | Sweden | Stockholm |
| Sweden/8882815866/2021 | EPI_ISL_2980231 | 6/30/2021 | Europe | Sweden | Stockholm |
| Sweden/DC107077411/2021 | EPI_ISL_1290189 | 2/8/2021 | Europe | Sweden | Skane |
| Sweden/SE00599654/2021 | EPI_ISL_3157621 | 7/10/2021 | Europe | Sweden | Skane |
| Sweden/SE00599655/2021 | EPI_ISL_3157627 | 7/10/2021 | Europe | Sweden | Skane |

| | | | | | |
|---|---|---|---|---|---|
| Sweden/SUS0001691/2021 | EPI_ISL_2103692 | 5/5/2021 | Europe | Sweden | Skane |
| Sweden/SUS0003284/2021 | EPI_ISL_3402080 | 8/3/2021 | Europe | Sweden | Skane |
| Sweden/SUS0003335/2021 | EPI_ISL_3402131 | 8/4/2021 | Europe | Sweden | Skane |
| Switzerland/AG-UZH-IMV-3ba4a760/2021 | EPI_ISL_2769123 | 6/9/2021 | Europe | Switzerland | Aargau |
| Switzerland/BE-ETHZ-33518975/2021 | EPI_ISL_3295670 | 7/17/2021 | Europe | Switzerland | Bern |
| Switzerland/BL-ETHZ-551599/2020 | EPI_ISL_1496178 | 3/23/2021 | Europe | Switzerland | Basel-Land |
| Switzerland/BL-ETHZ-610613/2021 | EPI_ISL_2152217 | 5/4/2021 | Europe | Switzerland | Basel-Land |
| Switzerland/BS-UHB-42221337/2020 | EPI_ISL_581862 | 3/5/2020 | Europe | Switzerland | Basel-Stadt |
| Switzerland/BS-UHB-42413789/2020 | EPI_ISL_581989 | 8/27/2020 | Europe | Switzerland | Basel-Stadt |
| Switzerland/BS-UHB-42419052/2020 | EPI_ISL_581998 | 8/31/2020 | Europe | Switzerland | Basel-Stadt |
| Switzerland/SG-CLM-04289106/2021 | EPI_ISL_1939271 | 4/28/2021 | Europe | Switzerland | Sankt Gallen |
| Switzerland/SG-CLM-04289130/2021 | EPI_ISL_1939272 | 4/28/2021 | Europe | Switzerland | Sankt Gallen |
| Switzerland/SG-ETHZ-660181/2021 | EPI_ISL_2724442 | 6/8/2021 | Europe | Switzerland | Sankt Gallen |
| Switzerland/SG-UZH-IMV-3ba4d925/2021 | EPI_ISL_3425887 | 8/5/2021 | Europe | Switzerland | Sankt Gallen |
| Switzerland/SO-ETHZ-521398/2020 | EPI_ISL_1260406 | 11/30/2020 | Europe | Switzerland | Solothurn |
| Switzerland/VD-CHUV-GEN6158/2021 | EPI_ISL_3475930 | 8/4/2021 | Europe | Switzerland | Vaud |
| Switzerland/VS-ETHZ-350373/2020 | EPI_ISL_1130727 | 10/31/2020 | Europe | Switzerland | Valais |
| Switzerland/ZH-ETHZ-33543674/2021 | EPI_ISL_3297398 | 7/21/2021 | Europe | Switzerland | Zvºrich |
| Switzerland/ZH-ETHZ-471117/2021 | EPI_ISL_1131043 | 1/27/2021 | Europe | Switzerland | Zvºrich |
| Switzerland/ZH-ETHZ-490260/2021 | EPI_ISL_1130960 | 2/5/2021 | Europe | Switzerland | Zvºrich |
| Switzerland/ZH-ETHZ-610384/2021 | EPI_ISL_2152208 | 5/4/2021 | Europe | Switzerland | Zvºrich |
| Taiwan/11102/2021 | EPI_ISL_3040152 | 6/5/2021 | Asia | Taiwan | Tainan |
| Taiwan/13435/2021 | EPI_ISL_3040140 | 6/24/2021 | Asia | Taiwan | Taipei City |
| Taiwan/2/2020 | EPI_ISL_406031 | 1/23/2020 | Asia | Taiwan | Taiwan |
| Taiwan/CGMH-CGU-01/2020 | EPI_ISL_411915 | 1/25/2020 | Asia | Taiwan | Taiwan |
| Taiwan/CGMH-CGU-50/2021 | EPI_ISL_956331 | 1/26/2021 | Asia | Taiwan | Taiwan |
| Taiwan/CGMH-CGU-67/2021 | EPI_ISL_2544703 | 5/20/2021 | Asia | Taiwan | Taiwan |

| | | | | | |
|---|---|---|---|---|---|
| Taiwan/TSGH-34/2020 | EPI_ISL_447593 | 4/1/2020 | Asia | Taiwan | New Taipei City |
| Taiwan/TSGH-39/2021 | EPI_ISL_2693004 | 4/25/2021 | Asia | Taiwan | Taiwan |
| Taiwan/TSGH-41/2021 | EPI_ISL_2693000 | 4/26/2021 | Asia | Taiwan | Taiwan |
| Taiwan/TSGH-43/2021 | EPI_ISL_2693006 | 5/7/2021 | Asia | Taiwan | Taiwan |
| Thailand/Bangkok-0088/2020 | EPI_ISL_447028 | 4/7/2020 | Asia | Thailand | Bangkok |
| Thailand/Bangkok-CONI-0692/2021 | EPI_ISL_2104730 | 4/20/2021 | Asia | Thailand | Bangkok |
| Thailand/Bangkok-CONI-0718/2021 | EPI_ISL_2104755 | 4/20/2021 | Asia | Thailand | Bangkok |
| Thailand/Bangkok-CONI-0881/2021 | EPI_ISL_2928534 | 6/29/2021 | Asia | Thailand | Bangkok |
| Thailand/DMSc-00393/2021 | EPI_ISL_2433295 | 5/15/2021 | Asia | Thailand | Bangkok |
| Thailand/DMSc-00647/2021 | EPI_ISL_2433287 | 5/23/2021 | Asia | Thailand | Bangkok |
| Thailand/PathumThani-CONI-0909/2021 | EPI_ISL_3152913 | 7/15/2021 | Asia | Thailand | Pathum Thani |
| Thailand/PathumThani-CONI-0918/2021 | EPI_ISL_3152922 | 7/14/2021 | Asia | Thailand | Pathum Thani |
| Thailand/SI202872-NT/2020 | EPI_ISL_437613 | 2/25/2020 | Asia | Thailand | Thailand |
| Thailand/SQ-CONI-0678/2021 | EPI_ISL_1538420 | 4/5/2021 | Asia | Thailand | Bangkok |
| Thailand/Tak-CONI-0860/2021 | EPI_ISL_2928514 | 6/15/2021 | Asia | Thailand | Tak |
| Togo/BMC-19125-20/2020 | EPI_ISL_1434443 | 6/4/2020 | Africa | Togo | Golfe |
| Togo/C5894/2021 | EPI_ISL_1508821 | 1/25/2021 | Africa | Togo | Golfe |
| TrinidadandTobago/1116/2020 | EPI_ISL_2230686 | 12/1/2020 | South America | Trinidad and Tobago | Trinidad and Tobago |
| TrinidadandTobago/37679/2020 | EPI_ISL_1490228 | 8/22/2020 | South America | Trinidad and Tobago | Trinidad and Tobago |
| TrinidadandTobago/55455/2021 | EPI_ISL_1591265 | 2/18/2021 | South America | Trinidad and Tobago | Trinidad and Tobago |
| TrinidadandTobago/60341/2021 | EPI_ISL_1588904 | 3/23/2021 | South America | Trinidad and Tobago | Trinidad and Tobago |
| TrinidadandTobago/65256/2021 | EPI_ISL_2649750 | 4/27/2021 | South America | Trinidad and Tobago | Trinidad and Tobago |
| TrinidadandTobago/65335/2021 | EPI_ISL_2631428 | 4/28/2021 | South America | Trinidad and Tobago | Trinidad and Tobago |
| TrinidadandTobago/71674/2021 | EPI_ISL_2756571 | 5/28/2021 | South America | Trinidad and Tobago | Trinidad and Tobago |

| | | | | | |
|---|---|---|---|---|---|
| TrinidadandTobago/71690/2021 | EPI_ISL_2756569 | 5/28/2021 | South America | Trinidad and Tobago | Trinidad and Tobago |
| TrinidadandTobago/72507/2021 | EPI_ISL_2756579 | 6/4/2021 | South America | Trinidad and Tobago | Trinidad and Tobago |
| TrinidadandTobago/72516/2021 | EPI_ISL_2756580 | 6/3/2021 | South America | Trinidad and Tobago | Trinidad and Tobago |
| TrinidadandTobago/TT44048/2020 | EPI_ISL_756311 | 11/18/2020 | South America | Trinidad and Tobago | Trinidad and Tobago |
| TrinidadandTobago/TT44092/2020 | EPI_ISL_756357 | 11/26/2020 | South America | Trinidad and Tobago | Trinidad and Tobago |
| Tunisia/11-MHT_12/2020 | EPI_ISL_855569 | 9/14/2020 | Africa | Tunisia | Tunis |
| Tunisia/19695/2020 | EPI_ISL_733500 | 7/12/2020 | Africa | Tunisia | Ben Arous |
| Tunisia/202129459/2021 | EPI_ISL_2154331 | 4/26/2021 | Africa | Tunisia | Gafsa |
| Tunisia/29100/2021 | EPI_ISL_2153433 | 4/24/2021 | Africa | Tunisia | Nabeul |
| Tunisia/36107/2021 | EPI_ISL_2896981 | 5/28/2021 | Africa | Tunisia | Tunis |
| Tunisia/9066/2020 | EPI_ISL_1116468 | 8/22/2020 | Africa | Tunisia | Sfax |
| Tunisia/SP-0089/2021 | EPI_ISL_2035753 | 1/29/2021 | Africa | Tunisia | Tunis |
| Turkey/20-Ankara-GUMV-50247/2020 | EPI_ISL_984745 | 11/19/2020 | Europe | Turkey | Ankara |
| Turkey/32-Ankara-GUMV-50630/2020 | EPI_ISL_984749 | 11/20/2020 | Europe | Turkey | Ankara |
| Turkey/CTF_01042021_M044/2021 | EPI_ISL_1662234 | 4/1/2021 | Europe | Turkey | Istanbul |
| Turkey/HSGM-200208/2020 | EPI_ISL_814077 | 4/24/2020 | Europe | Turkey | Turkey |
| Turkey/HSGM-7032/2021 | EPI_ISL_1533681 | 3/1/2021 | Europe | Turkey | Turkey |
| Turkey/HSGM-B11420/2021 | EPI_ISL_2158066 | 4/28/2021 | Europe | Turkey | Turkey |
| Turkey/HSGM-B11564/2021 | EPI_ISL_2158052 | 4/28/2021 | Europe | Turkey | Turkey |
| Turkey/HSGM-B13638/2021 | EPI_ISL_2403370 | 5/24/2021 | Europe | Turkey | Turkey |
| Turkey/HSGM-B14493/2021 | EPI_ISL_3066532 | 5/25/2021 | Europe | Turkey | Turkey |
| Turkey/HSGM-B18114/2021 | EPI_ISL_3032633 | 6/22/2021 | Europe | Turkey | Turkey |
| Turkey/HSGM-B19209/2021 | EPI_ISL_2987923 | 6/29/2021 | Europe | Turkey | Turkey |
| Turkey/HSGM-FS568/2021 | EPI_ISL_3402385 | 8/5/2021 | Europe | Turkey | Turkey |
| Turkey/HSGM-FS68/2021 | EPI_ISL_3402380 | 8/5/2021 | Europe | Turkey | Turkey |
| Turkey/HSGM-GA1/2021 | EPI_ISL_1073761 | 1/27/2021 | Europe | Turkey | Turkey |

| | | | | | |
|---|---|---|---|---|---|
| Turkey/HSGM-GF1865/2021 | EPI_ISL_3256534 | 7/15/2021 | Europe | Turkey | Turkey |
| Turkey/HSGM-GF7497/2021 | EPI_ISL_3306606 | 7/22/2021 | Europe | Turkey | Turkey |
| Uganda/UG053/2020 | EPI_ISL_737970 | 5/11/2020 | Africa | Uganda | Uganda |
| Uganda/UG056/2020 | EPI_ISL_737973 | 7/16/2020 | Africa | Uganda | Uganda |
| Uganda/UG087/2020 | EPI_ISL_737998 | 8/25/2020 | Africa | Uganda | Uganda |
| Uganda/UG226/2021 | EPI_ISL_1469324 | 1/20/2021 | Africa | Uganda | Uganda |
| Uganda/UG429/2021 | EPI_ISL_2346410 | 2/15/2021 | Africa | Uganda | Uganda |
| Uganda/UG461/2021 | EPI_ISL_2346427 | 4/17/2021 | Africa | Uganda | Uganda |
| Uganda/UG486/2021 | EPI_ISL_2690451 | 5/9/2021 | Africa | Uganda | Uganda |
| Uganda/UG489/2021 | EPI_ISL_2690459 | 4/20/2021 | Africa | Uganda | Uganda |
| Uganda/UG491/2021 | EPI_ISL_2690461 | 5/9/2021 | Africa | Uganda | Uganda |
| Uganda/UG511/2021 | EPI_ISL_3149345 | 6/4/2021 | Africa | Uganda | Uganda |
| Uganda/UG541/2021 | EPI_ISL_2690501 | 6/15/2021 | Africa | Uganda | Uganda |
| Uganda/UG619/2021 | EPI_ISL_3149360 | 7/2/2021 | Africa | Uganda | Uganda |
| Uganda/UG621/2021 | EPI_ISL_3149362 | 7/1/2021 | Africa | Uganda | Uganda |
| Ukraine/Dnipro-35/2021 | EPI_ISL_1315427 | 2/18/2021 | Europe | Ukraine | Dnipro |
| Ukraine/G1-00/2021 | EPI_ISL_2934574 | 6/18/2021 | Europe | Ukraine | Kyiv |
| Ukraine/HZ2-59758/2021 | EPI_ISL_2934570 | 5/21/2021 | Europe | Ukraine | Kherson |
| Ukraine/Kharkiv-782/2020 | EPI_ISL_582513 | 7/31/2020 | Europe | Ukraine | Kharkiv |
| Ukraine/Kyiv/F2-6/2021 | EPI_ISL_2966511 | 6/15/2021 | Europe | Ukraine | Kyiv |
| UnitedArabEmirates/0041/2020 | EPI_ISL_698250 | 5/17/2020 | Asia | United Arab Emirates | Abu Dhabi |
| UnitedArabEmirates/0168/2020 | EPI_ISL_698355 | 5/18/2020 | Asia | United Arab Emirates | Abu Dhabi |
| UnitedArabEmirates/0329/2020 | EPI_ISL_698505 | 5/18/2020 | Asia | United Arab Emirates | Abu Dhabi |
| UnitedArabEmirates/0382/2020 | EPI_ISL_698557 | 5/15/2020 | Asia | United Arab Emirates | Abu Dhabi |
| UnitedArabEmirates/4289/2020 | EPI_ISL_860057 | 12/27/2020 | Asia | United Arab Emirates | Abu Dhabi |
| UnitedArabEmirates/AZ-USAFSAM-S3096/2021 | EPI_ISL_2661466 | 4/27/2021 | Asia | United Arab Emirates | Abu Dhabi |
| UnitedArabEmirates/AZ-USAFSAM-S4144/2021 | EPI_ISL_3048168 | 6/23/2021 | Asia | United Arab Emirates | Abu Dhabi |

| | | | | | |
|---|---|---|---|---|---|
| UnitedArabEmirates/AZ-USAFSAM-S4149/2021 | EPI_ISL_3048171 | 6/23/2021 | Asia | United Arab Emirates | Abu Dhabi |
| UnitedArabEmirates/L0184/2020 | EPI_ISL_435121 | 2/25/2020 | Asia | United Arab Emirates | United Arab Emirates |
| UnitedArabEmirates/P1/2020 | EPI_ISL_463740 | 3/30/2020 | Asia | United Arab Emirates | United Arab Emirates |
| UnitedArabEmirates/skmc-920168117/2020 | EPI_ISL_582125 | 1/28/2020 | Asia | United Arab Emirates | United Arab Emirates |
| Uruguay/CHY-M479/2020 | EPI_ISL_2754203 | 12/14/2020 | South America | Uruguay | Rocha |
| Uruguay/CUY6-001069-UYSJ/2021 | EPI_ISL_2964655 | 4/19/2021 | South America | Uruguay | Departamento San Jose |
| Uruguay/CUY6-001290-UYSJ/2021 | EPI_ISL_2964682 | 4/18/2021 | South America | Uruguay | Departamento San Jose |
| Uruguay/RIV-M72/2020 | EPI_ISL_750163 | 6/5/2020 | South America | Uruguay | Rivera |
| Uruguay/RT01-UYMO/2020 | EPI_ISL_2965583 | 8/7/2020 | South America | Uruguay | Montevideo |
| Uruguay/RT59-UYMO/2020 | EPI_ISL_2965569 | 12/30/2020 | South America | Uruguay | Montevideo |
| Uruguay/UY-650/2020 | EPI_ISL_480338 | 5/25/2020 | South America | Uruguay | Rivera |
| Uruguay/UY-NYULH2033/2021 | EPI_ISL_2427611 | 2/27/2021 | South America | Uruguay | Montevideo |
| Uruguay/UY-NYUMC866/2020 | EPI_ISL_457962 | 3/30/2020 | South America | Uruguay | Montevideo |
| Uruguay/UY-NYUMC868/2020 | EPI_ISL_457964 | 4/1/2020 | South America | Uruguay | Montevideo |
| USA/AK-CDC-2-3845829/2021 | EPI_ISL_1094319 | 1/20/2021 | North America | USA | Alaska |
| USA/AK-PHL10134/2021 | EPI_ISL_3118596 | 4/26/2021 | North America | USA | Alaska |
| USA/AK-PHL10745/2021 | EPI_ISL_3270455 | 7/22/2021 | North America | USA | Alaska |
| USA/AK-PHL384/2020 | EPI_ISL_576111 | 6/1/2020 | North America | USA | Alaska |
| USA/AK-PHL6542/2020 | EPI_ISL_884216 | 12/1/2020 | North America | USA | Alaska |
| USA/AK-PHL6616/2020 | EPI_ISL_911694 | 12/13/2020 | North America | USA | Alaska |
| USA/AK-PHL8236/2021 | EPI_ISL_1789665 | 4/19/2021 | North America | USA | Alaska |

| | | | | | |
|---|---|---|---|---|---|
| USA/AK-PHL8694/2021 | EPI_ISL_2136439 | 5/6/2021 | North America | USA | Alaska |
| USA/AK-PHL8752/2021 | EPI_ISL_2154068 | 5/10/2021 | North America | USA | Alaska |
| USA/AK-PHL9342/2021 | EPI_ISL_2503978 | 6/1/2021 | North America | USA | Alaska |
| USA/AK-PHL9667/2021 | EPI_ISL_2790767 | 6/18/2021 | North America | USA | Alaska |
| USA/AK-PHL9886/2021 | EPI_ISL_2969589 | 7/2/2021 | North America | USA | Alaska |
| USA/AL-ADPH-ADPH_0137/2021 | EPI_ISL_3432475 | 8/2/2021 | North America | USA | Alabama |
| USA/AL-ADPH-ADPH_0140/2021 | EPI_ISL_3432476 | 8/2/2021 | North America | USA | Alabama |
| USA/AL-Baldwin-SL-21051360014/2021 | EPI_ISL_2958926 | 5/13/2021 | North America | USA | Alabama |
| USA/AL-CDC-2-4356444/2021 | EPI_ISL_2229281 | 4/27/2021 | North America | USA | Alabama |
| USA/AL-CDC-ASC210033401/2021 | EPI_ISL_2150763 | 4/26/2021 | North America | USA | Alabama |
| USA/AL-CDC-FG-035811/2021 | EPI_ISL_2525574 | 5/20/2021 | North America | USA | Alabama |
| USA/AL-CDC-FG-061972/2021 | EPI_ISL_3352371 | 7/29/2021 | North America | USA | Alabama |
| USA/AL-CDC-LC0020931/2021 | EPI_ISL_1222252 | 2/24/2021 | North America | USA | Alabama |
| USA/AL-CDC-LC0021197/2021 | EPI_ISL_1222419 | 3/1/2021 | North America | USA | Alabama |
| USA/AL-HGSC-JHJG/2021 | EPI_ISL_979474 | 1/25/2021 | North America | USA | Alabama |
| USA/AL-QDX-244/2020 | EPI_ISL_498695 | 4/28/2020 | North America | USA | Alabama |
| USA/AL-UAB-GX377/2021 | EPI_ISL_3050176 | 6/21/2021 | North America | USA | Alabama |
| USA/AL-UAB-GX380/2021 | EPI_ISL_3050179 | 6/24/2021 | North America | USA | Alabama |
| USA/AL-UAB-GX420/2021 | EPI_ISL_3417495 | 7/8/2021 | North America | USA | Alabama |
| USA/AL-USAFSAM-S268/2020 | EPI_ISL_812598 | 8/11/2020 | North America | USA | Alabama |

| | | | | | |
|---|---|---|---|---|---|
| USA/AL-USAFSAM-S295/2020 | EPI_ISL_812613 | 8/17/2020 | North America | USA | Alabama |
| USA/AR-CDC-4130844-001/2021 | EPI_ISL_2422437 | 5/6/2021 | North America | USA | Arkansas |
| USA/AR-CDC-4195/2020 | EPI_ISL_509678 | 7/9/2020 | North America | USA | Arkansas |
| USA/AR-CDC-9KXZ-8440/2020 | EPI_ISL_906848 | 12/28/2020 | North America | USA | Arkansas |
| USA/AR-CDC-ASC210035789/2021 | EPI_ISL_2043419 | 4/28/2021 | North America | USA | Arkansas |
| USA/AR-CDC-ASC210106884/2021 | EPI_ISL_2528356 | 6/3/2021 | North America | USA | Arkansas |
| USA/AR-CDC-QDX27249109/2021 | EPI_ISL_3396263 | 7/27/2021 | North America | USA | Arkansas |
| USA/AR-QDX-680/2020 | EPI_ISL_571409 | 3/18/2020 | North America | USA | Arkansas |
| USA/AR-UMGC-7561/2021 | EPI_ISL_3474026 | 7/28/2021 | North America | USA | Arkansas |
| USA/AR-UMGC-7727/2021 | EPI_ISL_3474061 | 8/2/2021 | North America | USA | Arkansas |
| USA/AR-UMGC-7963/2021 | EPI_ISL_3473983 | 8/2/2021 | North America | USA | Arkansas |
| USA/AR-UNM-BAP249456/2021 | EPI_ISL_3246867 | 5/22/2021 | North America | USA | Arkansas |
| USA/AR-UNM-BAPX250165/2021 | EPI_ISL_3274244 | 6/27/2021 | North America | USA | Arkansas |
| USA/AR-UNM-V00168/2021 | EPI_ISL_2484734 | 4/27/2021 | North America | USA | Arkansas |
| USA/AR-USAFSAM-S100/2020 | EPI_ISL_812308 | 3/19/2020 | North America | USA | Arkansas |
| USA/AZ-ASPHL-2727/2020 | EPI_ISL_1184569 | 10/26/2020 | North America | USA | Arizona |
| USA/AZ-ASU10287/2021 | EPI_ISL_3385657 | 8/3/2021 | North America | USA | Arizona |
| USA/AZ-ASU10291/2021 | EPI_ISL_3385528 | 8/2/2021 | North America | USA | Arizona |
| USA/AZ-CDC-LC0102431/2021 | EPI_ISL_3326784 | 7/12/2021 | North America | USA | Arizona |
| USA/AZ-CDC-QDX22554295/2021 | EPI_ISL_1291126 | 2/26/2021 | North America | USA | Arizona |

| | | | | | |
|---|---|---|---|---|---|
| USA/AZ-CDC-QDX24394333/2021 | EPI_ISL_2133764 | 4/26/2021 | North America | USA | Arizona |
| USA/AZ-CDC-QDX26344504/2021 | EPI_ISL_2929660 | 6/27/2021 | North America | USA | Arizona |
| USA/AZ-CDC-STM-000060822/2021 | EPI_ISL_2010057 | 4/22/2021 | North America | USA | Arizona |
| USA/AZ-TG280825/2020 | EPI_ISL_694171 | 4/21/2020 | North America | USA | Arizona |
| USA/AZ-TG476754/2020 | EPI_ISL_914502 | 7/7/2020 | North America | USA | Arizona |
| USA/AZ-TG666157/2020 | EPI_ISL_696238 | 5/2/2020 | North America | USA | Arizona |
| USA/AZ-TG717742/2021 | EPI_ISL_1036558 | 2/2/2021 | North America | USA | Arizona |
| USA/AZ-TG779744/2020 | EPI_ISL_1465097 | 11/6/2020 | North America | USA | Arizona |
| USA/AZ-TG832795/2021 | EPI_ISL_1909579 | 4/8/2021 | North America | USA | Arizona |
| USA/AZ-TG845984/2020 | EPI_ISL_2000922 | 11/5/2020 | North America | USA | Arizona |
| USA/AZ-TG909634/2021 | EPI_ISL_2491222 | 5/12/2021 | North America | USA | Arizona |
| USA/AZ-TG910121/2021 | EPI_ISL_2490991 | 5/18/2021 | North America | USA | Arizona |
| USA/AZ-TG935237/2020 | EPI_ISL_2769285 | 10/21/2020 | North America | USA | Arizona |
| USA/AZ-TG979194/2021 | EPI_ISL_3326835 | 6/24/2021 | North America | USA | Arizona |
| USA/AZ-TG989851/2021 | EPI_ISL_3460374 | 7/29/2021 | North America | USA | Arizona |
| USA/CA-ALSR-1430/2020 | EPI_ISL_494597 | 3/28/2020 | North America | USA | California |
| USA/CA-ALSR-7871/2021 | EPI_ISL_1366469 | 2/22/2021 | North America | USA | California |
| USA/CA-CDC-4097534-001/2020 | EPI_ISL_2101838 | 6/19/2020 | North America | USA | California |
| USA/CA-CDC-ASC210031917/2021 | EPI_ISL_2089959 | 4/22/2021 | North America | USA | California |
| USA/CA-CDC-FG-046228/2021 | EPI_ISL_3211576 | 7/20/2021 | North America | USA | California |

| | | | | | |
|---|---|---|---|---|---|
| USA/CA-CDC-FG-052442/2021 | EPI_ISL_3303432 | 7/28/2021 | North America | USA | California |
| USA/CA-CDC-FG-059582/2021 | EPI_ISL_3350701 | 8/3/2021 | North America | USA | California |
| USA/CA-CDC-FG-060550/2021 | EPI_ISL_3351287 | 8/3/2021 | North America | USA | California |
| USA/CA-CDC-LC0068268/2021 | EPI_ISL_2482167 | 5/28/2021 | North America | USA | California |
| USA/CA-CDPH-3000007591/2021 | EPI_ISL_2837944 | 4/8/2021 | North America | USA | California |
| USA/CA-CDPH-3000046154/2021 | EPI_ISL_2923608 | 4/27/2021 | North America | USA | California |
| USA/CA-CDPH-UC1/2020 | EPI_ISL_413557 | 2/26/2020 | North America | USA | Grand Princess |
| USA/CA-CDPH-UC11/2020 | EPI_ISL_413931 | 3/5/2020 | North America | USA | Grand Princess |
| USA/CA-CDPH-UC9/2020 | EPI_ISL_413928 | 3/5/2020 | North America | USA | Grand Princess |
| USA/CA-Curative-041773/2021 | EPI_ISL_2839804 | 6/25/2021 | North America | USA | California |
| USA/CA-CZB-10911/2020 | EPI_ISL_583140 | 8/27/2020 | North America | USA | California |
| USA/CA-CZB-13483/2020 | EPI_ISL_672046 | 10/29/2020 | North America | USA | California |
| USA/CA-LACPHL-AF01042/2021 | EPI_ISL_2391753 | 5/3/2021 | North America | USA | California |
| USA/CA-LACPHL-AF01467/2021 | EPI_ISL_2907575 | 6/25/2021 | North America | USA | California |
| USA/CA-SR0130/2020 | EPI_ISL_445114 | 3/28/2020 | North America | USA | California |
| USA/CO-CDC-ASC210032776/2021 | EPI_ISL_2090162 | 4/24/2021 | North America | USA | Colorado |
| USA/CO-CDC-FG-058346/2021 | EPI_ISL_3349220 | 8/2/2021 | North America | USA | Colorado |
| USA/CO-CDC-FG-062302/2021 | EPI_ISL_3397268 | 8/2/2021 | North America | USA | Colorado |
| USA/CO-CDC-LC0109361/2021 | EPI_ISL_3328566 | 7/7/2021 | North America | USA | Colorado |
| USA/CO-CDC-MMB08617707/2021 | EPI_ISL_2382126 | 5/3/2021 | North America | USA | Colorado |

| | | | | | |
|---|---|---|---|---|---|
| USA/CO-CDC-MMB08819580/2021 | EPI_ISL_2451038 | 5/15/2021 | North America | USA | Colorado |
| USA/CO-CDC-MMB09017080/2021 | EPI_ISL_2786017 | 6/1/2021 | North America | USA | Colorado |
| USA/CO-CDC-MMB09218058/2021 | EPI_ISL_2875423 | 6/22/2021 | North America | USA | Colorado |
| USA/CO-CDC-MMB09374559/2021 | EPI_ISL_3427261 | 7/14/2021 | North America | USA | Colorado |
| USA/CO-CDPHE-2004230658/2020 | EPI_ISL_677637 | 4/23/2020 | North America | USA | Colorado |
| USA/CO-CDPHE-2006050911/2020 | EPI_ISL_710266 | 6/4/2020 | North America | USA | Colorado |
| USA/CO-CDPHE-2009164052/2020 | EPI_ISL_710236 | 9/16/2020 | North America | USA | Colorado |
| USA/CO-CDPHE-2100111864/2020 | EPI_ISL_2310815 | 12/14/2020 | North America | USA | Colorado |
| USA/CO-CDPHE-2100247522/2021 | EPI_ISL_1038882 | 1/20/2021 | North America | USA | Colorado |
| USA/CO-CDPHE-2100529475/2021 | EPI_ISL_1234283 | 2/25/2021 | North America | USA | Colorado |
| USA/CO-CDPHE-2100993844/2021 | EPI_ISL_2309659 | 4/30/2021 | North America | USA | Colorado |
| USA/CO-CDPHE-2101210205/2021 | EPI_ISL_3160014 | 3/10/2021 | North America | USA | Colorado |
| USA/CT-CDC-ASC210110020/2021 | EPI_ISL_2784893 | 6/17/2021 | North America | USA | Connecticut |
| USA/CT-CDC-QDX23175097/2021 | EPI_ISL_1552394 | 3/21/2021 | North America | USA | Connecticut |
| USA/CT-CDC-QDX24365603/2021 | EPI_ISL_2133395 | 4/26/2021 | North America | USA | Connecticut |
| USA/CT-CDC-QDX25025302/2021 | EPI_ISL_2397597 | 5/10/2021 | North America | USA | Connecticut |
| USA/CT-CDC-QDX25472483/2021 | EPI_ISL_2529318 | 6/1/2021 | North America | USA | Connecticut |
| USA/CT-CDC-QDX27053335/2021 | EPI_ISL_3395649 | 7/18/2021 | North America | USA | Connecticut |
| USA/CT-CDCBI-CRSP_GS3KF2LXCISEOTL4/2021 | EPI_ISL_3408357 | 8/6/2021 | North America | USA | Connecticut |
| USA/CT-CDCBI-CRSP_R2K5UGASXXJ6EO72/2021 | EPI_ISL_3408111 | 8/5/2021 | North America | USA | Connecticut |

| | | | | | |
|---|---|---|---|---|---|
| USA/CT-Yale-1173/2020 | EPI_ISL_1017527 | 12/24/2020 | North America | USA | Connecticut |
| USA/CT-Yale-129/2020 | EPI_ISL_431092 | 4/13/2020 | North America | USA | Connecticut |
| USA/CT-Yale-1417/2021 | EPI_ISL_1091807 | 1/25/2021 | North America | USA | Connecticut |
| USA/CT-Yale-4223/2021 | EPI_ISL_2020702 | 4/20/2021 | North America | USA | Connecticut |
| USA/CT-Yale-4644/2021 | EPI_ISL_2159102 | 5/1/2021 | North America | USA | Connecticut |
| USA/CT-Yale-480/2020 | EPI_ISL_729765 | 9/18/2020 | North America | USA | Connecticut |
| USA/CT-Yale-620/2020 | EPI_ISL_730078 | 5/11/2020 | North America | USA | Connecticut |
| USA/CT-Yale-6772/2021 | EPI_ISL_3370156 | 7/21/2021 | North America | USA | Connecticut |
| USA/DC-CDC-2-4393173/2021 | EPI_ISL_2383998 | 5/5/2021 | North America | USA | Washington DC |
| USA/DC-CDC-FG-062559/2021 | EPI_ISL_3397627 | 8/4/2021 | North America | USA | Washington DC |
| USA/DC-CDC-LC0100575/2021 | EPI_ISL_3328398 | 7/16/2021 | North America | USA | Washington DC |
| USA/DC-DFS-PHL-0077/2020 | EPI_ISL_804924 | 5/29/2020 | North America | USA | Washington DC |
| USA/DC-DFS-PHL-0079/2020 | EPI_ISL_804863 | 6/26/2020 | North America | USA | Washington DC |
| USA/DC-DFS-PHL-0091/2020 | EPI_ISL_804931 | 7/21/2020 | North America | USA | Washington DC |
| USA/DC-DFS-PHL-0248/2020 | EPI_ISL_1295621 | 10/13/2020 | North America | USA | Washington DC |
| USA/DC-DFS-PHL-0539/2020 | EPI_ISL_2176257 | 10/2/2020 | North America | USA | Washington DC |
| USA/DC-DFS-PHL-0728/2021 | EPI_ISL_2854078 | 6/19/2021 | North America | USA | Washington DC |
| USA/DC-DFS-PHL-0755/2021 | EPI_ISL_2854101 | 3/17/2021 | North America | USA | Washington DC |
| USA/DC-DFS-PHL-0818/2021 | EPI_ISL_3010131 | 6/23/2021 | North America | USA | Washington DC |
| USA/DC-DFS-PHL-0889/2021 | EPI_ISL_3031755 | 5/7/2021 | North America | USA | Washington DC |

| | | | | | |
|---|---|---|---|---|---|
| USA/DC-DFS-PHL-0930/2021 | EPI_ISL_3369984 | 7/20/2021 | North America | USA | Washington DC |
| USA/DC-DFS-PHL-0955/2021 | EPI_ISL_3375650 | 4/17/2021 | North America | USA | Washington DC |
| USA/DC-DFS-PHL-0991/2021 | EPI_ISL_3481990 | 4/24/2021 | North America | USA | Washington DC |
| USA/DC-HP00080/2020 | EPI_ISL_438237 | 3/20/2020 | North America | USA | Washington DC |
| USA/DC-HP02379/2021 | EPI_ISL_1036358 | 1/8/2021 | North America | USA | Washington DC |
| USA/DC-HP13141-PIDOLQVETP/2020 | EPI_ISL_2691730 | 9/2/2020 | North America | USA | Washington DC |
| USA/DE-B1089749/2021 | EPI_ISL_2383701 | 5/15/2021 | North America | USA | Delaware |
| USA/DE-CDC-ASC210108586/2021 | EPI_ISL_2687440 | 6/10/2021 | North America | USA | Delaware |
| USA/DE-CDC-IBX166377103323/2021 | EPI_ISL_2323987 | 5/11/2021 | North America | USA | Delaware |
| USA/DE-CDC-LC0095671/2021 | EPI_ISL_3112472 | 7/6/2021 | North America | USA | Delaware |
| USA/DE-Curative-026510/2021 | EPI_ISL_3273251 | 7/9/2021 | North America | USA | Delaware |
| USA/DE-Curative-036915/2021 | EPI_ISL_2784141 | 6/5/2021 | North America | USA | Delaware |
| USA/DE-DHSS-B1084274/2021 | EPI_ISL_2107618 | 4/23/2021 | North America | USA | Delaware |
| USA/DE-DHSS-B1085846/2021 | EPI_ISL_2107768 | 4/25/2021 | North America | USA | Delaware |
| USA/DE-DHSS-F1051814/2021 | EPI_ISL_933764 | 1/16/2021 | North America | USA | Delaware |
| USA/DE-DHSS-F918049/2020 | EPI_ISL_593940 | 3/26/2020 | North America | USA | Delaware |
| USA/DE-DHSS-F970606/2020 | EPI_ISL_693712 | 6/12/2020 | North America | USA | Delaware |
| USA/DE-DHSS-F972766/2020 | EPI_ISL_693707 | 7/7/2020 | North America | USA | Delaware |
| USA/DE-DHSS-F979763/2020 | EPI_ISL_693697 | 7/30/2020 | North America | USA | Delaware |
| USA/FL_5091/2020 | EPI_ISL_419560 | 2/28/2020 | North America | USA | Florida |

| | | | | | |
|---|---|---|---|---|---|
| USA/FL-BPHL-0505/2020 | EPI_ISL_508708 | 6/10/2020 | North America | USA | Florida |
| USA/FL-BPHL-2382/2020 | EPI_ISL_848923 | 10/19/2020 | North America | USA | Florida |
| USA/FL-BPHL-2759/2020 | EPI_ISL_849039 | 9/15/2020 | North America | USA | Florida |
| USA/FL-BPHL-3992/2021 | EPI_ISL_2567083 | 4/23/2021 | North America | USA | Florida |
| USA/FL-CDC-ASC210030197/2021 | EPI_ISL_2149664 | 4/21/2021 | North America | USA | Florida |
| USA/FL-CDC-ASC210158782/2021 | EPI_ISL_3324200 | 7/17/2021 | North America | USA | Florida |
| USA/FL-CDC-FG-011556/2021 | EPI_ISL_1555458 | 3/12/2021 | North America | USA | Florida |
| USA/FL-CDC-KCDN-3707/2020 | EPI_ISL_812142 | 3/9/2020 | North America | USA | Florida |
| USA/FL-CDC-LC0069897/2021 | EPI_ISL_2611189 | 6/2/2021 | North America | USA | Florida |
| USA/FL-CDC-QDX26574713/2021 | EPI_ISL_3212965 | 7/5/2021 | North America | USA | Florida |
| USA/FL-CDC-STM-000073507/2021 | EPI_ISL_2440354 | 5/11/2021 | North America | USA | Florida |
| USA/FL-CDC-STM-125/2020 | EPI_ISL_802694 | 12/29/2020 | North America | USA | Florida |
| USA/FL-Curative-022307/2021 | EPI_ISL_2784033 | 5/18/2021 | North America | USA | Florida |
| USA/FL-TGH-1301/2021 | EPI_ISL_3399791 | 8/3/2021 | North America | USA | Florida |
| USA/FL-TGH-1353/2021 | EPI_ISL_3462366 | 8/8/2021 | North America | USA | Florida |
| USA/FL-UCF_NP_012/2021 | EPI_ISL_3031276 | 6/30/2021 | North America | USA | Florida |
| USA/FL-USAFSAM-S648/2020 | EPI_ISL_831848 | 12/4/2020 | North America | USA | Florida |
| USA/GA-CDC-2-4068562/2021 | EPI_ISL_1094284 | 1/4/2021 | North America | USA | Georgia |
| USA/GA-CDC-2-4106226/2020 | EPI_ISL_1533481 | 7/8/2020 | North America | USA | Georgia |
| USA/GA-CDC-2-4106332/2020 | EPI_ISL_1533550 | 11/17/2020 | North America | USA | Georgia |

| | | | | | |
|---|---|---|---|---|---|
| USA/GA-CDC-4131828-001/2021 | EPI_ISL_2441778 | 4/15/2021 | North America | USA | Georgia |
| USA/GA-CDC-5100/2020 | EPI_ISL_566088 | 6/25/2020 | North America | USA | Georgia |
| USA/GA-CDC-ASC210053585/2021 | EPI_ISL_1995073 | 4/7/2021 | North America | USA | Georgia |
| USA/GA-CDC-GA-EHC-117N/2020 | EPI_ISL_2790705 | 3/30/2020 | North America | USA | Georgia |
| USA/GA-CDC-LC0001032/2020 | EPI_ISL_1029390 | 12/26/2020 | North America | USA | Georgia |
| USA/GA-CDC-LC0065649/2021 | EPI_ISL_2480526 | 5/30/2021 | North America | USA | Georgia |
| USA/GA-CDC-LC0069905/2021 | EPI_ISL_2611203 | 6/3/2021 | North America | USA | Georgia |
| USA/GA-CDC-LC0099903/2021 | EPI_ISL_3329754 | 7/14/2021 | North America | USA | Georgia |
| USA/GA-CDC-MMB08883843/2021 | EPI_ISL_2647376 | 5/20/2021 | North America | USA | Georgia |
| USA/GA-CDC-MMB09275650/2021 | EPI_ISL_3065316 | 6/30/2021 | North America | USA | Georgia |
| USA/GA-CDC-MMB09398998/2021 | EPI_ISL_3427728 | 7/16/2021 | North America | USA | Georgia |
| USA/GA-CDC-STM-000060542/2021 | EPI_ISL_1990762 | 4/21/2021 | North America | USA | Georgia |
| USA/GA-EHC-1387K/2021 | EPI_ISL_2346151 | 4/30/2021 | North America | USA | Georgia |
| USA/GA-EHC-230X/2020 | EPI_ISL_1278064 | 5/1/2020 | North America | USA | Georgia |
| USA/GA-GPHL-0984/2021 | EPI_ISL_3435254 | 8/2/2021 | North America | USA | Georgia |
| USA/GA-GPHL-1002/2021 | EPI_ISL_3435093 | 8/2/2021 | North America | USA | Georgia |
| USA/GA-QDX-239/2020 | EPI_ISL_498716 | 4/27/2020 | North America | USA | Georgia |
| USA/GA-QDX-3463/2020 | EPI_ISL_876946 | 8/9/2020 | North America | USA | Georgia |
| USA/GA-USAFSAM-S508/2020 | EPI_ISL_831696 | 11/13/2020 | North America | USA | Georgia |
| USA/GU-CDC-2-3847017/2020 | EPI_ISL_3353743 | 11/27/2020 | North America | USA | Guam |

| | | | | | |
|---|---|---|---|---|---|
| USA/GU-CDC-2-4242957/2021 | EPI_ISL_1823583 | 4/5/2021 | North America | USA | Guam |
| USA/GU-CDC-2-4242998/2021 | EPI_ISL_1823588 | 4/5/2021 | North America | USA | Guam |
| USA/GU-CDC-2-4243086/2021 | EPI_ISL_1823587 | 4/11/2021 | North America | USA | Guam |
| USA/GU-CDC-2-4356427/2021 | EPI_ISL_2229096 | 4/21/2021 | North America | USA | Guam |
| USA/GU-CDC-2-4356439/2021 | EPI_ISL_2229087 | 4/20/2021 | North America | USA | Guam |
| USA/GU-CDC-2-4448976/2021 | EPI_ISL_2451659 | 5/4/2021 | North America | USA | Guam |
| USA/GU-CDC-2-4524556/2021 | EPI_ISL_2787627 | 5/24/2021 | North America | USA | Guam |
| USA/GU-CDC-2-4611186/2021 | EPI_ISL_3355185 | 6/8/2021 | North America | USA | Guam |
| USA/GU-CDC-2-4611269/2021 | EPI_ISL_3355210 | 6/14/2021 | North America | USA | Guam |
| USA/GU-CDC-2-4651356/2021 | EPI_ISL_3242000 | 7/2/2021 | North America | USA | Guam |
| USA/GU-CDC-2-4651485/2021 | EPI_ISL_3241994 | 7/6/2021 | North America | USA | Guam |
| USA/HI-CDC-2-3975978/2021 | EPI_ISL_1272649 | 2/3/2021 | North America | USA | Hawaii |
| USA/HI-CDC-ASC210108750/2021 | EPI_ISL_2687542 | 6/10/2021 | North America | USA | Hawaii |
| USA/HI-H200030/2020 | EPI_ISL_752616 | 3/19/2020 | North America | USA | Hawaii |
| USA/HI-H200046/2020 | EPI_ISL_752631 | 4/16/2020 | North America | USA | Hawaii |
| USA/HI-H200187/2020 | EPI_ISL_752746 | 7/31/2020 | North America | USA | Hawaii |
| USA/HI-H200476/2020 | EPI_ISL_753019 | 10/23/2020 | North America | USA | Hawaii |
| USA/HI-H200569/2020 | EPI_ISL_753096 | 10/24/2020 | North America | USA | Hawaii |
| USA/HI-H210920/2021 | EPI_ISL_967703 | 1/12/2021 | North America | USA | Hawaii |
| USA/HI-H211228/2021 | EPI_ISL_1292644 | 2/19/2021 | North America | USA | Hawaii |

| | | | | | |
|---|---|---|---|---|---|
| USA/HI-H211278/2021 | EPI_ISL_1292692 | 2/25/2021 | North America | USA | Hawaii |
| USA/HI-H211324/2021 | EPI_ISL_1292738 | 2/26/2021 | North America | USA | Hawaii |
| USA/HI-H211382/2021 | EPI_ISL_1292795 | 2/28/2021 | North America | USA | Hawaii |
| USA/HI-H211867/2021 | EPI_ISL_1771414 | 4/7/2021 | North America | USA | Hawaii |
| USA/HI-H211966/2021 | EPI_ISL_2007872 | 4/16/2021 | North America | USA | Hawaii |
| USA/HI-H212020/2021 | EPI_ISL_2007904 | 4/17/2021 | North America | USA | Hawaii |
| USA/HI-H212356/2021 | EPI_ISL_2724626 | 5/7/2021 | North America | USA | Hawaii |
| USA/HI-H212436/2021 | EPI_ISL_2724675 | 5/19/2021 | North America | USA | Hawaii |
| USA/HI-H212814/2021 | EPI_ISL_3033184 | 6/17/2021 | North America | USA | Hawaii |
| USA/HI-H213046/2021 | EPI_ISL_3189693 | 7/13/2021 | North America | USA | Hawaii |
| USA/HI-H213195/2021 | EPI_ISL_3307857 | 7/20/2021 | North America | USA | Hawaii |
| USA/IA_6401/2020 | EPI_ISL_424897 | 3/8/2020 | North America | USA | Iowa |
| USA/IA-CDC-2-4634466/2021 | EPI_ISL_3127990 | 6/30/2021 | North America | USA | Iowa |
| USA/IA-CDC-ASC210030890/2021 | EPI_ISL_2041463 | 4/21/2021 | North America | USA | Iowa |
| USA/IA-CDC-ASC210035003/2021 | EPI_ISL_2043148 | 4/27/2021 | North America | USA | Iowa |
| USA/IA-CDC-ASC210037723/2021 | EPI_ISL_2148482 | 5/1/2021 | North America | USA | Iowa |
| USA/IA-CDC-ASC210068512/2021 | EPI_ISL_1998540 | 4/13/2021 | North America | USA | Iowa |
| USA/IA-CDC-LC0056502/2021 | EPI_ISL_2046795 | 5/3/2021 | North America | USA | Iowa |
| USA/IA-GMF-17036/2020 | EPI_ISL_547735 | 8/24/2020 | North America | USA | Iowa |
| USA/IA-GMF-23647/2020 | EPI_ISL_660879 | 10/2/2020 | North America | USA | Iowa |

| | | | | | |
|---|---|---|---|---|---|
| USA/IA-GMF-B00892/2021 | EPI_ISL_2501015 | 6/3/2021 | North America | USA | Iowa |
| USA/IA-S21WGS2429/2021 | EPI_ISL_2983697 | 7/6/2021 | North America | USA | Iowa |
| USA/IA-SHL-1574135/2021 | EPI_ISL_1261788 | 2/1/2021 | North America | USA | Iowa |
| USA/IA-SHL-1761293/2021 | EPI_ISL_3398024 | 7/17/2021 | North America | USA | Iowa |
| USA/IA-UIHC-MO01/2020 | EPI_ISL_903312 | 6/8/2020 | North America | USA | Iowa |
| USA/IA-UIHC-MO29/2020 | EPI_ISL_1010733 | 5/11/2020 | North America | USA | Iowa |
| USA/IA-UW-662/2020 | EPI_ISL_491307 | 6/27/2020 | North America | USA | Illinois |
| USA/ID-BVAMC-724398/2021 | EPI_ISL_3010539 | 4/20/2021 | North America | USA | Idaho |
| USA/ID-BVAMC-737649/2021 | EPI_ISL_3090841 | 6/24/2021 | North America | USA | Idaho |
| USA/ID-BVAMC-742432/2021 | EPI_ISL_3404627 | 7/23/2021 | North America | USA | Idaho |
| USA/ID-CDC-2-3769290/2020 | EPI_ISL_903603 | 12/22/2020 | North America | USA | Idaho |
| USA/ID-CDC-ASC210044014/2021 | EPI_ISL_1753532 | 4/2/2021 | North America | USA | Idaho |
| USA/ID-CDC-FG-061703/2021 | EPI_ISL_3352155 | 8/2/2021 | North America | USA | Idaho |
| USA/ID-CDC-IBX416404394850/2021 | EPI_ISL_2324160 | 5/10/2021 | North America | USA | Idaho |
| USA/ID-CDC-LC0101210/2021 | EPI_ISL_3326233 | 7/17/2021 | North America | USA | Idaho |
| USA/ID-CDC-MMB09007027/2021 | EPI_ISL_2648001 | 5/18/2021 | North America | USA | Idaho |
| USA/ID-CDC-MMB09280784/2021 | EPI_ISL_3065344 | 6/22/2021 | North America | USA | Idaho |
| USA/ID-IBL-637445/2020 | EPI_ISL_3242950 | 7/30/2020 | North America | USA | Idaho |
| USA/ID-IBL-637508/2020 | EPI_ISL_3242929 | 7/30/2020 | North America | USA | Idaho |
| USA/ID-IBL-641208/2020 | EPI_ISL_3464630 | 8/10/2020 | North America | USA | Idaho |

| | | | | | |
|---|---|---|---|---|---|
| USA/ID-IBL-718911/2021 | EPI_ISL_1510265 | 3/23/2021 | North America | USA | Idaho |
| USA/ID-IBL-720493/2021 | EPI_ISL_1627289 | 3/24/2021 | North America | USA | Idaho |
| USA/ID-IBL-726445/2021 | EPI_ISL_2031630 | 4/27/2021 | North America | USA | Idaho |
| USA/ID-IBL-744608/2021 | EPI_ISL_3385927 | 8/2/2021 | North America | USA | Idaho |
| USA/ID-UNM-IBL_1663/2020 | EPI_ISL_1190732 | 9/4/2020 | North America | USA | Idaho |
| USA/ID-UNM-IBL049/2020 | EPI_ISL_2095694 | 10/21/2020 | North America | USA | Idaho |
| USA/ID-USAFSAM-S528/2020 | EPI_ISL_831809 | 11/17/2020 | North America | USA | Idaho |
| USA/IL-Abbott-7741/2020 | EPI_ISL_1597657 | 12/21/2020 | North America | USA | Illinois |
| USA/IL-C21WGS0164/2021 | EPI_ISL_1663087 | 3/19/2021 | North America | USA | Illinois |
| USA/IL-C21WGS1389/2021 | EPI_ISL_2800722 | 6/3/2021 | North America | USA | Illinois |
| USA/IL-C21WGS1550/2021 | EPI_ISL_3231894 | 6/19/2021 | North America | USA | Illinois |
| USA/IL-CDC-LC0053847/2021 | EPI_ISL_2044469 | 4/27/2021 | North America | USA | Illinois |
| USA/IL-CDC-QDX24633020/2021 | EPI_ISL_2179941 | 5/4/2021 | North America | USA | Illinois |
| USA/IL-CDC-QDX27127568/2021 | EPI_ISL_3396127 | 7/24/2021 | North America | USA | Illinois |
| USA/IL-IDPH-I-000127/2020 | EPI_ISL_848338 | 10/8/2020 | North America | USA | Illinois |
| USA/IL-IDPH-I-000258/2020 | EPI_ISL_848394 | 11/19/2020 | North America | USA | Illinois |
| USA/IL-IDPH-I-000516/2020 | EPI_ISL_962073 | 12/9/2020 | North America | USA | Illinois |
| USA/IL-IDPH-I-000663/2020 | EPI_ISL_848534 | 12/2/2020 | North America | USA | Illinois |
| USA/IL-IDPH-JAC-C-000565/2020 | EPI_ISL_1138456 | 5/1/2020 | North America | USA | Illinois |
| USA/IL-IDPH-KNO-S-0002031/2020 | EPI_ISL_1494283 | 8/29/2020 | North America | USA | Illinois |

| | | | | | |
|---|---|---|---|---|---|
| USA/IL-IDPH-PER-C-001758/2020 | EPI_ISL_1494387 | 7/26/2020 | North America | USA | Illinois |
| USA/IL-IDPH-WIL-I-0001169/2021 | EPI_ISL_1323430 | 1/28/2021 | North America | USA | Illinois |
| USA/IL-NM-0353/2020 | EPI_ISL_626381 | 3/27/2020 | North America | USA | Illinois |
| USA/IL-NM-3024/2020 | EPI_ISL_936609 | 11/6/2020 | North America | USA | Florida |
| USA/IL-RED-MCL-39967483/2021 | EPI_ISL_2319809 | 4/10/2021 | North America | USA | Illinois |
| USA/IL-RED-MCL-41006816/2021 | EPI_ISL_2319941 | 4/30/2021 | North America | USA | Illinois |
| USA/IL-RIPHL_10157_Y/2020 | EPI_ISL_3024697 | 3/27/2020 | North America | USA | Illinois |
| USA/IL-RIPHL_50082_G/2021 | EPI_ISL_2758240 | 5/8/2021 | North America | USA | Illinois |
| USA/IL-S21WGS3153/2021 | EPI_ISL_3371544 | 8/2/2021 | North America | USA | Illinois |
| USA/IL-S21WGS3285/2021 | EPI_ISL_3371611 | 8/3/2021 | North America | USA | Illinois |
| USA/IL-SHL-1763479/2021 | EPI_ISL_3374240 | 7/13/2021 | North America | USA | Illinois |
| USA/IN-CDC-LC0070587/2021 | EPI_ISL_2611603 | 6/3/2021 | North America | USA | Indiana |
| USA/IN-CDC-LC0095579/2021 | EPI_ISL_3112473 | 7/6/2021 | North America | USA | Indiana |
| USA/IN-CDC-STM-000064658/2021 | EPI_ISL_2144592 | 4/27/2021 | North America | USA | Indiana |
| USA/IN-GD-SID-C21172603/2021 | EPI_ISL_2696460 | 5/4/2021 | North America | USA | Indiana |
| USA/IN-GD-SID-C21202045/2021 | EPI_ISL_3461318 | 6/22/2021 | North America | USA | Indiana |
| USA/IN-INSPHL-01120/2021 | EPI_ISL_1994891 | 4/23/2021 | North America | USA | Indiana |
| USA/IN-INSPHL-01830/2021 | EPI_ISL_2439173 | 5/17/2021 | North America | USA | Indiana |
| USA/IN-INSPHL-02795/2021 | EPI_ISL_3007497 | 7/9/2021 | North America | USA | Indiana |
| USA/IN-INSPHL-06687/2021 | EPI_ISL_3461024 | 8/3/2021 | North America | USA | Indiana |

| | | | | | |
|---|---|---|---|---|---|
| USA/IN-INSPHL-06901/2021 | EPI_ISL_3461043 | 8/3/2021 | North America | USA | Indiana |
| USA/IN-QDX-182/2020 | EPI_ISL_494518 | 4/28/2020 | North America | USA | Indiana |
| USA/IN-SRL_254602760/2020 | EPI_ISL_1941720 | 8/20/2020 | North America | USA | Indiana |
| USA/IN-UND-00028796/2020 | EPI_ISL_2839513 | 11/18/2020 | North America | USA | Indiana |
| USA/KS-CDC-LC0008637/2021 | EPI_ISL_1032497 | 1/20/2021 | North America | USA | Kansas |
| USA/KS-CDC-QDX21643000/2021 | EPI_ISL_1087515 | 2/2/2021 | North America | USA | Kansas |
| USA/KS-GD-SID-21051631510/2021 | EPI_ISL_2613923 | 5/16/2021 | North America | USA | Kansas |
| USA/KS-KHEL-0001/2020 | EPI_ISL_875675 | 3/6/2020 | North America | USA | Kansas |
| USA/KS-KHEL-0021/2020 | EPI_ISL_883063 | 4/1/2020 | North America | USA | Kansas |
| USA/KS-KHEL-0033/2020 | EPI_ISL_883052 | 4/2/2020 | North America | USA | Kansas |
| USA/KS-KHEL-1014/2021 | EPI_ISL_1700774 | 4/19/2021 | North America | USA | Kansas |
| USA/KS-KHEL-1497/2021 | EPI_ISL_2023312 | 4/27/2021 | North America | USA | Kansas |
| USA/KS-KHEL-1549/2020 | EPI_ISL_2086256 | 7/28/2020 | North America | USA | Kansas |
| USA/KS-KHEL-1795/2021 | EPI_ISL_2227403 | 5/12/2021 | North America | USA | Kansas |
| USA/KS-KHEL-2285/2021 | EPI_ISL_2499943 | 6/7/2021 | North America | USA | Kansas |
| USA/KS-KHEL-2953/2021 | EPI_ISL_2833873 | 6/28/2021 | North America | USA | Kansas |
| USA/KS-KHEL-3527/2021 | EPI_ISL_3021312 | 7/14/2021 | North America | USA | Kansas |
| USA/KS-KHEL-4247/2021 | EPI_ISL_3235541 | 7/30/2021 | North America | USA | Kansas |
| USA/KS-KHEL-4356/2021 | EPI_ISL_3262635 | 8/2/2021 | North America | USA | Kansas |
| USA/KS-KHEL-4719/2021 | EPI_ISL_3462846 | 8/10/2021 | North America | USA | Kansas |

| | | | | | |
|---|---|---|---|---|---|
| USA/KS-KSU-2550/2020 | EPI_ISL_1423981 | 10/26/2020 | North America | USA | Kansas |
| USA/KY-CDC-2-3714580/2020 | EPI_ISL_751670 | 12/3/2020 | North America | USA | Kentucky |
| USA/KY-CDC-ASC210114318/2021 | EPI_ISL_3323141 | 7/13/2021 | North America | USA | Kentucky |
| USA/KY-CDC-LC0021471/2021 | EPI_ISL_1319063 | 2/25/2021 | North America | USA | Kentucky |
| USA/KY-CDC-LC0042771/2021 | EPI_ISL_1683544 | 4/5/2021 | North America | USA | Kentucky |
| USA/KY-CDC-LC0053689/2021 | EPI_ISL_2044392 | 4/27/2021 | North America | USA | Kentucky |
| USA/KY-CDC-LC0097964/2021 | EPI_ISL_3113692 | 7/5/2021 | North America | USA | Kentucky |
| USA/KY-CDC-STM-000004266/2021 | EPI_ISL_966677 | 1/21/2021 | North America | USA | Kentucky |
| USA/KY-GD-051121-21043082059/2021 | EPI_ISL_2602001 | 4/30/2021 | North America | USA | Kentucky |
| USA/KY-GD-051821-21050612007/2021 | EPI_ISL_2602328 | 5/6/2021 | North America | USA | Kentucky |
| USA/KY-GD-051821-21050612756/2021 | EPI_ISL_2602338 | 5/6/2021 | North America | USA | Kentucky |
| USA/KY-GD-SID-21060508203/2021 | EPI_ISL_3461188 | 6/4/2021 | North America | USA | Kentucky |
| USA/KY-GD-SID-21061015662/2021 | EPI_ISL_3461218 | 6/10/2021 | North America | USA | Kentucky |
| USA/KY-KSPHL-100042/2020 | EPI_ISL_966316 | 6/11/2020 | North America | USA | Kentucky |
| USA/KY-KSPHL-100078/2020 | EPI_ISL_982523 | 11/14/2020 | North America | USA | Kentucky |
| USA/KY-KSPHL-100182/2020 | EPI_ISL_1121995 | 5/17/2020 | North America | USA | Kentucky |
| USA/KY-KSPHL-100206/2020 | EPI_ISL_1184120 | 9/2/2020 | North America | USA | Kentucky |
| USA/KY-KSPHL-100436/2021 | EPI_ISL_2230745 | 4/5/2021 | North America | USA | Kentucky |
| USA/LA-2106170154/2021 | EPI_ISL_2658317 | 6/17/2021 | North America | USA | Louisiana |
| USA/LA-CDC-2-4594847/2021 | EPI_ISL_2987063 | 6/16/2021 | North America | USA | Louisiana |

| | | | | | |
|---|---|---|---|---|---|
| USA/LA-CDC-9KXK-8437/2020 | EPI_ISL_778865 | 12/26/2020 | North America | USA | Louisiana |
| USA/LA-CDC-ASC210113726/2021 | EPI_ISL_3220162 | 7/12/2021 | North America | USA | Louisiana |
| USA/LA-CDC-LC0055215/2021 | EPI_ISL_2045711 | 4/27/2021 | North America | USA | Louisiana |
| USA/LA-CDC-QDX24087618/2021 | EPI_ISL_1924732 | 4/18/2021 | North America | USA | Louisiana |
| USA/LA-CDC-QDX24799392/2021 | EPI_ISL_2367778 | 5/11/2021 | North America | USA | Louisiana |
| USA/LA-EVTL1781/2021 | EPI_ISL_889757 | 1/15/2021 | North America | USA | Louisiana |
| USA/LA-EVTL1869/2020 | EPI_ISL_1040083 | 10/31/2020 | North America | USA | Louisiana |
| USA/LA-EVTL319/2020 | EPI_ISL_578726 | 5/15/2020 | North America | USA | Louisiana |
| USA/LA-EVTL3890/2021 | EPI_ISL_3398174 | 8/2/2021 | North America | USA | Louisiana |
| USA/LA-EVTL3937/2021 | EPI_ISL_3398221 | 8/4/2021 | North America | USA | Louisiana |
| USA/LA-EVTL579/2020 | EPI_ISL_578983 | 6/30/2020 | North America | USA | Louisiana |
| USA/LA-GBCL-26629247/2020 | EPI_ISL_485235 | 4/21/2020 | North America | USA | Louisiana |
| USA/LA-OD-2421525/2021 | EPI_ISL_3161291 | 7/7/2021 | North America | USA | Louisiana |
| USA/LA-OD-2818933/2021 | EPI_ISL_2621529 | 5/24/2021 | North America | USA | Louisiana |
| USA/MA-CDC-QDX24476861/2021 | EPI_ISL_2143031 | 5/2/2021 | North America | USA | Massachusetts |
| USA/MA-CDC-QDX25497790/2021 | EPI_ISL_2599662 | 6/1/2021 | North America | USA | Massachusetts |
| USA/MA-CDCBI-CRSP_AJ2KSVEBL26UJ65M/2021 | EPI_ISL_2991587 | 6/8/2021 | North America | USA | Massachusetts |
| USA/MA-CDCBI-CRSP_NFLFHXV2MZBTYB2Y/2021 | EPI_ISL_3431429 | 8/9/2021 | North America | USA | Massachusetts |
| USA/MA-CDCBI-CRSP_P7UTNT5A4PDIGEPX/2021 | EPI_ISL_3084901 | 7/13/2021 | North America | USA | Massachusetts |
| USA/MA-CDCBI-CRSP_PB7VQMGLB7U7UQI6/2021 | EPI_ISL_1787074 | 4/17/2021 | North America | USA | Massachusetts |

| | | | | | |
|---|---|---|---|---|---|
| USA/MA-CDCBI-CRSP_UYSEYPEL3YAGOX5J/2021 | EPI_ISL_2096608 | 5/3/2021 | North America | USA | Massachusetts |
| USA/MA-CDCBI-CRSP_XVMG3DQHXLWSIQER/2021 | EPI_ISL_1971959 | 4/29/2021 | North America | USA | Massachusetts |
| USA/MA-CDCBI-CRSP_XWZFAFZGHKUD4A4K/2021 | EPI_ISL_3407662 | 8/2/2021 | North America | USA | Massachusetts |
| USA/MA-CDCBI-CRSP_YZHKKK2CCCCAB7T4/2021 | EPI_ISL_3085023 | 7/10/2021 | North America | USA | Massachusetts |
| USA/MA-MASPHL-00896/2020 | EPI_ISL_692844 | 6/4/2020 | North America | USA | Massachusetts |
| USA/MA-MGH-00187/2020 | EPI_ISL_460221 | 3/26/2020 | North America | USA | Massachusetts |
| USA/MA-MGH-00908/2020 | EPI_ISL_791818 | 4/10/2020 | North America | USA | Massachusetts |
| USA/MA-UMASSMED-P001G08/2020 | EPI_ISL_3410324 | 11/9/2020 | North America | USA | Massachusetts |
| USA/MD_NIDDL_2981/2020 | EPI_ISL_491908 | 4/7/2020 | North America | USA | Maryland |
| USA/MD-CDC-LC0027589/2021 | EPI_ISL_1339691 | 3/6/2021 | North America | USA | Maryland |
| USA/MD-CDC-LC0030598/2021 | EPI_ISL_1548904 | 3/13/2021 | North America | USA | Maryland |
| USA/MD-CDC-LC0067100/2021 | EPI_ISL_2481468 | 5/29/2021 | North America | USA | Maryland |
| USA/MD-HP01059/2020 | EPI_ISL_1405498 | 12/24/2020 | North America | USA | Maryland |
| USA/MD-HP03724/2021 | EPI_ISL_1468555 | 3/17/2021 | North America | USA | Maryland |
| USA/MD-HP07416-PIDJVUAIAN/2020 | EPI_ISL_3098657 | 7/15/2020 | North America | USA | Maryland |
| USA/MD-HP08069-PIDRUNZNRT/2021 | EPI_ISL_3373176 | 8/4/2021 | North America | USA | Maryland |
| USA/MD-HP08095-PIDVGZRPFL/2021 | EPI_ISL_3373241 | 8/1/2021 | North America | USA | Maryland |
| USA/MD-IGS-012113501484A/2021 | EPI_ISL_3430851 | 5/15/2021 | North America | USA | Maryland |
| USA/MD-IGS-142117600608A/2021 | EPI_ISL_3435330 | 6/25/2021 | North America | USA | Maryland |
| USA/MD-IGS-52110600392A/2021 | EPI_ISL_3011692 | 4/16/2021 | North America | USA | Maryland |

| | | | | | |
|---|---|---|---|---|---|
| USA/MD-IGS-52111911496A/2021 | EPI_ISL_3011976 | 4/29/2021 | North America | USA | Maryland |
| USA/MD-MDH-0244/2020 | EPI_ISL_602267 | 10/2/2020 | North America | USA | Maryland |
| USA/MD-MDH-1404/2021 | EPI_ISL_1336289 | 3/1/2021 | North America | USA | Maryland |
| USA/MD-MDH-2838/2021 | EPI_ISL_2790043 | 6/15/2021 | North America | USA | Maryland |
| USA/MD-MDH-3537/2021 | EPI_ISL_3235612 | 7/19/2021 | North America | USA | Maryland |
| USA/MD-MDH-3693/2021 | EPI_ISL_3304379 | 7/21/2021 | North America | USA | Maryland |
| USA/MD-NIH-00526/2020 | EPI_ISL_1675172 | 9/28/2020 | North America | USA | Maryland |
| USA/ME-CDC-2-4195195/2021 | EPI_ISL_1711763 | 1/2/2021 | North America | USA | Maine |
| USA/ME-CDC-QDX22257459/2021 | EPI_ISL_1194231 | 2/14/2021 | North America | USA | Maine |
| USA/ME-CDC-QDX24326492/2021 | EPI_ISL_2090527 | 4/19/2021 | North America | USA | Maine |
| USA/ME-CDC-QDX25089047/2021 | EPI_ISL_2439666 | 5/18/2021 | North America | USA | Maine |
| USA/ME-CDCBI-CRSP_5AR77JK5ZQIXHTNQ/2021 | EPI_ISL_3431632 | 8/8/2021 | North America | USA | Maine |
| USA/ME-HETL-H0073/2021 | EPI_ISL_1821058 | 4/16/2021 | North America | USA | Maine |
| USA/ME-HETL-H0146/2021 | EPI_ISL_2101013 | 4/22/2021 | North America | USA | Maine |
| USA/ME-HETL-H0227/2021 | EPI_ISL_2501174 | 5/17/2021 | North America | USA | Maine |
| USA/ME-HETL-J0176/2020 | EPI_ISL_755357 | 9/15/2020 | North America | USA | Maine |
| USA/ME-HETL-J1042/2021 | EPI_ISL_906660 | 1/18/2021 | North America | USA | Maine |
| USA/ME-HETL-J1564/2020 | EPI_ISL_1048621 | 6/3/2020 | North America | USA | Maine |
| USA/ME-HETL-J3544/2021 | EPI_ISL_2802783 | 6/1/2021 | North America | USA | Maine |
| USA/ME-HETL-J4072/2021 | EPI_ISL_3085988 | 6/8/2021 | North America | USA | Maine |

| | | | | | |
|---|---|---|---|---|---|
| USA/ME-HETL-J4394/2021 | EPI_ISL_3186525 | 7/12/2021 | North America | USA | Maine |
| USA/ME-HETL-J4765/2021 | EPI_ISL_3425437 | 7/19/2021 | North America | USA | Maine |
| USA/ME-HETL-J4961/2021 | EPI_ISL_3425545 | 8/3/2021 | North America | USA | Maine |
| USA/MI-CDC-2-4653296/2021 | EPI_ISL_3242033 | 7/1/2021 | North America | USA | Michigan |
| USA/MI-CDC-ASC210053204/2021 | EPI_ISL_1838170 | 4/9/2021 | North America | USA | Michigan |
| USA/MI-CDC-ASC210063163/2021 | EPI_ISL_2204591 | 5/13/2021 | North America | USA | Michigan |
| USA/MI-CDC-ASC210064370/2021 | EPI_ISL_2281083 | 5/17/2021 | North America | USA | Michigan |
| USA/MI-CDC-ASC210108327/2021 | EPI_ISL_2687150 | 6/8/2021 | North America | USA | Michigan |
| USA/MI-CDC-FG-058713/2021 | EPI_ISL_3349216 | 8/2/2021 | North America | USA | Michigan |
| USA/MI-CDC-QDX27212315/2021 | EPI_ISL_3395279 | 7/25/2021 | North America | USA | Michigan |
| USA/MI-CDC-STM-000060086/2021 | EPI_ISL_1991667 | 4/20/2021 | North America | USA | Michigan |
| USA/MI-CDC-STM-000063773/2021 | EPI_ISL_2097363 | 4/26/2021 | North America | USA | Michigan |
| USA/MI-MDHHS-SC20110/2020 | EPI_ISL_436822 | 3/18/2020 | North America | USA | Michigan |
| USA/MI-MDHHS-SC20801/2020 | EPI_ISL_471822 | 5/19/2020 | North America | USA | Michigan |
| USA/MI-MDHHS-SC20852/2020 | EPI_ISL_471873 | 5/8/2020 | North America | USA | Michigan |
| USA/MI-MDHHS-SC20881/2020 | EPI_ISL_471902 | 5/15/2020 | North America | USA | Michigan |
| USA/MI-MDHHS-SC21655/2020 | EPI_ISL_516382 | 7/6/2020 | North America | USA | Michigan |
| USA/MI-MDHHS-SC21755/2020 | EPI_ISL_529873 | 8/3/2020 | North America | USA | Michigan |
| USA/MI-MDHHS-SC24204/2021 | EPI_ISL_1158349 | 2/3/2021 | North America | USA | Michigan |
| USA/MI-MDHHS-SC24764/2021 | EPI_ISL_1289747 | 3/5/2021 | North America | USA | Michigan |

| | | | | | |
|---|---|---|---|---|---|
| USA/MI-MDHHS-SC26816/2021 | EPI_ISL_2246320 | 5/29/2020 | North America | USA | Michigan |
| USA/MI-MDHHS-SC31387/2021 | EPI_ISL_3184628 | 6/21/2021 | North America | USA | Michigan |
| USA/MI-MDHHS-SC31808/2021 | EPI_ISL_3446550 | 8/1/2021 | North America | USA | Michigan |
| USA/MI-UM-10037665894/2021 | EPI_ISL_873061 | 1/4/2021 | North America | USA | Michigan |
| USA/MI-UM-MHM1060/2020 | EPI_ISL_1731770 | 12/16/2020 | North America | USA | Michigan |
| USA/MN-CDC-6281/2020 | EPI_ISL_527692 | 5/27/2020 | North America | USA | Minnesota |
| USA/MN-CDC-6338/2020 | EPI_ISL_527705 | 5/19/2020 | North America | USA | Minnesota |
| USA/MN-CDC-IBX292921008427/2021 | EPI_ISL_2686862 | 6/5/2021 | North America | USA | Minnesota |
| USA/MN-CDC-IBX365497409952/2021 | EPI_ISL_2187049 | 4/30/2021 | North America | USA | Minnesota |
| USA/MN-CDC-IBX636778139438/2021 | EPI_ISL_2186570 | 5/3/2021 | North America | USA | Minnesota |
| USA/MN-CDC-IBX707019223263/2021 | EPI_ISL_2088189 | 4/21/2021 | North America | USA | Minnesota |
| USA/MN-CDC-IBX999591087413/2021 | EPI_ISL_2375780 | 5/12/2021 | North America | USA | Minnesota |
| USA/MN-CDC-QDX26765509/2021 | EPI_ISL_3214187 | 7/9/2021 | North America | USA | Minnesota |
| USA/MN-MDH-1432/2020 | EPI_ISL_514651 | 3/27/2020 | North America | USA | Minnesota |
| USA/MN-MDH-1736/2020 | EPI_ISL_576245 | 9/16/2020 | North America | USA | Minnesota |
| USA/MN-MDH-2107/2020 | EPI_ISL_683736 | 11/3/2020 | North America | USA | Minnesota |
| USA/MN-MDH-2568/2021 | EPI_ISL_913580 | 1/6/2021 | North America | USA | Minnesota |
| USA/MN-MDH-3453/2021 | EPI_ISL_1224887 | 2/8/2021 | North America | USA | Minnesota |
| USA/MN-MDH-4975/2021 | EPI_ISL_1578571 | 1/11/2021 | North America | USA | Minnesota |
| USA/MN-MDH-8471/2021 | EPI_ISL_2958948 | 6/30/2021 | North America | USA | Minnesota |

| | | | | | |
|---|---|---|---|---|---|
| USA/MN-MDH-8715/2021 | EPI_ISL_3118534 | 7/1/2021 | North America | USA | Minnesota |
| USA/MN-MDH-9765/2021 | EPI_ISL_3392718 | 8/3/2021 | North America | USA | Minnesota |
| USA/MN-MDH-9840/2021 | EPI_ISL_3447642 | 8/8/2021 | North America | USA | Minnesota |
| USA/MO-CDC-2-4044649/2020 | EPI_ISL_903898 | 12/8/2020 | North America | USA | Missouri |
| USA/MO-CDC-ASC210065909/2021 | EPI_ISL_2370200 | 5/20/2021 | North America | USA | Missouri |
| USA/MO-CDC-ASC210153893/2021 | EPI_ISL_2875729 | 6/23/2021 | North America | USA | Missouri |
| USA/MO-CDC-QDX21673667/2021 | EPI_ISL_1087607 | 2/1/2021 | North America | USA | Missouri |
| USA/MO-CDC-QDX26343573/2021 | EPI_ISL_2929460 | 6/28/2021 | North America | USA | Missouri |
| USA/MO-CDC-QDX26598366/2021 | EPI_ISL_3213224 | 7/6/2021 | North America | USA | Missouri |
| USA/MO-MSPHL-000211/2020 | EPI_ISL_1362759 | 9/9/2020 | North America | USA | Missouri |
| USA/MO-MSPHL-000252/2020 | EPI_ISL_1362800 | 10/6/2020 | North America | USA | Missouri |
| USA/MO-MSPHL-000496/2020 | EPI_ISL_1392856 | 4/15/2020 | North America | USA | Missouri |
| USA/MO-MSPHL-000509/2020 | EPI_ISL_1392869 | 4/18/2020 | North America | USA | Missouri |
| USA/MO-MSPHL-000541/2020 | EPI_ISL_1489854 | 5/4/2020 | North America | USA | Missouri |
| USA/MO-MSPHL-000864/2021 | EPI_ISL_1791402 | 3/31/2021 | North America | USA | Missouri |
| USA/MO-MSPHL-002111/2021 | EPI_ISL_2304222 | 4/28/2021 | North America | USA | Missouri |
| USA/MO-MSPHL-002504/2021 | EPI_ISL_2753701 | 4/30/2021 | North America | USA | Missouri |
| USA/MO-MSPHL-002672/2021 | EPI_ISL_3010967 | 5/21/2021 | North America | USA | Missouri |
| USA/MO-MSPHL-003022/2021 | EPI_ISL_3161252 | 7/12/2021 | North America | USA | Missouri |
| USA/MO-QDX-3376/2020 | EPI_ISL_884365 | 9/30/2020 | North America | USA | Missouri |

| | | | | | |
|---|---|---|---|---|---|
| USA/MO-QDX-3492/2020 | EPI_ISL_876908 | 9/4/2020 | North America | USA | Missouri |
| USA/MO-QDX-4514/2020 | EPI_ISL_937082 | 10/9/2020 | North America | USA | Missouri |
| USA/MO-UMGC-7088/2021 | EPI_ISL_3346618 | 8/1/2021 | North America | USA | Missouri |
| USA/MO-UMGC-7154/2021 | EPI_ISL_3346657 | 8/1/2021 | North America | USA | Missouri |
| USA/MP-CDC-2-3831005/2020 | EPI_ISL_3353675 | 7/16/2020 | North America | USA | Northern Mariana Islands |
| USA/MP-CDC-2-3831034/2021 | EPI_ISL_3353689 | 1/11/2021 | North America | USA | Northern Mariana Islands |
| USA/MS-ASU10293/2021 | EPI_ISL_3385766 | 8/2/2021 | North America | USA | Mississippi |
| USA/MS-CDC-6571/2020 | EPI_ISL_648023 | 3/13/2020 | North America | USA | Mississippi |
| USA/MS-CDC-6581/2020 | EPI_ISL_648032 | 4/1/2020 | North America | USA | Mississippi |
| USA/MS-CDC-ASC210061574/2021 | EPI_ISL_2180711 | 5/9/2021 | North America | USA | Mississippi |
| USA/MS-CDC-ASC210154930/2021 | EPI_ISL_3321344 | 6/29/2021 | North America | USA | Mississippi |
| USA/MS-CDC-LC0052158/2021 | EPI_ISL_1929487 | 4/20/2021 | North America | USA | Mississippi |
| USA/MS-CDC-LC0093882/2021 | EPI_ISL_3111463 | 7/6/2021 | North America | USA | Mississippi |
| USA/MS-UMMC-M531D11-504819/2021 | EPI_ISL_2134974 | 4/23/2021 | North America | USA | Mississippi |
| USA/MS-UMMC-M653C10-505302/2021 | EPI_ISL_3062766 | 7/12/2021 | North America | USA | Mississippi |
| USA/MS-UMMC-X10D10-210999024272/2021 | EPI_ISL_2920881 | 6/28/2021 | North America | USA | Mississippi |
| USA/MS-UMMC-X3D7-210927000276/2021 | EPI_ISL_2224375 | 5/4/2021 | North America | USA | Mississippi |
| USA/MS-UT2043/2020 | EPI_ISL_1912988 | 6/3/2020 | North America | USA | Mississippi |
| USA/MT-BHDH-1945/2020 | EPI_ISL_900715 | 6/6/2020 | North America | USA | Montana |
| USA/MT-BHDH-245/2021 | EPI_ISL_3330425 | 2/22/2021 | North America | USA | Montana |

| | | | | | |
|---|---|---|---|---|---|
| USA/MT-BHDH-428/2021 | EPI_ISL_3232234 | 4/22/2021 | North America | USA | Montana |
| USA/MT-BHDH-442/2021 | EPI_ISL_2886142 | 4/28/2021 | North America | USA | Montana |
| USA/MT-CDC-2-4504153/2021 | EPI_ISL_3355067 | 5/29/2021 | North America | USA | Montana |
| USA/MT-MTPHL-21410148/2021 | EPI_ISL_1577213 | 4/5/2021 | North America | USA | Montana |
| USA/MT-MTPHL-21410163/2021 | EPI_ISL_1761455 | 4/8/2021 | North America | USA | Montana |
| USA/MT-MTPHL-3805525/2021 | EPI_ISL_2376148 | 5/17/2021 | North America | USA | Montana |
| USA/MT-MTPHL-3815522/2021 | EPI_ISL_2628031 | 6/2/2021 | North America | USA | Montana |
| USA/MT-MTPHL-3816860/2021 | EPI_ISL_2533897 | 6/4/2021 | North America | USA | Montana |
| USA/MT-MTPHL-3834051/2021 | EPI_ISL_2988442 | 7/13/2021 | North America | USA | Montana |
| USA/MT-MTPHL-3836788/2021 | EPI_ISL_3133152 | 7/18/2021 | North America | USA | Montana |
| USA/MT-MTPHL-3844822/2021 | EPI_ISL_3319084 | 8/3/2021 | North America | USA | Montana |
| USA/MT-MTPHL-3844946/2021 | EPI_ISL_3319098 | 8/2/2021 | North America | USA | Montana |
| USA/MT-RML-23/2020 | EPI_ISL_3050744 | 11/30/2020 | North America | USA | Montana |
| USA/MT-UMGC-00174/2020 | EPI_ISL_3333718 | 10/23/2020 | North America | USA | Montana |
| USA/MT-UMGC-00254/2020 | EPI_ISL_3333796 | 10/26/2020 | North America | USA | Montana |
| USA/MT-UMGC-00267/2020 | EPI_ISL_3333807 | 9/8/2020 | North America | USA | Montana |
| USA/NC-CDC-LC0036963/2021 | EPI_ISL_1610079 | 3/25/2021 | North America | USA | North Carolina |
| USA/NC-CDC-LC0050259/2021 | EPI_ISL_1926440 | 4/20/2021 | North America | USA | North Carolina |
| USA/NC-CDC-LC0061763/2021 | EPI_ISL_2306841 | 5/13/2021 | North America | USA | North Carolina |
| USA/NC-CDC-LC0096127/2021 | EPI_ISL_3112893 | 7/11/2021 | North America | USA | North Carolina |

| | | | | | |
|---|---|---|---|---|---|
| USA/NC-CDC-MMB08920237/2021 | EPI_ISL_2647559 | 5/23/2021 | North America | USA | North Carolina |
| USA/NC-CDC-QDX26343170/2021 | EPI_ISL_2929765 | 6/28/2021 | North America | USA | North Carolina |
| USA/NC-CDC-STM-000061115/2021 | EPI_ISL_2010315 | 4/20/2021 | North America | USA | North Carolina |
| USA/NC-CO-HOST_12-1/2020 | EPI_ISL_3088343 | 6/5/2020 | North America | USA | North Carolina |
| USA/NC-NCSLPH_0046/2020 | EPI_ISL_1036271 | 9/18/2020 | North America | USA | North Carolina |
| USA/NC-SLPH-0187/2021 | EPI_ISL_2657665 | 6/8/2021 | North America | USA | North Carolina |
| USA/NC-SLPH-0296/2021 | EPI_ISL_3254961 | 7/17/2021 | North America | USA | North Carolina |
| USA/NC-UNC-0077/2021 | EPI_ISL_965168 | 1/24/2021 | North America | USA | North Carolina |
| USA/NC-UNC-LCCC0353/2020 | EPI_ISL_1334522 | 5/16/2020 | North America | USA | North Carolina |
| USA/NC-UNC-LCCC0375/2020 | EPI_ISL_1334543 | 7/11/2020 | North America | USA | North Carolina |
| USA/ND-NDDH-0014/2020 | EPI_ISL_812246 | 5/8/2020 | North America | USA | North Dakota |
| USA/ND-NDDH-0016/2020 | EPI_ISL_812248 | 4/25/2020 | North America | USA | North Dakota |
| USA/ND-NDDH-01246/2021 | EPI_ISL_2151452 | 4/26/2021 | North America | USA | North Dakota |
| USA/ND-NDDH-01489/2021 | EPI_ISL_2339939 | 5/3/2021 | North America | USA | North Dakota |
| USA/ND-NDDH-02419/2021 | EPI_ISL_2858732 | 6/2/2021 | North America | USA | North Dakota |
| USA/ND-NDDH-02670/2021 | EPI_ISL_2920119 | 5/6/2021 | North America | USA | North Dakota |
| USA/ND-NDDH-2902/2021 | EPI_ISL_3062272 | 4/26/2021 | North America | USA | North Dakota |
| USA/ND-NDDH-3201/2021 | EPI_ISL_3245900 | 6/28/2021 | North America | USA | North Dakota |
| USA/ND-NDDH-3219/2021 | EPI_ISL_3245918 | 7/9/2021 | North America | USA | North Dakota |
| USA/ND-NDDH-3305/2020 | EPI_ISL_3268319 | 7/1/2020 | North America | USA | North Dakota |

| | | | | | |
|---|---|---|---|---|---|
| USA/ND-NDDH-3351/2021 | EPI_ISL_3268271 | 7/31/2021 | North America | USA | North Dakota |
| USA/ND-NDDH-3379/2020 | EPI_ISL_3370353 | 7/16/2020 | North America | USA | North Dakota |
| USA/ND-NDDH-3401/2020 | EPI_ISL_3370375 | 10/12/2020 | North America | USA | North Dakota |
| USA/ND-NDDH-3430/2021 | EPI_ISL_3370402 | 8/2/2021 | North America | USA | North Dakota |
| USA/ND-NDDH-3432/2021 | EPI_ISL_3370404 | 8/1/2021 | North America | USA | North Dakota |
| USA/ND-USAFSAM-S240/2020 | EPI_ISL_812575 | 7/27/2020 | North America | USA | North Dakota |
| USA/NE-001-17/2020 | EPI_ISL_732820 | 9/26/2020 | North America | USA | Nebraska |
| USA/NE-9200147940/2020 | EPI_ISL_2006773 | 12/2/2020 | North America | USA | Nebraska |
| USA/NE-CDC-LC0079129/2021 | EPI_ISL_2872754 | 6/18/2021 | North America | USA | Nebraska |
| USA/NE-MP_NCOV20-34162/2021 | EPI_ISL_2597946 | 7/14/2020 | North America | USA | Nebraska |
| USA/NE-NCOV21-19688/2021 | EPI_ISL_2023409 | 4/28/2021 | North America | USA | Nebraska |
| USA/NE-NCOV21-21765/2021 | EPI_ISL_2308529 | 5/24/2021 | North America | USA | Nebraska |
| USA/NE-NCOV21-22231/2021 | EPI_ISL_2382437 | 4/27/2021 | North America | USA | Nebraska |
| USA/NE-NCOV21-24925/2021 | EPI_ISL_3150787 | 7/22/2021 | North America | USA | Nebraska |
| USA/NE-NCOV21-25116/2021 | EPI_ISL_3234981 | 7/10/2021 | North America | USA | Nebraska |
| USA/NE-NCOV21-26517/2021 | EPI_ISL_3385207 | 8/6/2021 | North America | USA | Nebraska |
| USA/NE-NCOV21-27104/2021 | EPI_ISL_3447353 | 8/11/2021 | North America | USA | Nebraska |
| USA/NE-NPHL-20915/2021 | EPI_ISL_2206999 | 5/10/2021 | North America | USA | Nebraska |
| USA/NE-TESTNE_A2UKYS7/2021 | EPI_ISL_3020995 | 6/26/2021 | North America | USA | Nebraska |
| USA/NE-USAFSAM-S236/2020 | EPI_ISL_812571 | 7/27/2020 | North America | USA | Nebraska |

| | | | | | |
|---|---|---|---|---|---|
| USA/NH-CDC-2-3714179/2020 | EPI_ISL_751582 | 11/6/2020 | North America | USA | New Hampshire |
| USA/NH-CDC-2-3714279/2020 | EPI_ISL_751587 | 11/2/2020 | North America | USA | New Hampshire |
| USA/NH-CDC-2-4282739/2021 | EPI_ISL_1937846 | 4/19/2021 | North America | USA | New Hampshire |
| USA/NH-CDC-LC0019732/2021 | EPI_ISL_1298334 | 2/20/2021 | North America | USA | New Hampshire |
| USA/NH-CDC-LC0037832/2021 | EPI_ISL_1610962 | 3/30/2021 | North America | USA | New Hampshire |
| USA/NH-CDC-LC0052232/2021 | EPI_ISL_1930239 | 4/21/2021 | North America | USA | New Hampshire |
| USA/NH-CDC-LC0065701/2021 | EPI_ISL_2480521 | 5/30/2021 | North America | USA | New Hampshire |
| USA/NH-CDC-LC0104427/2021 | EPI_ISL_3330514 | 7/13/2021 | North America | USA | New Hampshire |
| USA/NH-CDC-QDX24632996/2021 | EPI_ISL_2269138 | 5/4/2021 | North America | USA | New Hampshire |
| USA/NH-CDC-QDX25806490/2021 | EPI_ISL_3091678 | 6/9/2021 | North America | USA | New Hampshire |
| USA/NH-CDC-QDX26076031/2021 | EPI_ISL_2869179 | 6/21/2021 | North America | USA | New Hampshire |
| USA/NH-CDCBI-CRSP_DF2AZEOPBJL56ER2/2021 | EPI_ISL_3408455 | 8/6/2021 | North America | USA | New Hampshire |
| USA/NH-CDCBI-CRSP_DREAP5D2K3BIWWQ3/2021 | EPI_ISL_3085049 | 7/14/2021 | North America | USA | New Hampshire |
| USA/NH-CDCBI-CRSP_WOC364GRIPTYIJO2/2021 | EPI_ISL_3408175 | 8/6/2021 | North America | USA | New Hampshire |
| USA/NH-UW-5500353/2020 | EPI_ISL_2773920 | 7/1/2020 | North America | USA | New Hampshire |
| USA/NH-UW-5500796/2020 | EPI_ISL_2773917 | 6/28/2020 | North America | USA | New Hampshire |
| USA/NH-Yale-1068/2020 | EPI_ISL_1067644 | 5/1/2020 | North America | USA | New Hampshire |
| USA/NJ-CDC-7300/2020 | EPI_ISL_452117 | 3/10/2020 | North America | USA | New Jersey |
| USA/NJ-CDC-ASC210056453/2021 | EPI_ISL_1996069 | 4/11/2021 | North America | USA | New Jersey |
| USA/NJ-CDC-ASC210074044/2021 | EPI_ISL_2039227 | 4/17/2021 | North America | USA | New Jersey |

| | | | | | |
|---|---|---|---|---|---|
| USA/NJ-CDC-FG-034197/2021 | EPI_ISL_2179918 | 5/3/2021 | North America | USA | New Jersey |
| USA/NJ-CDC-LC0010155/2021 | EPI_ISL_1030692 | 1/24/2021 | North America | USA | New Jersey |
| USA/NJ-CDC-LC0022220/2021 | EPI_ISL_1319693 | 3/1/2021 | North America | USA | New Jersey |
| USA/NJ-CDC-LC0054262/2021 | EPI_ISL_2044968 | 4/29/2021 | North America | USA | New Jersey |
| USA/NJ-CDC-LC0057051/2021 | EPI_ISL_2182577 | 5/7/2021 | North America | USA | New Jersey |
| USA/NJ-CDC-LC0111153/2021 | EPI_ISL_3327734 | 7/15/2021 | North America | USA | New Jersey |
| USA/NJ-CDC-QDX27347745/2021 | EPI_ISL_3456337 | 7/31/2021 | North America | USA | New Jersey |
| USA/NJ-MSHSPSP-PV17505/2020 | EPI_ISL_802202 | 8/26/2020 | North America | USA | New Jersey |
| USA/NJ-NYGC-NJ-BioR-420-Ampliseq/2020 | EPI_ISL_2193476 | 9/23/2020 | North America | USA | New Jersey |
| USA/NJ-NYUMC627/2020 | EPI_ISL_444744 | 4/2/2020 | North America | USA | New Jersey |
| USA/NJ-PHEL-20-14942/2020 | EPI_ISL_1164769 | 11/30/2020 | North America | USA | New Jersey |
| USA/NJ-PHEL-21-15987/2021 | EPI_ISL_2649852 | 6/3/2021 | North America | USA | New Jersey |
| USA/NJ-PHEL-V21006096/2021 | EPI_ISL_3477149 | 8/1/2021 | North America | USA | New Jersey |
| USA/NJ-PHEL-V21006335/2021 | EPI_ISL_3398094 | 8/2/2021 | North America | USA | New Jersey |
| USA/NJ-UW-41000002BN05/2020 | EPI_ISL_1233170 | 4/12/2020 | North America | USA | New Jersey |
| USA/NM-CDC-FG-062291/2021 | EPI_ISL_3397188 | 8/2/2021 | North America | USA | New Mexico |
| USA/NM-CDC-IBX983770229146/2021 | EPI_ISL_3353582 | 8/2/2021 | North America | USA | New Mexico |
| USA/NM-CDC-QDX25284120/2021 | EPI_ISL_2440880 | 5/26/2021 | North America | USA | New Mexico |
| USA/NM-CDC-QDX26573456/2021 | EPI_ISL_3303896 | 7/5/2021 | North America | USA | New Mexico |
| USA/NM-NMDOH-2021114435/2021 | EPI_ISL_2322817 | 5/13/2021 | North America | USA | New Mexico |

| | | | | | |
|---|---|---|---|---|---|
| USA/NM-NMDOH-2021130503/2021 | EPI_ISL_2928089 | 6/20/2021 | North America | USA | New Mexico |
| USA/NM-NMDOH-2021131578/2021 | EPI_ISL_2928120 | 6/23/2021 | North America | USA | New Mexico |
| USA/NM-NMDOH-2021135791/2021 | EPI_ISL_3127132 | 7/3/2021 | North America | USA | New Mexico |
| USA/NM-UNM-00034/2020 | EPI_ISL_467615 | 3/15/2020 | North America | USA | New Mexico |
| USA/NM-UNM-00039/2020 | EPI_ISL_467620 | 3/18/2020 | North America | USA | New Mexico |
| USA/NM-UNM-00177/2020 | EPI_ISL_467586 | 4/4/2020 | North America | USA | New Mexico |
| USA/NM-UNM-00646/2020 | EPI_ISL_560334 | 5/9/2020 | North America | USA | New Mexico |
| USA/NM-UNM-TC239891/2021 | EPI_ISL_2228240 | 4/13/2021 | North America | USA | New Mexico |
| USA/NM-UNM-TC240722/2021 | EPI_ISL_2562758 | 4/26/2021 | North America | USA | New Mexico |
| USA/NMDOH-2020117134/2020 | EPI_ISL_542013 | 6/9/2020 | North America | USA | New Mexico |
| USA/NMDOH-2020320960/2020 | EPI_ISL_542067 | 9/1/2020 | North America | USA | New Mexico |
| USA/NMDOH-2020324052/2020 | EPI_ISL_542093 | 9/1/2020 | North America | USA | New Mexico |
| USA/NMDOH-2021107917/2021 | EPI_ISL_2211335 | 4/27/2021 | North America | USA | New Mexico |
| USA/NV-CDC-ASC210035063/2021 | EPI_ISL_2043180 | 4/28/2021 | North America | USA | Nevada |
| USA/NV-CDC-LC0002828/2020 | EPI_ISL_1029913 | 12/31/2020 | North America | USA | Nevada |
| USA/NV-CDC-LC0055980/2021 | EPI_ISL_2046324 | 4/27/2021 | North America | USA | Nevada |
| USA/NV-CDC-LC0068100/2021 | EPI_ISL_2482260 | 5/29/2021 | North America | USA | Nevada |
| USA/NV-CDC-LC0105547/2021 | EPI_ISL_3327299 | 7/12/2021 | North America | USA | Nevada |
| USA/NV-CDC-QDX22062013/2021 | EPI_ISL_1139482 | 2/16/2021 | North America | USA | Nevada |
| USA/NV-CDC-QDX26257762/2021 | EPI_ISL_2875137 | 6/28/2021 | North America | USA | Nevada |

| | | | | | |
|---|---|---|---|---|---|
| USA/NV-CDC-QDX26446241/2021 | EPI_ISL_3018585 | 7/3/2021 | North America | USA | Nevada |
| USA/NV-CDC-QDX27347905/2021 | EPI_ISL_3456110 | 8/1/2021 | North America | USA | Nevada |
| USA/NV-NSPHL-378058/2021 | EPI_ISL_3371856 | 8/2/2021 | North America | USA | Nevada |
| USA/NV-NSPHL-A0132/2020 | EPI_ISL_515400 | 4/29/2020 | North America | USA | Nevada |
| USA/NV-NSPHL-A0157/2020 | EPI_ISL_515421 | 5/14/2020 | North America | USA | Nevada |
| USA/NV-NSPHL-A0200/2020 | EPI_ISL_515454 | 5/27/2020 | North America | USA | Nevada |
| USA/NV-NSPHL-A0206/2020 | EPI_ISL_515460 | 6/2/2020 | North America | USA | Nevada |
| USA/NV-NSPHL-A0207/2020 | EPI_ISL_514674 | 6/5/2020 | North America | USA | Nevada |
| USA/NV-NSPHL-NV2028003/2020 | EPI_ISL_1663895 | 9/17/2020 | North America | USA | Nevada |
| USA/NV-NSPHL-NV2032543/2020 | EPI_ISL_1136005 | 10/12/2020 | North America | USA | Nevada |
| USA/NV-NSPHL-NV2044665/2021 | EPI_ISL_1136406 | 2/19/2021 | North America | USA | Nevada |
| USA/NV-NSPHL-NV2052866/2020 | EPI_ISL_2032401 | 12/7/2020 | North America | USA | Nevada |
| USA/NV-SNPHL-347525/2021 | EPI_ISL_2484866 | 5/21/2021 | North America | USA | Nevada |
| USA/NV-SNPHL-360268/2021 | EPI_ISL_2716939 | 6/15/2021 | North America | USA | Nevada |
| USA/NY-CDC-2-4309841/2021 | EPI_ISL_2018433 | 4/19/2021 | North America | USA | New York |
| USA/NY-CDC-LC0078351/2021 | EPI_ISL_2872877 | 6/15/2021 | North America | USA | New York |
| USA/NY-CDC-QDX24980937/2021 | EPI_ISL_2372109 | 5/10/2021 | North America | USA | New York |
| USA/NY-CUIMC-NP-3552/2020 | EPI_ISL_3232382 | 11/28/2020 | North America | USA | New York |
| USA/NY-MSHSPSP-PV14657/2020 | EPI_ISL_802096 | 5/24/2020 | North America | USA | New York |
| USA/NY-MSHSPSP-PV17526/2020 | EPI_ISL_801886 | 8/24/2020 | North America | USA | New York |

| | | | | | |
|---|---|---|---|---|---|
| USA/NY-NYCPHL-000954/2020 | EPI_ISL_632991 | 10/1/2020 | North America | USA | New York |
| USA/NY-NYCPHL-004858/2021 | EPI_ISL_2758641 | 6/10/2021 | North America | USA | New Jersey |
| USA/NY-NYUMC742/2020 | EPI_ISL_451457 | 3/21/2020 | North America | USA | New York |
| USA/NY-PRL-2021_0426_02F24/2021 | EPI_ISL_1828070 | 4/24/2021 | North America | USA | New York |
| USA/NY-PRL-2021_0628_00A16/2021 | EPI_ISL_2827492 | 6/23/2021 | North America | USA | New York |
| USA/NY-PRL-2021_0808_01I08/2021 | EPI_ISL_3394575 | 8/3/2021 | North America | USA | New York |
| USA/NY-PRL-2021_0809_01A08/2021 | EPI_ISL_3394759 | 8/3/2021 | North America | USA | New York |
| USA/NY-UB-00529/2021 | EPI_ISL_1400987 | 1/6/2021 | North America | USA | New York |
| USA/NY-Wadsworth-194579-01/2020 | EPI_ISL_677035 | 8/7/2020 | North America | USA | New York |
| USA/NY-Wadsworth-21020483-01/2021 | EPI_ISL_1227530 | 2/3/2021 | North America | USA | New York |
| USA/NY1-PV08001/2020 | EPI_ISL_414476 | 2/29/2020 | North America | USA | New York |
| USA/OH-CDC-2-4567984/2021 | EPI_ISL_2840335 | 4/29/2021 | North America | USA | Ohio |
| USA/OH-CDC-FG-058399/2021 | EPI_ISL_3349289 | 8/1/2021 | North America | USA | Ohio |
| USA/OH-CDC-FG-068399/2021 | EPI_ISL_3456666 | 8/6/2021 | North America | USA | Ohio |
| USA/OH-CDC-LC0022234/2021 | EPI_ISL_1319558 | 2/26/2021 | North America | USA | Ohio |
| USA/OH-CDC-MMB09384834/2021 | EPI_ISL_3427736 | 7/15/2021 | North America | USA | Ohio |
| USA/OH-CDC-QDX23356009/2021 | EPI_ISL_1526048 | 3/25/2021 | North America | USA | Ohio |
| USA/OH-CDC-QDX26076145/2021 | EPI_ISL_2868694 | 6/19/2021 | North America | USA | Ohio |
| USA/OH-CDC-QDX27053522/2021 | EPI_ISL_3352779 | 7/19/2021 | North America | USA | Ohio |
| USA/OH-GD-SID-21050713664/2021 | EPI_ISL_2614022 | 5/7/2021 | North America | USA | Ohio |

| | | | | | |
|---|---|---|---|---|---|
| USA/OH-GD-SID-21052042281/2021 | EPI_ISL_2536019 | 5/20/2021 | North America | USA | Ohio |
| USA/OH-ODH-SC0063830/2020 | EPI_ISL_2644673 | 7/20/2020 | North America | USA | Ohio |
| USA/OH-ODH-SC0156996/2020 | EPI_ISL_2557247 | 11/12/2020 | North America | USA | Ohio |
| USA/OH-ODH-SC034594/2020 | EPI_ISL_1389161 | 4/20/2020 | North America | USA | Ohio |
| USA/OH-ODH-SC037802/2020 | EPI_ISL_2307083 | 5/16/2020 | North America | USA | Ohio |
| USA/OH-ODH-SC22228/2020 | EPI_ISL_940860 | 3/27/2020 | North America | USA | Ohio |
| USA/OH-ODH-SC35013/2020 | EPI_ISL_765209 | 4/23/2020 | North America | USA | Ohio |
| USA/OH-OSU-21AM-113CO02762/2021 | EPI_ISL_1911097 | 4/23/2021 | North America | USA | Ohio |
| USA/OH-SRL_254602759/2020 | EPI_ISL_1941717 | 9/8/2020 | North America | USA | Ohio |
| USA/OH-USAFSAM-S264/2020 | EPI_ISL_812594 | 8/7/2020 | North America | USA | Ohio |
| USA/OH-USAFSAM-S4045/2021 | EPI_ISL_2966740 | 6/23/2021 | North America | USA | Ohio |
| USA/OH-USAFSAM-S421/2020 | EPI_ISL_812708 | 9/17/2020 | North America | USA | Ohio |
| USA/OH-USAFSAM-S480/2020 | EPI_ISL_831775 | 10/15/2020 | North America | USA | Ohio |
| USA/OK-ADDL-01/2020 | EPI_ISL_535364 | 4/10/2020 | North America | USA | Oklahoma |
| USA/OK-ADDL-02/2020 | EPI_ISL_535361 | 5/27/2020 | North America | USA | Oklahoma |
| USA/OK-CDC-2-3693479/2020 | EPI_ISL_747158 | 11/17/2020 | North America | USA | Oklahoma |
| USA/OK-CDC-2-4195490/2021 | EPI_ISL_1711803 | 4/5/2021 | North America | USA | Oklahoma |
| USA/OK-CDC-4114731-001/2021 | EPI_ISL_2295286 | 4/26/2021 | North America | USA | Oklahoma |
| USA/OK-CDC-4130814-001/2021 | EPI_ISL_2422447 | 5/10/2021 | North America | USA | Oklahoma |
| USA/OK-CDC-4158200-001/2021 | EPI_ISL_2562736 | 5/27/2021 | North America | USA | Oklahoma |

| | | | | | |
|---|---|---|---|---|---|
| USA/OK-CDC-ASC210075358/2021 | EPI_ISL_2041760 | 4/18/2021 | North America | USA | Oklahoma |
| USA/OK-CDC-FG-059258/2021 | EPI_ISL_3350539 | 8/2/2021 | North America | USA | Oklahoma |
| USA/OK-CDC-LC0079883/2021 | EPI_ISL_2873133 | 6/13/2021 | North America | USA | Oklahoma |
| USA/OK-CDC-LC0092946/2021 | EPI_ISL_3111369 | 7/3/2021 | North America | USA | Oklahoma |
| USA/OK-CDC-LC0095126/2021 | EPI_ISL_3111987 | 7/11/2021 | North America | USA | Oklahoma |
| USA/OK-CDC-QDX22523967/2021 | EPI_ISL_1267256 | 2/25/2021 | North America | USA | Oklahoma |
| USA/OK-CDC-QDX25662884/2021 | EPI_ISL_2652341 | 6/7/2021 | North America | USA | Oklahoma |
| USA/OK-CDC-STM-A100101/2020 | EPI_ISL_850942 | 12/31/2020 | North America | USA | Oklahoma |
| USA/OK-KHEL-4782/2021 | EPI_ISL_3477543 | 8/11/2021 | North America | USA | Oklahoma |
| USA/OK-QDX-4502/2020 | EPI_ISL_937101 | 10/3/2020 | North America | USA | Oklahoma |
| USA/OK-QDX-749/2020 | EPI_ISL_571551 | 3/17/2020 | North America | USA | Oklahoma |
| USA/OK-USAFSAM-S409/2020 | EPI_ISL_812699 | 9/11/2020 | North America | USA | Oklahoma |
| USA/OR-CDC-FG-035603/2021 | EPI_ISL_2525962 | 5/17/2021 | North America | USA | Oregon |
| USA/OR-CDC-FG-040577/2021 | EPI_ISL_3016668 | 7/6/2021 | North America | USA | Oregon |
| USA/OR-CDC-QDX25848198/2021 | EPI_ISL_2871666 | 6/13/2021 | North America | USA | Oregon |
| USA/OR-CDC-QDX26765257/2021 | EPI_ISL_3214291 | 7/13/2021 | North America | USA | Oregon |
| USA/OR-MAP000456/2021 | EPI_ISL_2304025 | 5/4/2021 | North America | USA | Oregon |
| USA/OR-OHSU-11218/2021 | EPI_ISL_2080862 | 4/16/2021 | North America | USA | Oregon |
| USA/OR-OHSU-11444/2021 | EPI_ISL_2462103 | 4/23/2021 | North America | USA | Oregon |
| USA/OR-OHSU-11559/2021 | EPI_ISL_2462206 | 4/28/2021 | North America | USA | Oregon |

| | | | | | |
|---|---|---|---|---|---|
| USA/OR-OHSU-9018/2020 | EPI_ISL_1171293 | 12/23/2020 | North America | USA | Oregon |
| USA/OR-OHSU-9570/2021 | EPI_ISL_1122213 | 1/9/2021 | North America | USA | Oregon |
| USA/OR-OSPHL01013/2021 | EPI_ISL_2693022 | 6/17/2021 | North America | USA | Oregon |
| USA/OR-OSPHL01677/2021 | EPI_ISL_3370939 | 8/3/2021 | North America | USA | Oregon |
| USA/OR-OSPHL01729/2021 | EPI_ISL_3404415 | 8/1/2021 | North America | USA | Oregon |
| USA/OR-UOGC3F-000118/2021 | EPI_ISL_1508888 | 2/22/2021 | North America | USA | Oregon |
| USA/PA_1802/2020 | EPI_ISL_424881 | 3/5/2020 | North America | USA | Pennsylvania |
| USA/PA-CDC-9N0A-8975/2021 | EPI_ISL_1379693 | 1/8/2021 | North America | USA | Pennsylvania |
| USA/PA-CDC-ASC210110076/2021 | EPI_ISL_2785164 | 6/18/2021 | North America | USA | Pennsylvania |
| USA/PA-CDC-FG-062264/2021 | EPI_ISL_3397262 | 8/2/2021 | North America | USA | Pennsylvania |
| USA/PA-CDC-LC0000619/2020 | EPI_ISL_1029252 | 12/27/2020 | North America | USA | Pennsylvania |
| USA/PA-CDC-LC0043334/2021 | EPI_ISL_1684188 | 4/8/2021 | North America | USA | Pennsylvania |
| USA/PA-CDC-LC0059453/2021 | EPI_ISL_2185406 | 5/7/2021 | North America | USA | Pennsylvania |
| USA/PA-CDC-LC0110686/2021 | EPI_ISL_3327558 | 7/13/2021 | North America | USA | Pennsylvania |
| USA/PA-CDC-MMB09382065/2021 | EPI_ISL_3427516 | 7/14/2021 | North America | USA | Pennsylvania |
| USA/PA-CDC-QDX26380626/2021 | EPI_ISL_3018362 | 6/27/2021 | North America | USA | Pennsylvania |
| USA/PA-CDC-QDX27388114/2021 | EPI_ISL_3456143 | 8/1/2021 | North America | USA | Pennsylvania |
| USA/PA-CDC-STM-000007819/2021 | EPI_ISL_1016672 | 1/30/2021 | North America | USA | Pennsylvania |
| USA/PA-CDC-STM-000061445/2021 | EPI_ISL_2010538 | 4/22/2021 | North America | USA | Pennsylvania |
| USA/PA-CDC-STM-000067024/2021 | EPI_ISL_2160540 | 4/30/2021 | North America | USA | Pennsylvania |

| | | | | | |
|---|---|---|---|---|---|
| USA/PA-CDC-STM-000071746/2021 | EPI_ISL_2321053 | 5/9/2021 | North America | USA | Pennsylvania |
| USA/PA-MGEL-00500/2020 | EPI_ISL_682026 | 8/24/2020 | North America | USA | Pennsylvania |
| USA/PA-MGEL-00971/2020 | EPI_ISL_853324 | 11/14/2020 | North America | USA | Pennsylvania |
| USA/PA-MGEL-01080/2020 | EPI_ISL_1137218 | 11/14/2020 | North America | USA | Pennsylvania |
| USA/PA-MGEL-01940/2021 | EPI_ISL_1732405 | 1/19/2021 | North America | USA | Pennsylvania |
| USA/PA-MGEL-02220/2021 | EPI_ISL_1732504 | 4/16/2021 | North America | USA | Pennsylvania |
| USA/PA-VSP0100/2020 | EPI_ISL_1511440 | 5/8/2020 | North America | USA | Pennsylvania |
| USA/PA-VSP0639/2021 | EPI_ISL_1511521 | 1/21/2021 | North America | USA | Pennsylvania |
| USA/PR-07252021-PRI4979/2021 | EPI_ISL_3342546 | 7/25/2021 | North America | USA | Puerto Rico |
| USA/PR-07272021-PR-0072/2021 | EPI_ISL_3342529 | 7/27/2021 | North America | USA | Puerto Rico |
| USA/PR-08012021-PRI4985/2021 | EPI_ISL_3342551 | 8/1/2021 | North America | USA | Puerto Rico |
| USA/PR-08022021-PRI4492/2021 | EPI_ISL_3342499 | 8/2/2021 | North America | USA | Puerto Rico |
| USA/PR-AGN5/2020 | EPI_ISL_513292 | 6/29/2020 | North America | USA | Puerto Rico |
| USA/PR-B40A/2020 | EPI_ISL_527387 | 8/6/2020 | North America | USA | Puerto Rico |
| USA/PR-CDC-ASC210017760/2021 | EPI_ISL_2242160 | 3/23/2021 | North America | USA | Puerto Rico |
| USA/PR-CDC-ASC210060057/2021 | EPI_ISL_2146636 | 5/6/2021 | North America | USA | Puerto Rico |
| USA/PR-CDC-ASC210060406/2021 | EPI_ISL_2147653 | 5/7/2021 | North America | USA | Puerto Rico |
| USA/PR-CDC-ASC210073271/2021 | EPI_ISL_2202575 | 4/16/2021 | North America | USA | Puerto Rico |
| USA/PR-CDC-LC0016907/2021 | EPI_ISL_1163003 | 2/10/2021 | North America | USA | Puerto Rico |
| USA/PR-CDC-QDX23269210/2021 | EPI_ISL_1552938 | 3/24/2021 | North America | USA | Puerto Rico |

| | | | | | |
|---|---|---|---|---|---|
| USA/PR-CDC-QDX24236457/2021 | EPI_ISL_2047164 | 4/23/2021 | North America | USA | Puerto Rico |
| USA/PR-CDC-S142/2021 | EPI_ISL_1168667 | 1/14/2021 | North America | USA | Puerto Rico |
| USA/PR-CDC-S34/2020 | EPI_ISL_940907 | 6/11/2020 | North America | USA | Puerto Rico |
| USA/PR-CDC-S54/2020 | EPI_ISL_940927 | 7/23/2020 | North America | USA | Puerto Rico |
| USA/PR-Yale-5807/2021 | EPI_ISL_2860323 | 6/21/2021 | North America | USA | Puerto Rico |
| USA/PR-Yale-6001/2021 | EPI_ISL_3025394 | 6/10/2021 | North America | USA | Puerto Rico |
| USA/RI_0702/2020 | EPI_ISL_424887 | 3/5/2020 | North America | USA | Rhode Island |
| USA/RI_RKL_001_55410_AM623/2020 | EPI_ISL_2000598 | 6/18/2020 | North America | USA | Rhode Island |
| USA/RI-Broad_RIDOH-00045/2020 | EPI_ISL_872708 | 11/30/2020 | North America | USA | Rhode Island |
| USA/RI-CDC-2-4679265/2021 | EPI_ISL_3305805 | 7/11/2021 | North America | USA | Rhode Island |
| USA/RI-CDC-9KXU-8439/2020 | EPI_ISL_778866 | 12/21/2020 | North America | USA | Rhode Island |
| USA/RI-CDC-LC0052415/2021 | EPI_ISL_1930140 | 4/21/2021 | North America | USA | Rhode Island |
| USA/RI-CDC-LC0061767/2021 | EPI_ISL_2306762 | 5/12/2021 | North America | USA | Rhode Island |
| USA/RI-CDC-LC0082480/2021 | EPI_ISL_2926496 | 6/26/2021 | North America | USA | Rhode Island |
| USA/RI-CDCBI-CRSP_PMK5JHE3DE3BQ6FA/2021 | EPI_ISL_3408527 | 8/3/2021 | North America | USA | Rhode Island |
| USA/RI-CDCBI-CRSP_QWNI6FALAN5GN2XM/2021 | EPI_ISL_1825304 | 4/25/2021 | North America | USA | Rhode Island |
| USA/RI-CDCBI-CRSP_TXUGAGSURIDWE2UU/2021 | EPI_ISL_3407440 | 8/2/2021 | North America | USA | Rhode Island |
| USA/RI-CDCBI-RIDOH_00700/2021 | EPI_ISL_1407387 | 2/25/2021 | North America | USA | Rhode Island |
| USA/RI-CDCBI-RIDOH_10119/2021 | EPI_ISL_2362392 | 5/11/2021 | North America | USA | Rhode Island |
| USA/RI-CDCBI-RIDOH_10457/2021 | EPI_ISL_2713271 | 6/1/2021 | North America | USA | Rhode Island |

| | | | | | |
|---|---|---|---|---|---|
| USA/RI-RISHL-077926/2021 | EPI_ISL_3398884 | 7/24/2021 | North America | USA | Rhode Island |
| USA/RI-RKL_001_RK003_AM635/2020 | EPI_ISL_3032075 | 8/20/2020 | North America | USA | Rhode Island |
| USA/SC-CDC-2-4309029/2021 | EPI_ISL_3354864 | 4/22/2021 | North America | USA | South Carolina |
| USA/SC-CDC-ASC210002997/2021 | EPI_ISL_2241805 | 3/7/2021 | North America | USA | South Carolina |
| USA/SC-CDC-MMB09265565/2021 | EPI_ISL_3065156 | 6/28/2021 | North America | USA | South Carolina |
| USA/SC-CDC-MMB09358025/2021 | EPI_ISL_3427907 | 7/12/2021 | North America | USA | South Carolina |
| USA/SC-CDC-MMB09382037/2021 | EPI_ISL_3427294 | 7/14/2021 | North America | USA | South Carolina |
| USA/SC-DHEC-0021/2021 | EPI_ISL_873187 | 1/3/2021 | North America | USA | South Carolina |
| USA/SC-DHEC-0074/2020 | EPI_ISL_529176 | 4/1/2020 | North America | USA | South Carolina |
| USA/SC-DHEC-0141/2021 | EPI_ISL_1303255 | 1/29/2021 | North America | USA | South Carolina |
| USA/SC-DHEC-1424/2021 | EPI_ISL_2289158 | 4/16/2021 | North America | USA | South Carolina |
| USA/SC-DHEC-1708/2021 | EPI_ISL_2312997 | 5/4/2021 | North America | USA | South Carolina |
| USA/SC-DHEC-1736/2021 | EPI_ISL_2354486 | 5/5/2021 | North America | USA | South Carolina |
| USA/SC-DHEC-2089/2021 | EPI_ISL_2758626 | 6/10/2021 | North America | USA | South Carolina |
| USA/SC-MCRDPI_20_1342-T42/2020 | EPI_ISL_2894765 | 8/4/2020 | North America | USA | South Carolina |
| USA/SC-MCRDPI_20_2106-T28/2020 | EPI_ISL_2894832 | 9/8/2020 | North America | USA | South Carolina |
| USA/SC-MCRDPI_20_3171-T56/2020 | EPI_ISL_2894852 | 10/27/2020 | North America | USA | South Carolina |
| USA/SC-MUSC-153/2020 | EPI_ISL_1482661 | 12/9/2020 | North America | USA | South Carolina |
| USA/SC-USAFSAM-S518/2020 | EPI_ISL_831804 | 11/16/2020 | North America | USA | South Carolina |
| USA/SC-USC-COM0003/2020 | EPI_ISL_493129 | 4/4/2020 | North America | USA | South Carolina |

| | | | | | |
|---|---|---|---|---|---|
| USA/SD-CDC-2-4243052/2021 | EPI_ISL_1823580 | 4/14/2021 | North America | USA | South Dakota |
| USA/SD-CDC-2-4307105/2021 | EPI_ISL_2018209 | 4/18/2021 | North America | USA | South Dakota |
| USA/SD-CDC-2-4462337/2021 | EPI_ISL_2645283 | 5/27/2021 | North America | USA | South Dakota |
| USA/SD-CDC-2-4594849/2021 | EPI_ISL_2987036 | 6/8/2021 | North America | USA | South Dakota |
| USA/SD-CDC-2-4611168/2021 | EPI_ISL_3355170 | 6/24/2021 | North America | USA | South Dakota |
| USA/SD-CDC-2-4634888/2021 | EPI_ISL_3127976 | 7/8/2021 | North America | USA | South Dakota |
| USA/SD-CDC-2-4679376/2021 | EPI_ISL_3305789 | 7/21/2021 | North America | USA | South Dakota |
| USA/SD-SDPHL-0001/2020 | EPI_ISL_569609 | 4/3/2020 | North America | USA | South Dakota |
| USA/SD-SDPHL-0005/2020 | EPI_ISL_569613 | 7/7/2020 | North America | USA | South Dakota |
| USA/SD-SDPHL-0106/2020 | EPI_ISL_1314862 | 11/18/2020 | North America | USA | South Dakota |
| USA/SD-SDPHL-0107/2020 | EPI_ISL_1314863 | 11/18/2020 | North America | USA | South Dakota |
| USA/SD-SDPHL-0330/2021 | EPI_ISL_2102011 | 4/27/2021 | North America | USA | South Dakota |
| USA/SD-SDPHL-0351/2021 | EPI_ISL_2427233 | 5/3/2021 | North America | USA | South Dakota |
| USA/SD-SDPHL-0425/2020 | EPI_ISL_3047985 | 8/26/2020 | North America | USA | South Dakota |
| USA/SD-UMGC_111/2020 | EPI_ISL_1016994 | 5/19/2020 | North America | USA | South Dakota |
| USA/SD-UW-MAYO73/2020 | EPI_ISL_500517 | 4/2/2020 | North America | USA | South Dakota |
| USA/TN-CDC-2-4069707/2020 | EPI_ISL_1711079 | 11/27/2020 | North America | USA | Tennessee |
| USA/TN-CDC-3981282-001/2020 | EPI_ISL_2859128 | 9/9/2020 | North America | USA | Tennessee |
| USA/TN-CDC-4167625-001/2021 | EPI_ISL_2710041 | 1/13/2021 | North America | USA | Tennessee |
| USA/TN-CDC-4184422-002/2021 | EPI_ISL_3064747 | 6/11/2021 | North America | USA | Tennessee |

| | | | | | |
|---|---|---|---|---|---|
| USA/TN-CDC-ASC210031871/2021 | EPI_ISL_2370295 | 4/24/2021 | North America | USA | Tennessee |
| USA/TN-CDC-ASC210067093/2021 | EPI_ISL_2489505 | 5/27/2021 | North America | USA | Tennessee |
| USA/TN-CDC-ASC210106215/2021 | EPI_ISL_2530942 | 5/30/2021 | North America | USA | Tennessee |
| USA/TN-CDC-ASC210110574/2021 | EPI_ISL_2832536 | 6/21/2021 | North America | USA | Tennessee |
| USA/TN-CDC-ASC210161182/2021 | EPI_ISL_3324909 | 7/20/2021 | North America | USA | Tennessee |
| USA/TN-CDC-FG-014286/2021 | EPI_ISL_1556077 | 3/23/2021 | North America | USA | Tennessee |
| USA/TN-CDC-FG-061791/2021 | EPI_ISL_3352153 | 8/2/2021 | North America | USA | Tennessee |
| USA/TN-CDC-FG-069061/2021 | EPI_ISL_3457266 | 8/6/2021 | North America | USA | Tennessee |
| USA/TN-CDC-JHTP-9956/2020 | EPI_ISL_1373905 | 6/18/2020 | North America | USA | Tennessee |
| USA/TN-CDC-JHU8-1597/2020 | EPI_ISL_1373884 | 7/13/2020 | North America | USA | Tennessee |
| USA/TN-CDC-JHWD-0396/2020 | EPI_ISL_1373992 | 12/10/2020 | North America | USA | Tennessee |
| USA/TN-CDC-LC0048396/2021 | EPI_ISL_1802023 | 4/19/2021 | North America | USA | Tennessee |
| USA/TN-CDC-QDX26763429/2021 | EPI_ISL_3214010 | 7/9/2021 | North America | USA | Tennessee |
| USA/TN-QDX-2517/2020 | EPI_ISL_604842 | 3/14/2020 | North America | USA | Tennessee |
| USA/TX-BSWTemple-ILL-R8-0082/2021 | EPI_ISL_2083079 | 5/3/2021 | North America | USA | Texas |
| USA/TX-CDC-4175344-001/2020 | EPI_ISL_2754937 | 12/9/2020 | North America | USA | Texas |
| USA/TX-CDC-ASC210075810/2021 | EPI_ISL_2041158 | 4/17/2021 | North America | USA | Texas |
| USA/TX-CDC-FG-057505/2021 | EPI_ISL_3349477 | 7/30/2021 | North America | USA | Texas |
| USA/TX-CDC-FG-057543/2021 | EPI_ISL_3460992 | 8/1/2021 | North America | USA | Texas |
| USA/TX-CDC-FG-068192/2021 | EPI_ISL_3456636 | 8/6/2021 | North America | USA | Texas |

| | | | | | |
|---|---|---|---|---|---|
| USA/TX-CDC-LC0078376/2021 | EPI_ISL_2873074 | 6/20/2021 | North America | USA | Texas |
| USA/TX-CDC-QDX26115952/2021 | EPI_ISL_2869232 | 6/20/2021 | North America | USA | Texas |
| USA/TX-CDC-QDX26488130/2021 | EPI_ISL_3114459 | 7/3/2021 | North America | USA | Texas |
| USA/TX-CDC-STM-000045138/2021 | EPI_ISL_1592745 | 3/26/2021 | North America | USA | Texas |
| USA/TX-DSHS-6312/2021 | EPI_ISL_2017922 | 4/20/2021 | North America | USA | Texas |
| USA/TX-HHD-210520C566/2021 | EPI_ISL_2210479 | 5/17/2021 | North America | USA | Texas |
| USA/TX-HMH-2119/2020 | EPI_ISL_545130 | 5/20/2020 | North America | USA | Texas |
| USA/TX-HMH-MCoV-16050/2020 | EPI_ISL_785734 | 10/27/2020 | North America | USA | Texas |
| USA/TX-HMH-MCoV-29955/2021 | EPI_ISL_1304509 | 1/3/2021 | North America | USA | Texas |
| USA/TX-HMH-MCoV-40326/2020 | EPI_ISL_2223508 | 8/3/2020 | North America | USA | Texas |
| USA/TX-TCH-TCMC04004/2020 | EPI_ISL_1446537 | 4/27/2020 | North America | USA | Texas |
| USA/TX1/2020 | EPI_ISL_411956 | 2/11/2020 | North America | USA | Texas |
| USA/UT-01601/2020 | EPI_ISL_449835 | 4/6/2020 | North America | USA | Utah |
| USA/UT-CDC-2-3973453/2021 | EPI_ISL_1272833 | 2/16/2021 | North America | USA | Utah |
| USA/UT-CDC-ASC210028951/2021 | EPI_ISL_2041427 | 4/18/2021 | North America | USA | Utah |
| USA/UT-QDX-2302/2020 | EPI_ISL_604616 | 3/13/2020 | North America | USA | Utah |
| USA/UT-UPHL-2104848080/2021 | EPI_ISL_2496645 | 2/1/2021 | North America | USA | Utah |
| USA/UT-UPHL-210528169309/2021 | EPI_ISL_2458406 | 5/15/2021 | North America | USA | Utah |
| USA/UT-UPHL-2105285192/2021 | EPI_ISL_2292857 | 4/27/2021 | North America | USA | Utah |
| USA/UT-UPHL-2105452715/2021 | EPI_ISL_2290175 | 1/15/2021 | North America | USA | Utah |

| | | | | | |
|---|---|---|---|---|---|
| USA/UT-UPHL-210609852534/2021 | EPI_ISL_2856093 | 5/10/2021 | North America | USA | Utah |
| USA/UT-UPHL-210617258358/2021 | EPI_ISL_2856290 | 6/4/2021 | North America | USA | Utah |
| USA/UT-UPHL-210716546212/2021 | EPI_ISL_3268814 | 6/30/2021 | North America | USA | Utah |
| USA/UT-UPHL-210721947137/2021 | EPI_ISL_3269630 | 7/12/2021 | North America | USA | Utah |
| USA/UT-UPHL-210729610418/2021 | EPI_ISL_3263079 | 7/13/2021 | North America | USA | Utah |
| USA/UT-UPHL-210814516196/2021 | EPI_ISL_3450526 | 8/2/2021 | North America | USA | Utah |
| USA/UT-UPHL-210814710707/2021 | EPI_ISL_3450228 | 8/2/2021 | North America | USA | Utah |
| USA/VA-CDC-LC0000672/2020 | EPI_ISL_1027671 | 12/27/2020 | North America | USA | Virginia |
| USA/VA-CDC-LC0008252/2021 | EPI_ISL_1020626 | 1/20/2021 | North America | USA | Virginia |
| USA/VA-CDC-LC0050751/2021 | EPI_ISL_1926847 | 4/21/2021 | North America | USA | Virginia |
| USA/VA-CDC-LC0055210/2021 | EPI_ISL_2045519 | 4/27/2021 | North America | USA | Virginia |
| USA/VA-CDC-LC0057909/2021 | EPI_ISL_2184111 | 5/2/2021 | North America | USA | Virginia |
| USA/VA-CDC-LC0065978/2021 | EPI_ISL_2480610 | 5/17/2021 | North America | USA | Virginia |
| USA/VA-CDC-LC0070442/2021 | EPI_ISL_2611484 | 6/1/2021 | North America | USA | Virginia |
| USA/VA-CDC-QDX27388044/2021 | EPI_ISL_3456396 | 8/1/2021 | North America | USA | Virginia |
| USA/VA-CDC-QDX27388047/2021 | EPI_ISL_3456404 | 8/1/2021 | North America | USA | Virginia |
| USA/VA-DCLS-0014/2020 | EPI_ISL_419708 | 3/12/2020 | North America | USA | Virginia |
| USA/VA-DCLS-0768/2020 | EPI_ISL_526884 | 6/9/2020 | North America | USA | Virginia |
| USA/VA-DCLS-2508/2020 | EPI_ISL_734283 | 11/16/2020 | North America | USA | Virginia |
| USA/VA-DCLS-2915/2020 | EPI_ISL_978188 | 12/23/2020 | North America | USA | Virginia |

| | | | | | |
|---|---|---|---|---|---|
| USA/VA-DCLS-5766/2021 | EPI_ISL_3025855 | 6/27/2021 | North America | USA | Virginia |
| USA/VA-DCLS-5792/2021 | EPI_ISL_3025877 | 7/8/2021 | North America | USA | Virginia |
| USA/VA-DCLS-6411/2021 | EPI_ISL_3447409 | 7/24/2021 | North America | USA | Virginia |
| USA/VI-CDC-2-4122528/2021 | EPI_ISL_3354588 | 3/1/2021 | North America | USA | Virgin Islands |
| USA/VI-CDC-2-4420272/2021 | EPI_ISL_2384232 | 4/29/2021 | North America | USA | Virgin Islands |
| USA/VI-CDC-3943/2020 | EPI_ISL_450806 | 4/8/2020 | North America | USA | Virgin Islands |
| USA/VI-Yale-5624/2021 | EPI_ISL_3045994 | 5/27/2021 | North America | USA | Virgin Islands |
| USA/VI-Yale-5683/2021 | EPI_ISL_3046022 | 6/3/2021 | North America | USA | Virgin Islands |
| USA/VI-Yale-5697/2021 | EPI_ISL_3046031 | 5/20/2021 | North America | USA | Virgin Islands |
| USA/VI-Yale-5830/2021 | EPI_ISL_3045998 | 4/23/2021 | North America | USA | Virgin Islands |
| USA/VI-Yale-5851/2021 | EPI_ISL_3046013 | 4/8/2021 | North America | USA | Virgin Islands |
| USA/VI-Yale-6044/2021 | EPI_ISL_3045948 | 6/28/2021 | North America | USA | Virgin Islands |
| USA/VI-Yale-6525/2021 | EPI_ISL_3236506 | 7/7/2021 | North America | USA | Virgin Islands |
| USA/VI-Yale-7016/2021 | EPI_ISL_3347342 | 7/29/2021 | North America | USA | Virgin Islands |
| USA/VT-CDC-2-4044713/2020 | EPI_ISL_3354359 | 11/27/2020 | North America | USA | Vermont |
| USA/VT-CDC-ASC210003672/2021 | EPI_ISL_1446175 | 3/8/2021 | North America | USA | Vermont |
| USA/VT-CDC-QDX21490895/2021 | EPI_ISL_1087035 | 1/24/2021 | North America | USA | Vermont |
| USA/VT-CDCBI-CRSP_7OHT5DL7QJHEEMCN/2021 | EPI_ISL_3007474 | 6/29/2021 | North America | USA | Vermont |
| USA/VT-CDCBI-CRSP_B2EN724IAKSGDTOM/2021 | EPI_ISL_3399621 | 7/29/2021 | North America | USA | Vermont |
| USA/VT-CDCBI-CRSP_EPHRB54R5NRQLUW2/2021 | EPI_ISL_3007477 | 6/24/2021 | North America | USA | Vermont |

| | | | | | |
|---|---|---|---|---|---|
| USA/VT-CDCBI-CRSP_HNUBHECBNFZKQ2VF/2021 | EPI_ISL_3408768 | 8/3/2021 | North America | USA | Vermont |
| USA/VT-CDCBI-CRSP_LH2S4RQEWGEMDTDM/2021 | EPI_ISL_3431664 | 8/7/2021 | North America | USA | Vermont |
| USA/VT-CDCBI-CRSP_MMHAHMW66IKRFFQQ/2021 | EPI_ISL_2312472 | 5/14/2021 | North America | USA | Vermont |
| USA/VT-CDCBI-CRSP_NIJIBCF3VX4DFQJY/2021 | EPI_ISL_1787299 | 4/16/2021 | North America | USA | Vermont |
| USA/VT-CDCBI-CRSP_NM3UUPB4VXOQN6Z3/2021 | EPI_ISL_3407005 | 7/27/2021 | North America | USA | Vermont |
| USA/VT-CDCBI-CRSP_SHK5GDQTTXNLKWZD/2021 | EPI_ISL_1825578 | 4/23/2021 | North America | USA | Vermont |
| USA/VT-CDCBI-CRSP_W3PGYO4AWK3PQPIO/2021 | EPI_ISL_2991991 | 5/25/2021 | North America | USA | Vermont |
| USA/VT-CDCBI-CRSP_W77IN3OVTI5EAZNY/2021 | EPI_ISL_1825284 | 4/24/2021 | North America | USA | Vermont |
| USA/WA-Altius-ALTCOV-K57SJFXJJXUMP2DJ/2021 | EPI_ISL_2378410 | 4/26/2021 | North America | USA | Washington |
| USA/WA-Altius-ALTCOV-SWQGHWVTJTAJWBES/2021 | EPI_ISL_2473168 | 4/23/2021 | North America | USA | Washington |
| USA/WA-CDC-2-4502930/2021 | EPI_ISL_2690106 | 5/24/2021 | North America | USA | Washington |
| USA/WA-CDC-8384/2020 | EPI_ISL_594459 | 2/26/2020 | North America | USA | Washington |
| USA/WA-CDC-UW21061342786/2021 | EPI_ISL_2693293 | 6/13/2021 | North America | USA | Washington |
| USA/WA-CDC-UW21071739737/2021 | EPI_ISL_3149690 | 7/17/2021 | North America | USA | Washington |
| USA/WA-CDC-UW21080135761/2021 | EPI_ISL_3411157 | 8/1/2021 | North America | USA | Washington |
| USA/WA-CDC-UW21080278512/2021 | EPI_ISL_3449126 | 8/2/2021 | North America | USA | Washington |
| USA/WA-ISB23/2020 | EPI_ISL_2504182 | 4/6/2020 | North America | USA | Washington |
| USA/WA-S3253/2020 | EPI_ISL_849408 | 10/13/2020 | North America | USA | Washington |
| USA/WA-S9618/2021 | EPI_ISL_3104862 | 6/30/2021 | North America | USA | Washington |
| USA/WA-S9813/2021 | EPI_ISL_3346910 | 7/12/2021 | North America | USA | Washington |

| | | | | | |
|---|---|---|---|---|---|
| USA/WA-UW-21050572929/2021 | EPI_ISL_2299359 | 5/5/2021 | North America | USA | Washington |
| USA/WA-UW-5359/2020 | EPI_ISL_430901 | 4/5/2020 | North America | USA | Washington |
| USA/WA-UW-6535/2020 | EPI_ISL_437859 | 4/4/2020 | North America | USA | Washington |
| USA/WI-CDC-FG-038575/2021 | EPI_ISL_2785883 | 6/19/2021 | North America | USA | Wisconsin |
| USA/WI-CDC-FG-058421/2021 | EPI_ISL_3349288 | 8/2/2021 | North America | USA | Wisconsin |
| USA/WI-CDC-FG-062235/2021 | EPI_ISL_3397174 | 8/2/2021 | North America | USA | Wisconsin |
| USA/WI-CDC-QDX27212236/2021 | EPI_ISL_3395287 | 7/24/2021 | North America | USA | Wisconsin |
| USA/WI-CDC-QDX27212301/2021 | EPI_ISL_3397100 | 7/25/2021 | North America | USA | Wisconsin |
| USA/WI-MHDL-68797/2021 | EPI_ISL_2273857 | 5/6/2021 | North America | USA | Wisconsin |
| USA/WI-UW-4229/2021 | EPI_ISL_1827550 | 4/2/2021 | North America | USA | Wisconsin |
| USA/WI-UW-5355/2021 | EPI_ISL_2833909 | 6/24/2021 | North America | USA | Wisconsin |
| USA/WI-WSLH-200403/2020 | EPI_ISL_578569 | 6/7/2020 | North America | USA | Wisconsin |
| USA/WI-WSLH-200484/2020 | EPI_ISL_578625 | 5/20/2020 | North America | USA | Wisconsin |
| USA/WI-WSLH-214565/2021 | EPI_ISL_2279239 | 4/19/2021 | North America | USA | Wisconsin |
| USA/WI-WSLH-214827/2021 | EPI_ISL_2279457 | 4/27/2021 | North America | USA | Wisconsin |
| USA/WI-WSLH-214897/2021 | EPI_ISL_2558105 | 5/3/2021 | North America | USA | Wisconsin |
| USA/WI1/2020 | EPI_ISL_408670 | 1/31/2020 | North America | USA | Wisconsin |
| USA/WV-CDC-2-4044445/2020 | EPI_ISL_1446719 | 11/13/2020 | North America | USA | West Virginia |
| USA/WV-CDC-LC0045539/2021 | EPI_ISL_1799600 | 4/16/2021 | North America | USA | West Virginia |
| USA/WV-CDC-LC0059997/2021 | EPI_ISL_2305170 | 5/12/2021 | North America | USA | West Virginia |

| | | | | | |
|---|---|---|---|---|---|
| USA/WV-CDC-LC0079872/2021 | EPI_ISL_2873092 | 6/9/2021 | North America | USA | West Virginia |
| USA/WV-CDC-LC0097344/2021 | EPI_ISL_3113467 | 7/1/2021 | North America | USA | West Virginia |
| USA/WV-CDC-LC0102156/2021 | EPI_ISL_3326385 | 7/8/2021 | North America | USA | West Virginia |
| USA/WV-CDC-MMB08661076/2021 | EPI_ISL_2285100 | 5/5/2021 | North America | USA | West Virginia |
| USA/WV-QDX-1284/2020 | EPI_ISL_572053 | 3/17/2020 | North America | USA | West Virginia |
| USA/WV-WVU-WV000860/2020 | EPI_ISL_2195496 | 11/16/2020 | North America | USA | West Virginia |
| USA/WV-WVU-WV002665/2020 | EPI_ISL_2195510 | 11/30/2020 | North America | USA | West Virginia |
| USA/WV-WVU-WV056806/2021 | EPI_ISL_2195573 | 3/31/2021 | North America | USA | West Virginia |
| USA/WV-WVU-WV064593/2021 | EPI_ISL_1624816 | 12/31/2020 | North America | USA | West Virginia |
| USA/WV-WVU-WV118690/2020 | EPI_ISL_2195722 | 5/4/2020 | North America | USA | West Virginia |
| USA/WV-WVU-WV118882/2021 | EPI_ISL_2195864 | 4/20/2021 | North America | USA | West Virginia |
| USA/WV-WVU-WV118930/2020 | EPI_ISL_2195918 | 10/14/2020 | North America | USA | West Virginia |
| USA/WV-WVU-WV118938/2020 | EPI_ISL_2195924 | 4/6/2020 | North America | USA | West Virginia |
| USA/WV-WVU-WV120893/2021 | EPI_ISL_2758364 | 6/7/2021 | North America | USA | West Virginia |
| USA/WY-CDC-IBX749918140367/2021 | EPI_ISL_2282350 | 5/5/2021 | North America | USA | Wyoming |
| USA/WY-UNM-00147/2020 | EPI_ISL_614248 | 5/26/2020 | North America | USA | Wyoming |
| USA/WY-UNM-WY00377/2020 | EPI_ISL_1626992 | 9/24/2020 | North America | USA | Wyoming |
| USA/WY-WPHL-21030504/2021 | EPI_ISL_2248720 | 4/21/2021 | North America | USA | Wyoming |
| USA/WY-WYPHL-00026/2020 | EPI_ISL_462919 | 3/21/2020 | North America | USA | Wyoming |
| USA/WY-WYPHL-00043/2020 | EPI_ISL_462935 | 3/25/2020 | North America | USA | Wyoming |

| | | | | | |
|---|---|---|---|---|---|
| USA/WY-WYPHL-00069/2020 | EPI_ISL_462961 | 3/27/2020 | North America | USA | Wyoming |
| USA/WY-WYPHL-20015627/2020 | EPI_ISL_540921 | 5/18/2020 | North America | USA | Wyoming |
| USA/WY-WYPHL-20114197/2020 | EPI_ISL_2841562 | 10/30/2020 | North America | USA | Wyoming |
| USA/WY-WYPHL-21022714/2021 | EPI_ISL_2090999 | 3/10/2021 | North America | USA | Wyoming |
| USA/WY-WYPHL-21028658/2021 | EPI_ISL_2423041 | 4/12/2021 | North America | USA | Wyoming |
| USA/WY-WYPHL-21031318/2021 | EPI_ISL_2842516 | 4/23/2021 | North America | USA | Wyoming |
| USA/WY-WYPHL-21039589A/2021 | EPI_ISL_2566087 | 5/19/2021 | North America | USA | Wyoming |
| USA/WY-WYPHL-21043353/2021 | EPI_ISL_2712934 | 6/21/2021 | North America | USA | Wyoming |
| USA/WY-WYPHL-21045605/2021 | EPI_ISL_2960292 | 6/21/2021 | North America | USA | Wyoming |
| USA/WY-WYPHL-21046420/2021 | EPI_ISL_3099556 | 7/2/2021 | North America | USA | Wyoming |
| USA/WY-WYPHL-21046993/2021 | EPI_ISL_3038876 | 7/6/2021 | North America | USA | Wyoming |
| USA/WY-WYPHL-21054770/2021 | EPI_ISL_3478315 | 8/2/2021 | North America | USA | Wyoming |
| USA/WY-WYPHL-21057332/2021 | EPI_ISL_3462413 | 8/9/2021 | North America | USA | Wyoming |
| USA/WY-WYPHL21012869/2021 | EPI_ISL_1857421 | 2/6/2021 | North America | USA | Wyoming |
| Uzbekistan/334/2021 | EPI_ISL_3188970 | 6/25/2021 | Asia | Uzbekistan | Tashkent |
| Uzbekistan/353/2021 | EPI_ISL_3188976 | 6/25/2021 | Asia | Uzbekistan | Tashkent |
| Uzbekistan/Tashkent-CGB-17/2020 | EPI_ISL_1477047 | 12/7/2020 | Asia | Uzbekistan | Tashkent |
| Venezuela/Ama1935/2021 | EPI_ISL_2628303 | 4/19/2021 | South America | Venezuela | Amazonas VE |
| Venezuela/Ara6800/2021 | EPI_ISL_2628306 | 5/10/2021 | South America | Venezuela | Aragua |
| Venezuela/DeA1365/2021 | EPI_ISL_2628302 | 4/17/2021 | South America | Venezuela | Delta Amacuro |

| | | | | | |
|---|---|---|---|---|---|
| Venezuela/Zul6567/2021 | EPI_ISL_2628310 | 5/8/2021 | South America | Venezuela | Zulia |
| Vietnam/HCMC-05109069-05/2021 | EPI_ISL_2372266 | 5/21/2021 | Asia | Vietnam | Ho Chi Minh City |
| Vietnam/VN5722/2021 | EPI_ISL_2455494 | 4/22/2021 | Asia | Vietnam | Kien Giang |
| Vietnam/VN5725/2021 | EPI_ISL_2455496 | 4/21/2021 | Asia | Vietnam | Yen Bai |
| Vietnam/VN5984/2021 | EPI_ISL_2455585 | 5/4/2021 | Asia | Vietnam | Red River Delta |
| Wales/PHWC-PYHYFD/2021 | EPI_ISL_3297090 | 7/26/2021 | Europe | United Kingdom | Wales |
| Wuhan/Hu-1/2019 | EPI_ISL_402125 | 12/26/2019 | Asia | China | Hubei |
| Wuhan/IPBCAMS-WH-02/2019 | EPI_ISL_403931 | 12/30/2019 | Asia | China | Hubei |
| Wuhan/WH01/2019 | EPI_ISL_406798 | 12/26/2019 | Asia | China | Hubei |
| Yunnan/YN-09/2021 | EPI_ISL_2840855 | 6/26/2021 | Asia | China | Yunnan |
| Yunnan/YN-10/2021 | EPI_ISL_2840856 | 6/25/2021 | Asia | China | Yunnan |
| Yunnan/YN-16/2021 | EPI_ISL_2863929 | 7/5/2021 | Asia | China | Yunnan |
| Yunnan/YN-41/2021 | EPI_ISL_2896225 | 7/8/2021 | Asia | China | Yunnan |
| Zambia/RC-138/2020 | EPI_ISL_2678308 | 12/21/2020 | Africa | Zambia | Lusaka |
| Zambia/ZMB-1324/2020 | EPI_ISL_2803648 | 4/9/2020 | Africa | Zambia | Lusaka |
| Zambia/ZMB-85345/2021 | EPI_ISL_2803568 | 2/10/2021 | Africa | Zambia | Lusaka |
| Zambia/ZMB-94233/2021 | EPI_ISL_2803635 | 4/27/2021 | Africa | Zambia | Lusaka |
| Zambia/ZMB-94239/2021 | EPI_ISL_2803629 | 4/28/2021 | Africa | Zambia | Lusaka |
| Zambia/ZMB-97951/2021 | EPI_ISL_2803677 | 5/27/2021 | Africa | Zambia | Lusaka |
| Zambia/ZMB-CH14212/2021 | EPI_ISL_2803450 | 5/2/2021 | Africa | Zambia | Lusaka |
| Zhejiang/zhoushan37/2021 | EPI_ISL_1911197 | 4/24/2021 | Asia | China | Zhejiang |
| Zimbabwe/CERI-KRISP-K011670/2020 | EPI_ISL_2492708 | 11/30/2020 | Africa | Zimbabwe | Masvingo |
| Zimbabwe/ZW-28979/2020 | EPI_ISL_1191818 | 9/1/2020 | Africa | Zimbabwe | Zimbabwe |
| Zimbabwe/ZW-29055/2020 | EPI_ISL_1191822 | 9/1/2020 | Africa | Zimbabwe | Zimbabwe |

**Table S4. Spike mutation statistics of sites with enriched mutation rates in vaccine breakthrough infections compared to unvaccinated controls.**

| Spike_pos | 19 | 19 | 19 | 97 | 98 | 112 | 142 | Ins | 156 | 157 | 158 | 222 | 289 | 452 | 478 | 485 | 493 | 677 | 681 | 722 | 879 | 950 | 1104 | 1199 | 1237 |
|---|---|---|---|---|---|---|---|---|---|---|---|---|---|---|---|---|---|---|---|---|---|---|---|---|---|
| hCoV-19/Wuhan-Hu-1 | T | K | D | K | S | S | G | - | E | F | R | A | V | L | T | G | Q | Q | P | V | A | D | V | D | M |
| spike mutation | T19R | K77N/T | D80G | K97E | S98F | S112L | G142D | -InsT | E156Δ | F157Δ/S | R158G | A222V | V289I | L452R | T478K | G485R | Q493P/R | Q677H/P | P681R/H | V722I | A879S | D950N/H | V1104L | D1199G | M1237I |
| important sites | | | | | | | | | del | | | | RBD (aa 331 - 524) | | S1/S2 cleavage | | | | | | | | | | |
| Unvacc-ctr WT (%) | 56.22 | 97.88 | 97.86 | 99.64 | 98.55 | 99.27 | 49.08 | 98.58 | 47.81 | 46.35 | 49.44 | 90.71 | 98.53 | 53.17 | 52.67 | 100 | 100 | 98.93 | 22.78 | 100 | 97.75 | 51.48 | 98.56 | 99.64 | 100 |
| Unvacc-ctr mut (%) | 43.78 | 2.12 | 2.14 | 0.36 | 1.45 | 0.73 | 50.92 | 1.42 | 52.19 | 53.65 | 50.56 | 9.29 | 1.47 | 46.83 | 47.33 | 0 | 0 | 1.07 | 77.22 | 0 | 2.25 | 48.52 | 1.44 | 0.36 | 0 |
| Vacc WT (%) | 29.07 | 95.45 | 97.69 | 98.46 | 98.46 | 89.13 | 22.66 | 97.73 | 21.71 | 19.38 | 22.4 | 84.85 | 96.9 | 25.45 | 24.39 | 97.98 | 97.98 | 98.48 | 9.85 | 98.48 | 97.66 | 20.18 | 96.95 | 98.47 | 98.48 |
| Vacc mut (%) | 70.93 | 4.55 | 2.31 | 1.54 | 1.54 | 10.87 | 77.34 | 2.27 | 78.29 | 80.62 | 77.6 | 15.15 | 3.1 | 74.55 | 75.61 | 2.02 | 2.02 | 1.52 | 90.15 | 1.52 | 2.34 | 79.82 | 3.05 | 1.53 | 1.52 |
| Δ (Vacc mut % minus Unvacc mut %) | 27.15 | 2.43 | 0.17 | 1.18 | 0.09 | 10.14 | 26.42 | 0.85 | 26.1 | 26.97 | 27.04 | 5.86 | 1.63 | 27.72 | 28.28 | 2.02 | 2.02 | 0.45 | 12.93 | 1.52 | 0.09 | 31.3 | 1.61 | 1.17 | 1.52 |
| p (Fisher) | 2.3E-05 | 0.21 | 1.00 | 0.24 | 1.00 | 4.1E-03 | 4.2E-07 | 0.68 | 4.2E-07 | 1.0E-07 | 3.2E-07 | 0.09 | 0.28 | 1.1E-06 | 1.7E-07 | 0.09 | 0.09 | 0.66 | 1.7E-03 | 0.10 | 1.00 | 1.8E-08 | 0.27 | 0.24 | 0.10 |
| q (multiplicity correction, BH) | 7.1E-05 | 0.33 | 1 | 0.34 | 1 | 0.01 | 1.7E-06 | 0.78 | 1.7E-06 | 1.3E-06 | 1.7E-06 | 0.17 | 0.34 | 4.0E-06 | 1.4E-06 | 0.17 | 0.17 | 0.78 | 4.7E-03 | 0.17 | 1 | 4.6E-07 | 0.34 | 0.34 | 0.17 |

**Table S5. Delta variant spike mutation statistics of sites with enriched mutation rates in Delta vaccine breakthrough infections compared to Delta infections in unvaccinated controls.**

| Spike_pos | 19 | 97 | 112 | 222 | 289 | 452 | 485 | 1104 | 1199 | 1237 |
|---|---|---|---|---|---|---|---|---|---|---|
| hCoV-19/Wuhan-Hu-1 | T | K | S | A | V | L | G | V | D | M |
| spike mutation | T19R | K97E | S112L | A222V | V289I | L452R | G485R | V1104L | D1199G | M1237I |
| important sites | | | | | | RBD (aa 331 - 524) | | | | |
| Unvacc-ctr WT (%) | 1.05 | 99.26 | 97.44 | 81.88 | 97.01 | 2.78 | 100 | 97.1 | 100 | 100 |
| Unvacc-ctr mut (%) | 98.95 | 0.74 | 2.56 | 18.12 | 2.99 | 97.22 | 0 | 2.9 | 0 | 0 |
| Vacc WT (%) | 0 | 98 | 81.48 | 80.2 | 95.92 | 0 | 97.26 | 96 | 98 | 98.02 |
| Vacc mut (%) | 100 | 2 | 18.52 | 19.8 | 4.08 | 100 | 2.74 | 4 | 2 | 1.98 |
| Δ (Vacc mut % minus Unvacc mut %) | 1.05 | 1.26 | 15.96 | 1.68 | 1.09 | 2.78 | 2.74 | 1.1 | 2 | 1.98 |
| p (Fisher) | 1 | 0.58 | 0.04 | 0.74 | 0.72 | 0.26 | 0.17 | 0.72 | 0.18 | 0.18 |
| q (multiplicity correction, BH) | 1 | 0.82 | 0.38 | 0.82 | 0.82 | 0.53 | 0.45 | 0.82 | 0.45 | 0.45 |

**Table S6. Delta variant full genome mutation statistics of all sites with significantly different mutation rates in Delta vaccine breakthrough infections compared to Delta infections in unvaccinated controls.**

| Full genome position | 1191 | 1267 | 12946 | 14014 | 18176 | 19160 | 20262 | 21897 | 27739 |
|---|---|---|---|---|---|---|---|---|---|
| hCoV-19/Wuhan-Hu-1 | c | c | t | t | c | c | a | c | c |
| bp mutation | c1191t | c1267t | t12946c | t14014g | c18176t | c19160t | a20262g | c21897t | c27739t |
| aa mutation | nsp2:P129L | (nsp2:G154G) | (nsp9:Y87Y) | nsp12:F192V | nsp14:P46L | nsp14:S374F | (nsp15:L214L) | S:S112L | ORF7a:L116F |
| Unvacc-ctr WT (%) | 79.9 | 79.9 | 83.5 | 96.4 | 84.4 | 85.2 | 82.2 | 97.4 | 83.9 |
| Unvacc-ctr mut (%) | 20.14 | 20.14 | 16.55 | 3.6 | 15.56 | 14.81 | 17.83 | 2.56 | 16.06 |
| Vacc WT (%) | 90.1 | 90.1 | 93.1 | 89.1 | 94.1 | 95.0 | 92.6 | 81.5 | 93.1 |
| Vacc mut (%) | 9.9 | 9.9 | 6.93 | 10.89 | 5.94 | 5.05 | 7.37 | 18.52 | 6.93 |
| Δ (Vacc mut % minus Unvacc mut %) | -10.2 | -10.2 | -9.6 | 7.3 | -9.6 | -9.8 | -10.5 | 16.0 | -9.1 |
| p (Fisher) | 0.03 | 0.03 | 0.03 | 0.03 | 0.02 | 0.02 | 0.03 | 0.04 | 0.04 |
| q (multiplicity correction, BH) | 0.04 | 0.04 | 0.04 | 0.04 | 0.04 | 0.04 | 0.04 | 0.04 | 0.04 |

**Table S7. Contingency tables and chi-square tests of unmatched and 1:1 matched data from vaccinated and unvaccinated SARS-CoV-2$^{+}$ participants.**

| unmatched | | | | | | | | |
|---|---|---|---|---|---|---|---|---|
| | Delta | Alpha | Iota | Gamma | Other | total | | |
| vaccinated | 101 | 39 | 29 | 5 | 34 | 208 | Test | Chi-square |
| unvaccinated | 143 | 371 | 363 | 27 | 425 | 1329 | P value | <0.0001 |
| total | 244 | 410 | 392 | 32 | 459 | | | **** |
| 1:1 matched | | | | | | | | |
| | Delta | Alpha | Iota | Gamma | Other | total | | |
| vaccinated | 101 | 39 | 29 | 5 | 34 | 208 | Test | Chi-square |
| unvaccinated | 99 | 45 | 30 | 6 | 28 | 208 | P value | 0.89 |
| total | 200 | 84 | 59 | 11 | 62 | | | ns |

**Table S8. Estimated model comparing the probability of a positive Delta test for the vaccinated/unvaccinated groups, adjusted for sex, age (centered and standardized), and month of test.**

| Characteristic | Vaccinated | Sex | Age | March | April | May | June | July |
|---|---|---|---|---|---|---|---|---|
| Beta estimate | 0.02 | 0 | 0 | NA | 0.01 | 0.06 | 0.37 | 0.88 |
| 95% CI | -0.01, 0.06 | -0.03, 0.02 | -0.01, 0.01 | NA | -0.02, 0.03 | 0.03, 0.10 | 0.31, 0.43 | 0.85, 0.92 |
| p value | 0.13 | 0.8 | 0.8 | NA | 0.7 | <0.001 | <0.001 | <0.001 |

CI: confidence interval.
